# Trends in hospitalization rates for ocular diseases in Brazil

**DOI:** 10.64898/2026.05.18.26353540

**Authors:** Iago Dutra, Vinícius Rodrigues Soares, Luiz Max Carvalho

## Abstract

This study mapped the age- and region-specific risks of eye diseases in the Brazilian population, evaluating temporal trends and geographical inequalities in access to healthcare. Secondary data from DATASUS, covering the 27 Brazilian federative units from 2010 to 2024, were used, employing hierarchical negative binomial regression. A significant national increase in hospital admission rates was observed during the studied period, with increases of 160.8% for retinopathy, 126.4% for eye and appendage diseases, and 122.8% for glaucoma. State-level heterogeneity was extreme, with variations spanning from -93.1% to +3588% for glaucoma, for example. Even so, regional disparities were observed throughout the period; the South region reported an average 43.2% higher than the national average for retinopathies, and the Southeast 28.5% higher for eye and adnexal diseases, while the North region reported the lowest rates. Projections up to 2036 predict a further national increase of up to +377.0% for retinopathies, with interventions covering more than an order of magnitude. In addition to the temporal projection, rates in state, age, and year components on a logarithmic scale with calibrated uncertainty were verified. Out-of-sample tests show that the chosen modeling outperforms the last observed value maintenance method and naive linear extrapolation in all three diseases considered. Thus, the escalating, age-driven burden of ophthalmological diseases and profound geographic disparities highlight an urgent need to decentralize specialized care and target resource allocation within the public health system.

## 1. Introduction

Visual impairment and blindness constitute major global public health and socioeconomic challenges. Worldwide, hundreds of millions of individuals experience moderate to severe visual impairment; however, the World Health Organization (WHO) estimates that approximately 75% of all blindness cases are preventable or treatable ^1^. The leading ophthalmic conditions driving the global burden of vision loss and legal blindness are cataracts, uncorrected refractive errors, glaucoma, age-related macular degeneration (AMD), and diabetic retinopathy ^2,3^. Specifically for retinopathies, the global epidemiological burden is rising rapidly. For example, the estimated global prevalence of diabetic retinopathy among patients with diabetes mellitus is 22.27% ^4^. In 2020, this accounted for approximately 103.12 million affected adults worldwide, of whom about 28.5 million had vision-threatening disease ^4^. Furthermore, diabetic macular edema affects over 18 million individuals, representing the leading cause of moderate vision loss among patients with type 2 diabetes ^4,5^. Additionally, AMD affects approximately 8.69% of the global population aged 45 to 85 years, historically representing the leading cause of irreversible vision loss in developed countries ^6,7^. Retinal detachment also represents a highly morbid ophthalmic emergency within this group, leading to permanent visual impairment if not treated promptly ^8^.

The development, incidence, and progression of these ophthalmic conditions are strongly associated with various demographic, genetic, and primarily clinical determinants. Age remains the most significant non-modifiable risk factor across virtually all ocular epidemiology ^6^. The risk of chronic eye diseases, such as cataracts, glaucoma, and AMD, increases exponentially due to cumulative metabolic, oxidative, and environmental damage to ocular structures over time ^9^. For instance, the prevalence of late and exudative AMD rises sharply after 75 years of age ^7^. Regarding modifiable risk factors, systemic comorbidities are central to retinal pathogenesis. For diabetic retinopathy, prolonged diabetes duration, chronic hyperglycemia, and systemic arterial hypertension are the most consistent predictors in the literature for the onset of microvascular lesions. Poor glycemic control induces capillary wall damage and secondary ischemia, whereas arterial hypertension amplifies this risk by increasing intraluminal pressure, promoting vascular leakage, precipitating hemorrhages, and inducing severe retinal ischemia ^3,5^. Additional evidence-based factors include smoking, elevated body mass index (BMI), genetic predisposition, and ethnicity; for example, populations of European ancestry show a higher prevalence of early AMD ^6,7,10^.

Driven by rapid global population aging and the rising burden of non-communicable diseases, projections for increased ophthalmic demand are critical. Globally, the number of individuals with AMD is projected to increase from 196 million in 2020 to 288 million by 2040 ^7^. Similarly, the global number of adults with diabetic retinopathy is projected to grow by over 55%, reaching 160.5 million by 2045. This increase is largely driven by rising rates of obesity and physical inactivity worldwide ^7^. This scenario indicates a profound impact in Brazil, a country undergoing an accelerated and complex demographic transition. Official estimates predict that the Brazilian elderly population will double by 2042 ^11^ and could account for approximately 35% of the total population by 2070 ^9^. This imminent aging, coupled with the high prevalence of diabetes in Brazil, affecting 9.2% of adults with a significant underreporting rate ^12^ will exert immense pressure on the healthcare system regarding the demand for ocular interventions ^9,11^.

Within the Brazilian public health system, managing complex ocular pathologies is challenged by structural, geographic, and socioeconomic disparities. An estimated 65% of the Brazilian population (over 160 million individuals) relies exclusively on the Unified Health System (SUS) for ophthalmic consultations, screenings, and surgeries ^11^. However, the provision of specialized care is limited and heavily concentrated in major economic centers, leading to significant healthcare inequities. For instance, while the Southeast region has one ophthalmologist per 7,599 inhabitants, the North region has only one per 12,084 inhabitants ^13^. Data from the DATASUS clearly reflect these practical inequities. Between 2010 and 2019, official records showed a sharp increase (1,088%) in intravitreal injections for macular edema and AMD, alongside a substantial rise in laser photocoagulation ^13^. Nevertheless, most of these procedures were performed in the Southeast and South regions, highlighting the access deficit in the North and Northeast ^13^. For acute conditions such as retinal detachment, incidence rates and hospitalization volumes follow the same geographic concentration, illustrating that patients outside major urban centers face critical barriers to timely diagnosis and vision-saving interventions ^8,13-15^. The inadequate provision of essential preventive exams, such as funduscopy in primary care units, exacerbates these disparities and perpetuates a cycle where patients seek emergency care only after visual damage is severe or permanent ^12^. These barriers have driven the implementation of teleophthalmology initiatives aimed at reducing extensive waiting lists and promoting effective, democratized screening within primary care ^11,16,17^.

Brazil possesses one of the world’s largest public health reporting systems, and recent epidemiological literature has effectively utilized DATASUS to map aging related problems^18^, chronic conditions (e.g., leukemia, diabetes, Alzheimer’s ^19-23^) and infectious outbreaks (e.g., chikungunya, syphilis ^24-26^), ophthalmological public health has been largely overlooked. Aside from the tracking of highly specific procedural interventions like intravitreal injections and photocoagulation ^8,9,13^. Valuable studies have mapped specific interventions, such as the rising volume of intravitreal injections, or focused on the localized burden of isolated pathologies like retinal detachment and age-related cataracts^8,9,13^. Building upon this foundational work, there is a growing opportunity to integrate these disease-specific and age-restricted analyses into a comprehensive, nationwide framework that systematically correlates ocular health with underlying demographic and regional shifts. While absolute case counts are vital for evaluating local hospital capacity, relative indicators are essential to accurately assess epidemiological risk across populations of varying sizes and densities. Transitioning from absolute occurrence data to integrated relative indicators allows for a clearer visualization of regional inequities. By cross-referencing multiple ocular diseases by age and geographic macro-region alongside robust predictive modeling, this study aims to refine the current understanding of the disease burden and provide actionable projections for future healthcare planning.

In this context, establishing robust baselines derived from relative risk indicators is essential to inform public health policies, decentralize healthcare services, optimize telemedicine initiatives, and promote sustainable patient education guidelines ^8,11,27,28^. Given the need for resource optimization amidst the budgetary constraints of the SUS and rapid population aging, this investigation is highly warranted. Therefore, the primary objective of this study is to map the age and region-specific risks of ocular diseases in the Brazilian population using DATASUS records. Specifically, this research aims to: (1) describe the prevalence and relative incidence rates of major ophthalmic disorders; (2) evaluate the association between advancing age and the increased risk of requiring hospital and outpatient interventions; and (3) quantify between-state and between-region heterogeneity in hospitalization rates for these conditions, which combines underlying disease burden with healthcare-supply factors and cannot be cleanly decomposed into “access” versus “incidence” with administrative data alone.

## 2. Methods

### 2.1. Study Design and Data Source

Nationwide secondary data from the DATASUS, covering all 27 federative units (26 states plus the Federal District) over the period 2010-2024 were collected. Hospital admission counts were extracted from the Hospital Information SIH/SUS for three groupings of ICD-10 codes: glaucoma (H40-H42); diseases of the retina, choroid, and vitreous body (H30-H36), hereafter referred to as “retinopathies”; and the broader category of diseases of the eye and appendages (H00-H59). Data were aggregated by state, calendar year, and seven age strata (25-34, 35-44, 45-54, 55-64, 65-74, 75-89, and 90+ years), yielding up to 2,835 state-year-age cells per disease (27 × 15 × 7). Cells with counts below the SIH/SUS reporting threshold (recorded as missing in the public extract) were excluded from estimation; sensitivity to this choice was assessed in a parallel analysis treating these cells as zeros. The year 2025 was excluded due to incomplete reporting at the time of extraction. As a complementary test, we held out the most recent two complete years (2023 and 2024) and refit the model on 2010-2022 to assess the model’s predictive capacity. The model’s posterior predictive distribution recovered observed 2023-2024 hospitalization rates with MAE 9.3 (glaucoma), 39.2 (retinopathy) and 182.1 (eye and appendages) per million, outperforming naïve last-value-carried-forward (LVCF) and linear-extrapolation baselines on retinopathy and eye-and-appendage diseases (Supplementary Table 5). For glaucoma, for which yearly variation is smaller, LVCF performed marginally better, indicating the temporal structure adds little value at the national level for this condition.

Population denominators were obtained from the official 2024 revision of the Brazilian Institute of Geography and Statistics (IBGE) population projections ^29^, aggregated to match the seven age strata. Hospitalization rates were expressed as cases per million inhabitants. We use the term “administrative prevalence” throughout to indicate that these rates capture healthcare utilization recorded in SIH/SUS rather than true epidemiological prevalence.

### 2.2. Statistical Model

A series of statistical models was tested to represent the data, with different temporal dynamics, dispersion structures, and age and state effect modeling. Specifications of every studied variant can be found in the Supplementary Material. Model selection was based on the leave-one-out information criterion (LOO-IC). For each disease, two variants tied within one LOO-IC standard error, and the simpler specification was chosen on grounds of parsimony. Convergence and posterior predictive checks are reported in the Supplementary Material.

The selected final model was a hierarchical negative binomial regression on the count data, with the logarithm of the population as offset and a categorical age structure as a fixed effect. Let *S* and *A* be the set of states and the set of age strata, respectively. Let *s* ∈ *S* be a state, *a* ∈ *A* be an age stratum and *t* be the year, the model is:

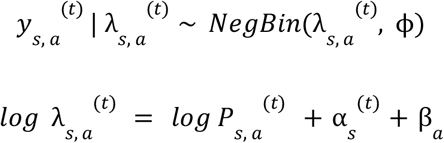

where *P* _*s, a*_ ^(*t*)^ is the population denominator (offset), α _*s*_ ^(*t*)^ is a state-specific time-varying intercept, and β _*a*_ is the deflection for the age stratum *a*. Identifiability between α _*s*_ ^(*t*)^ and the age effects was enforced via a sum-to-zero constraint on the seven age coefficients 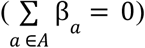,implemented through a deflection coding parameterization in Stan. Temporal smoothing was imposed by a first-order Gaussian random walk on the state intercepts:

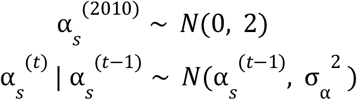

Weakly informative priors were used throughout: ϕ ∼ *Half* – *Normal*(0, 10) and σ_α_ ∼ *Half* – *Normal*(0, 1). Posterior inference used Hamiltonian Monte Carlo via Stan ^30^, with four chains of 4,000 iterations each (2,000 warmup), adapt_delta = 0.8, and maximum tree depth = 12. Convergence was assessed by R-hat < 1.01 and minimum effective sample size > 400 across all monitored parameters.

Robustness of the inferred parameters to the priors was assessed by refitting the baseline model under five alternative initializations: *Half* – *Normal*(0, 10), *Half* – *Normal*(0, 3), *Gamma*(2, 0. 1), *Exponential*(0. 1) and *Half* – *Cauchy*(0, 5). Two further sensitivity analyses re-estimated the selected model after (a) excluding the year 2024 to test whether incomplete recent reporting biased the trend estimates, and (b) recording suppressed cells as zero counts rather than missing. The invariance of final predictions to those perturbations is shown in the Supplementary Material.

To assess forecast calibration, the selected model was refit on data restricted to 2010-2022 and used to predict the held-out 2023-2024 observations. Predictions were compared against two non-Bayesian baselines: a last-value-carried-forward predictor using the 2022 state-level rate, and a state-specific linear extrapolation fit on 2018-2022 data. Metrics included mean absolute error (MAE), mean relative error (MRE), root mean squared error (RMSE), bias on the rate scale, and empirical coverage of the 80% and 95% posterior predictive intervals, summarised in Table 1 and Table 2.

**Table 1.** Model Fitness Metrics for Age Subgroups.

| Disease | Subgroup | MRE | MAE | RMSE | Bias |
| --- | --- | --- | --- | --- | --- |
| Glaucoma | 25-34 | 0.64 | 1.55 | 2.5 | -0.09 |
|  | 35-44 | 0.56 | 1.87 | 2.74 | -0.01 |
|  | 45-54 | 0.73 | 3.33 | 5.1 | 0.18 |
|  | 55-64 | 1.11 | 6.17 | 9.77 | 0.42 |
|  | 65-74 | 2.05 | 12.58 | 19.86 | -0.5 |
|  | 75-89 | 3.10 | 24.7 | 38.47 | -1.22 |
|  | 90+ | 3.57 | 23.94 | 38.01 | 1.99 |
| Retinopathy | 25-34 | 0.61 | 6.08 | 8.7 | 0.69 |
|  | 35-44 | 0.52 | 6.99 | 10.08 | 1.11 |
|  | 45-54 | 0.63 | 12.31 | 17.93 | 2.11 |
|  | 55-64 | 1.47 | 30.62 | 45.04 | -0.41 |
|  | 65-74 | 1.95 | 46.79 | 72.25 | -5.26 |
|  | 75-89 | 3.30 | 72.62 | 112.87 | -8.92 |
|  | 90+ | 2.77 | 66.39 | 101.86 | -1.97 |
| Eye & App. | 25-34 | 0.55 | 32.17 | 47.26 | 9.67 |
|  | 35-44 | 0.46 | 36.07 | 53.55 | 13.28 |
|  | 45-54 | 0.43 | 52.31 | 80.55 | 18.66 |
|  | 55-64 | 0.90 | 102.4 | 139.78 | 19.23 |
|  | 65-74 | 2.27 | 309.02 | 475.86 | -83.43 |
|  | 75-89 | 4.04 | 685.26 | 1109.49 | -240.27 |
|  | 90+ | 5.85 | 480.33 | 759.58 | -121.4 |

**Table 2.** Model Fitness Metrics for Region Subgroups.

| Disease | Subgroup | MRE | MAE | RMSE | Bias |
| --- | --- | --- | --- | --- | --- |
| Glaucoma | North | 3.98 | 8.29 | 17.55 | 0.65 |
|  | Northeast | 1.22 | 9.23 | 20.96 | 0.13 |
|  | Center-West | 1.48 | 15.09 | 28.5 | -1.93 |
|  | Southeast | 0.59 | 8.04 | 17.05 | -3.72 |
|  | South | 0.40 | 15.1 | 27.03 | 6.67 |
| Retinopathy | North | 5.10 | 26.11 | 68.55 | -7.59 |
|  | Northeast | 0.39 | 32.07 | 55.37 | 0.93 |
|  | Center-West | 0.82 | 43.41 | 76.35 | -1.67 |
|  | Southeast | 0.17 | 31.67 | 52.48 | 1.03 |
|  | South | 0.18 | 51.71 | 88.35 | -0.76 |
| Eye & App. | North | 7.14 | 209.06 | 567.11 | -90.52 |
|  | Northeast | 0.32 | 201.37 | 393.99 | -32.71 |
|  | Center-West | 0.32 | 385.37 | 864.98 | -114.47 |
|  | Southeast | 0.24 | 274.15 | 492.52 | -118.33 |
|  | South | 0.28 | 211.31 | 344.25 | 125.69 |

State-level posterior projections of the hospitalization rate from 2025 to 2036 were obtained by forward-simulating the random walk on α _*s*_ ^(*t*)^ for each posterior draw, holding the categorical age coefficients fixed and using IBGE projected populations as future denominators. For each draw, predicted state-year cases were summed across age groups; regional and national rates were obtained by aggregating predicted cases across the relevant states and dividing by the aggregated regional or national population. Posterior summaries are reported as means with 80% and 95% credible intervals.

## 3. Results

### 3.1. Temporal Trends of Retinopathy Prevalence by Region

Over the analyzed period from 2010 to 2024, the data revealed a nationwide increase in the prevalence of hospitalizations and interventions for retinal and other ocular disorders (Eye: 126.4%; Retina: 160.8%; Glaucoma: 122.8% national increase in 2024 compared to 2010). These pooled national rates conceal substantial between-state heterogeneity over the same period: observed 2010-to-2024 changes ranged from -36.4% (Espírito Santo) to +7,612% (Rondônia) for retinopathies, from -68.6% (Tocantins) to +1,719% (Rondônia) for eye-and-appendage diseases, and from -93.1% (Alagoas) to +3,588% (Pará) for glaucoma. Between two and five states recorded decreases over the period for each disease, while a handful of small-population Northern states drove the upper tail through very low 2010 baselines. Median state-level changes were +202.1% (retinopathy), +138.6% (eye and appendage) and +126.4% (glaucoma), with interquartile ranges of roughly 67-796%, 18-282% and 19-342% respectively. State-by-state observed changes are tabulated in Supplementary Material. Regional stratification highlighted significant geographical disparities, with the South region demonstrating a consistently higher prevalence of hospitalizations for retinopathy, maintaining a constant trend above the national average (South: 252.0 vs 176.0, 43.2% higher) (Fig 1A). This geographical concentration in the South and Southeast also extended to broader categories, showing a higher prevalence of hospitalizations for general diseases of the eye and appendage diseases in Southeast (Southeast: 934.2 vs 727.0, 28.5% higher) (Fig 1B). Furthermore, hospitalizations for glaucoma were notably more prevalent in the South, Southeast, and Center-West regions, with the South consistently tracking above the national average (South: +8.9%; Southeast: +11.5%; Center-West: +15.0%) (Fig 1C). In contrast, the North region historically reported the lowest volume of cases (Fig 1A-C, red lines), usually remaining below the national average prevalence of hospitalizations throughout the studied period (Eye: 61.0% below; Retina: 53.0% below; Glaucoma: 72.2% below). Meanwhile, the Northeast region exhibited a steady pattern, recording prevalence rates that were very close to the national average across all evaluated ocular diseases (Eye: 21.0% difference; Retina: 17.1% difference; Glaucoma: 9.9% difference).

**Figure 1.**
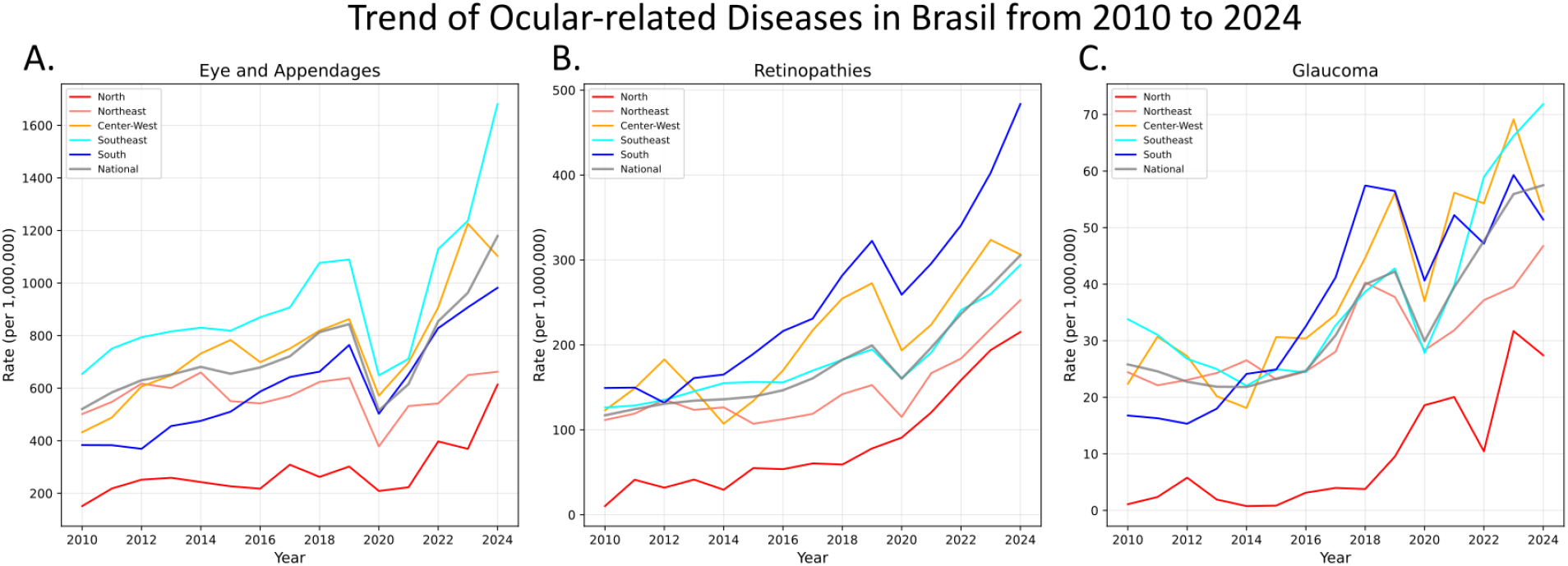
Temporal trends of ocular-related diseases in Brazil (2010-2024). The line charts illustrate the annual incidence rates (per million population) for (A) Eye and Appendages, (B) Retinopathies, and (C) Glaucoma over a 15-year observational period. The data is stratified by geographic macro-region: North (red), Northeast (salmon), Center-West (orange), Southeast (cyan), and South (blue). The overall national trend is represented by the solid gray line. The y-axis scales are adjusted independently for each panel to accommodate the differing prevalence magnitudes among the specific disease categories.

### 3.2. Age-Stratified Temporal Dynamics of Retinopathy Prevalence

Hospitalization rates for all ocular diseases demonstrated a pronounced, age-dependent increase across all regions (Fig 2A-C). Nationally in 2024, comparing the 25-34 and 90+ age cohorts, glaucoma surged 42.9-fold (3.6 to 154.3 per million), followed by eye and appendages diseases (18.4-fold; 161.0 to 2956.7) (Fig 2A), and retinopathies (14.6-fold; 32.6 to 476.7) (Fig 2B). Regional ordering remained strictly consistent across all age strata for eye-and-appendage diseases, with the highest rates in the Southeast and the lowest in the North. Observed 2024 rates in the Southeast rise from 222.1 per million in the 25-34 stratum (posterior mean 250.1, 95% CI 214.8-293.2) to 4,100.9 in the 90+ stratum (posterior mean 3,684.9, 95% CI 3,188.1-4,303.5), an 18.5-fold between-cohort age difference within the same region (Fig 2A). Retinopathy trajectories largely mirrored the non-stratified findings, particularly in the South, Southeast, and Center-West (Fig 2B). In the South, observed 2024 rates ranged from 49.7 per million in the 25-34 stratum (posterior mean 61.2, 95% CI 56.1-66.5) to 798.6 in the 90+ stratum (posterior mean 639.0, 95% CI 586.4-695.7). Similar age-related increases were observed in the Southeast, from 34.7 (posterior mean 38.0, 95% CI 34.8-41.4) to 429.6 (posterior mean 409.6, 95% CI 376.0-446.9), and in the Center-West, from 42.0 (posterior mean 45.0, 95% CI 41.1-49.5) to 444.2 (posterior mean 483.8, 95% CI 441.2-533.8). However, a distinct divergence emerged in the North among individuals older than 45 years. Between 2020 and 2024, this demographic showed progressive rate increases. For example, the North’s 45-54 cohort rose from 77.2 to 107.7 per million. Although the national weighted average also increased (104.2 to 150.2), the absolute gap between the North and the national rate widened from 27.0 to 42.5 (Fig 2B), a divergence that remained consistent across older age strata. Glaucoma rates were highest in the Center-West across all age strata, increasing from 4.9 per million at 25-34 (posterior mean 5.4, 95% CI 4.6-6.4) to a peak of 246.8 at 75-89 (posterior mean 224.7, 95% CI 193.0-260.7), before declining to 134.9 at 90+ (posterior mean 143.6, 95% CI 123.0-166.9) (Fig 2C). This non-monotonic age pattern was consistent across regions and is likely explained by survivor selection. The model captured the decline well, although posterior uncertainty increased substantially in the oldest strata.

**Figure 2.**
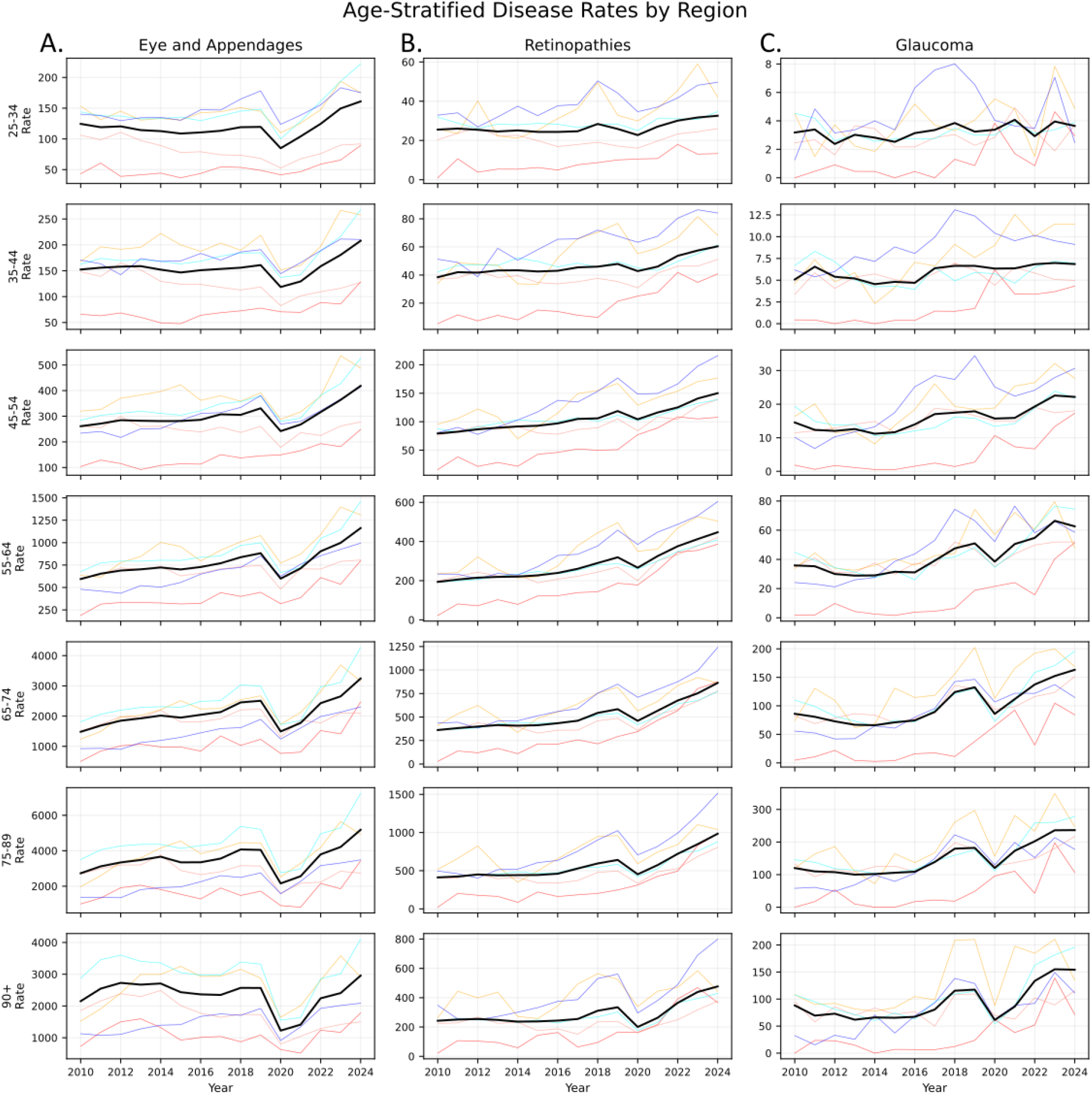
Age-stratified historical ocular disease rates by region (2010-2024). The panels display the historical incidence rates for (A) Eye and Appendages, (B) Retinopathies, and (C) Glaucoma, comprehensively stratified by age group. The grid’s rows represent distinct age cohorts in ascending order, from 25-34 years (top row) to 90+ years (bottom row). The x-axis covers the observational period from 2010 to 2024. Within each individual panel, the thick black line denotes the overall national rate, while the thinner, lighter-colored lines represent the respective regional rates (North, Northeast, Center-West, Southeast, and South). The y-axis scales vary independently across both disease categories and age cohorts to accommodate the changing magnitude of disease prevalence in older populations.

### 3.3. Evaluation of Predictive Model Performance Across Demographic Subgroups

The epidemiological profiles of ophthalmic diseases demonstrate marked demographic and geographic heterogeneity, with disease trajectories exhibiting increased variance and complexity with advancing patient age and across diverse regional landscapes. The Eye and Appendages category highlights this demographic complexity most prominently: the magnitude of variation escalates sharply in older populations, reflecting a substantial concentration of clinical burden in elderly cohorts (mean observed rate ≈ 3,507 per million in the 75-89 stratum; absolute bias -244.2 per million, relative bias -7.0%, indicating mild under-prediction relative to the underlying rate). Spatial analysis further reveals pronounced regional disparities, with localized disease dynamics presenting pronounced variations in the Center-West (RMSE: 864.98) and a distinct directional variance in the South (bias: 125.69). Retinopathy trends closely mirror this age-dependent progression of clinical complexity, where the relative stability of disease indicators observed in younger demographics (MRE: 0.61 in ages 25-34) broadens significantly in older, more vulnerable populations (MRE: 3.30 in ages 75-89), compounded by spatial disparities that result in notable regional variance. In contrast, Glaucoma profiles suggest a less demographically and geographically volatile disease progression across the population, maintaining a relatively stable distribution across all age brackets and regions (MRE < 4.0).

### 3.4. Long-Term State-Level Forecasting of Retinopathy Subtypes to 2036

We projected age-standardized rates (per million inhabitants) and 95% Credible Intervals (CI) for major ocular diseases from 2025 to 2036 (Fig 3A-C). Nationally, retinopathy may experience the largest relative increase (+377.0%), rising from 226.2 in 2024 to 1,078.8 (95% CI: 7.8-7,241.7) by 2036. Glaucoma follows with a 346.1% increase (from 31.2 to 139.1; 95% CI: 1.4-873.9), while Eye and Appendage diseases are projected to grow by 124.4% (from 727.6 to 1,632.7; 95% CI: 71.5-8,444.8). In every case, uncertainty increases markedly over the long-term forecasting horizon. Regionally, the epicenters of projected growth vary by condition, the Center-West leads in Eye and Appendage diseases (+134.6%, reaching 2,765.3), the South in retinopathies (+394.1%, reaching 2,326.8), and the Southeast in glaucoma (+425.5%, reaching 286.4). In stark contrast, the North consistently exhibits the lowest relative growth across all three categories. These heterogeneous trajectories may reflect distinct regional risk factor profiles, diagnostic capacities, and healthcare access patterns.

**Figure 3.**
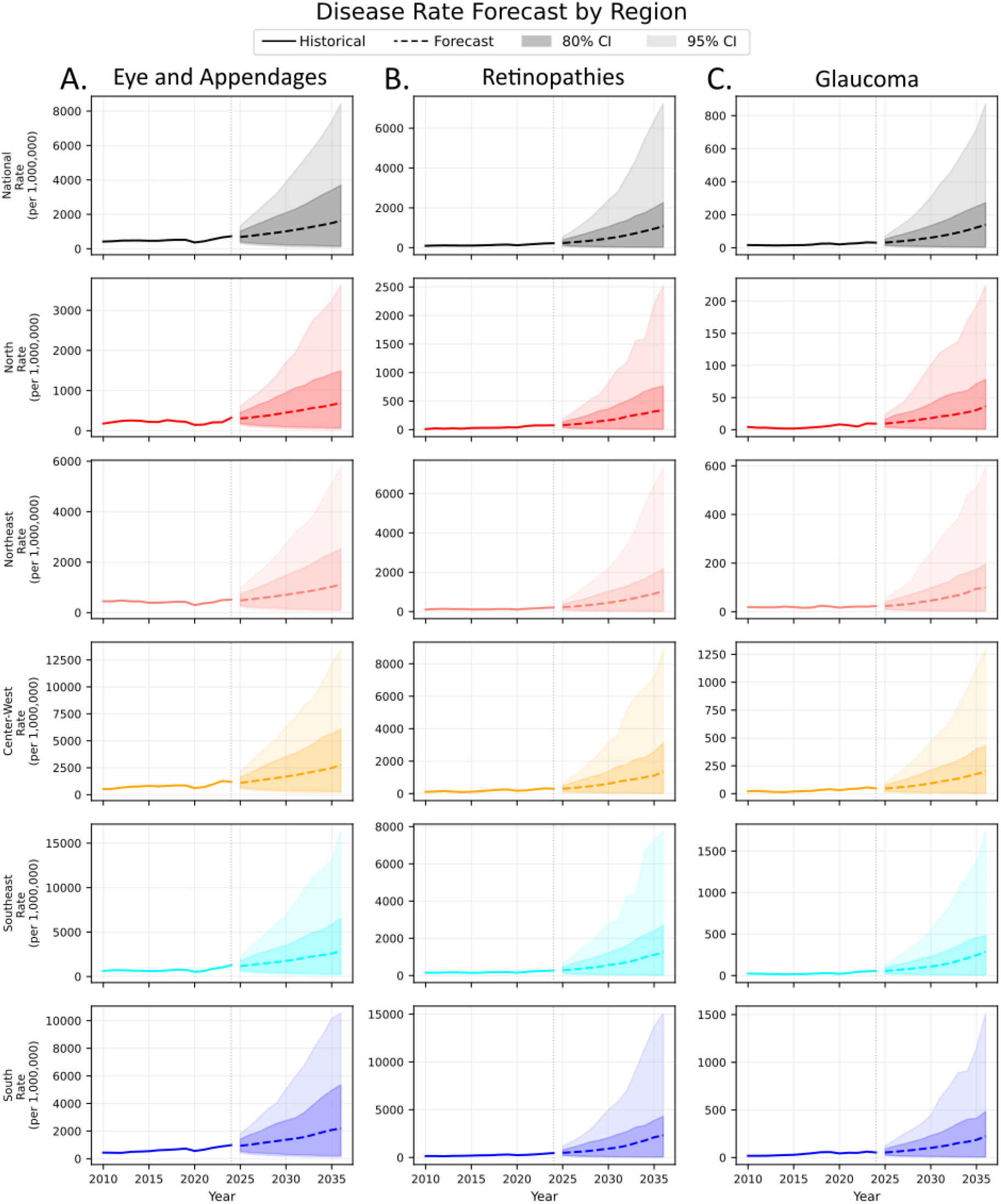
Regional forecasts for ocular disease rates. The panels display the historical and projected incidence rates per million individuals for Eye and Appendages (A), Retinopathies (B), and Glaucoma (C). Rows correspond to varying geographic regions: National (black), North (red), Northeast (salmon), Center-West (orange), Southeast (cyan), and South (blue). Solid lines indicate historical observed data, while dashed lines represent the forecasted rates. The darker and lighter shaded areas indicate the 80% and 95% credible intervals (CIs) of the forecasts, respectively. A vertical dotted line marks the transition from historical data to the forecast period. The y-axis scales vary independently across both disease categories to accommodate the changing magnitude of disease prevalence in regions.

## 4. Discussion

To the best of our knowledge, this study provides the first comprehensive, long-term forecasting of hospitalization rates for major ocular diseases within the SUS up to 2036. Our projections reveal an escalation in the ophthalmic disease burden, driven primarily by a nearly five-fold expected increase in retinopathies and a significant rise in glaucoma admissions over the next decade. Furthermore, we identified stark geographic inequalities that mirror global epidemiological variations. Our findings underscore the growing burden of eye diseases in Brazil, particularly retinopathy and glaucoma, which may soon require substantial increases in screening capacity, specialized care, and surgical resources. Regional projections highlight the Center-West and Southeast as hotspots for Eye & Appendage Diseases and Glaucoma, respectively, while the South faces the fastest growth in retinopathy. These projections provide critical evidence for targeted health system planning and resource allocation through 2036.

The epidemiological profiles of ophthalmic diseases demonstrate marked demographic heterogeneity and a clear age-dependence, with disease trajectories exhibiting increased variance and clinical complexity in advancing age. The “Eye and Appendages” category highlights this demographic shift most prominently, this clinical burden is overwhelmingly concentrated in elderly cohorts ^9^. Retinopathy trends closely mirror this age-dependent progression, the relative stability of disease indicators observed in younger demographics broadens into significantly higher predictive variance in older, more vulnerable populations. This escalating retinopathy burden in older adults aligns with the natural history of specific posterior segment diseases, such as retinal detachment, which sees a distinct hospitalization peak in the 60-69 age group due to age-associated vitreoretinal degeneration ^8^, and age-related macular degeneration (AMD), which exhibits a rapid surge in global prevalence and severity after 65 to 75 years of age ^6,7^.

Beyond national tracking, understanding the epidemiology of ocular diseases requires recognizing spatial disparities, just as prevalence rates fluctuate heavily across global continents, our data reveals profound regional heterogeneity within Brazil. In international perspective, a meta-analysis of the European Eye Epidemiology Consortium found that glaucoma prevalence varies within Europe itself ^31^. Studies from Eastern Europe and Southern Europe showed a significantly higher prevalence of glaucoma compared to Western Europe ^31^. In another meta-analysis, diabetic retinopathy prevalence varied drastically worldwide, peaking in regions like Africa (35.90%) and North America, while remaining significantly lower in South and Central America (13.37%) ^4^. For example, the Western Pacific region currently has the largest absolute number of people with diabetic retinopathy (31.50 million in 2020) and is projected to maintain this lead through 2045 due to the sheer size of its diabetic population ^4^. For glaucoma, based on recent projections, the open-angle glaucoma burden in Tropical Latin America, the epidemiological region encompassing Brazil, is estimated to affect 3.14 million people (with a prevalence of 3.5%) in 2024, and this number is projected to more than double to reach 6.66 million affected individuals by 2060 ^32^, contrasting with our data of hospitalization with a fourfold increase in the next ten years.

Similarly, AMD prevalence in Asia is less than half of that observed in Europe and North America ^7^. These international patterns underscore the heavy influence of geographic and demographic factors on ophthalmic disease burden, a phenomenon closely reflected in our national data, which demonstrates that the disproportionately high rates of ocular interventions in the South and Southeast contrast sharply with the lower volumes in the North.

Furthermore, our spatial analysis indicates that while the South, Southeast, and Center-West maintained consistent retinopathy trendlines across age groups, the North region exhibited a significant increase in hospitalizations specifically for the 45 to 90+ demographic over the last five years, bringing its rates close to the national average. In contrast, while the overall prevalence of glaucoma increases substantially with advancing age, rising from 0.22% in patients in their early 40s to over 10% in populations over 85 ^31^, its regional distribution remains a relatively stable across all age brackets.

A critical limitation when interpreting these epidemiological profiles is the profound underdiagnosis prevalent in the North region, which skews the true burden of asymptomatic diseases like glaucoma. The North suffers from marked healthcare disparities, characterized by a severe scarcity of specialists ^13^ and alarming general underreporting rates, reaching up to 72.8% for conditions like diabetes ^12^. Because early-stage glaucoma is asymptomatic and its proper detection requires complex ^33^, specialized examinations, up to 90% of affected individuals in Brazil may remain completely unaware of their condition^1^. Consequently, the historically low volume of glaucoma hospitalizations recorded in the North and Northeast likely reflects this massive diagnostic deficit rather than a genuinely lower disease prevalence. The artificially low hospitalization rates in the Northeast may also be exacerbated by higher overall mortality in diabetic patients, who develop other complications (such as kidney failure or amputations) that cause them to postpone ocular care ^13^. Moreover, DATASUS relies on administrative records that lack granular clinical details and entirely exclude data from the private healthcare sector. Data accuracy is further compromised by reporting biases, administrative input errors, and the grouping of distinct conditions under generic diagnostic codes ^34^. Finally, because the system records hospitalization events rather than individual patients, readmissions frequently inflate the actual number of cases ^20,25,34^.

Acknowledging this data limitation is of paramount importance, as it calls for the decentralization of specialized eye care and the implementation of targeted, opportunistic screening programs in these vulnerable regions to prevent irreversible blindness.

## 5. Conclusion

The epidemiological burden of ocular diseases in Brazil is rising rapidly, driven by demographic aging and rising diabetes prevalence, Brazil faces an escalating public health challenge from ocular diseases. Hospitalizations for retinopathies, glaucoma, and appendage diseases surged between 2010 and 2024, and are projected to grow by 2036, though long-horizon forecasts carry substantial uncertainty. This rising demand exposes stark regional inequities, with specialized care concentrated in the South and Southeast, leaving the North and Northeast with critical deficits in access. Mitigating widespread preventable vision loss requires the decentralization of ophthalmic care within the SUS, prioritizing teleophthalmology and equitable resource distribution, prioritizing targeted, state-specific resource allocation rather than uniform expansion, especially in regions with documented underdiagnosis.

## Data Availability

All used data was collected from public sources mentioned in the text and are available as clean, preprocessed datasets in the project repository [https://github.com/DutraIRS/Ocular-Pathologies-in-DataSUS].

https://github.com/DutraIRS/Ocular-Pathologies-in-DataSUS

https://datasus.saude.gov.br/informacoes-de-saude-tabnet/

## Software and reproducibility

All analyses were conducted in R version 4 using the loo^21^, rstan^22^ and bridgesampling^23^ packages. The complete source code, intermediate diagnostic outputs, posterior summaries, and chart-data CSVs are publicly available at the project repository [https://github.com/DutraIRS/Ocular-Pathologies-in-DataSUS].

## Ethical considerations

This study used exclusively de-identified, publicly available aggregate data from DATASUS and IBGE. In accordance with Resolution 510/2016 of the Brazilian National Health Council, research using public-domain secondary data does not require Research Ethics Committee approval.

## Contributor Roles

VS: Conceptualization, Investigation, Writing - Original Draft. ID: Software, Conceptualization, Investigation, Writing - Original Draft. LC: Supervision, Validation, Writing - Review & Editing.

## Data Availability

All used data was collected from public sources mentioned in the text and are available as clean, preprocessed datasets in the project repository.

## Acknowledgements

The authors acknowledge the Departamento de Informática do Sistema Único de Saúde (DATASUS) and the Brazilian Ministry of Health for their ongoing commitment to open science and public health surveillance. Furthermore, we appreciate the tireless efforts of the healthcare workers, hospital administrators, and local health departments across all Brazilian macro-regions who meticulously register and compile this vital epidemiological information.

