## Supplemental Material for "Trends in hospitalization rates for ocular diseases in Brazil"

May 19, 2026

**3 Model Selection** **11**

**4 Out-of-sample Validation** **13**

**5 Subgroup Performance of the Selected Model** **13**

**6 Posterior Predictive Checks** **15**

**9 Observed State-level Change 2010-2024** **46**

**10 Posterior Credible Intervals on 2024 Age × Region Cells** **47**

**11 Forecasts** **50**

### 1 Model Development

#### 1.1 Common framework

Let $y_{s,t,a}$ be the observed count of hospitalizations for a single disease in state $s = 1, \ldots, 27$, year $t = 1, \ldots, 15$ (2010-2024), and age stratum $a \in \{25\text{-}34, 35\text{-}44, 45\text{-}54, 55\text{-}64, 65\text{-}74, 75\text{-}89, 90+\}$. Population denominators $P_{s,t,a}$ come from IBGE projections. Exploratory analysis revealed marginal variances much larger than the means at every level of disaggregation, so a Poisson likelihood was ruled out. We therefore work throughout with a Negative Binomial of the second kind,

$$y_{s,t,a} \mid \lambda_{s,t,a}, \phi \;\sim\; \text{NegBin}_2(\lambda_{s,t,a}, \phi), \qquad \text{Var}(y) = \lambda + \lambda^2/\phi,$$

with the standard log link and a population offset:

$$\log \lambda_{s,t,a} \;=\; \log P_{s,t,a} \;+\; \alpha_{s,t} \;+\; \mathcal{A}_a.$$

The state-year intercept $\alpha_{s,t}$ carries all of the spatial and temporal structure, and $\mathcal{A}_a$ is an age contribution that we vary across specifications. All models use the common prior $\phi \sim \mathcal{N}^+(0, 10)$ on the overdispersion parameter, with sensitivity to this choice reported in Section 7.

#### 1.2 Initial exploration: nine baseline structures

Before settling on a final pool of candidates, we screened nine baseline structures (`nb0-nb8`) covering the natural progression from a global intercept to fully state-specific dynamics in both intercept and age components. Each model retains the Negative Binomial likelihood and the log-population offset; they differ only in how $\alpha_{s,t}$ and $\mathcal{A}_a$ are parameterized:

- **nb0** (intercept only): $\log \lambda_{s,t,a} = \log P_{s,t,a} + \beta_0$, with global $\beta_0$ and $\phi$ only.
- **nb1**: state-specific intercepts $\alpha_s$ and a global linear age effect $\beta x_a$, $x_a = (\text{age}_a - 60)/10$.
- **nb2**: state-specific intercepts $\alpha_s$ *and* state-specific age slopes $\beta_s$.
- **nb3**: state-fixed intercepts with a global *time-varying* age effect, $\beta_t \sim \mathcal{N}(\beta_{t-1}, \sigma_\beta)$.
- **nb4**: state-and-time-varying age effect $\beta_{s,t} \sim \mathcal{N}(\beta_{s,t-1}, \sigma_\beta)$.
- **nb5**: a single global, time-invariant age slope with a state-level random walk on the intercept, $\alpha_{s,t} \sim \mathcal{N}(\alpha_{s,t-1}, \sigma_\alpha)$.
- **nb6**: as nb5 but with state-specific (time-constant) age slopes $\beta_s$.
- **nb7**: dynamic state intercepts *and* a time-varying global age effect.
- **nb8**: fully dynamic - both $\alpha_{s,t}$ and $\beta_{s,t}$ evolve as state-specific random walks.

Performance summaries for the nine baseline structures on the glaucoma series are shown in Table 1; analogous patterns hold for retinopathy and eye-and-appendage diseases. A clear pattern emerges. `nb4` (27 state-specific age-slope walks on a fixed-intercept backbone) and `nb8` (state-specific walks on both intercept and slope) fail to mix on $T = 15$ years of data: the level and slope components of `nb8` are not jointly identified at this temporal resolution, and `nb4` is over-parameterized for the available time series. Their LOOIC values are not interpretable and are reported for completeness only. `nb7` (dynamic state intercepts plus a global time-varying age slope) mixes cleanly but offers no LOOIC improvement over `nb5`, indicating that the additional temporal flexibility on the age effect is not supported by the data once the state-level walk is in place. Among the structures that mix cleanly, `nb5` achieved the lowest LOOIC by a wide margin (over 700 points below `nb1-nb4`). `nb6` achieved a marginally lower LOOIC than `nb5` (13 998 vs. 14 080 for glaucoma) but at the cost of 26 additional state-specific slope parameters; the same gap appears for the other two diseases.

Table 1: Initial screening of nine baseline structures on the glaucoma series. Analogous rankings hold for retinopathy and eye-and-appendage diseases.

| **Model** | **LOOIC** | $LOOIC_{SE}$ | **WAIC** | $WAIC_{SE}$ | $\log ML$ | $\log ML_{SE}$ | **Max** $\hat{R}$ | **Min** $N_{eff}$ |
|---|---|---|---|---|---|---|---|---|
| nb0 | 18098.68 | 221.35 | 18098.68 | 221.35 | -9055.67 | ¡0.001 | 1.0013 | 6264 |
| nb1 | 14918.31 | 204.61 | 14918.23 | 204.61 | -7891.40 | ¡0.001 | 1.0013 | 5519 |
| nb2 | 14874.24 | 204.65 | 14873.27 | 204.62 | -7883.37 | ¡0.001 | 1.0017 | 2900 |
| nb3 | 14919.92 | 204.64 | 14919.82 | 204.64 | -7882.81 | 0.001 | 1.0061 | 312 |
| nb4 | 14878.18 | 204.72 | 14877.08 | 204.69 | -7541.71 | 0.357 | 1.3696 | 12 |
| nb5 | 14080.10 | 197.96 | 14074.73 | 197.82 | -7228.77 | 0.078 | 1.0029 | 2564 |
| nb6 | 13998.36 | 198.34 | 13991.67 | 198.19 | -7209.09 | 0.171 | 1.0031 | 2082 |
| nb7 | 14083.89 | 197.99 | 14078.54 | 197.85 | -7219.41 | 0.598 | 1.0085 | 544 |
| nb8 | 14001.07 | 198.44 | 13994.19 | 198.28 | -6859.11 | 0.978 | 1.2839 | 17 |

We therefore adopt `nb5` as the *trunk* structure for the remainder of the development: a state-level Gaussian random walk on the intercept, a single global age effect, and a Negative Binomial likelihood. Subsequent variants modify this trunk by changing the prior process on $\alpha_{s,t}$ (toward drift, autoregression, local linear trend) and by changing the functional form of $\mathcal{A}_a$ (toward quadratic or categorical). The numeral *5* that subsequent variants carry as a prefix is therefore a label, not a rank - it indicates that the variant inherits the `nb5` likelihood and offset structure.

#### 1.3 Diagnosing the trunk: why nb5 alone is not enough

Although `nb5` dominates the initial nine-model screening on LOOIC, its subgroup performance (Table 2) shows substantial systematic bias concentrated in the oldest age strata. For all three diseases the bias in the 90+ stratum is one to three orders of magnitude larger than in any of the four youngest strata; for retinopathy the model under-predicts the 90+ rate by 656 per million on average (MAE 660, RMSE 818), and for eye-and-appendage diseases the bias reaches +2067 per million in the 90+ cohort. The pattern reflects the linearity of the age effect: a single global slope cannot accommodate the non-monotonic age gradient (rise to a peak in 75-89, plateau or modest decline at 90+) that the data exhibit consistently across regions and diseases. This motivated a structured exploration of alternative age specifications and alternative temporal dynamics, described next.

Table 2: Subgroup performance of the trunk `nb5` model. Note the large bias and RMSE in the 75-89 and 90+ strata for all three diseases - the motivation for developing the 19-variant pool of refinements (Section 1.4).

| **Disease** | **Group** | **Subgroup** | **N** | **MAE** | **RMSE** | **Bias** | **Cov_80** | **Cov_95** |
|---|---|---|---|---|---|---|---|---|
| *Glaucoma* | Age | 25-34 | 345 | 2.39 | 3.65 | 1.58 | 92.2 | 98.0 |
| | Age | 35-44 | 390 | 3.19 | 4.88 | 2.45 | 97.9 | 99.7 |
| | Age | 45-54 | 390 | 3.65 | 5.51 | 1.36 | 98.7 | 100.0 |
| | Age | 55-64 | 375 | 8.93 | 14.44 | -6.39 | 95.5 | 98.9 |
| | Age | 65-74 | 405 | 28.17 | 47.81 | -24.83 | 84.4 | 98.8 |
| | Age | 75-89 | 375 | 33.18 | 58.65 | -21.88 | 94.9 | 99.5 |
| | Age | 90+ | 345 | 136.38 | 192.36 | 134.55 | 77.1 | 95.4 |
| *Retinopathy* | Age | 25-34 | 405 | 14.65 | 19.89 | 13.83 | 93.3 | 99.8 |
| | Age | 35-44 | 405 | 20.82 | 28.52 | 20.25 | 98.3 | 99.8 |
| | Age | 45-54 | 405 | 14.44 | 20.45 | 6.51 | 100.0 | 100.0 |
| | Age | 55-64 | 405 | 87.35 | 119.49 | -84.57 | 87.7 | 98.3 |
| | Age | 65-74 | 405 | 174.24 | 238.45 | -170.35 | 81.5 | 99.5 |
| | Age | 75-89 | 405 | 86.15 | 140.69 | -35.25 | 99.0 | 99.5 |
| | Age | 90+ | 390 | 659.58 | 818.19 | 655.95 | 42.3 | 88.7 |
| *Eye & App. Dis.* | Age | 25-34 | 405 | 35.59 | 52.84 | 17.14 | 90.4 | 99.0 |
| | Age | 35-44 | 405 | 69.72 | 103.10 | 63.43 | 91.1 | 99.0 |
| | Age | 45-54 | 405 | 95.51 | 141.96 | 85.57 | 98.0 | 100.0 |
| | Age | 55-64 | 405 | 103.22 | 144.43 | -50.06 | 98.5 | 100.0 |
| | Age | 65-74 | 405 | 619.69 | 942.16 | -597.09 | 73.8 | 96.3 |
| | Age | 75-89 | 405 | 930.55 | 1613.11 | -800.54 | 81.7 | 96.3 |
| | Age | 90+ | 405 | 2073.55 | 2532.44 | 2067.34 | 52.1 | 89.1 |

##### 1.4 Refined candidate pool: nineteen variants

To address the limitations of the trunk model and to assess the sensitivity of inference to plausible alternative specifications, we developed a refined pool of nineteen variants. All inherit the Negative Binomial likelihood and the population offset from `nb5`; they differ in two orthogonal axes.

*Axis 1: temporal dynamics of* $\alpha_{s,t}$.

- **Family A - single-level random walk**: `nb5` (baseline RW on $\alpha_{s,t}$ with $\sigma_\alpha \sim \mathcal{N}^+(0, 1)$); `nb5_expo` ($\sigma_\alpha \sim$ Exp(1)); `nb5_tight_sigma` ($\sigma_\alpha \sim$ Exp(5)); `nb5_drift`, `nb5_drift_expo`, `nb5_drift_tight` (with a non-negative shared drift $\mu \sim$ Exp(1)).
- **Family B - local linear trend (two-level RW)**: `nb5_llt`, `nb5_llt_expo`, `nb5_llt_drift`, `nb5_llt_drift_expo`, `nb5_llt_damped` (Holt-Winters damping $\varphi \sim$ Beta(8, 2)).
- **Family C - stationary autoregressive**: `nb5_ar1` (mean-reverting AR(1) to a state-specific level $\mu_s$, $\rho \sim$ Beta(8, 2)); `nb5_ar1_hier` (hierarchical pooling on $\mu_s$); `nb5_ar1_trend` (AR(1) around a state intercept plus a global linear trend); `nb5_ar2` (AR(2) parameterized via partial autocorrelations).

*Axis 2: functional form of $\mathcal{A}_a$.*

- **Linear** (default for most variants): $\mathcal{A}_a = \beta\, x_a$, $x_a = (\text{age}_a - 60)/10$, $\beta \sim \mathcal{N}(0, 2)$.
- **Quadratic** (`_age_quad` suffix): $\mathcal{A}_a = \beta_1 x_a + \beta_2 x_a^2$. Variants in the pool: `nb5_age_quad`, `nb5_ar1_age_quad`.
- **Categorical** (`_age_cat` suffix, sum-to-zero deflection coding): $\mathcal{A}_a = \beta_{\text{age},j(a)}$ with $\sum_j \beta_{\text{age},j} = 0$. Variants in the pool: `nb5_age_cat`, `nb5_ar1_age_cat`.

Family B (local linear trend) variants fail to converge cleanly on $T = 15$ years for the same identification reason that prevented `nb4`, `nb7` and `nb8` from mixing in the initial screening: the level and slope components of a two-level random walk are not jointly identified on a short series. We report these variants in the cross-model comparison (Section 3, Table 3) for completeness, marked with an asterisk; they are not retained as selection candidates because their LOOIC values are not interpretable. All other Family A and Family C variants converged cleanly.

#### 2 Convergence Diagnostics for the Selected Model

We report MCMC convergence diagnostics for the selected `nb5_age_cat` fits used in the main analysis: autocorrelation function (ACF) for global parameters across chains, and trace plots for the same parameters. Maximum $\hat{R}$ and minimum effective sample size for every parameter are summarized in Table 3 (selected-model row). Complete reproduction code and additional checks can be found in the project repository.

#### 2.1 Glaucoma

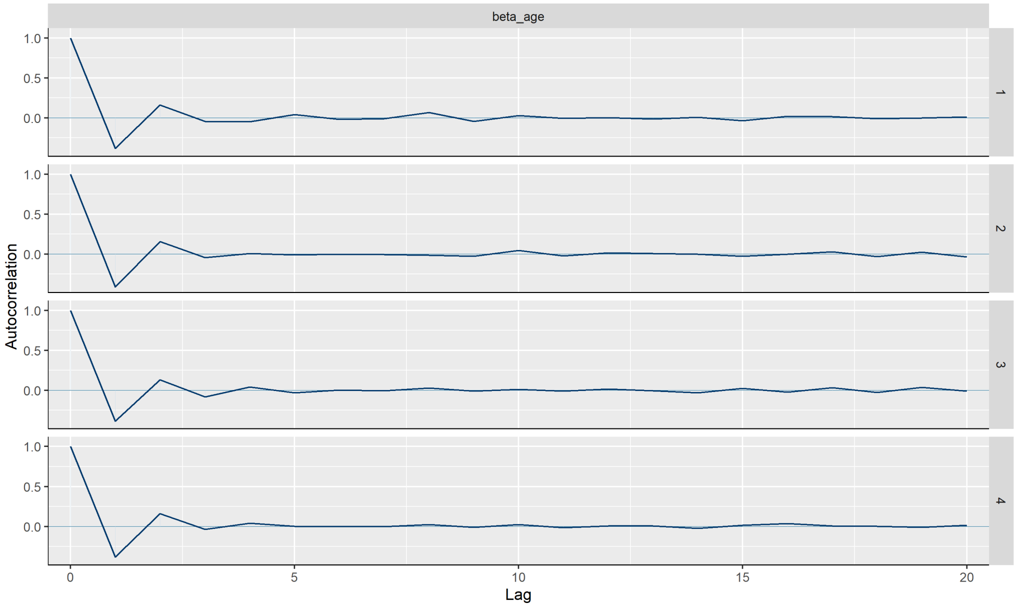

Figure 1: Auto-Correlation Function - Glaucoma.

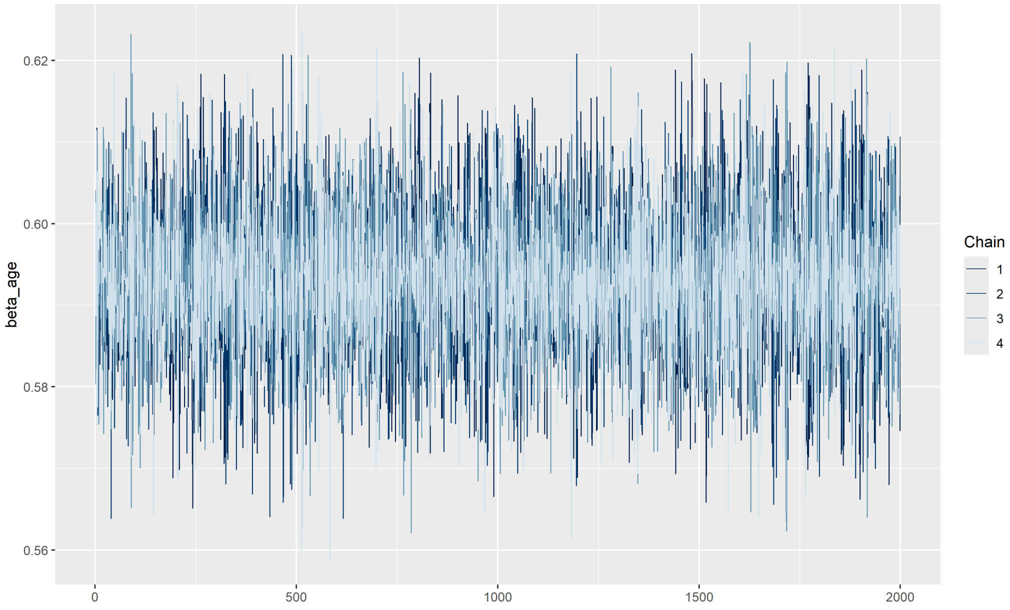

Figure 2: Trace Plot - Glaucoma.

#### 2.2 Retinopathy

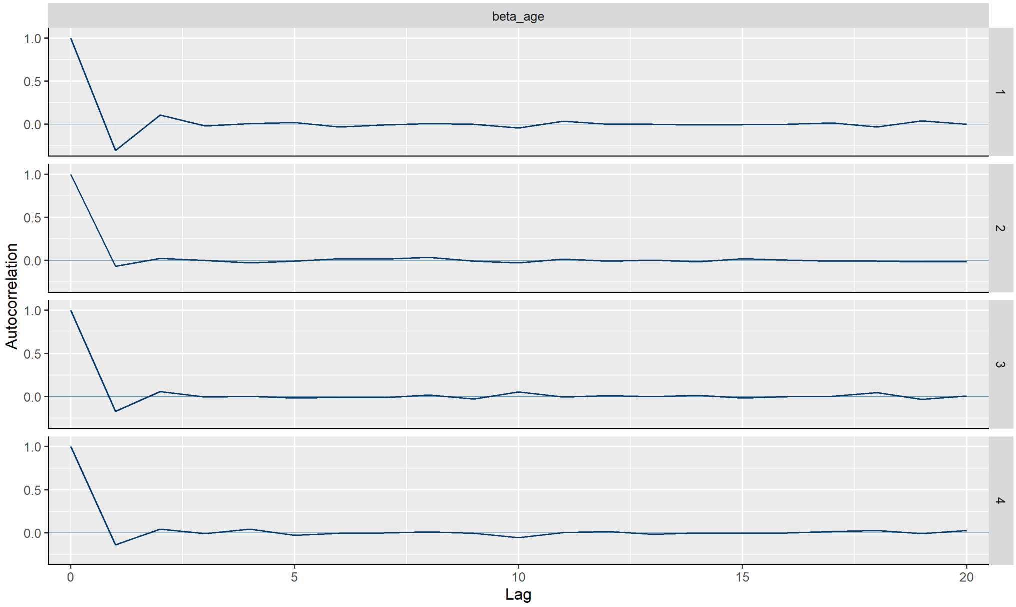

Figure 3: Auto-Correlation Function - Retinopathy.

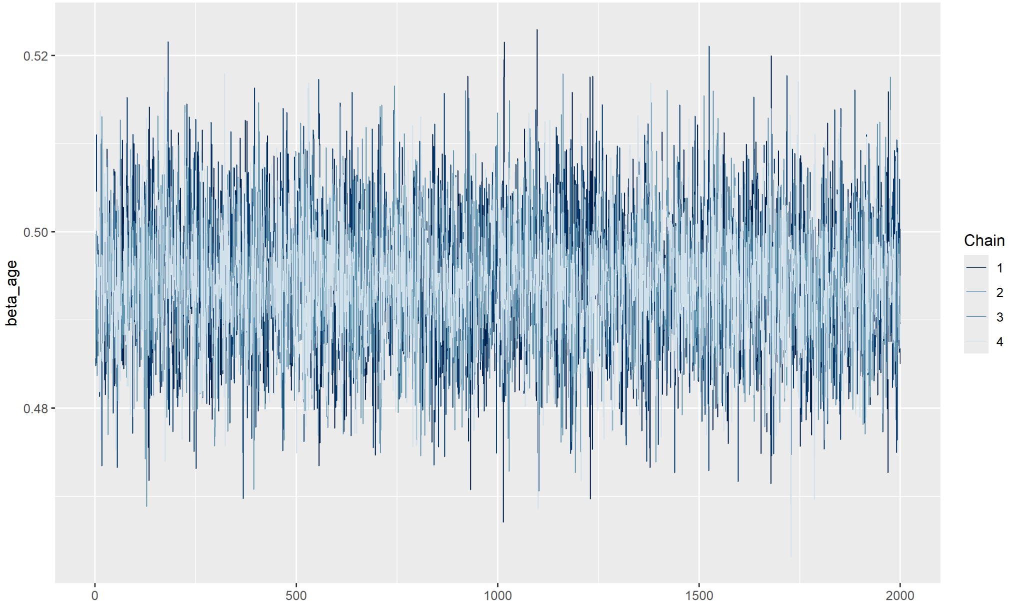

Figure 4: Trace Plot - Retinopathy.

#### 2.3 Eye & Appendage Diseases

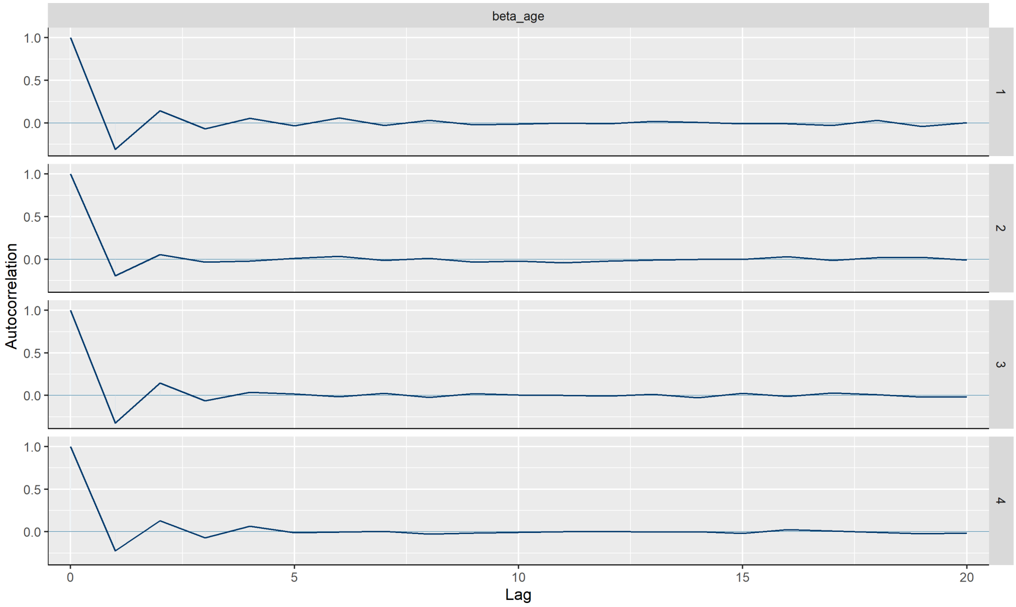

Figure 5: Auto-Correlation Function - Eye & Appendage Diseases.

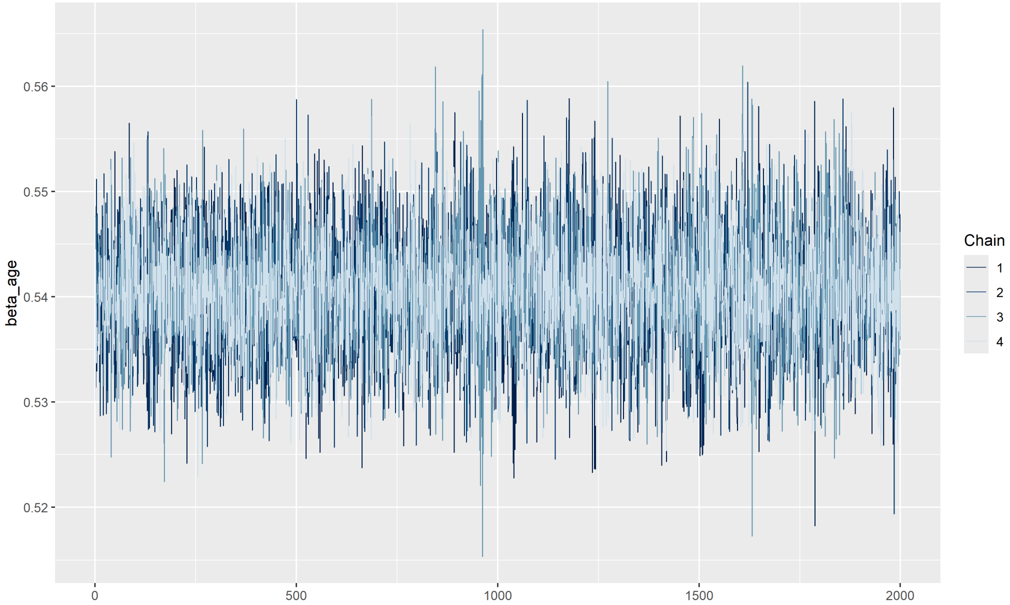

Figure 6: Trace Plot - Eye & Appendage Diseases.

### 3 Model Selection

Selection from the refined pool of 19 variants (Section 1.4) followed the leave-one-out information criterion (LOO-IC), with ties defined by overlap of LOO-IC $\pm 1$ SE intervals and broken by structural parsimony. Convergence was *not* used as a selection criterion - variants with marginal MCMC mixing remain in the comparison and are flagged in Table 3 with an asterisk, but as discussed in Section 1.4 the LOOIC values for the non-identified Family B variants are uninterpretable in any case. Their presence in the table is for transparency only.

Table 3 reports LOO-IC, MRE, MAE, RMSE, coverage and convergence summaries for the 19 variants on all three diseases. For each disease, two variants tied within one LOO-IC standard error: `nb5_ar1_age_cat` achieved the numerically lowest LOO-IC, and `nb5_age_cat` was within $|\Delta\text{LOO-IC}| = 3.2$-$34.9$ versus SE = 174-192 on all three diseases. The simpler `nb5_age_cat` (state-level Gaussian random walk with sum-to-zero categorical age effects) was chosen on grounds of parsimony.

Table 3: Cross-model predictive performance for the 19 candidate variants across all three diseases, ordered by LOO-IC within disease. Asterisk (*) flags variants with marginal MCMC convergence ($\hat{R} \geq 1.01$ or $N_{eff} < 400$); these are reported for transparency but, given the LOO-IC gaps in each disease, none of them are among the candidates that tied with the selected model.

| **Variant** | **Disease** | **LOOIC** | $_{SE}$ | **MAE** | **MRE** | **RMSE** | **Cov**$_{80}$ | **Cov**$_{95}$ | **Max** $\hat{R}$ | **Min** $N_{eff}$ |
|---|---|---|---|---|---|---|---|---|---|---|
| nb5_ar1_age_cat | Glaucoma | 12354.8 | 176.1 | 10.16 | 1.42 | 21.59 | 91.1 | 97.5 | 1.0033 | 1049 |
| nb5_age_cat | Glaucoma | 12389.7 | 174.4 | 10.47 | 1.66 | 21.95 | 90.8 | 97.8 | 1.0034 | 2539 |
| nb5_ar1_age_quad | Glaucoma | 12961.4 | 187.6 | 13.08 | 1.52 | 28.85 | 90.4 | 97.8 | 1.0040 | 1404 |
| nb5_age_quad | Glaucoma | 12993.4 | 186.1 | 13.27 | 1.75 | 28.79 | 90.3 | 98.1 | 1.0028 | 2581 |
| nb5_ar1_hier* | Glaucoma | 14046.0 | 199.5 | 29.26 | 2.50 | 75.26 | 91.9 | 98.7 | 1.0167 | 209 |
| nb5_ar1_trend | Glaucoma | 14046.1 | 199.8 | 29.13 | 2.46 | 74.88 | 91.0 | 98.7 | 1.0049 | 1071 |
| nb5_ar1 | Glaucoma | 14046.8 | 199.8 | 29.19 | 2.45 | 75.01 | 91.4 | 98.6 | 1.0061 | 1120 |
| nb5_drift | Glaucoma | 14077.6 | 197.9 | 29.67 | 2.80 | 75.72 | 90.7 | 98.8 | 1.0029 | 2418 |
| nb5_drift_expo | Glaucoma | 14078.8 | 198.0 | 29.66 | 2.79 | 76.00 | 91.6 | 98.6 | 1.0049 | 2171 |
| nb5_drift_tight | Glaucoma | 14079.3 | 197.9 | 29.72 | 2.77 | 76.27 | 91.4 | 98.9 | 1.0019 | 2308 |
| nb5_expo | Glaucoma | 14079.6 | 197.9 | 29.59 | 2.77 | 75.41 | 91.2 | 98.8 | 1.0031 | 2057 |
| nb5 | Glaucoma | 14080.6 | 198.0 | 29.62 | 2.78 | 75.88 | 91.1 | 98.7 | 1.0028 | 2313 |
| nb5_tight_sigma | Glaucoma | 14081.2 | 198.0 | 29.62 | 2.78 | 75.98 | 91.2 | 98.9 | 1.0027 | 2431 |
| nb5_llt_damped* | Glaucoma | 14082.3 | 197.9 | 29.67 | 2.81 | 75.72 | 91.2 | 98.5 | 1.4894 | 11 |
| nb5_llt* | Glaucoma | 14088.0 | 198.1 | 29.63 | 2.81 | 75.76 | 91.5 | 98.6 | 1.2968 | 20 |
| nb5_llt_expo* | Glaucoma | 14089.0 | 198.1 | 29.62 | 2.81 | 75.95 | 91.2 | 98.9 | 1.4324 | 9 |
| nb5_llt_drift* | Glaucoma | 14089.0 | 198.1 | 29.84 | 2.84 | 76.44 | 91.0 | 98.7 | 1.2197 | 16 |
| nb5_llt_drift_expo* | Glaucoma | 14091.6 | 198.1 | 29.67 | 2.81 | 75.93 | 91.3 | 98.8 | 1.1225 | 21 |
| nb5_ar2 | Glaucoma | 14126.0 | 200.2 | 29.60 | 2.47 | 78.26 | 93.0 | 99.0 | 1.0066 | 874 |
| nb5_ar1_age_cat | Retinopathy | 20624.6 | 193.0 | 34.13 | 1.46 | 65.81 | 86.4 | 97.1 | 1.0036 | 2237 |
| nb5_age_cat | Retinopathy | 20653.2 | 191.9 | 34.37 | 1.60 | 66.16 | 86.5 | 97.0 | 1.0045 | 2367 |
| nb5_ar1_age_quad | Retinopathy | 22257.2 | 208.4 | 50.17 | 1.55 | 91.51 | 88.0 | 97.7 | 1.0037 | 2263 |
| nb5_age_quad | Retinopathy | 22287.4 | 207.3 | 49.89 | 1.69 | 90.55 | 88.3 | 97.9 | 1.0039 | 2510 |
| nb5_ar2 | Retinopathy | 24897.0 | 223.6 | 148.35 | 2.96 | 329.26 | 86.7 | 98.3 | 1.0029 | 1501 |
| nb5_ar1_trend | Retinopathy | 24910.4 | 223.4 | 146.85 | 2.99 | 322.00 | 86.2 | 98.2 | 1.0040 | 2699 |
| nb5_ar1 | Retinopathy | 24911.0 | 223.6 | 146.54 | 3.00 | 319.80 | 85.8 | 98.3 | 1.0040 | 2848 |
| nb5_ar1_hier* | Retinopathy | 24912.1 | 223.5 | 146.47 | 3.03 | 319.61 | 86.5 | 98.4 | 1.0110 | 230 |
| nb5_llt* | Retinopathy | 24935.6 | 222.1 | 147.66 | 3.26 | 324.74 | 86.8 | 97.9 | 1.7274 | 4 |
| nb5_llt_expo* | Retinopathy | 24937.8 | 222.1 | 147.94 | 3.25 | 324.13 | 86.7 | 97.9 | 1.3159 | 12 |
| nb5_drift_tight | Retinopathy | 24939.4 | 221.9 | 148.13 | 3.28 | 324.61 | 86.1 | 98.0 | 1.0030 | 2109 |
| nb5_llt_drift* | Retinopathy | 24939.6 | 222.1 | 147.96 | 3.26 | 325.50 | 86.5 | 98.0 | 1.3684 | 8 |
| nb5_drift_expo | Retinopathy | 24940.2 | 222.0 | 148.22 | 3.25 | 325.12 | 86.4 | 98.1 | 1.0028 | 1691 |
| nb5_drift | Retinopathy | 24940.8 | 221.9 | 148.32 | 3.27 | 325.69 | 86.5 | 97.8 | 1.0023 | 2100 |
| nb5_llt_drift_expo* | Retinopathy | 24941.3 | 222.1 | 147.91 | 3.28 | 325.27 | 86.6 | 97.9 | 1.1870 | 18 |
| nb5 | Retinopathy | 24941.4 | 222.1 | 148.22 | 3.28 | 324.75 | 86.6 | 98.2 | 1.0025 | 2257 |
| nb5_expo | Retinopathy | 24942.1 | 222.1 | 148.34 | 3.28 | 325.43 | 86.7 | 98.2 | 1.0020 | 2686 |
| nb5_llt_damped* | Retinopathy | 24942.2 | 222.0 | 148.26 | 3.28 | 325.88 | 86.6 | 98.2 | 1.3879 | 9 |
| nb5_tight_sigma | Retinopathy | 24944.5 | 222.0 | 148.40 | 3.25 | 325.69 | 86.1 | 97.9 | 1.0027 | 2431 |
| nb5_ar1_age_cat | Eye & App. Dis. | 30292.8 | 190.7 | 242.21 | 2.05 | 541.62 | 85.4 | 96.4 | 1.0044 | 2468 |
| nb5_age_cat | Eye & App. Dis. | 30296.0 | 190.4 | 242.51 | 2.09 | 543.20 | 85.4 | 96.3 | 1.0038 | 2341 |
| nb5_ar1_age_quad | Eye & App. Dis. | 31783.2 | 205.6 | 348.41 | 2.30 | 747.83 | 84.9 | 97.4 | 1.0022 | 2771 |
| nb5_age_quad | Eye & App. Dis. | 31785.0 | 204.8 | 347.63 | 2.35 | 741.74 | 84.7 | 97.2 | 1.0024 | 3087 |
| nb5_ar2 | Eye & App. Dis. | 32471.0 | 206.7 | 562.94 | 3.40 | 1192.29 | 83.6 | 96.5 | 1.0049 | 624 |
| nb5_llt_damped* | Eye & App. Dis. | 32473.0 | 206.8 | 563.18 | 3.36 | 1193.29 | 83.2 | 96.9 | 1.1158 | 37 |
| nb5_ar1 | Eye & App. Dis. | 32479.0 | 207.7 | 557.67 | 3.20 | 1179.01 | 83.5 | 97.1 | 1.0027 | 2956 |
| nb5_ar1_trend | Eye & App. Dis. | 32479.3 | 207.7 | 557.23 | 3.24 | 1180.57 | 83.8 | 97.2 | 1.0030 | 2840 |
| nb5_ar1_hier* | Eye & App. Dis. | 32480.1 | 207.7 | 557.49 | 3.27 | 1179.78 | 83.5 | 97.1 | 1.0174 | 298 |
| nb5_drift_tight | Eye & App. Dis. | 32481.3 | 207.2 | 561.26 | 3.31 | 1193.46 | 83.7 | 97.1 | 1.0029 | 2958 |
| nb5_drift_expo | Eye & App. Dis. | 32481.5 | 207.2 | 560.74 | 3.30 | 1189.38 | 83.7 | 97.1 | 1.0026 | 3010 |
| nb5_expo | Eye & App. Dis. | 32483.4 | 207.2 | 558.46 | 3.30 | 1181.49 | 84.1 | 97.0 | 1.0023 | 2995 |
| nb5_drift | Eye & App. Dis. | 32483.4 | 207.2 | 558.25 | 3.30 | 1181.84 | 83.7 | 97.2 | 1.0025 | 3123 |
| nb5_tight_sigma | Eye & App. Dis. | 32483.6 | 207.2 | 558.43 | 3.35 | 1183.74 | 83.6 | 96.9 | 1.0032 | 2881 |
| nb5_llt_expo* | Eye & App. Dis. | 32483.7 | 207.2 | 559.40 | 3.31 | 1186.60 | 83.8 | 97.3 | 1.3173 | 18 |
| nb5_llt* | Eye & App. Dis. | 32484.3 | 207.1 | 561.15 | 3.27 | 1192.99 | 83.7 | 97.1 | 1.2013 | 21 |
| nb5 | Eye & App. Dis. | 32485.5 | 207.2 | 558.80 | 3.29 | 1179.92 | 84.2 | 97.0 | 1.0031 | 2544 |
| nb5_llt_drift* | Eye & App. Dis. | 32486.2 | 207.1 | 560.82 | 3.28 | 1192.04 | 83.3 | 97.3 | 1.1517 | 37 |
| nb5_llt_drift_expo* | Eye & App. Dis. | 32486.4 | 207.1 | 560.99 | 3.31 | 1193.28 | 83.5 | 97.1 | 1.6578 | 4 |

### 4 Out-of-sample Validation

To assess predictive performance under temporal hold-out, we refit `nb5_age_cat` on data restricted to 2010-2022 and forecasted state-year rates for the held-out 2023 and 2024 (54 state-year cells per disease). Performance was compared against two naive baselines: last-value-carried-forward (LVCF) using 2022 as the predictor, and a state-specific linear extrapolation fit to 2018-2022. Results are reported in Table 4.

The fitted hierarchical model outperformed both naive baselines on retinopathy and eye-and-appendage diseases across all summary metrics. For glaucoma, where year-on-year variation at the national level is small, LVCF marginally beat the hierarchical model on MAE (8.62 vs. 9.28 per million) and MRE (0.40 vs. 0.52), indicating that the temporal structure adds substantive value primarily where rates exhibit material trend rather than near-stationarity. Coverage of 95% predictive intervals from the model ranged from 90.7% (glaucoma) to 94.4% (retinopathy), close to the nominal value.

Table 4: Out-of-sample (2023-2024) holdout performance of `nb5_age_cat` vs. naive baselines. Training set: 2010-2022. $N_{obs} = 54$ state-year cells per row.

| **Variant** | **Disease** | **MAE** | **MRE** | **RMSE** | **Bias** | **Cov$_{80}$** | **Cov$_{95}$** |
|---|---|---|---|---|---|---|---|
| nb5_age_cat | Glaucoma | 9.28 | 0.52 | 14.93 | -0.04 | 74.1 | 90.7 |
| naive_LVCF | Glaucoma | 8.62 | 0.40 | 14.09 | -5.38 | - | - |
| naive_linear | Glaucoma | 11.44 | 1.06 | 16.67 | -7.22 | - | - |
| nb5_age_cat | Retinopathy | 39.18 | 0.50 | 61.59 | 0.79 | 87.0 | 94.4 |
| naive_LVCF | Retinopathy | 54.05 | 0.48 | 76.16 | -37.08 | - | - |
| naive_linear | Retinopathy | 60.94 | 0.69 | 89.54 | -40.71 | - | - |
| nb5_age_cat | Eye & App. Dis. | 182.07 | 0.33 | 315.39 | -137.29 | 83.3 | 92.6 |
| naive_LVCF | Eye & App. Dis. | 194.37 | 0.36 | 308.08 | -157.81 | - | - |
| naive_linear | Eye & App. Dis. | 309.17 | 0.64 | 449.60 | -262.20 | - | - |

### 5 Subgroup Performance of the Selected Model

Subgroup performance metrics for the selected `nb5_age_cat` model are reported in Tables 5 and 6. The mean relative error (MRE) is reported alongside MAE to enable scale-invariant comparison across age and region strata; relative bias is reported as $100 \times \text{Bias}/\overline{\text{obs}}$, where $\overline{\text{obs}}$ is the mean observed rate in the stratum.

Table 5: Subgroup performance for the selected model `nb5_age_cat` (Part 1: Error Metrics). All rates are expressed per million inhabitants.

| Disease | Subgroup | N | MAE | MRE | RMSE |
|---|---|---|---|---|---|
| *Glaucoma* | 25–34 | 345 | 1.56 | 0.64 | 2.50 |
| | 35–44 | 390 | 1.88 | 0.56 | 2.76 |
| | 45–54 | 390 | 3.33 | 0.73 | 5.13 |
| | 55–64 | 375 | 6.10 | 1.11 | 9.74 |
| | 65–74 | 405 | 12.48 | 2.05 | 19.75 |
| | 75–89 | 375 | 24.59 | 3.10 | 38.38 |
| | 90+ | 345 | 24.10 | 3.57 | 37.93 |
| | North | 570 | 8.31 | 3.98 | 17.71 |
| | Northeast | 900 | 9.24 | 1.22 | 20.88 |
| | Center-West | 420 | 15.11 | 1.48 | 28.41 |
| | Southeast | 420 | 7.95 | 0.59 | 16.81 |
| | South | 315 | 14.98 | 0.40 | 26.96 |
| | All | 2625 | 10.46 | 1.66 | 21.89 |
| *Retinopathy* | 25–34 | 405 | 6.06 | 0.61 | 8.70 |
| | 35–44 | 405 | 6.98 | 0.52 | 10.09 |
| | 45–54 | 405 | 12.31 | 0.63 | 18.04 |
| | 55–64 | 405 | 30.48 | 1.47 | 44.47 |
| | 65–74 | 405 | 47.14 | 1.95 | 73.01 |
| | 75–89 | 405 | 72.35 | 3.30 | 112.40 |
| | 90+ | 390 | 66.32 | 2.77 | 101.12 |
| | North | 720 | 25.98 | 5.10 | 68.37 |
| | Northeast | 945 | 32.03 | 0.39 | 55.29 |
| | Center-West | 420 | 43.58 | 0.82 | 76.15 |
| | Southeast | 420 | 31.87 | 0.17 | 52.22 |
| | South | 315 | 51.44 | 0.18 | 87.86 |
| | All | 2820 | 34.35 | 1.60 | 65.96 |
| *Eye & App. Dis.* | 25–34 | 405 | 32.00 | 0.55 | 46.60 |
| | 35–44 | 405 | 36.02 | 0.46 | 53.42 |
| | 45–54 | 405 | 52.23 | 0.43 | 80.65 |
| | 55–64 | 405 | 102.93 | 0.90 | 141.33 |
| | 65–74 | 405 | 306.81 | 2.27 | 472.32 |
| | 75–89 | 405 | 691.30 | 4.04 | 1119.44 |
| | 90+ | 405 | 483.46 | 5.85 | 766.00 |
| | North | 735 | 209.36 | 7.14 | 566.33 |
| | Northeast | 945 | 204.29 | 0.32 | 398.09 |
| | Center-West | 420 | 384.68 | 0.32 | 875.74 |
| | Southeast | 420 | 274.04 | 0.24 | 495.27 |
| | South | 315 | 212.16 | 0.28 | 345.71 |
| | All | 2835 | 243.54 | 2.07 | 547.00 |

Table 6: Subgroup performance for the selected model `nb5_age_cat` (Part 2: Bias and Coverage). All rates are expressed per million inhabitants.

| Disease | Subgroup | Bias | Mean obs. | Rel. bias (%) | $\mathrm{Cov}_{80}$ | $\mathrm{Cov}_{95}$ |
|---|---|---|---|---|---|---|
| *Glaucoma* | 25–34 | -0.09 | 3.3 | -2.8 | 82.9 | 92.8 |
| | 35–44 | -0.02 | 6.0 | -0.3 | 90.8 | 97.7 |
| | 45–54 | 0.19 | 15.8 | 1.2 | 92.3 | 97.9 |
| | 55–64 | 0.45 | 42.1 | 1.1 | 93.6 | 98.7 |
| | 65–74 | -0.61 | 101.0 | -0.6 | 94.8 | 100.0 |
| | 75–89 | -1.14 | 148.2 | -0.8 | 93.9 | 98.4 |
| | 90+ | 1.72 | 93.0 | 1.8 | 88.4 | 98.0 |
| | North | 0.65 | 10.3 | 6.3 | 95.6 | 99.1 |
| | Northeast | 0.08 | 30.9 | 0.3 | 92.0 | 98.1 |
| | Center-West | -1.96 | 40.0 | -4.9 | 90.5 | 98.3 |
| | Southeast | -3.61 | 38.4 | -9.4 | 90.7 | 96.9 |
| | South | 6.42 | 37.8 | 17.0 | 81.9 | 94.3 |
| | All | 0.05 | 34.6 | 0.1 | 91.1 | 97.7 |
| *Retinopathy* | 25–34 | 0.69 | 26.6 | 2.6 | 74.6 | 90.9 |
| | 35–44 | 1.08 | 46.3 | 2.3 | 84.4 | 96.3 |
| | 45–54 | 2.11 | 105.7 | 2.0 | 92.3 | 99.8 |
| | 55–64 | -0.34 | 281.3 | -0.1 | 93.8 | 98.8 |
| | 65–74 | -5.09 | 514.5 | -1.0 | 94.6 | 99.3 |
| | 75–89 | -8.43 | 561.3 | -1.5 | 88.1 | 98.5 |
| | 90+ | -1.89 | 290.7 | -0.7 | 76.7 | 94.6 |
| | North | -7.43 | 88.2 | -8.4 | 87.9 | 95.3 |
| | Northeast | 0.80 | 147.7 | 0.5 | 87.3 | 98.0 |
| | Center-West | -1.66 | 210.0 | -0.8 | 87.9 | 98.1 |
| | Southeast | 1.81 | 182.2 | 1.0 | 84.0 | 96.4 |
| | South | -0.76 | 257.3 | -0.3 | 81.6 | 96.2 |
| | All | -1.69 | 179.0 | -0.9 | 86.4 | 96.9 |
| *Eye & App. Dis.* | 25–34 | 9.59 | 119.1 | 8.1 | 77.8 | 92.3 |
| | 35–44 | 13.31 | 156.1 | 8.5 | 82.0 | 96.0 |
| | 45–54 | 17.91 | 300.0 | 6.0 | 93.6 | 99.3 |
| | 55–64 | 19.99 | 777.1 | 2.6 | 96.3 | 99.3 |
| | 65–74 | -82.91 | 2109.3 | -3.9 | 87.9 | 97.5 |
| | 75–89 | -244.22 | 3507.0 | -7.0 | 78.8 | 95.3 |
| | 90+ | -121.08 | 2355.5 | -5.1 | 79.0 | 94.3 |
| | North | -90.07 | 290.9 | -31.0 | 78.5 | 91.8 |
| | Northeast | -33.68 | 574.7 | -5.9 | 87.4 | 98.0 |
| | Center-West | -115.31 | 768.7 | -15.0 | 85.7 | 96.9 |
| | Southeast | -120.43 | 944.0 | -12.8 | 90.2 | 98.3 |
| | South | 127.40 | 616.8 | 20.7 | 85.4 | 98.1 |
| | All | -55.34 | 734.5 | -7.5 | 85.0 | 96.3 |

### 6 Posterior Predictive Checks

Posterior predictive checks (PPCs) assess goodness-of-fit by comparing summary statistics of the observed data with the distribution of those statistics under replicated datasets simulated from the fitted model. For each disease, we drew posterior predictive samples

$$y_n^{\text{rep}} \mid \alpha^{(s)}, \beta_{\text{age}}^{(s)}, \phi^{(s)} \sim \text{NegBin}_2\Big(\exp\big(\log P_n + \alpha_{s(n),\,t(n)}^{(s)} + \beta_{\text{age},\,j(n)}^{(s)}\big),\, \phi^{(s)}\Big)\,,$$

where $(s)$ indexes posterior draws of the selected `nb5_age_cat` model, $n$ indexes the $N$ observed state-year-age cells, and $\{s(n), t(n), j(n)\}$ are the state, year and age-stratum indices of cell $n$. We computed five test statistics on each replicate dataset and compared to the corresponding statistic on the observed data: the marginal density of counts (*density*), the sample mean (*mean*), the sample standard deviation (*sd*), the sample maximum (*max*), and the proportion of zero-count cells (*zeros*).

In each figure below the posterior predictive distribution is shown as overlapping light draws and the observed-data summary as a dark line or point. Adequate fit corresponds to the observed statistic falling within the bulk of the predictive distribution; systematic discrepancies indicate likelihood or structural misspecification. Two specific failure modes are worth flagging in advance: an observed sample standard deviation outside the predictive bulk would suggest the Negative Binomial likelihood underestimates dispersion (or that residual structure is absorbed by $\phi$); an observed proportion of zeros above the predictive upper bound would suggest the data exhibit zero-inflation beyond what a continuous-mean Negative Binomial can produce. The checks are reported separately for the three diseases in the subsections that follow.

#### 6.1 Glaucoma

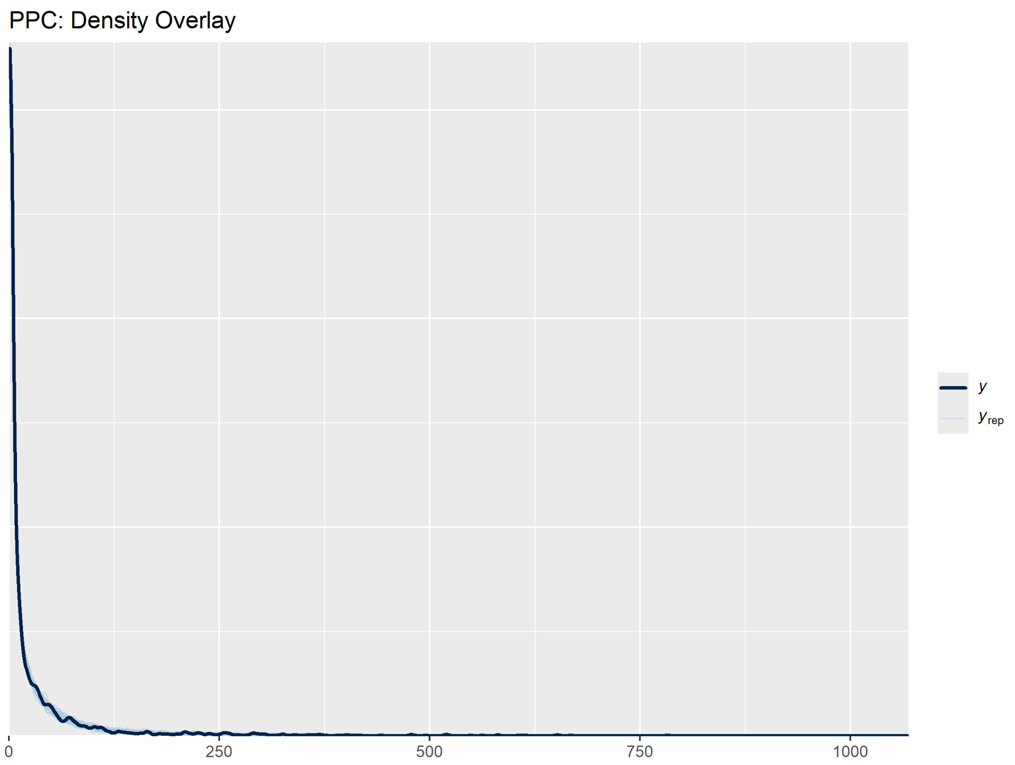

Figure 7: Posterior Predictive Check - Density - Glaucoma.

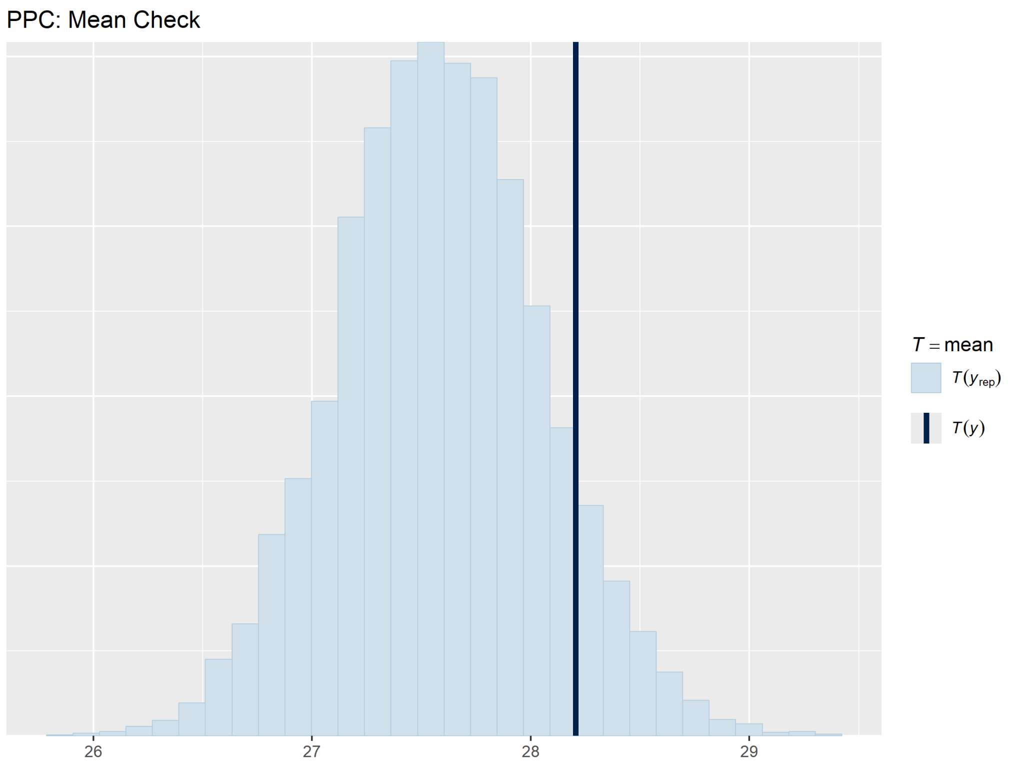

Figure 8: Posterior Predictive Check - Mean - Glaucoma.

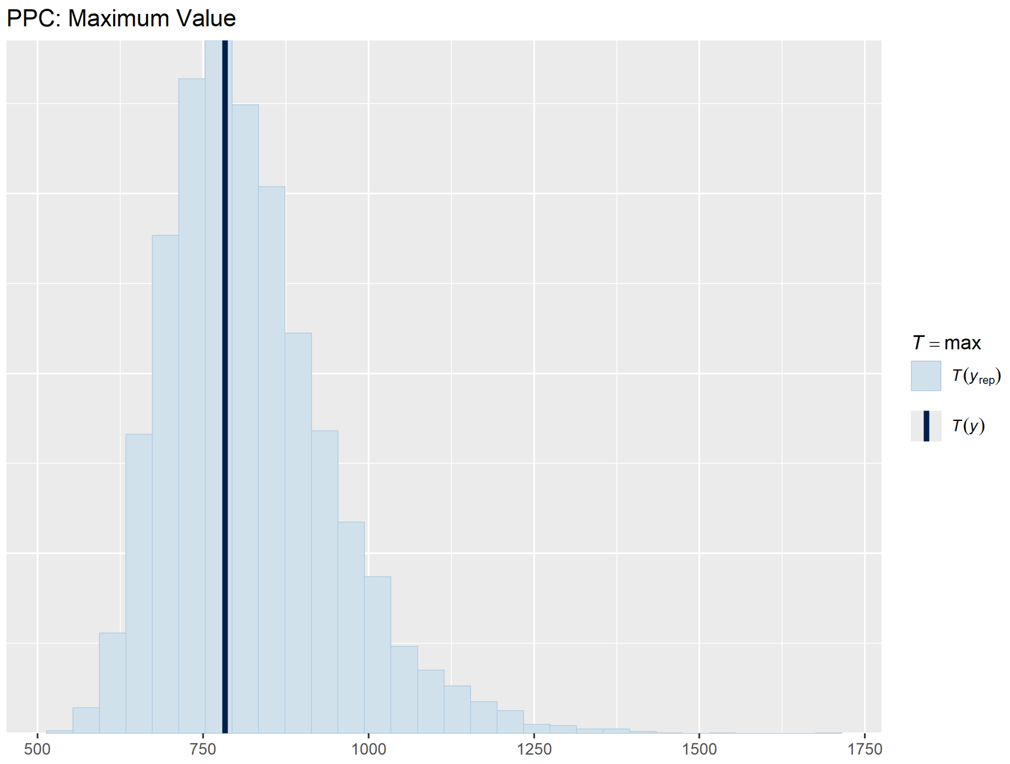

Figure 9: Posterior Predictive Check - Sample Maximum - Glaucoma.

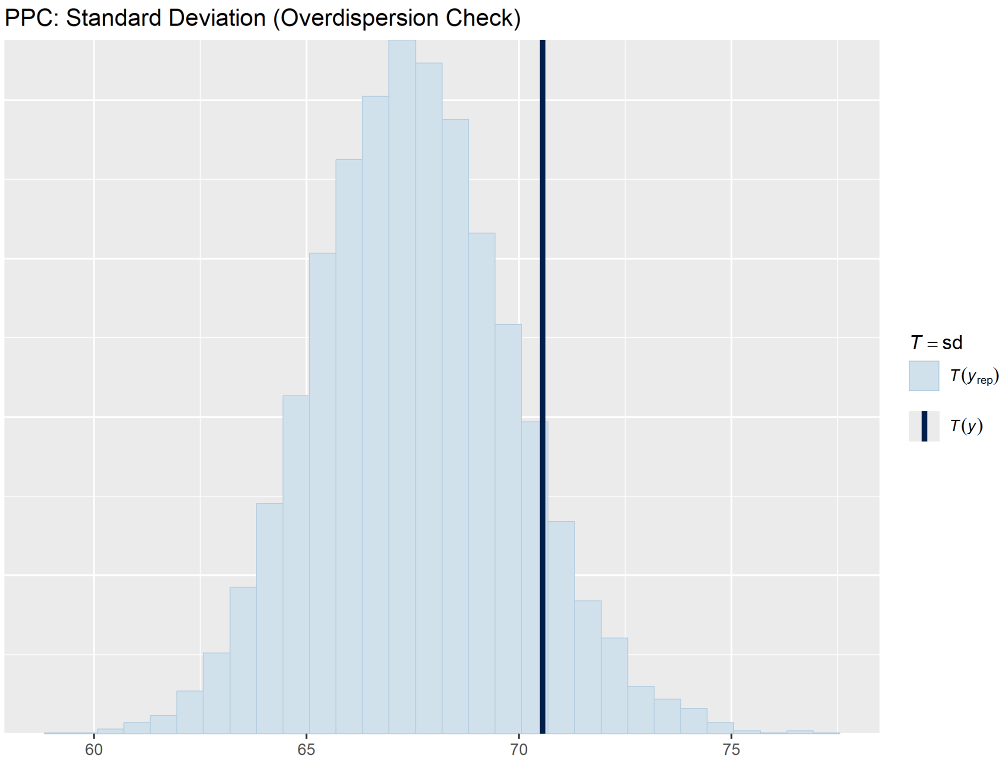

Figure 10: Posterior Predictive Check - Standard Deviation - Glaucoma.

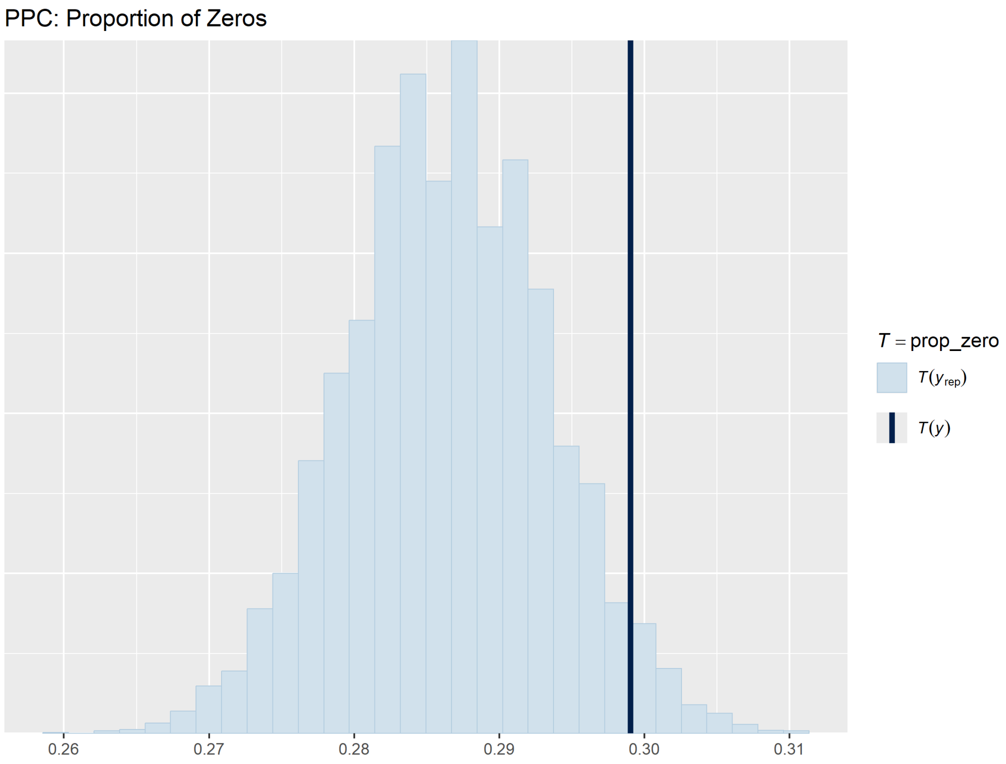

Figure 11: Posterior Predictive Check - Zeros - Glaucoma.

#### 6.2 Retinopathy

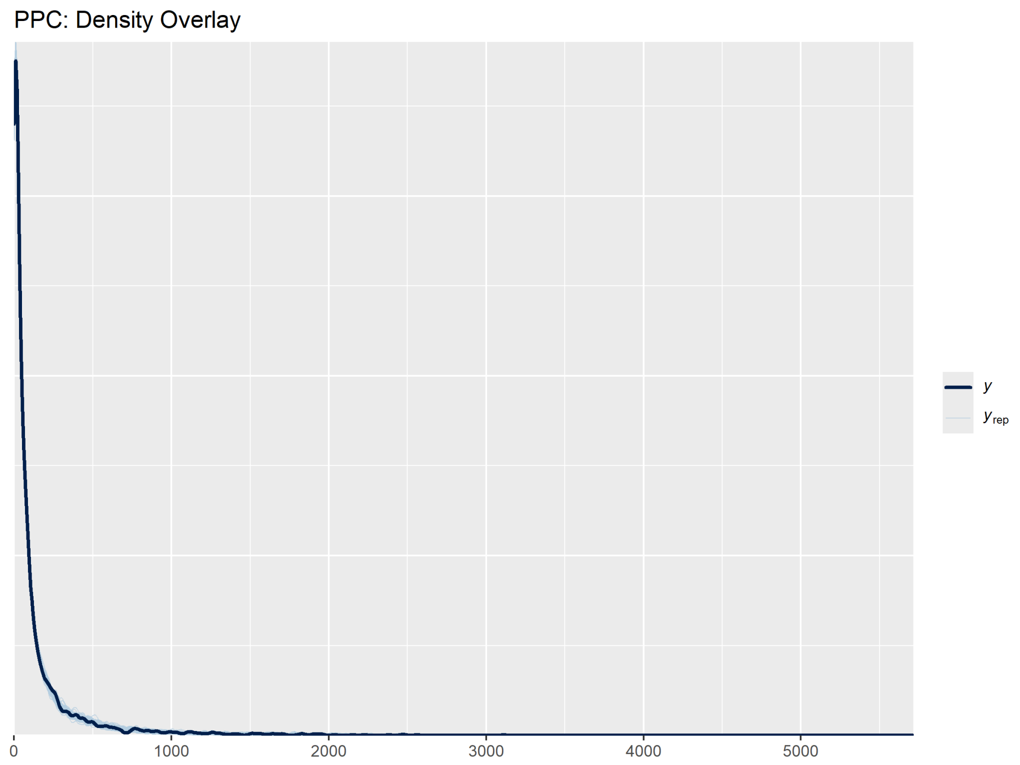

Figure 12: Posterior Predictive Check - Density - Retinopathy.

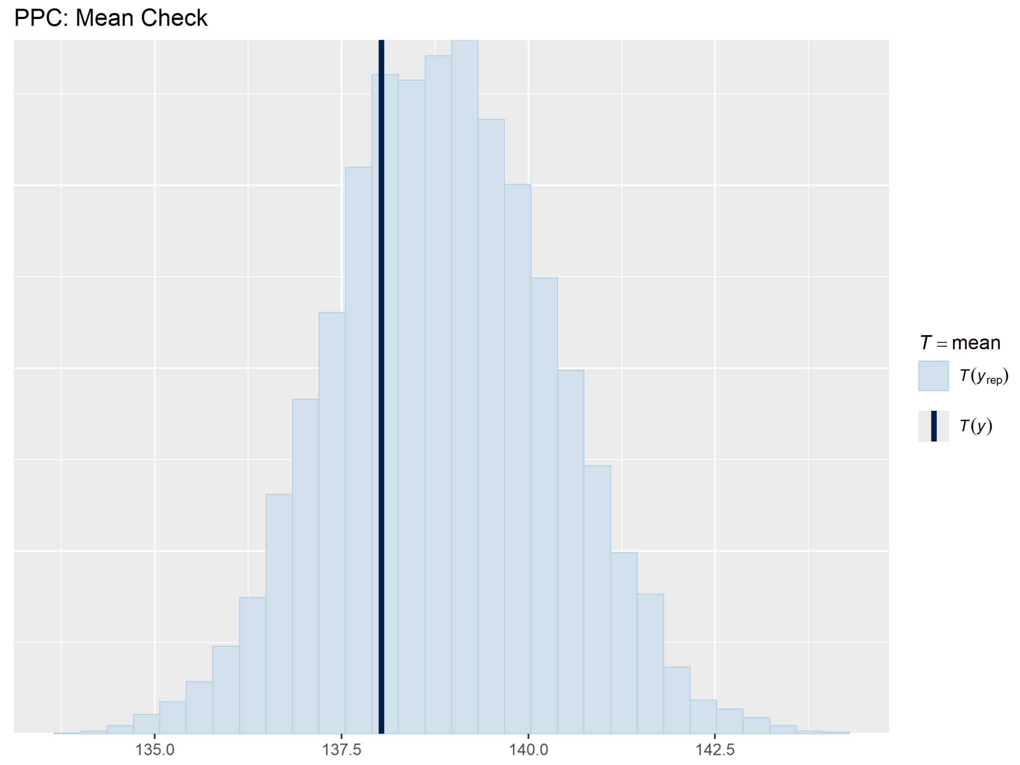

Figure 13: Posterior Predictive Check - Mean - Retinopathy.

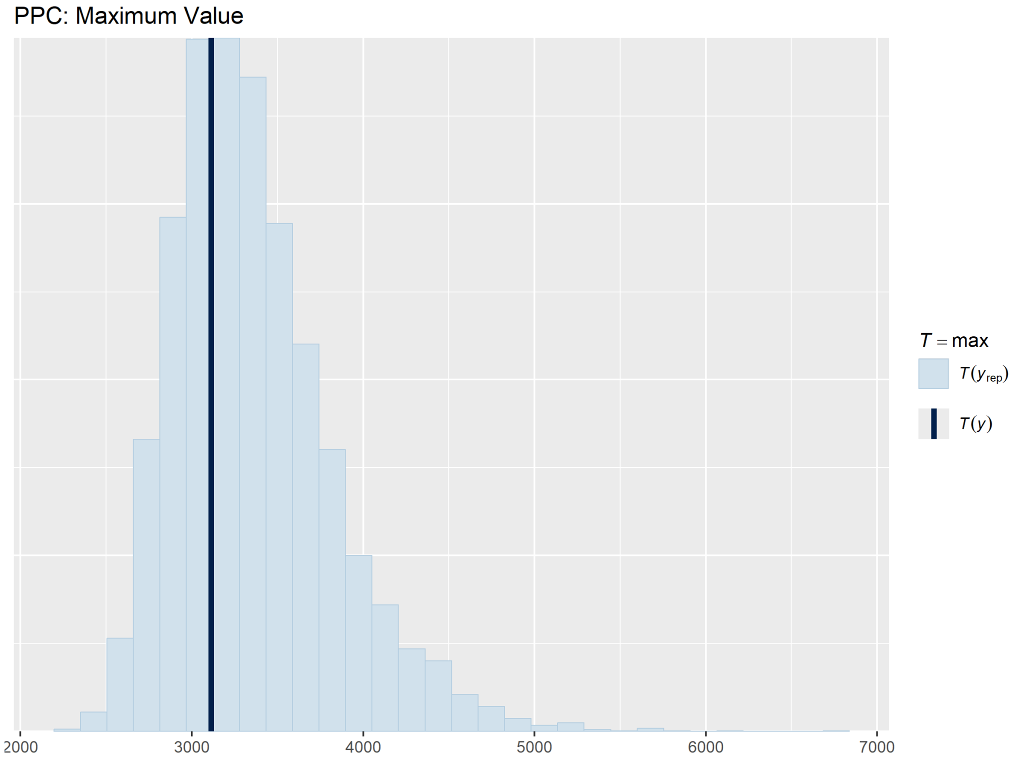

Figure 14: Posterior Predictive Check - Sample Maximum - Retinopathy.

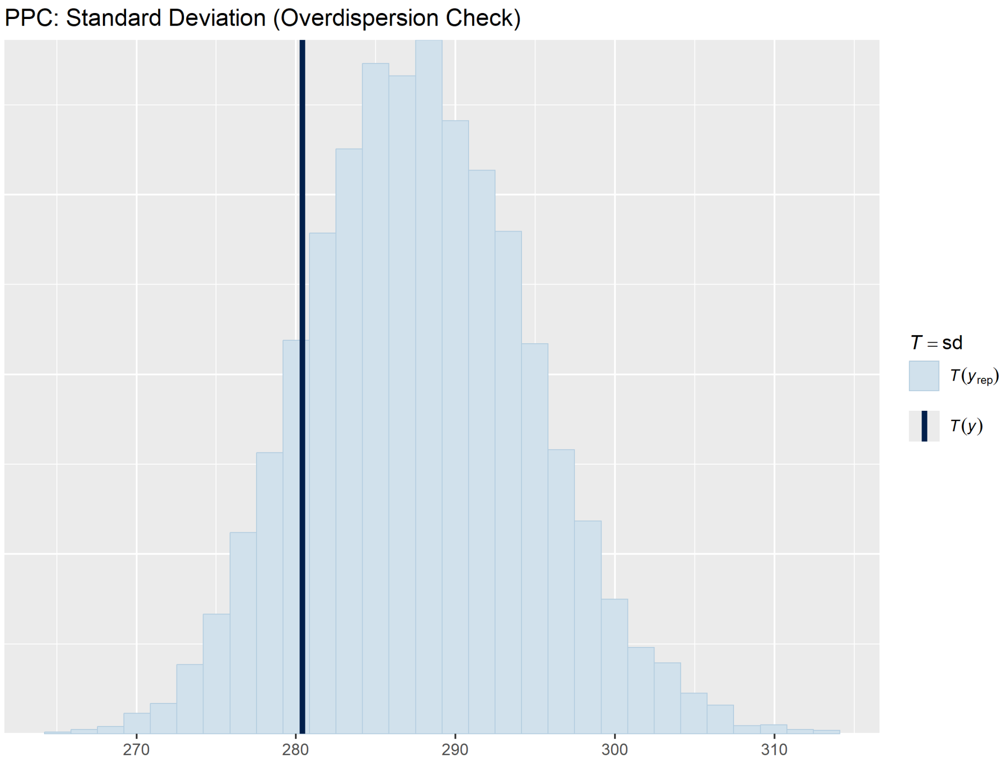

Figure 15: Posterior Predictive Check - Standard Deviation - Retinopathy.

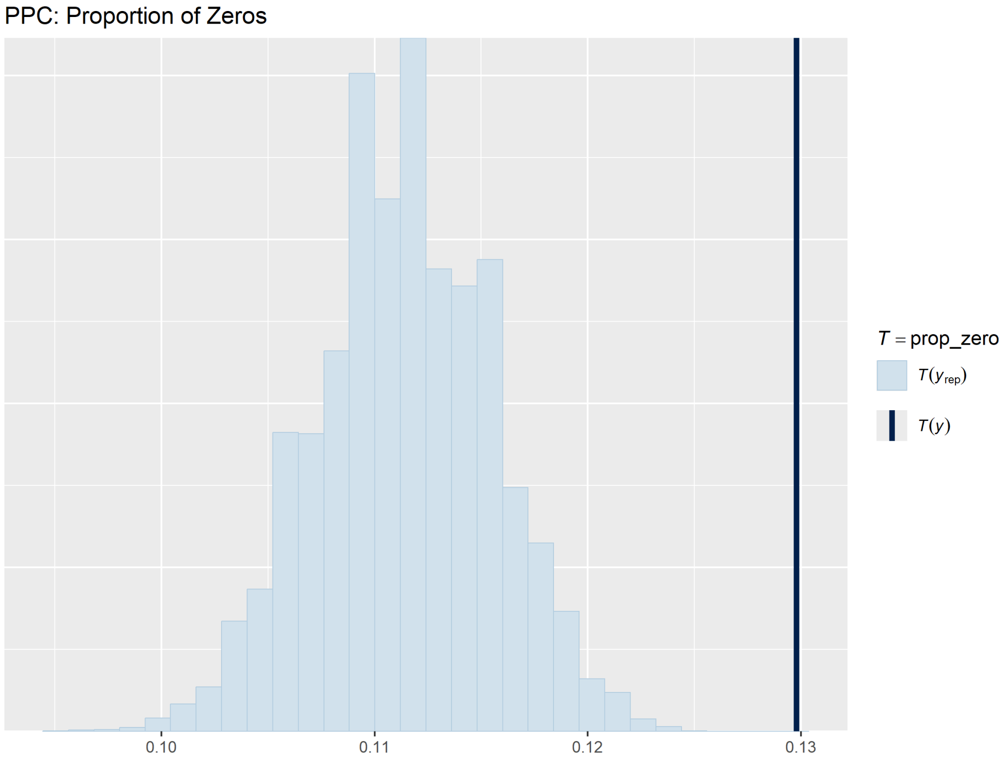

Figure 16: Posterior Predictive Check - Zeros - Retinopathy.

#### 6.3 Eye & Appendage Diseases

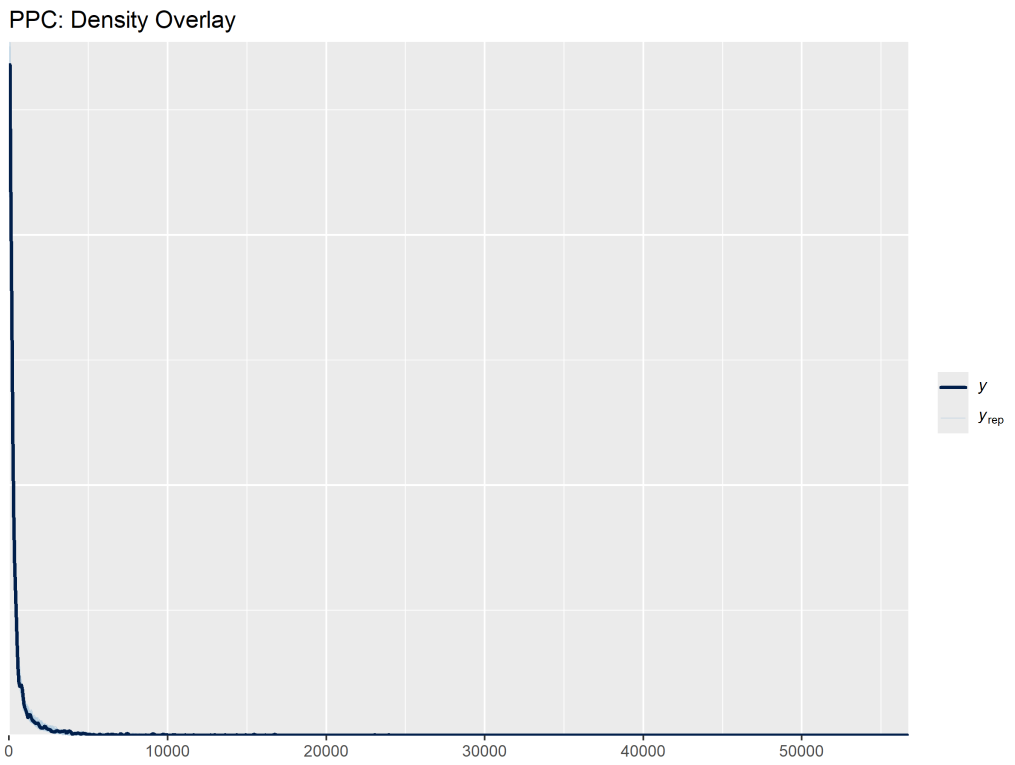

Figure 17: Posterior Predictive Check - Density - Eye & Appendage Diseases.

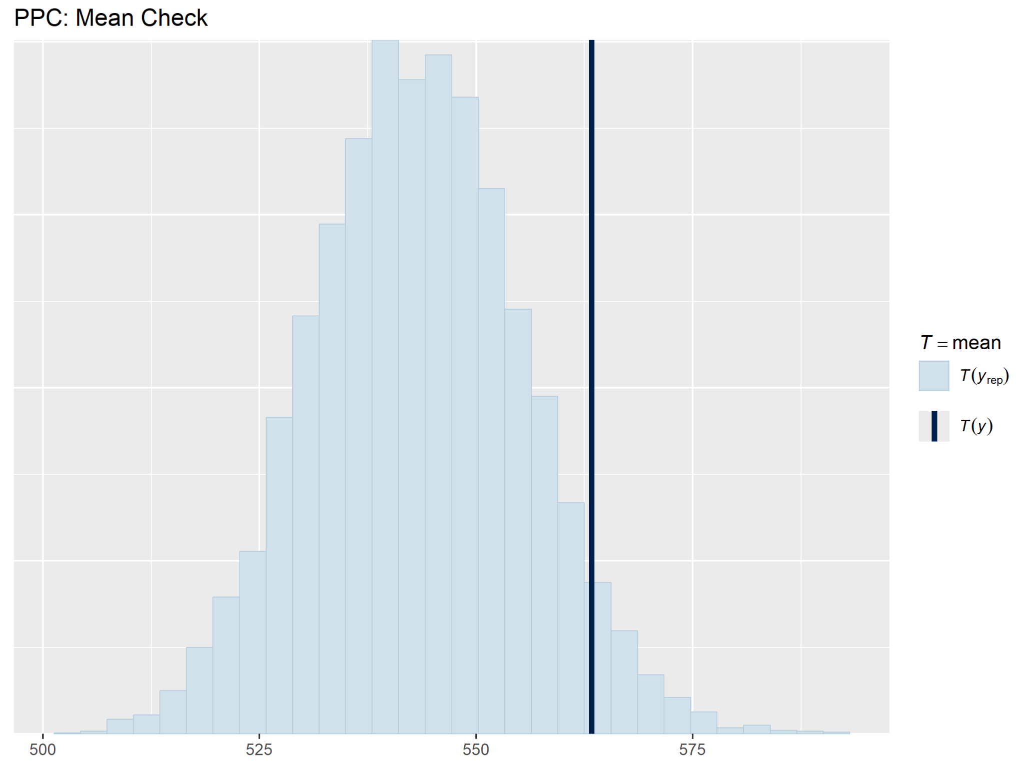

Figure 18: Posterior Predictive Check - Mean - Eye & Appendage Diseases.

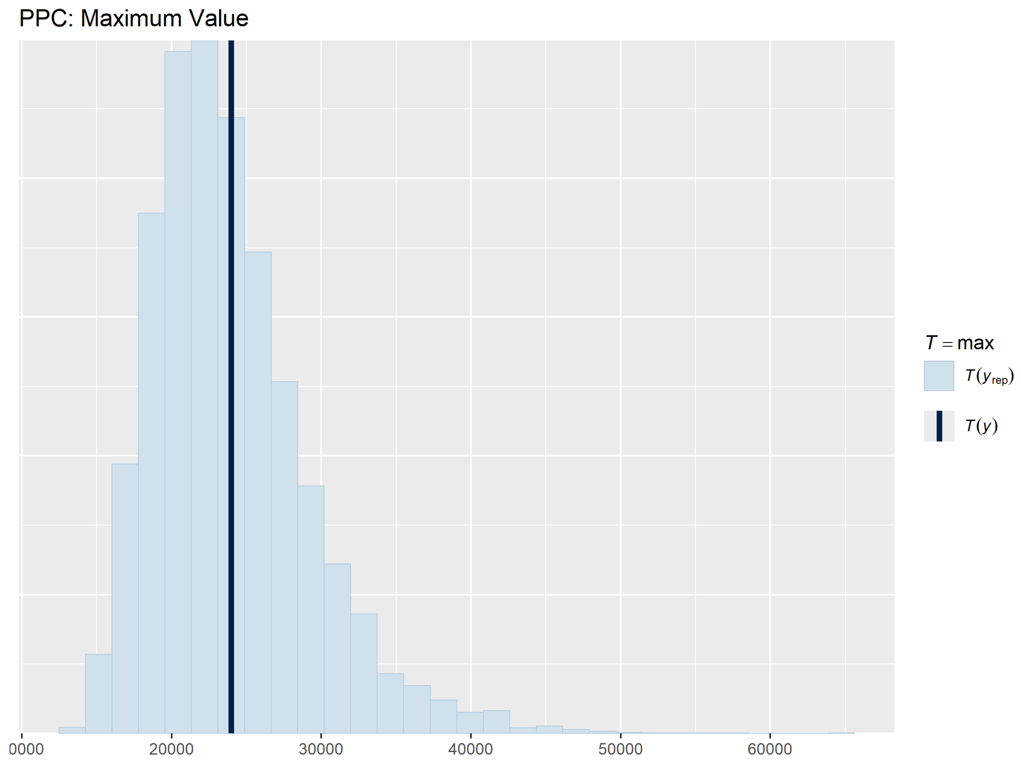

Figure 19: Posterior Predictive Check - Sample Maximum - Eye & Appendage Diseases.

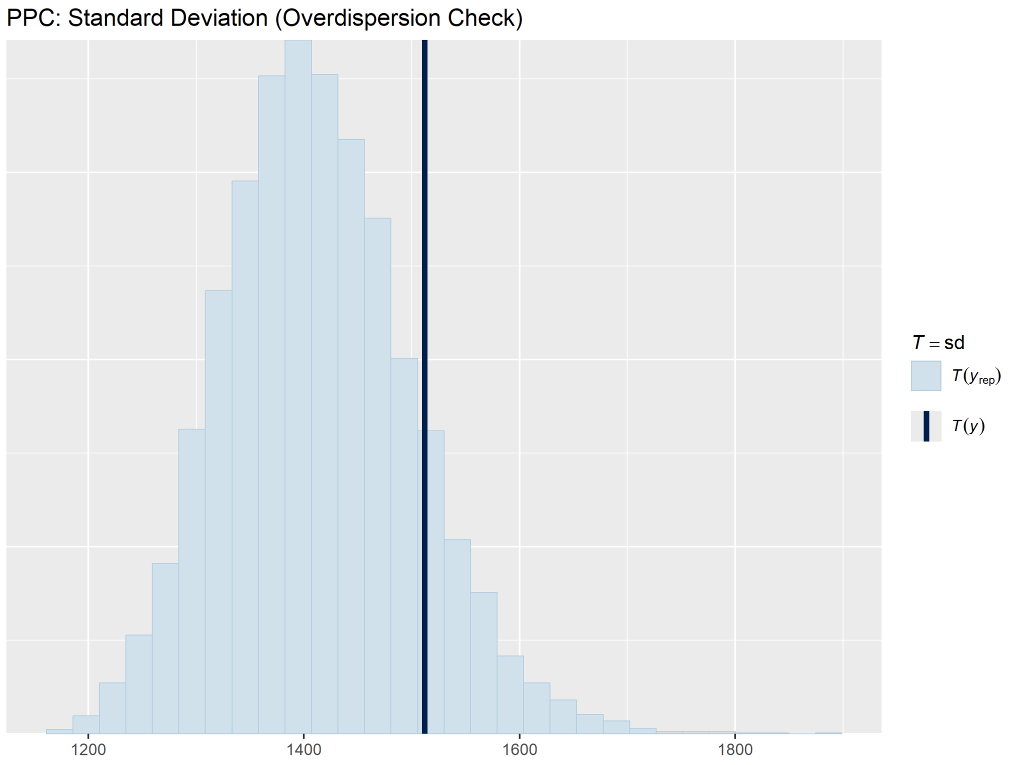

Figure 20: Posterior Predictive Check - Standard Deviation - Eye & Appendage Diseases.

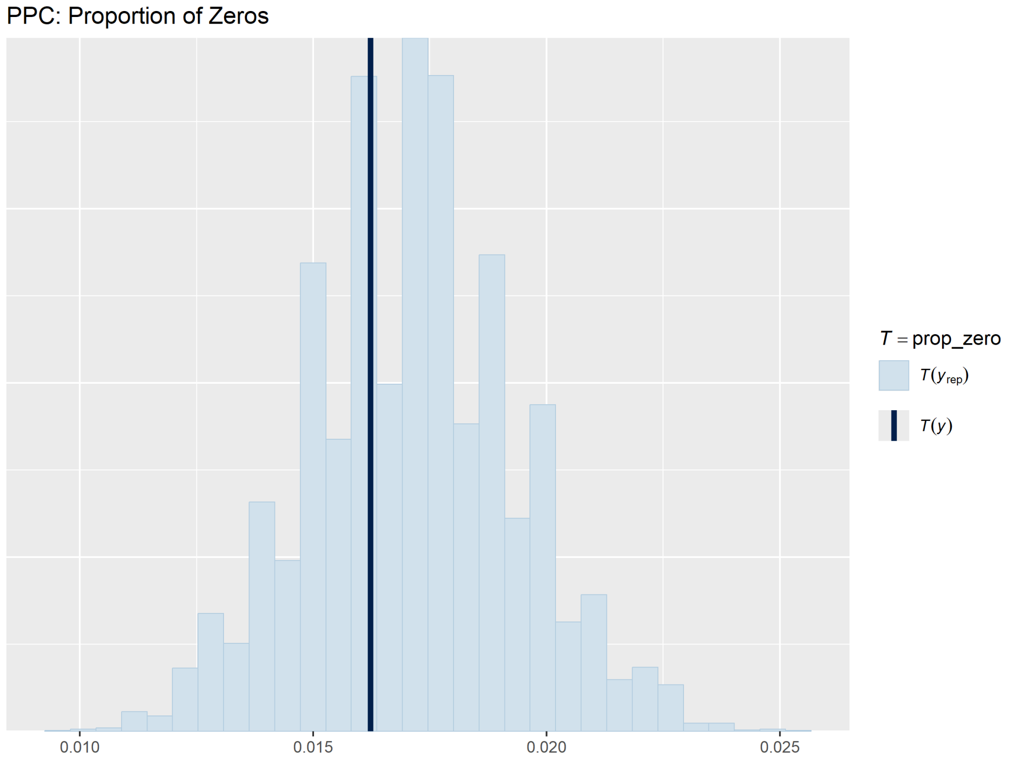

Figure 21: Posterior Predictive Check - Zeros - Eye & Appendage Diseases.

### 7 Sensitivity Analysis

We assessed robustness of the selected `nb5_age_cat` model along five $\phi$-prior variants (Figs. 22-24 and Table 7). Two additional data-scenario tests - excluding 2024 from the training window, and treating cells with missing counts as zero - were also run on the simpler `nb5` structure as a coarse-grained robustness check; results are reported in Table 8.

Across the five $\phi$ priors ($\mathcal{N}^+(0,10)$ baseline, $\mathcal{N}^+(0,3)$ tight, Gamma$(2,0.1)$, Exp$(0.1)$, and half-Cauchy(0, 5)) inference on $\phi$ is robust for eye-and-appendage diseases (posterior mean within $\pm1\%$ across all priors), but materially sensitive under the tighter $\mathcal{N}^+(0,3)$ prior for glaucoma ($\phi$ drops from 17.53 to 15.09, a 14% reduction) and for retinopathy ($\phi$ drops from 31.04 to 25.85, a 17% reduction); the four remaining priors agree to within 2% of the baseline for all three diseases. The tighter half-normal prior places substantial mass below the data-supported $\phi$ range and effectively regularizes the overdispersion estimate downward; we therefore retain the weakly informative $\mathcal{N}^+(0,10)$ baseline, which agrees with three of the four alternative priors and is data-dominated. Excluding 2024 from the training data leaves $\phi$, $\beta_{\text{age,mean}}$ and $\sigma_{\text{rw,mean}}$ unchanged to within Monte Carlo error while reducing LOO-IC proportionally to the loss of one year of data, confirming that 2024 reporting did not unduly influence the posterior. Treating missing cells as zero changes the parameters by less than 0.5% across all three diseases and slightly worsens LOO-IC, supporting the decision to treat missing as missing in the main analysis.

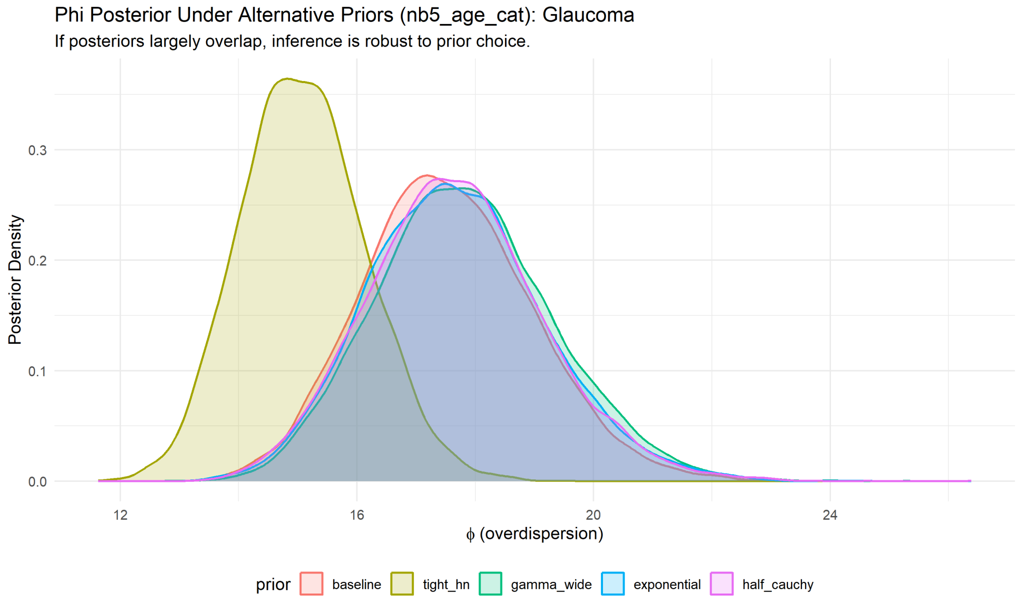

Figure 22: Posterior of $\phi$ under five alternative priors - Glaucoma, selected model `nb5_age_cat`.

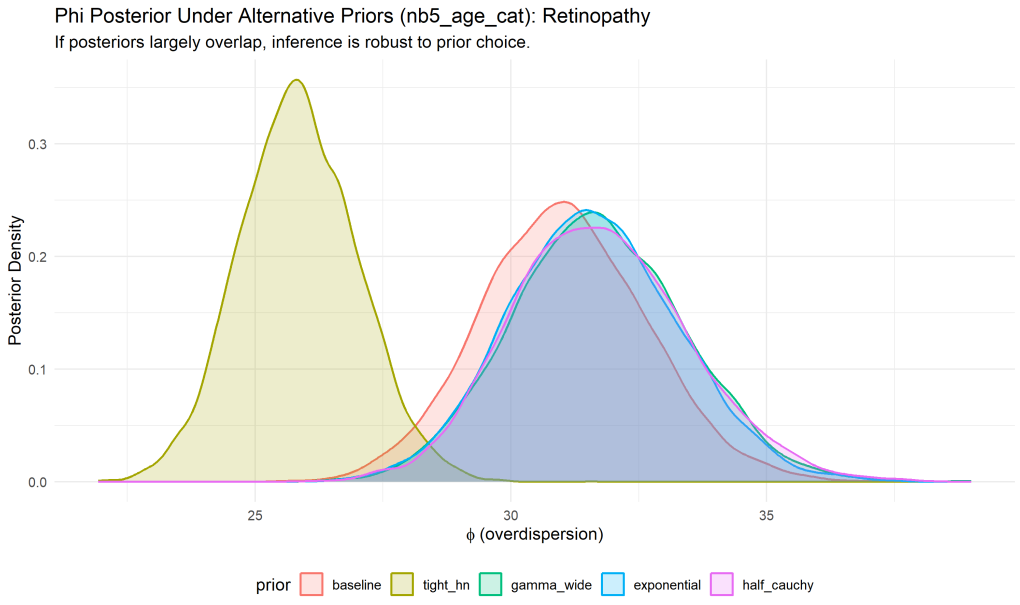

Figure 23: Posterior of $\phi$ under five alternative priors - Retinopathy, selected model `nb5_age_cat`.

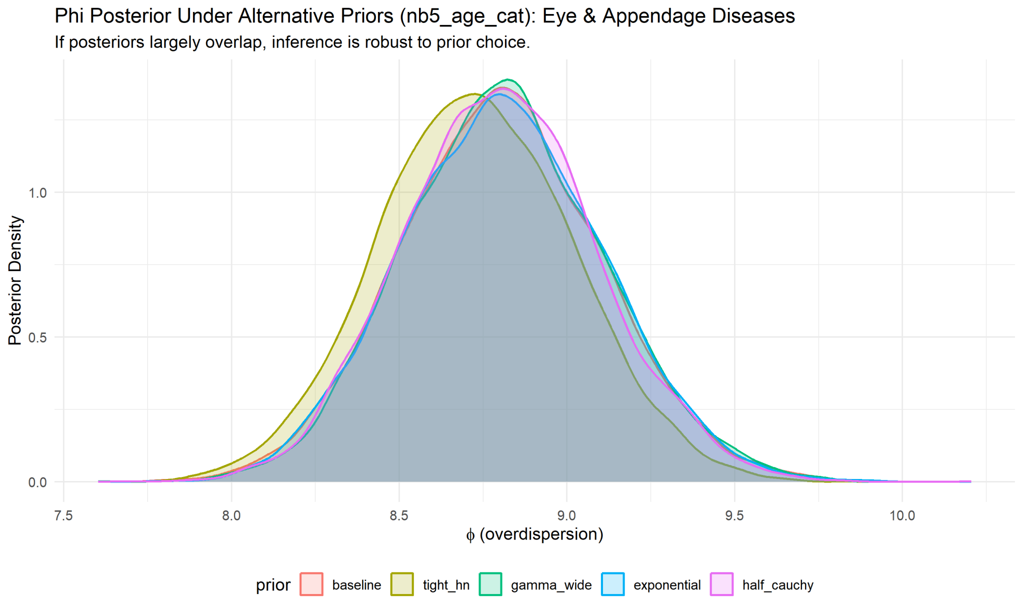

Figure 24: Posterior of $\phi$ under five alternative priors - Eye and Appendage Diseases, selected model `nb5_age_cat`.

Table 7: Posterior of $\phi$ under five alternative priors, for the selected model `nb5_age_cat`. The tight half-normal prior materially shifts $\phi$ for glaucoma and retinopathy but not for eye-and-appendage diseases; the four other priors agree to within ~2% of the baseline on all diseases.

| **Disease** | **Prior on $\phi$** | $\phi_{\text{mean}}$ | $\phi_{\text{q025}}$ | $\phi_{\text{q975}}$ | **Max $\hat{R}$** | **Min $N_{\text{eff}}$** |
|---|---|---|---|---|---|---|
| Glaucoma | $\mathcal{N}^+(0, 10)$ baseline | 17.534 | 14.901 | 20.518 | 1.0027 | 2126 |
| Glaucoma | $\mathcal{N}^+(0, 3)$ tight | 15.089 | 13.168 | 17.220 | 1.0026 | 2152 |
| Glaucoma | Gamma(2, 0.1) | 17.827 | 15.095 | 20.950 | 1.0028 | 2040 |
| Glaucoma | Exponential(0.1) | 17.680 | 14.986 | 20.795 | 1.0056 | 1406 |
| Glaucoma | half-Cauchy(0, 5) | 17.667 | 14.930 | 20.758 | 1.0019 | 2175 |
| Retinopathy | $\mathcal{N}^+(0, 10)$ baseline | 31.036 | 27.893 | 34.353 | 1.0039 | 2175 |
| Retinopathy | $\mathcal{N}^+(0, 3)$ tight | 25.849 | 23.645 | 28.158 | 1.0039 | 2390 |
| Retinopathy | Gamma(2, 0.1) | 31.713 | 28.491 | 35.084 | 1.0034 | 2433 |
| Retinopathy | Exponential(0.1) | 31.614 | 28.458 | 34.927 | 1.0035 | 2378 |
| Retinopathy | half-Cauchy(0, 5) | 31.733 | 28.527 | 35.331 | 1.0032 | 2335 |
| Eye & App. Dis. | $\mathcal{N}^+(0, 10)$ baseline | 8.803 | 8.222 | 9.407 | 1.0031 | 2313 |
| Eye & App. Dis. | $\mathcal{N}^+(0, 3)$ tight | 8.727 | 8.163 | 9.304 | 1.0030 | 2476 |
| Eye & App. Dis. | Gamma(2, 0.1) | 8.813 | 8.256 | 9.416 | 1.0026 | 2297 |
| Eye & App. Dis. | Exponential(0.1) | 8.809 | 8.234 | 9.408 | 1.0036 | 2334 |
| Eye & App. Dis. | half-Cauchy(0, 5) | 8.800 | 8.253 | 9.382 | 1.0039 | 2452 |

Table 8: Data-scenario sensitivity on the simpler `nb5` base model: excluding 2024 from training, and treating missing cell counts as zero. Posterior means of the key parameters are reported. Both scenarios change the parameters by less than 2% relative to the baseline, supporting the analytic choices in the main paper.

| **Disease** | **Scenario** | $\phi_{\text{mean}}$ | $\beta_{\text{age,mean}}$ | $\sigma_{\text{rw,mean}}$ | **LOOIC** | **Max $\hat{R}$** | **Min $N_{\text{eff}}$** |
|---|---|---|---|---|---|---|---|
| Glaucoma | Baseline | 2.736 | 0.5931 | 0.4157 | 14080.1 | 1.0029 | 2564 |
| Retinopathy | Baseline | 2.700 | 0.4945 | 0.4199 | 24942.6 | 1.0029 | 2035 |
| Eye & App. | Baseline | 3.477 | 0.5406 | 0.3289 | 32482.9 | 1.0026 | 2664 |
| Glaucoma | Exclude 2024 | 2.769 | 0.5917 | 0.4328 | 12996.2 | 1.0024 | 2271 |
| Retinopathy | Exclude 2024 | 2.746 | 0.4900 | 0.4047 | 23000.8 | 1.0021 | 2265 |
| Eye & App. | Exclude 2024 | 3.501 | 0.5400 | 0.3201 | 30099.4 | 1.0027 | 3128 |
| Glaucoma | Missing as zero | 2.719 | 0.5883 | 0.4173 | 14153.8 | 1.0029 | 2427 |
| Retinopathy | Missing as zero | 2.700 | 0.4938 | 0.4215 | 24946.6 | 1.0022 | 2364 |
| Eye & App. | Missing as zero | 3.474 | 0.5407 | 0.3291 | 32485.0 | 1.0028 | 2665 |

### 8 Posterior Effects of the Selected Model

This section reports posterior summaries for the key parameters of the selected `nb5_age_cat` model: overdispersion ($\phi$), random-walk standard deviation ($\sigma_{\alpha,\text{rw}}$), categorical age effects ($\beta_{\text{age},j}$), and final-year state-level intercepts ($\alpha_{s,2024}$).

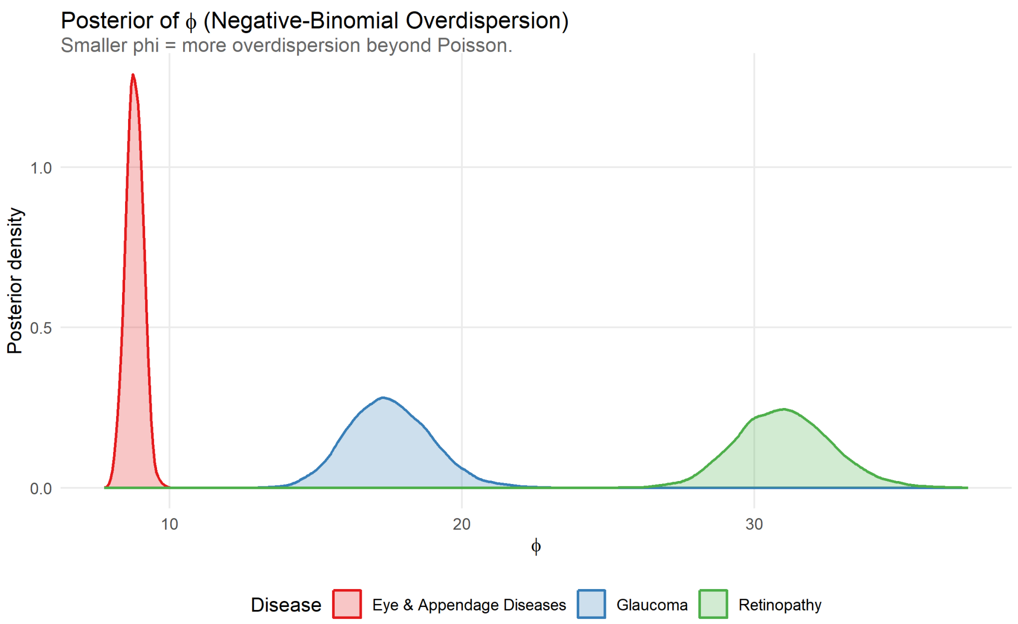

Figure 25: Phi Posterior per Disease.

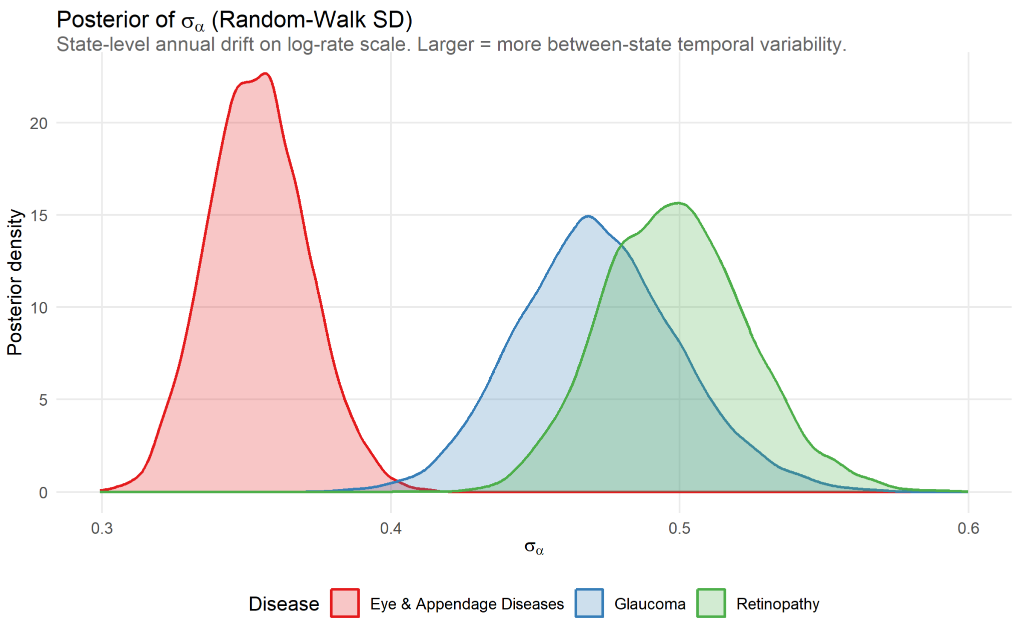

Figure 26: Sigma Posterior per Disease.

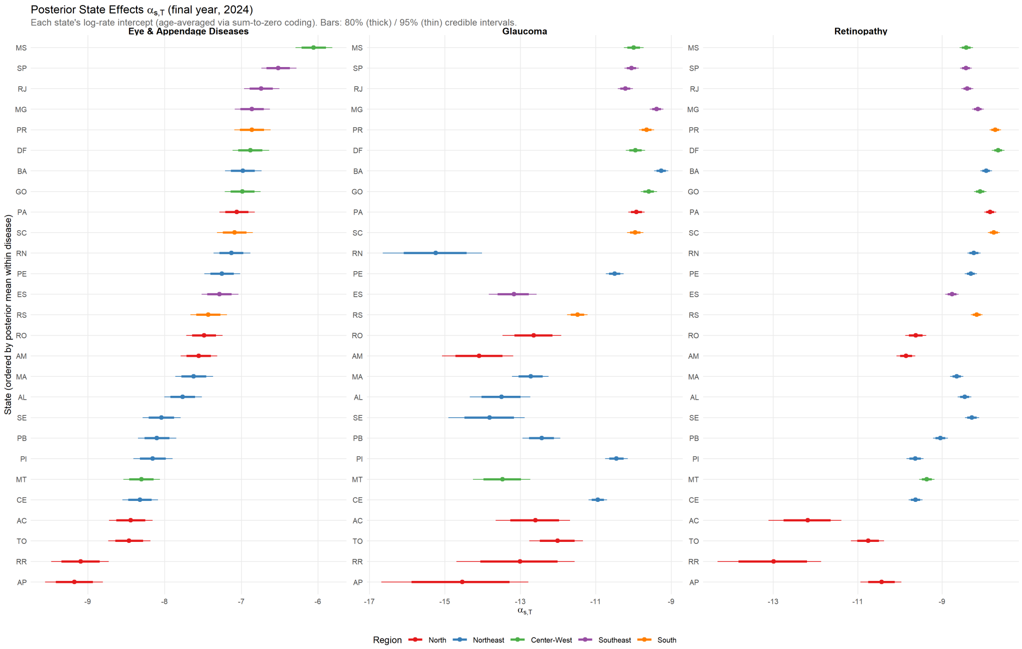

Figure 27: Posterior State Effects for 2024 per Disease.

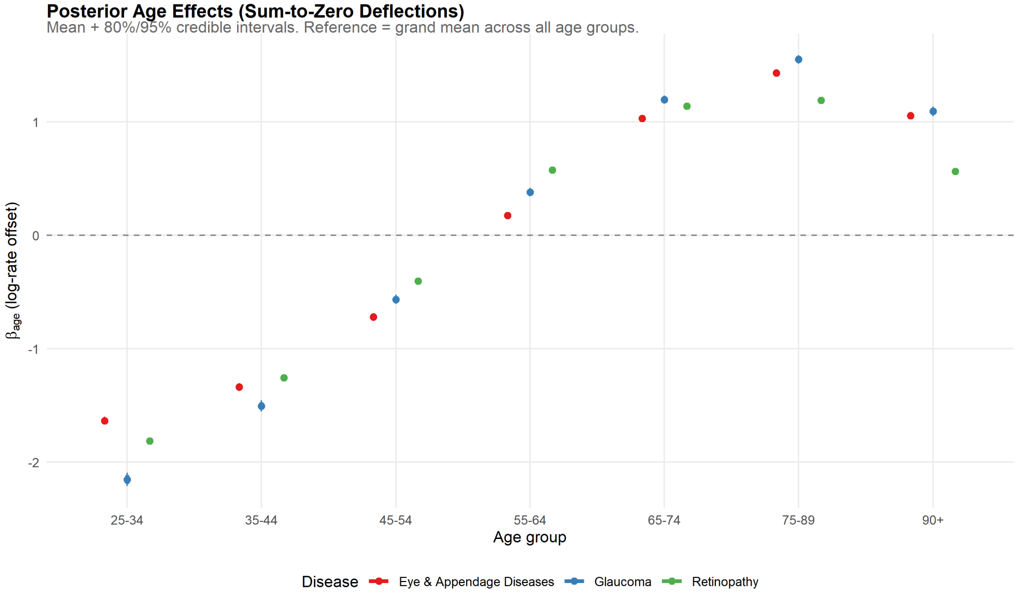

Figure 28: Posterior Age Effects per Disease.

#### 8.1 State-Year Trajectories and Regional Aggregates

We present the population-weighted regional aggregate of the state-year intercepts $\alpha_{s,t}$ (Fig. 29), followed by per-state temporal trajectories for each of the 27 federative units. Each per-state figure shows the posterior mean and 95% credible interval of $\alpha_{s,t}$ across the 2010-2024 fitting window for all three diseases.

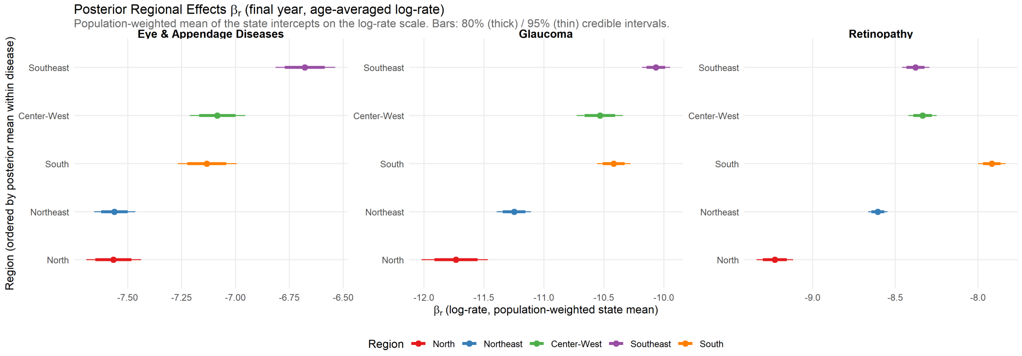

Figure 29: Region Posterior per Disease.

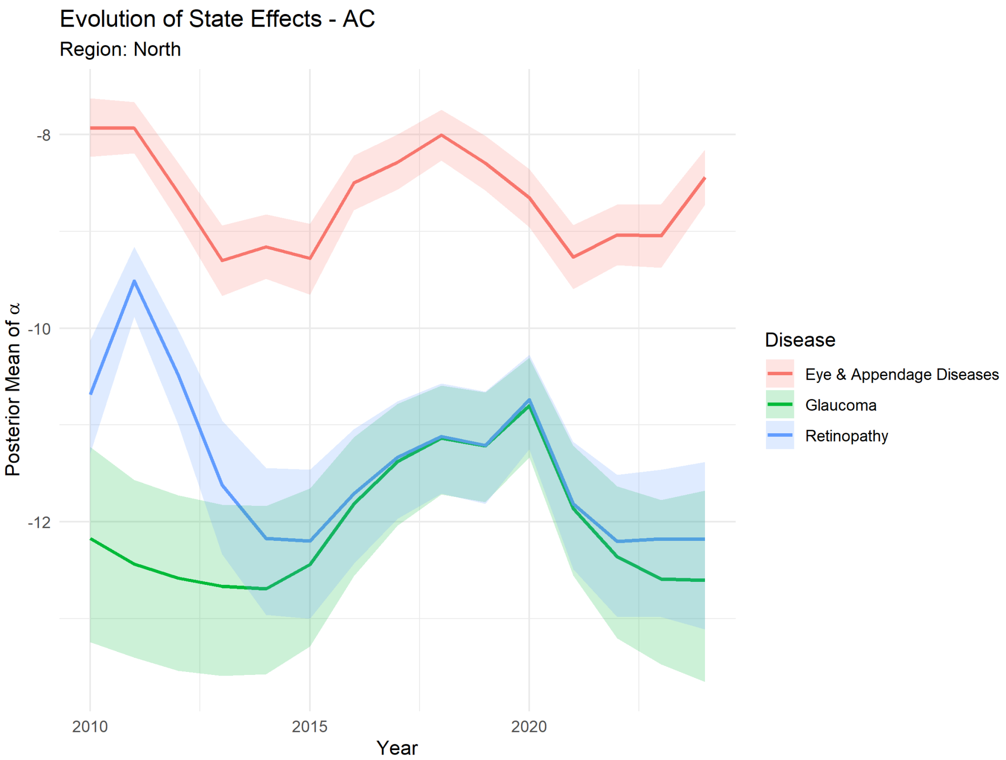

Figure 30: Acre Posterior per Disease.

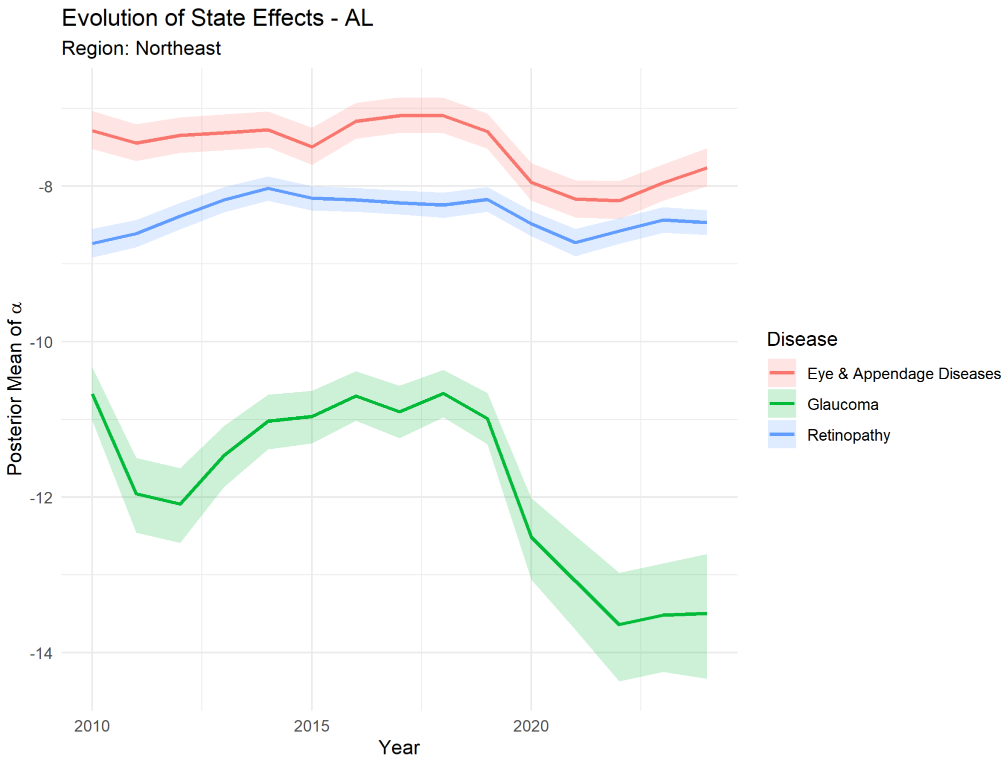

Figure 31: Alagoas Posterior per Disease.

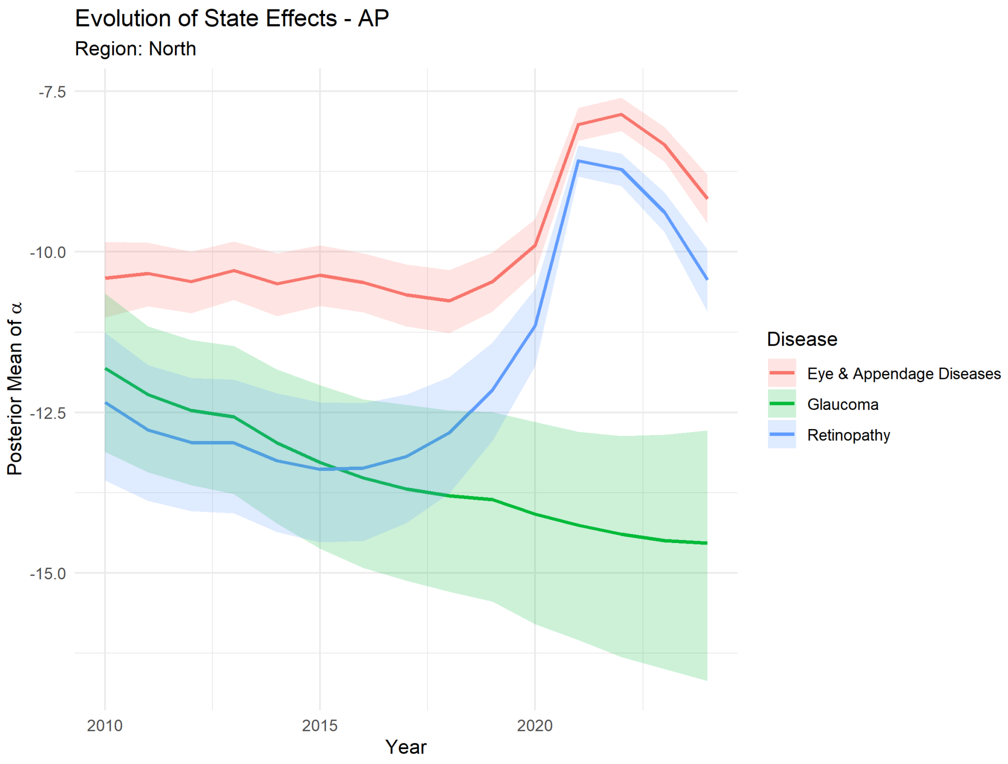

Figure 32: Amapá Posterior per Disease.

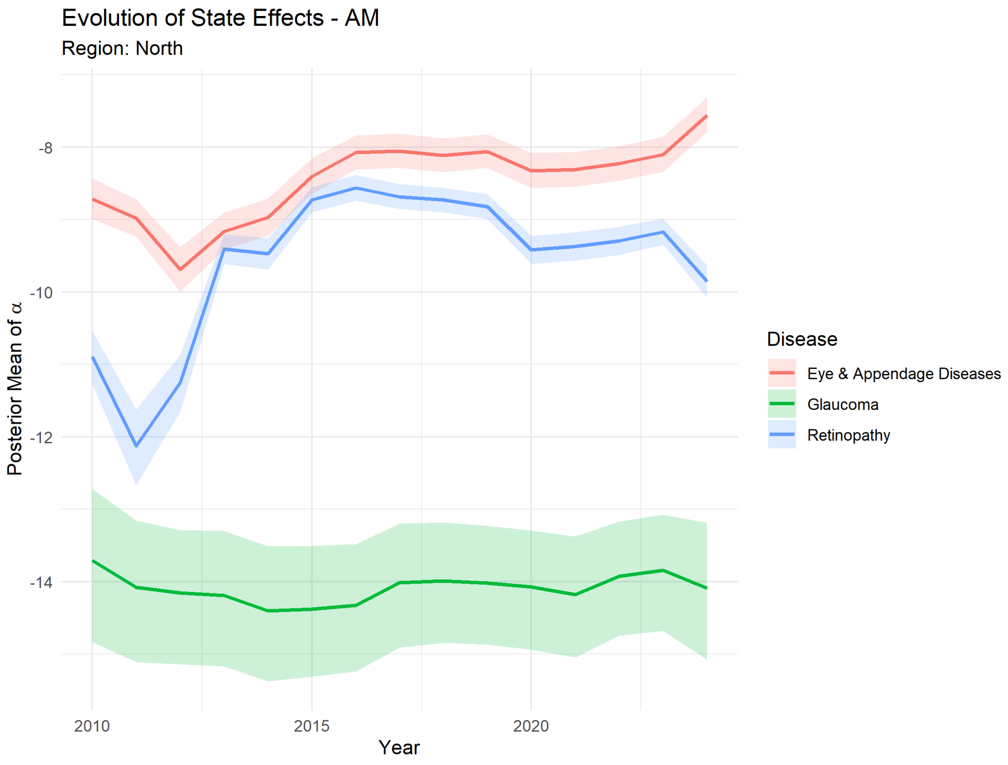

Figure 33: Amazonas Posterior per Disease.

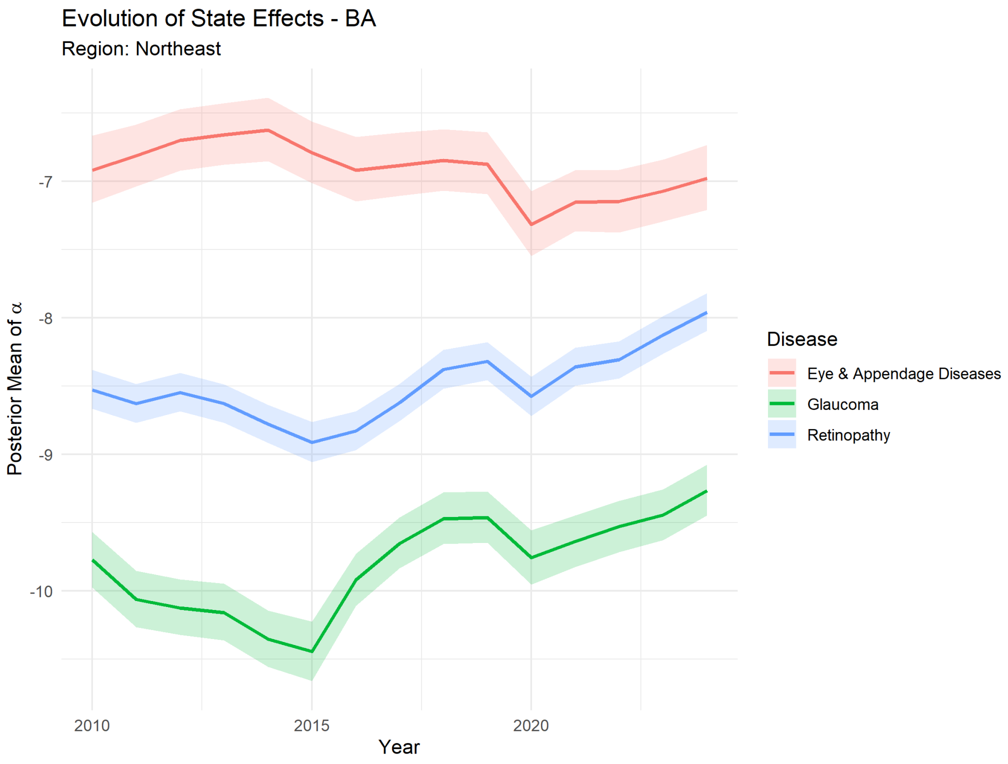

Figure 34: Bahia Posterior per Disease.

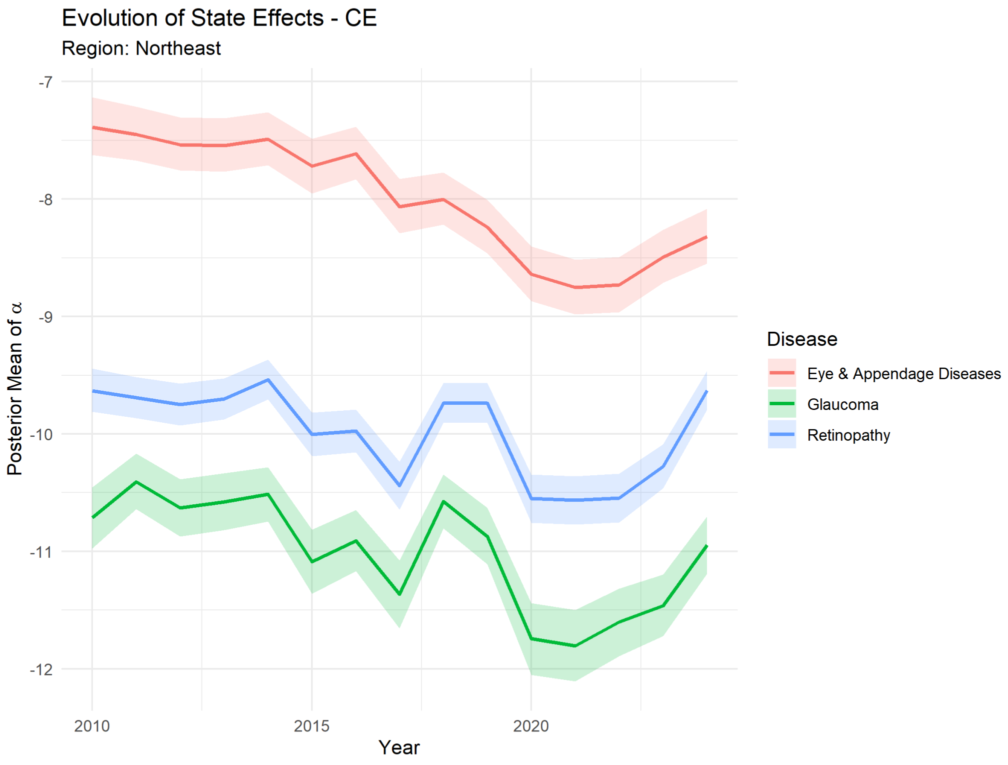

Figure 35: Ceará Posterior per Disease.

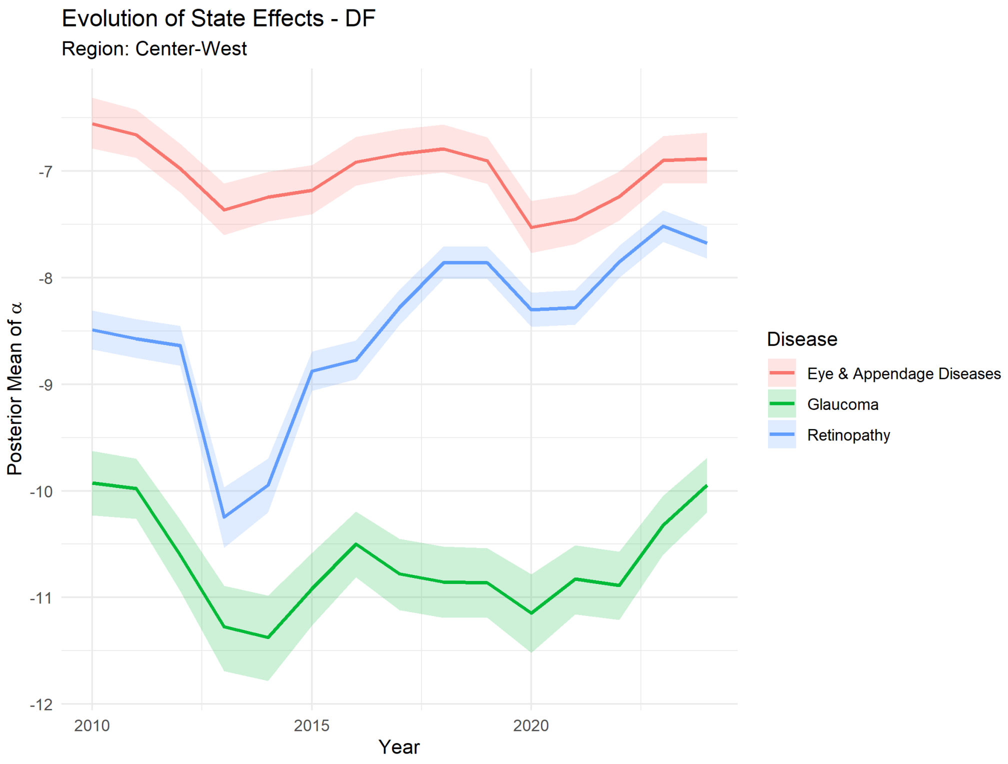

Figure 36: Distrito Federal Posterior per Disease.

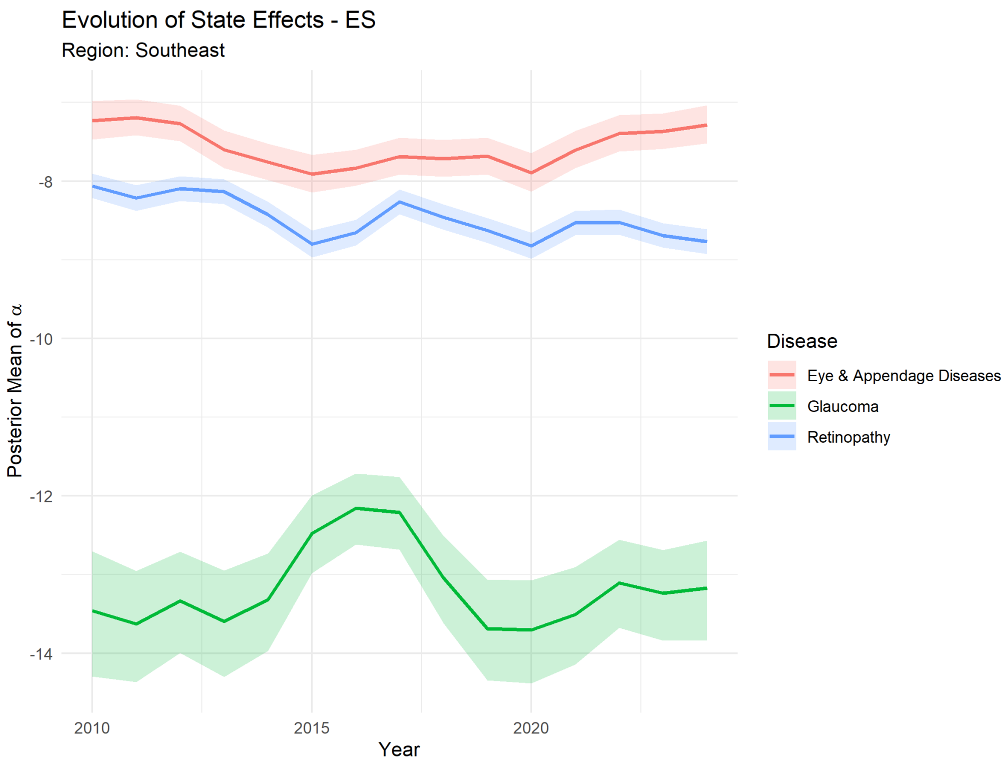

Figure 37: Espírito Santo Posterior per Disease.

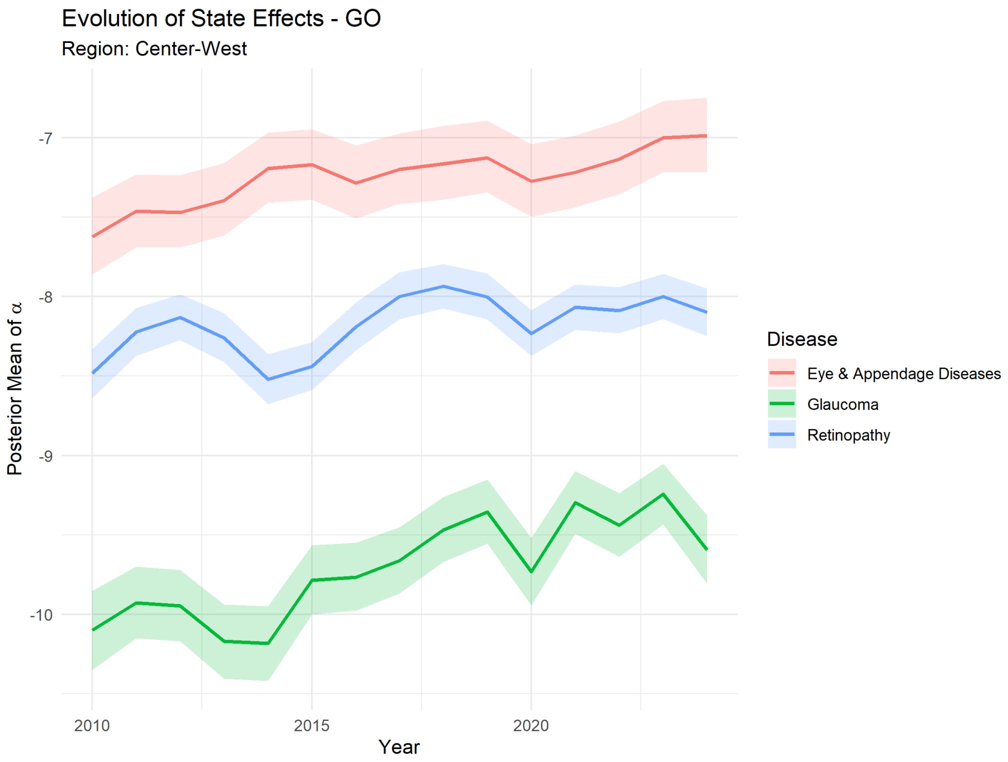

Figure 38: Goiás Posterior per Disease.

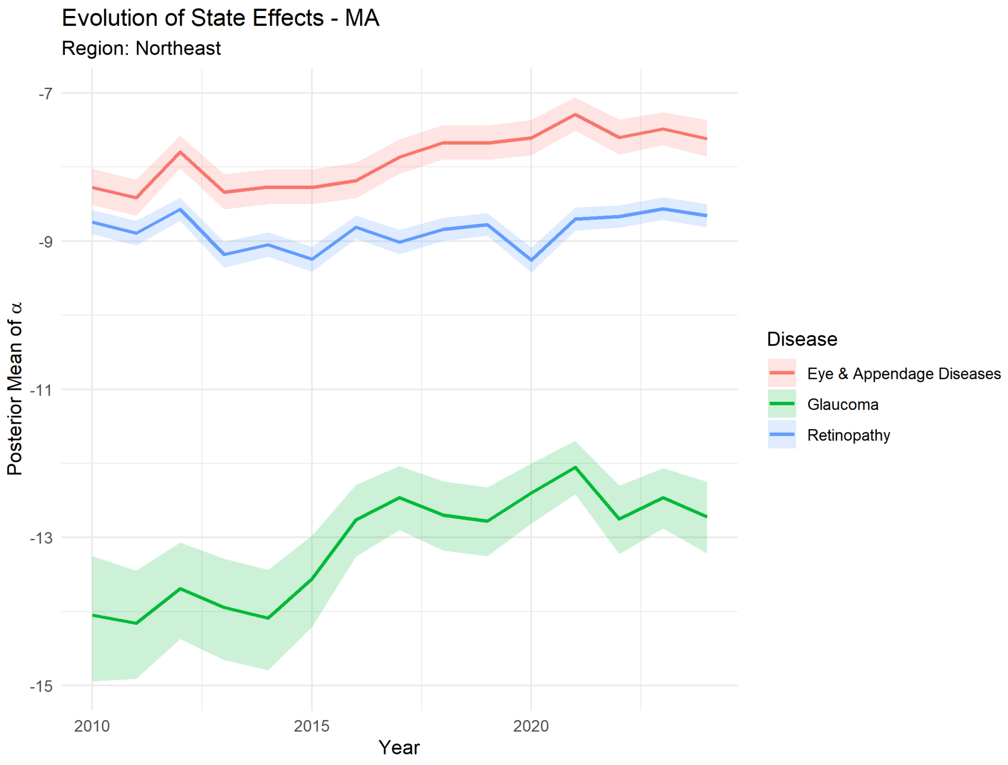

Figure 39: Maranhão Posterior per Disease.

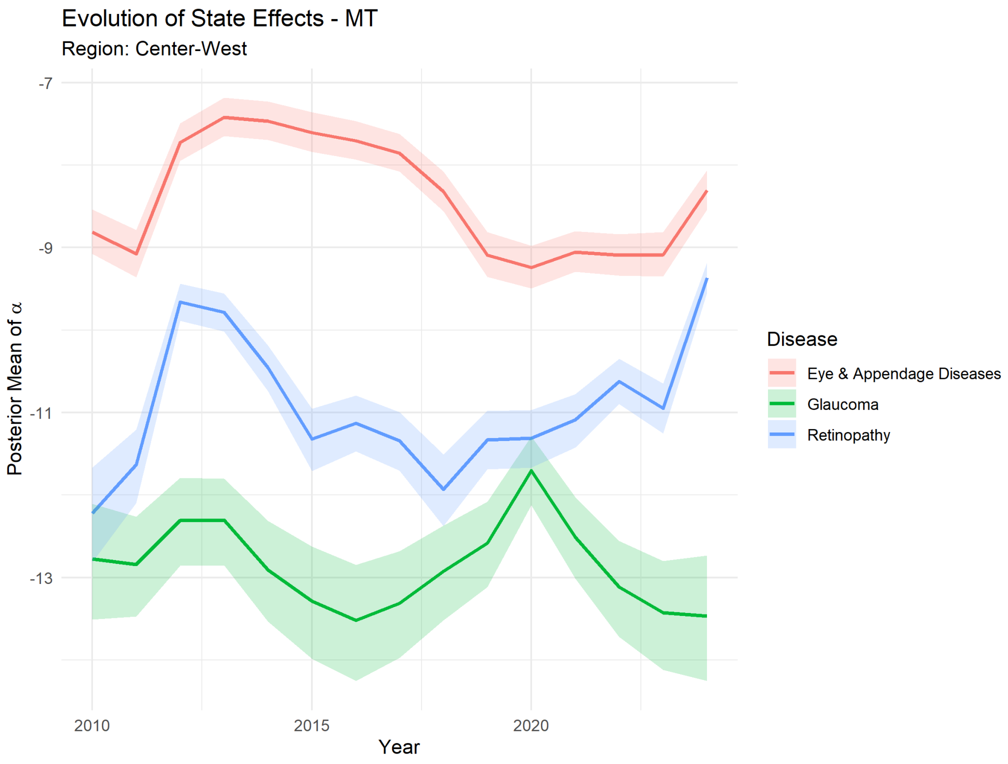

Figure 40: Mato Grosso Posterior per Disease.

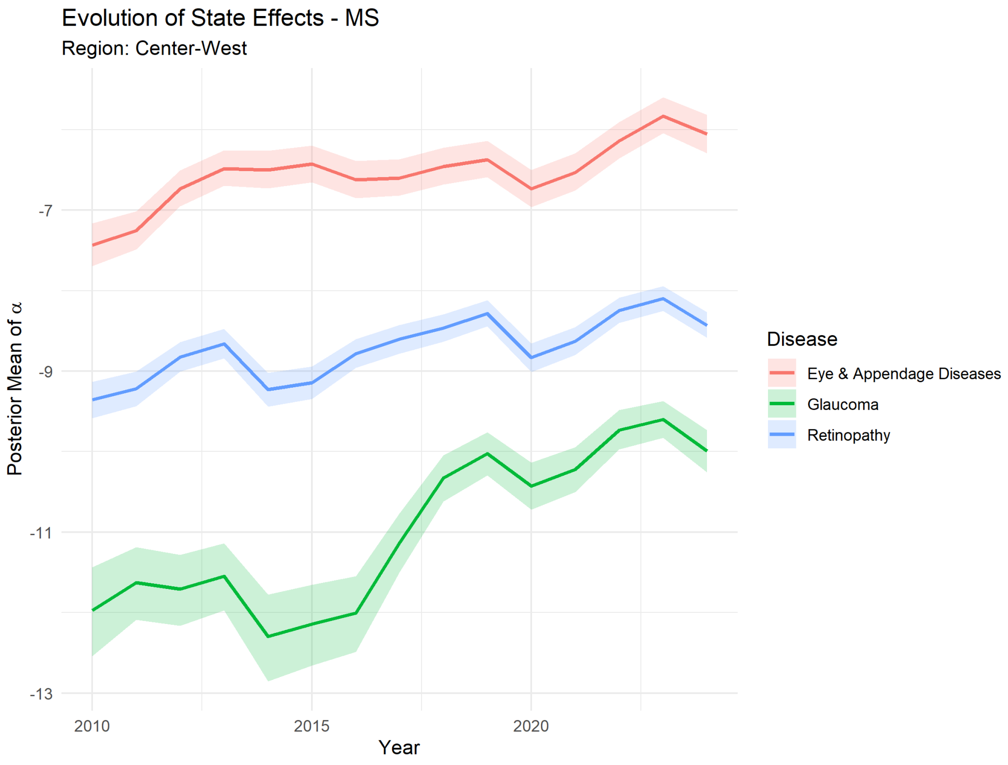

Figure 41: Mato Grosso do Sul Posterior per Disease.

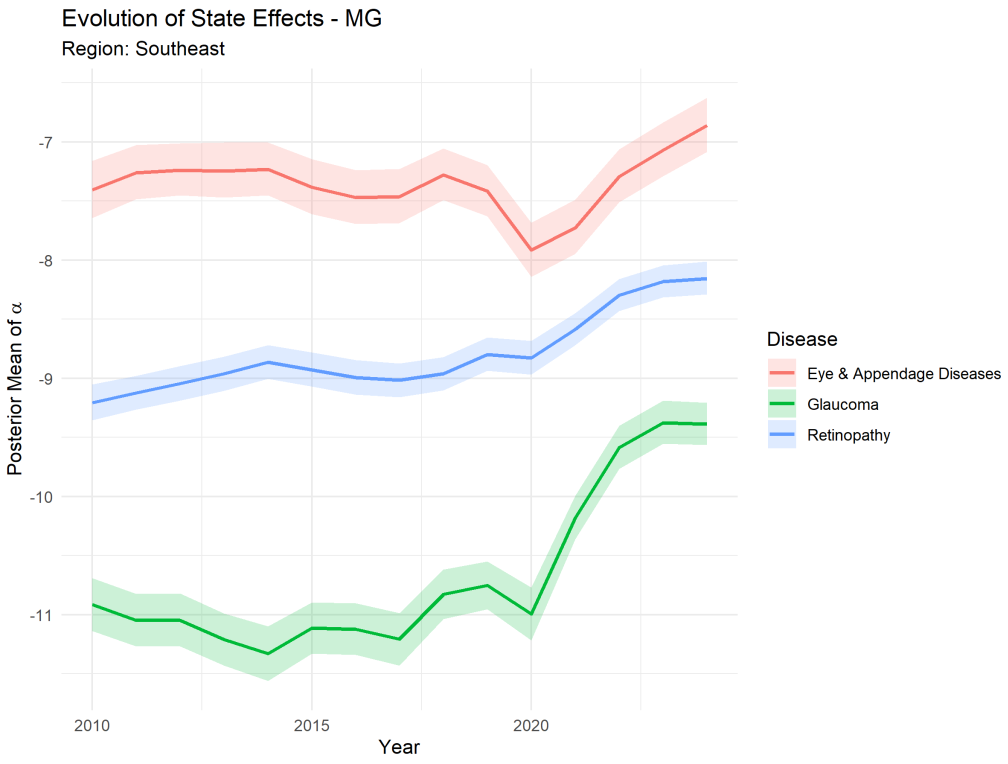

Figure 42: Minas Gerais Posterior per Disease.

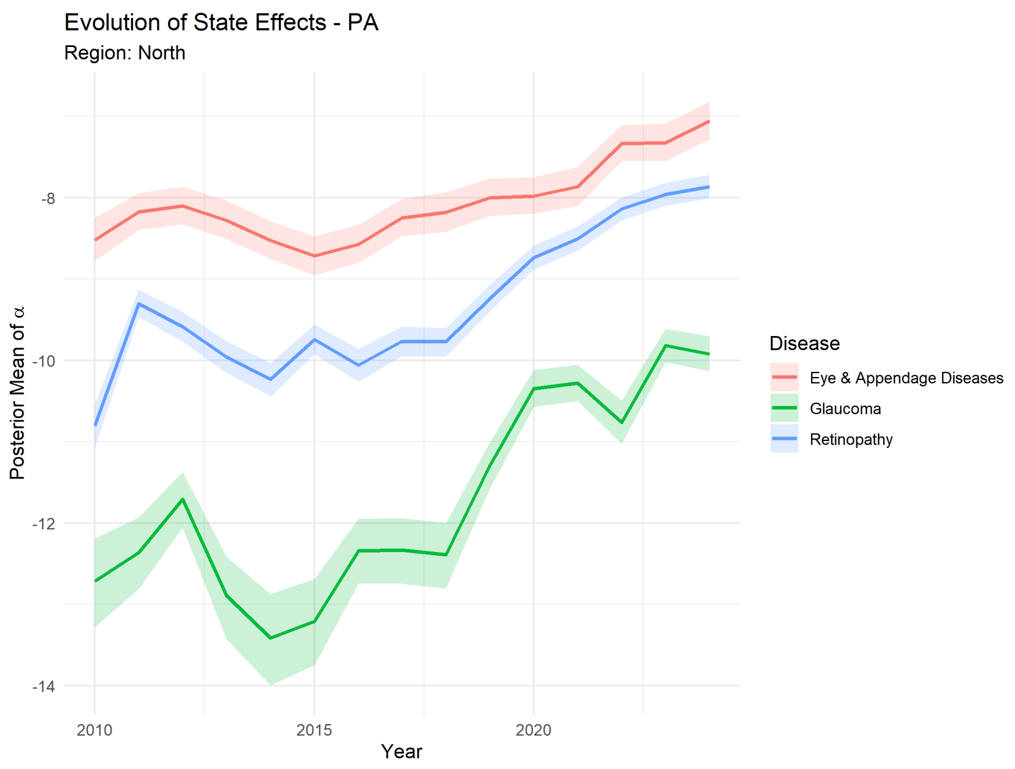

Figure 43: Pará Posterior per Disease.

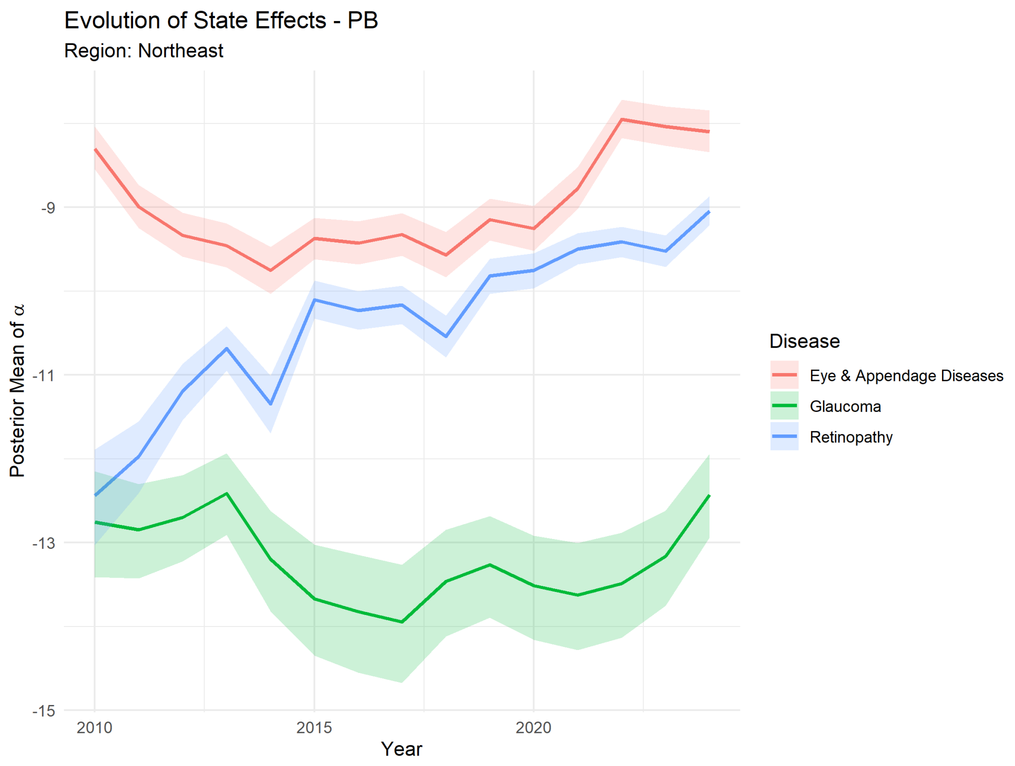

Figure 44: Paraíba Posterior per Disease.

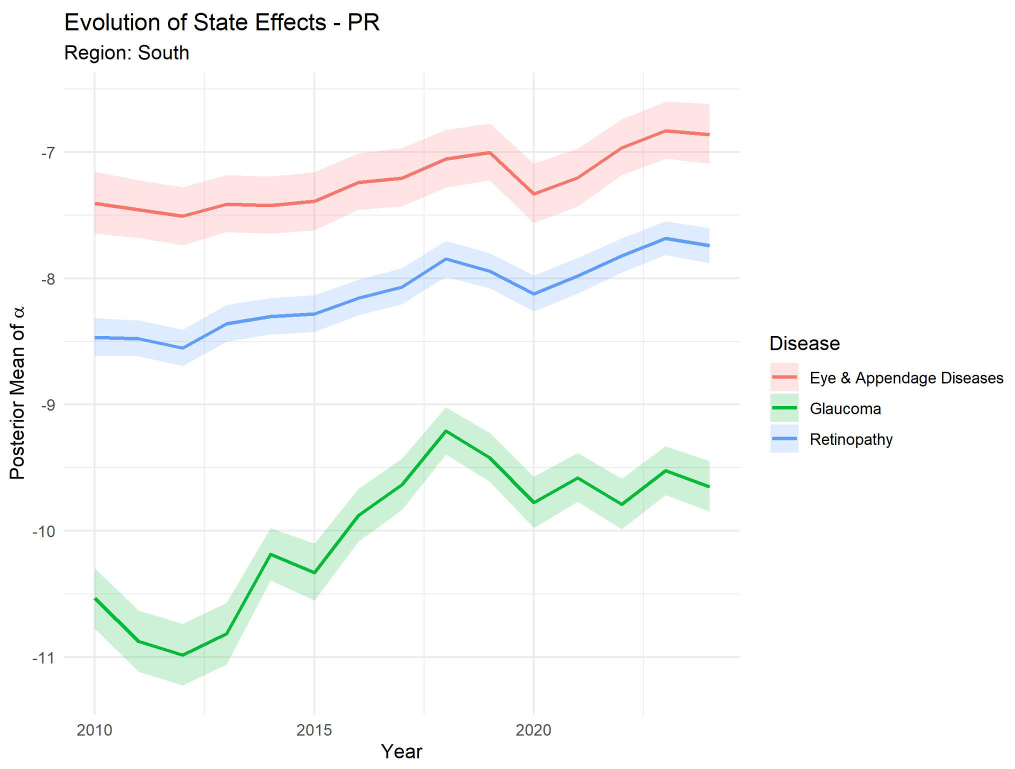

Figure 45: Paraná Posterior per Disease.

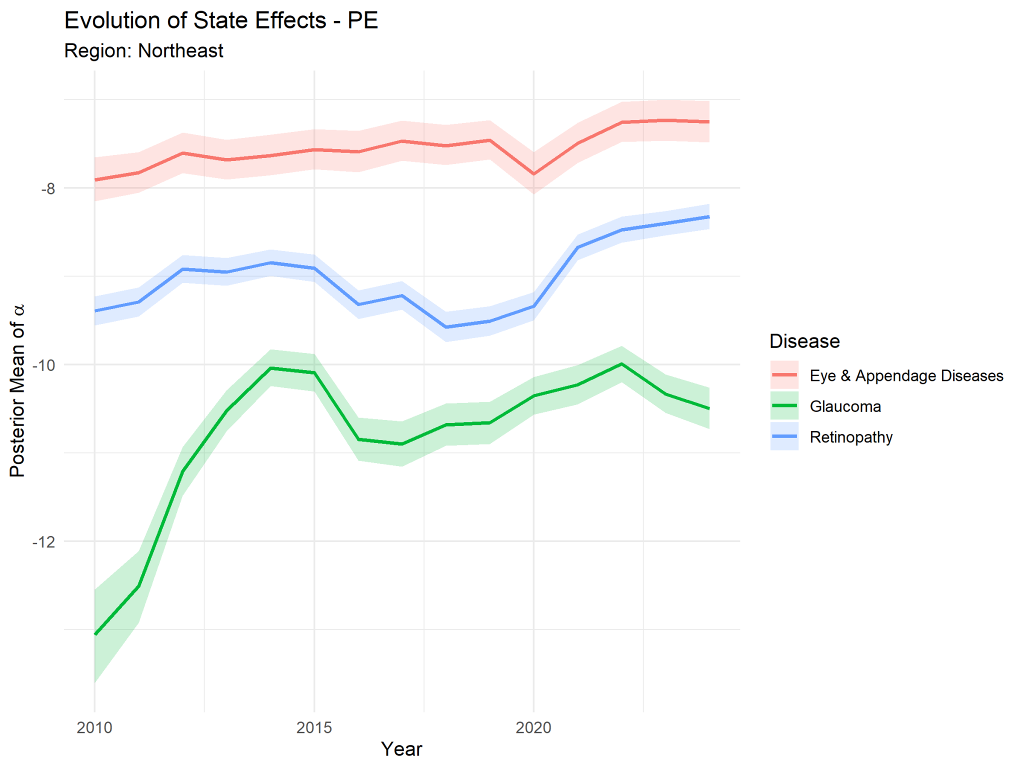

Figure 46: Pernambuco Posterior per Disease.

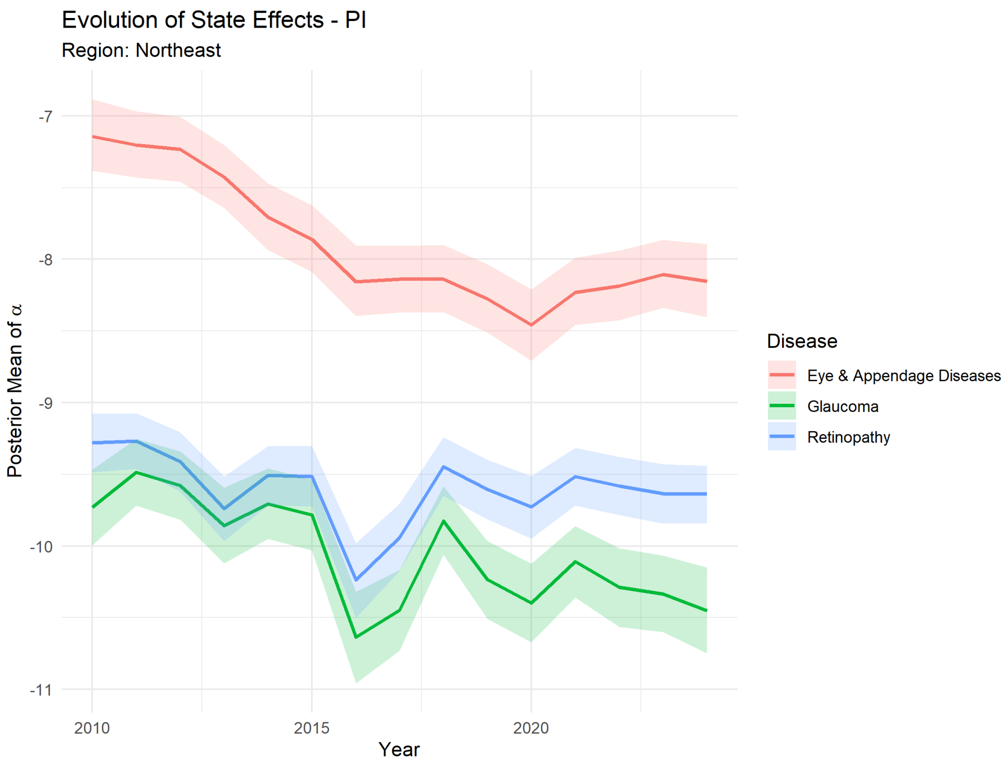

Figure 47: Piauí Posterior per Disease.

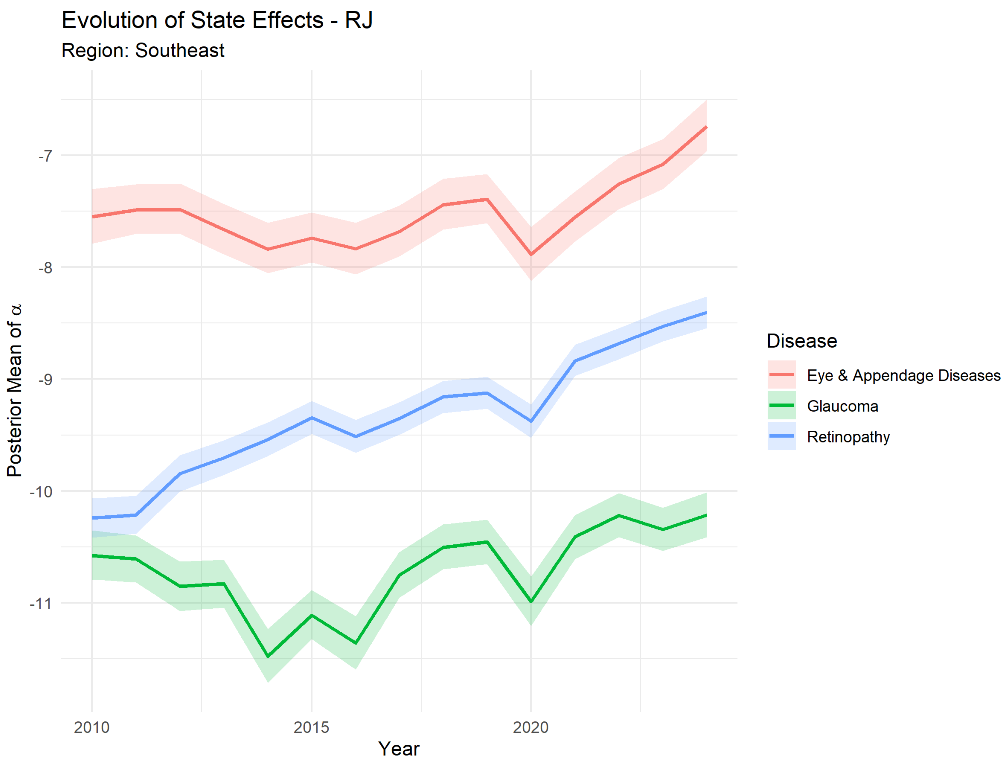

Figure 48: Rio de Janeiro Posterior per Disease.

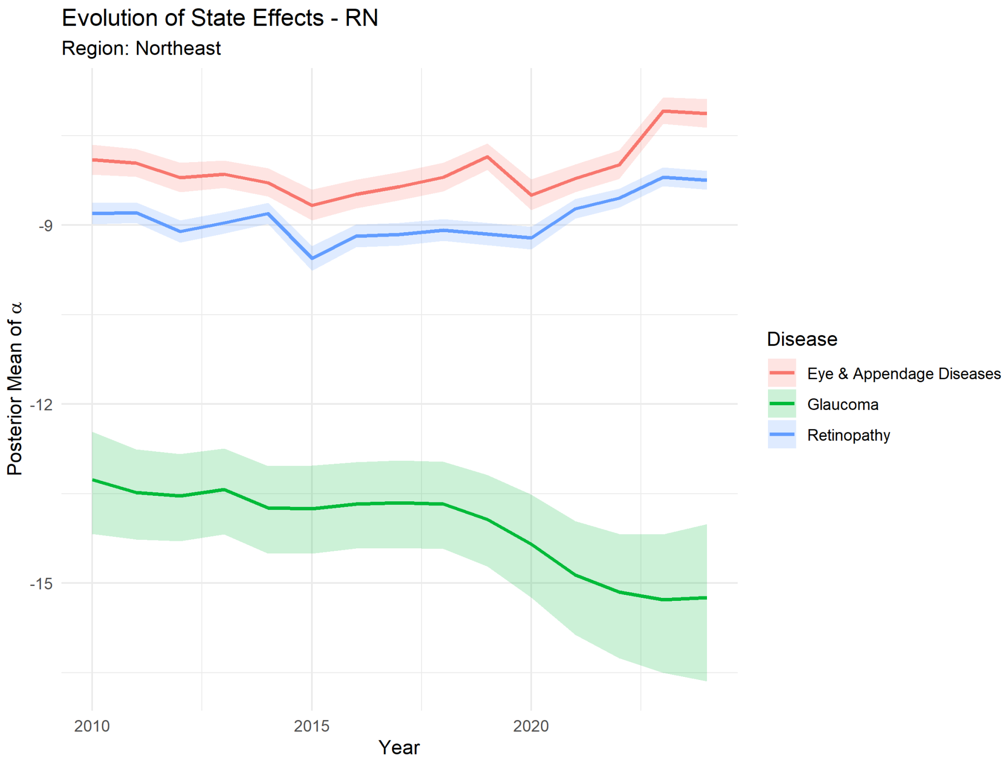

Figure 49: Rio Grande do Norte Posterior per Disease.

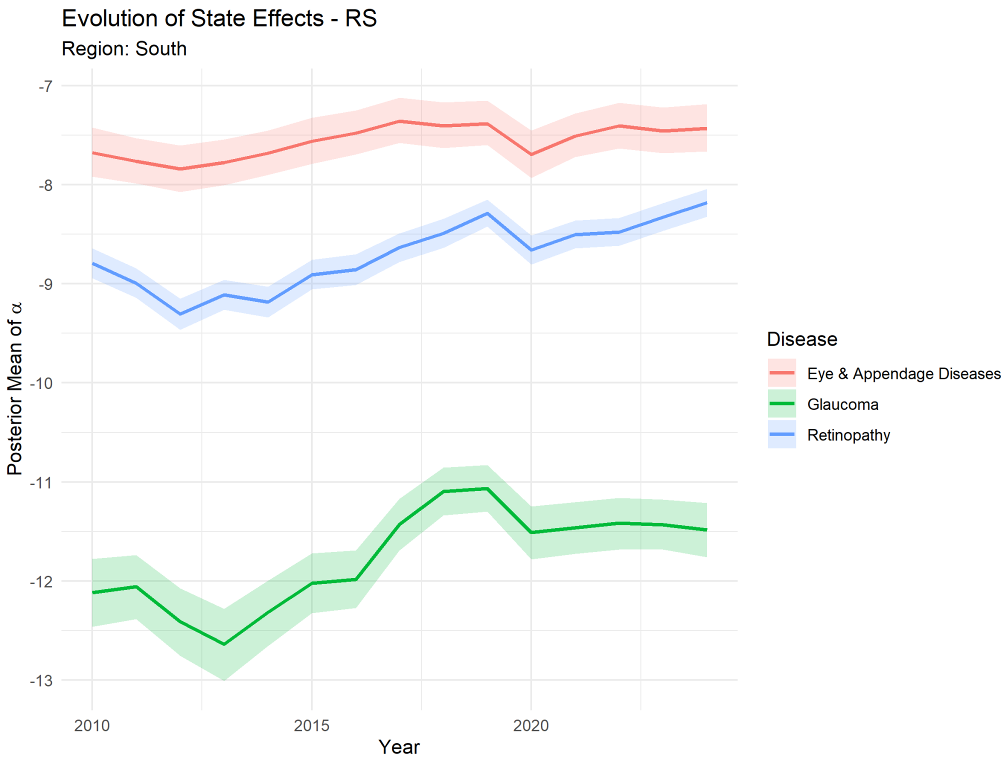

Figure 50: Rio Grande do Sul Posterior per Disease.

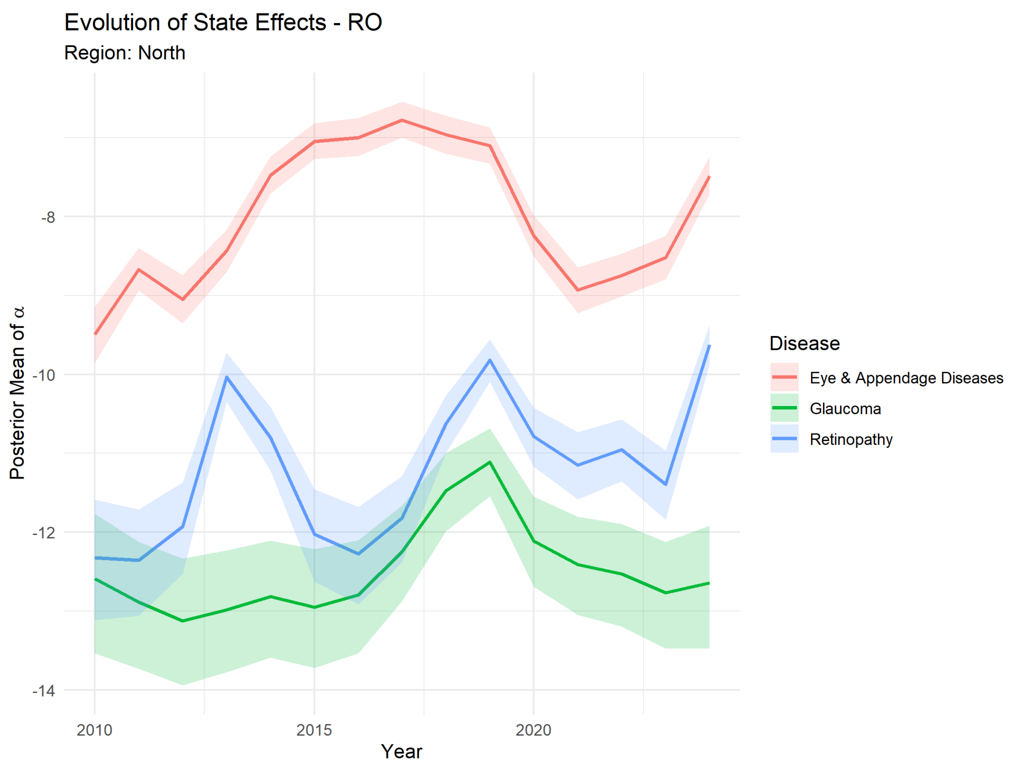

Figure 51: Rondônia Posterior per Disease.

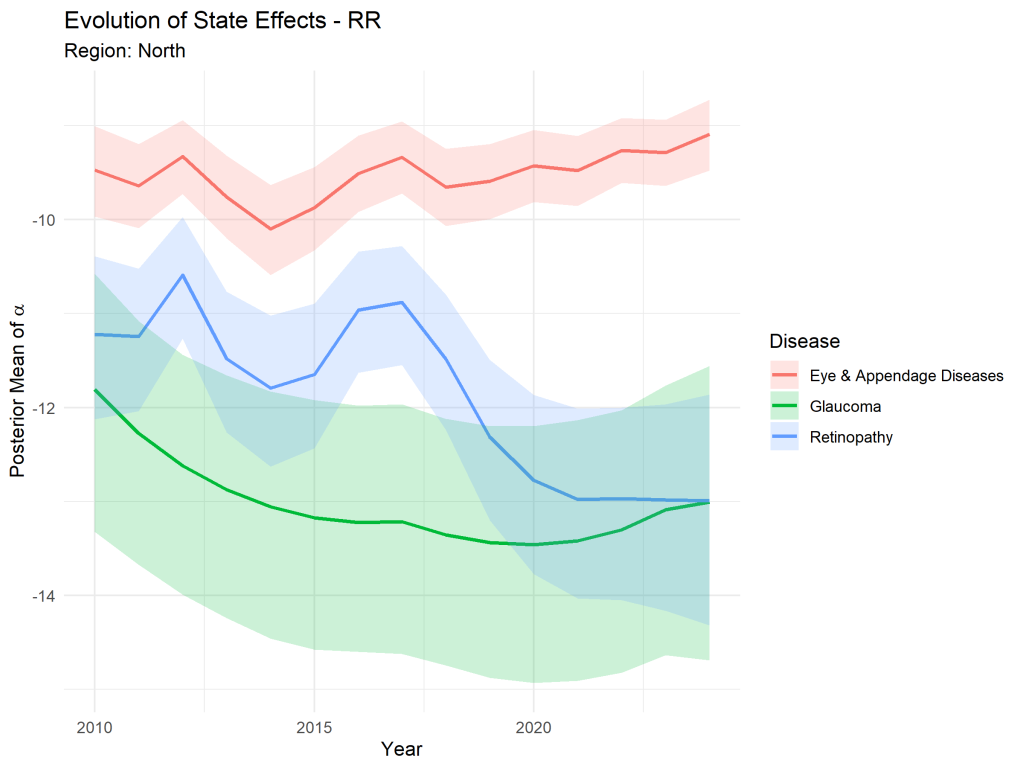

Figure 52: Roraima Posterior per Disease.

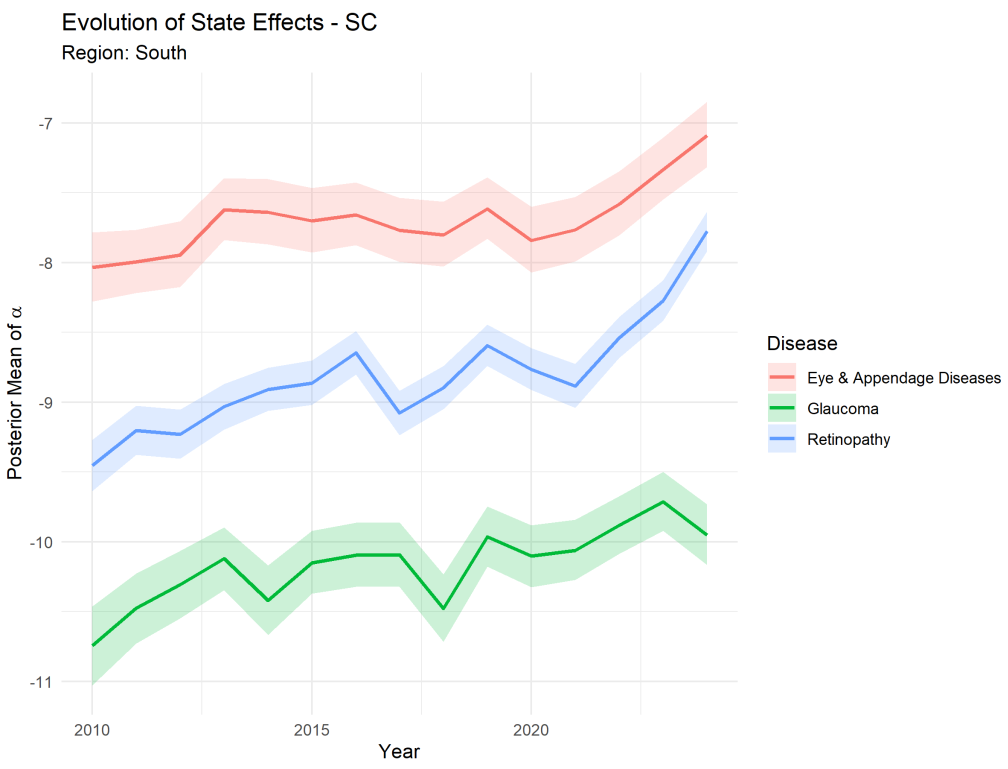

Figure 53: Santa Catarina Posterior per Disease.

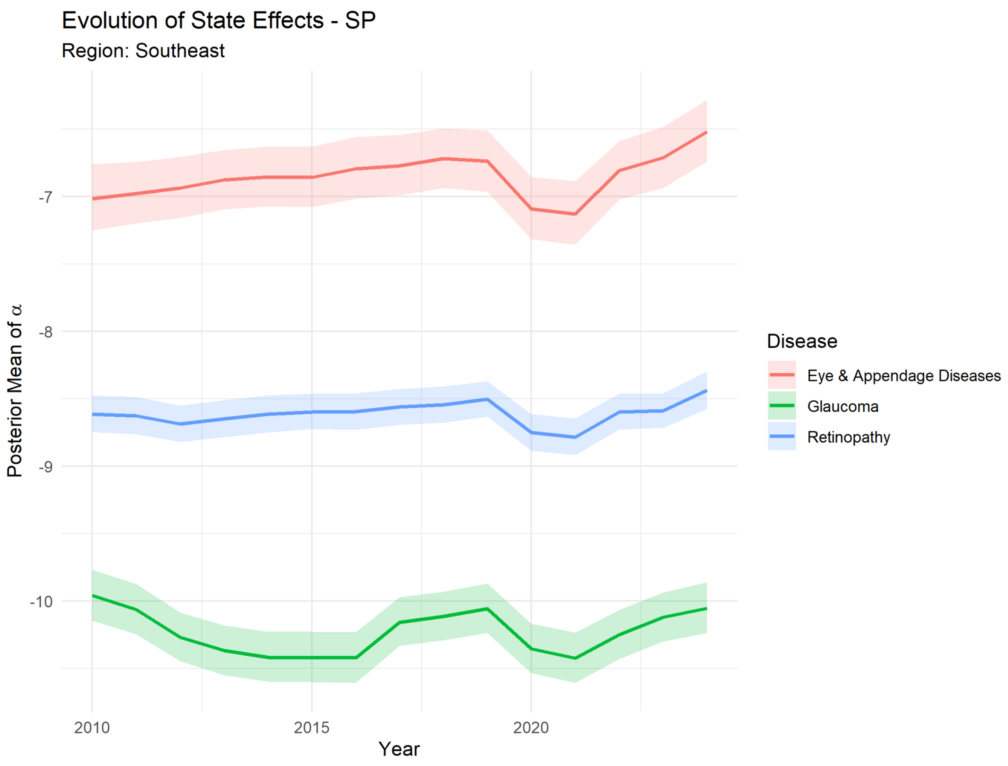

Figure 54: São Paulo Posterior per Disease.

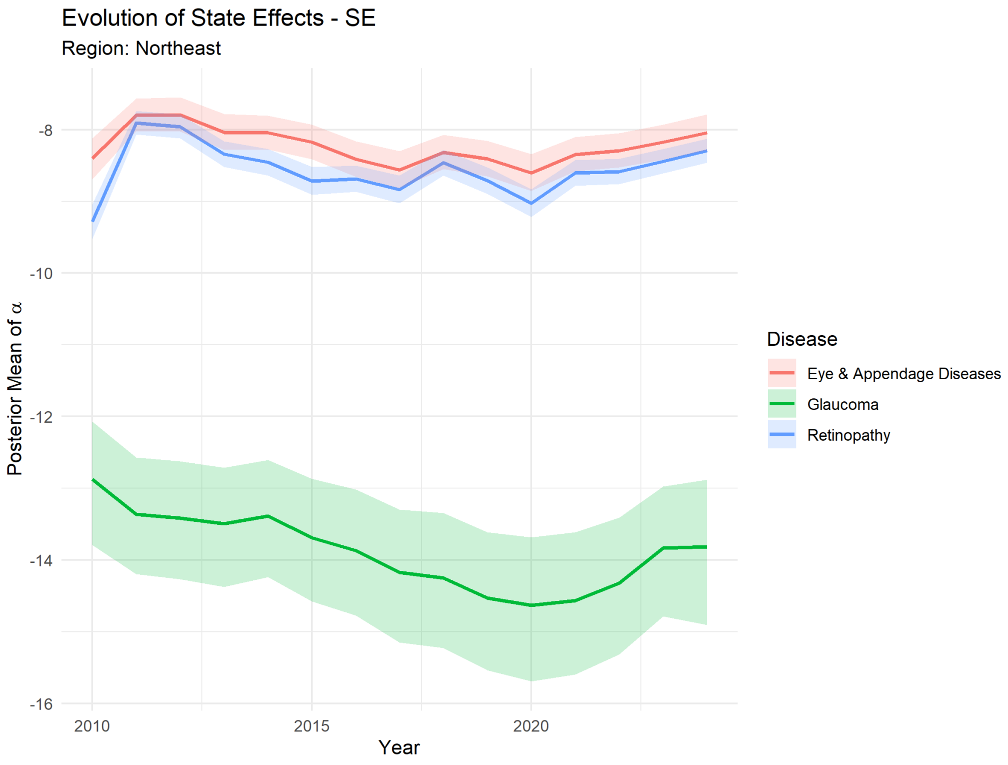

Figure 55: Sergipe Posterior per Disease.

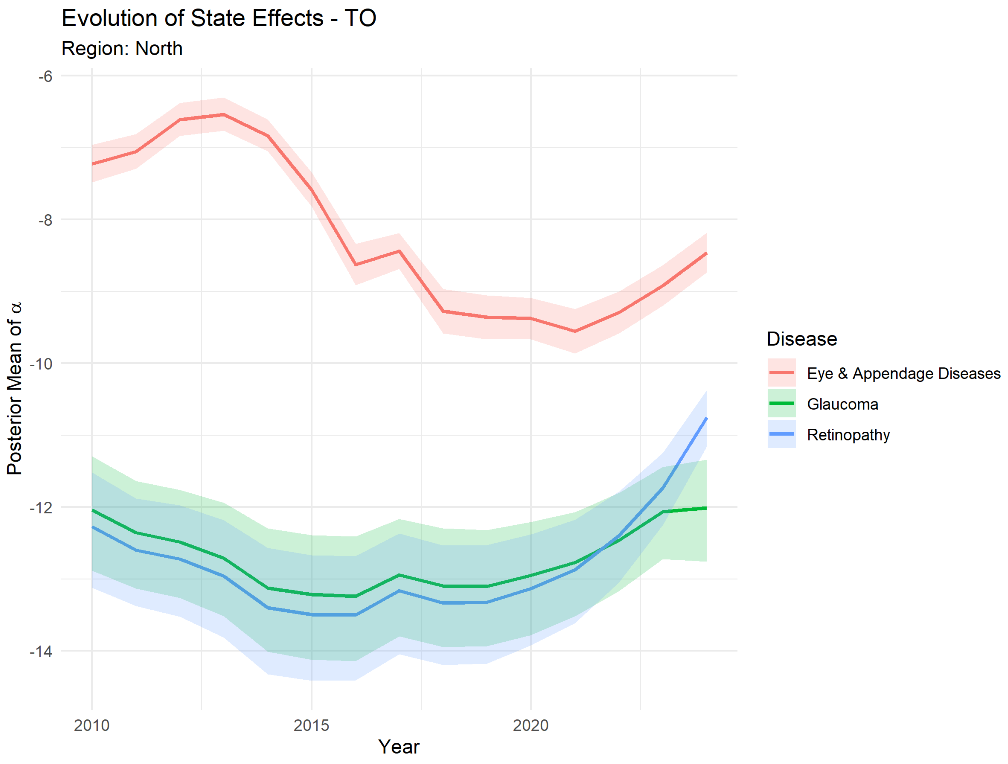

Figure 56: Tocantins Posterior per Disease.

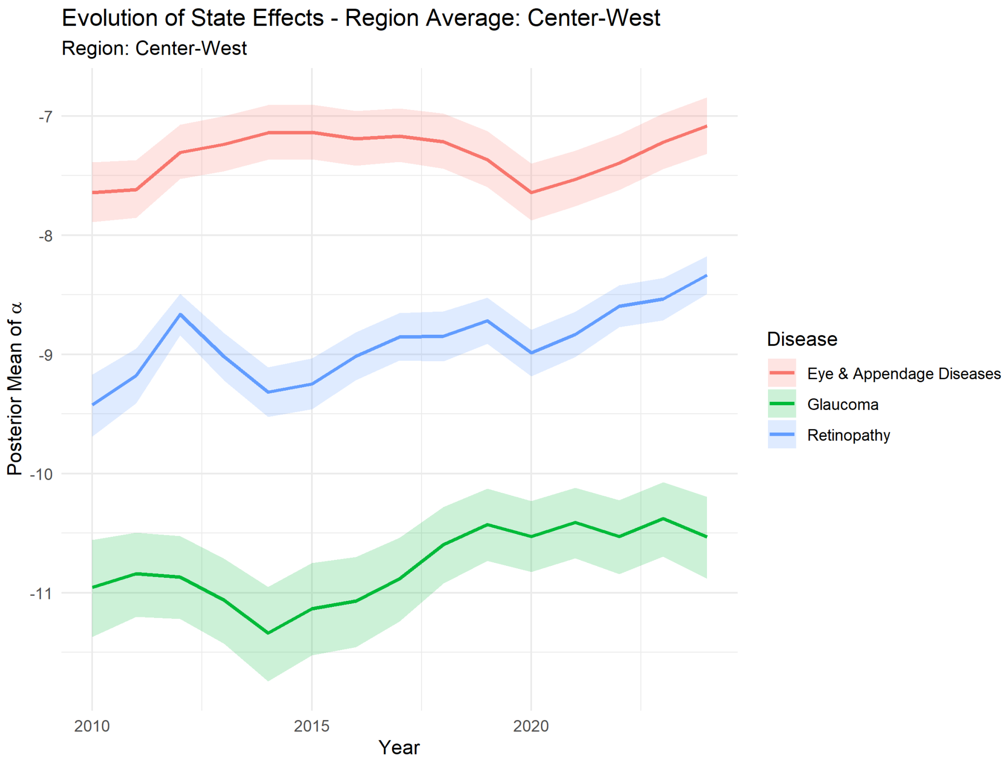

Figure 57: Center-West Posterior per Disease.

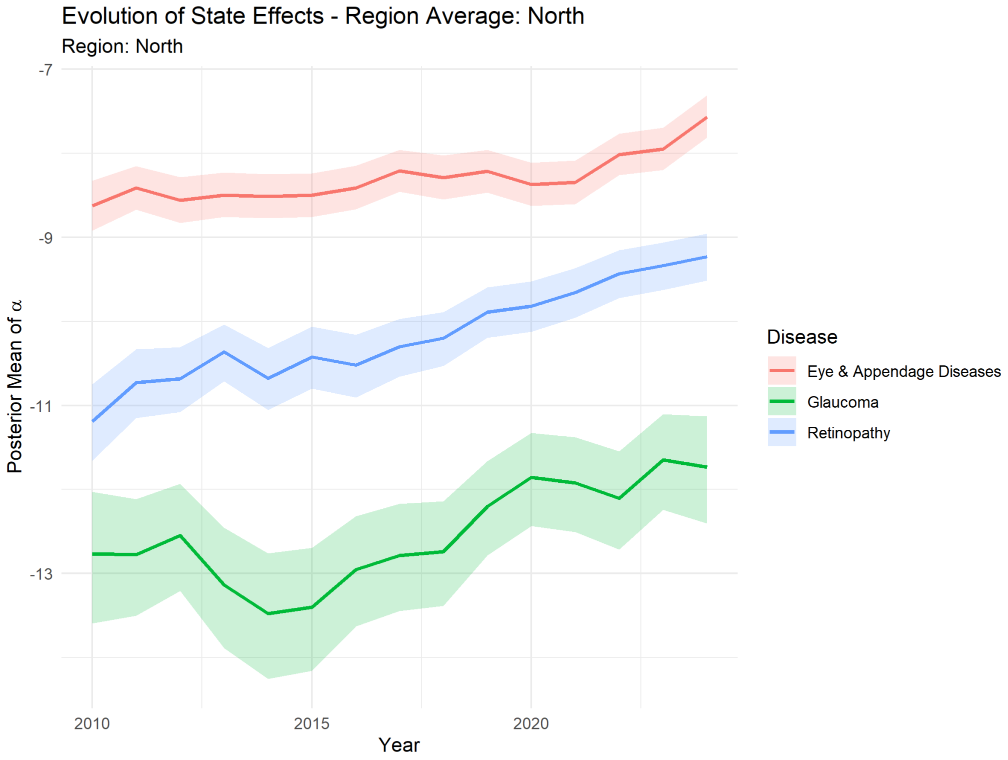

Figure 58: North Posterior per Disease.

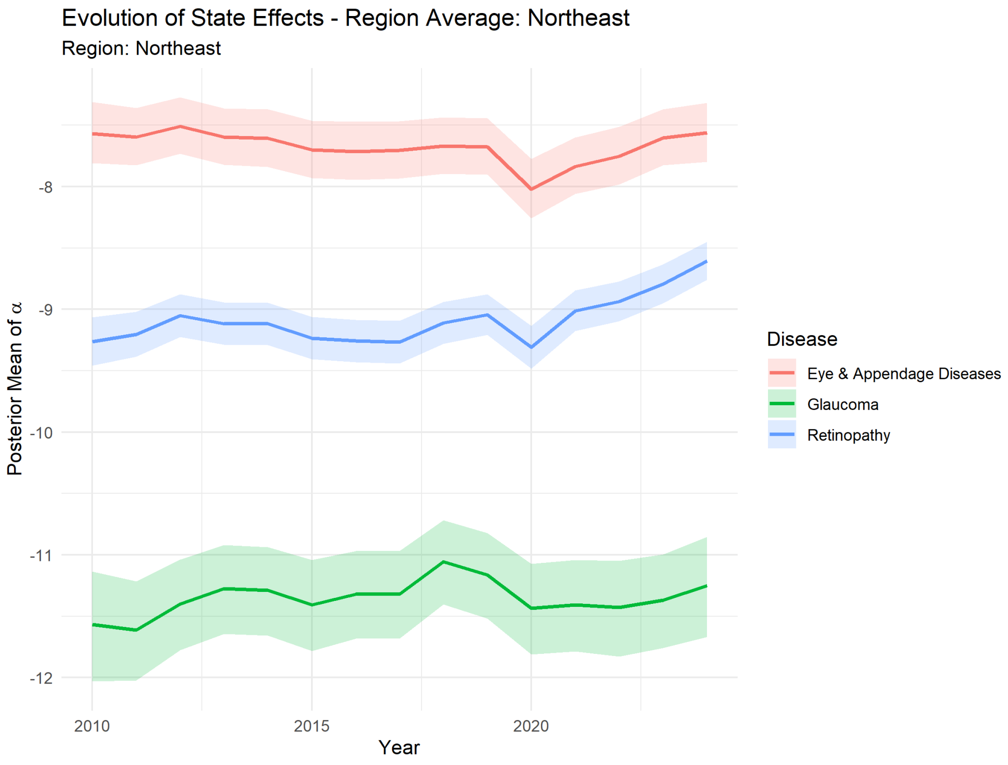

Figure 59: Northeast Posterior per Disease.

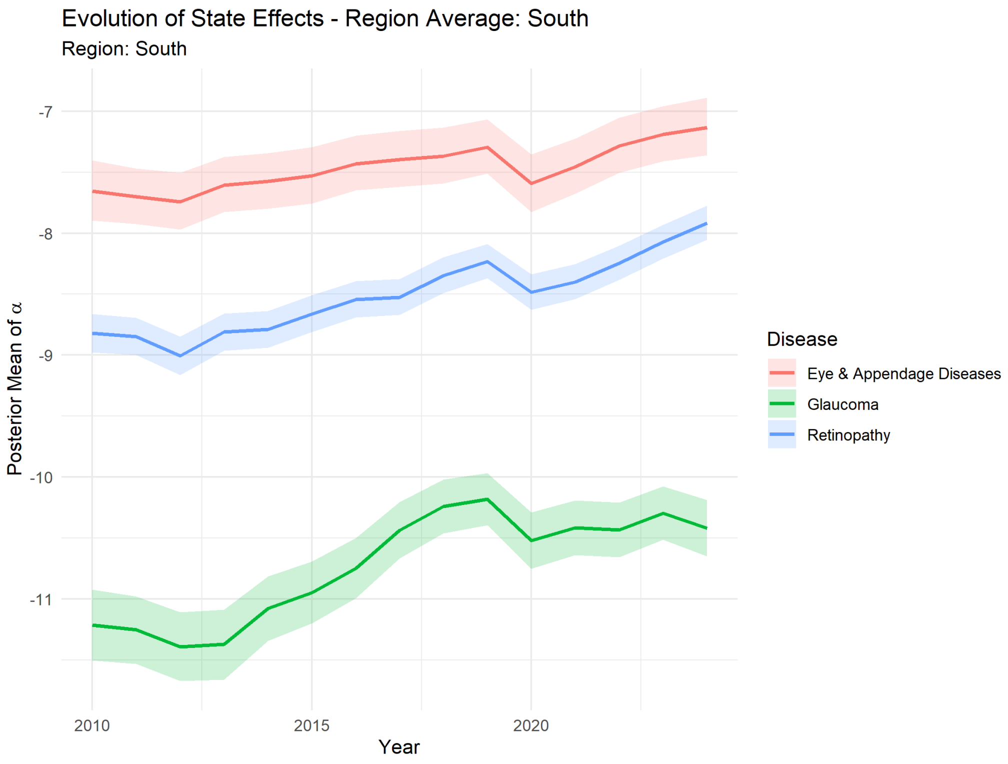

Figure 60: South Posterior per Disease.

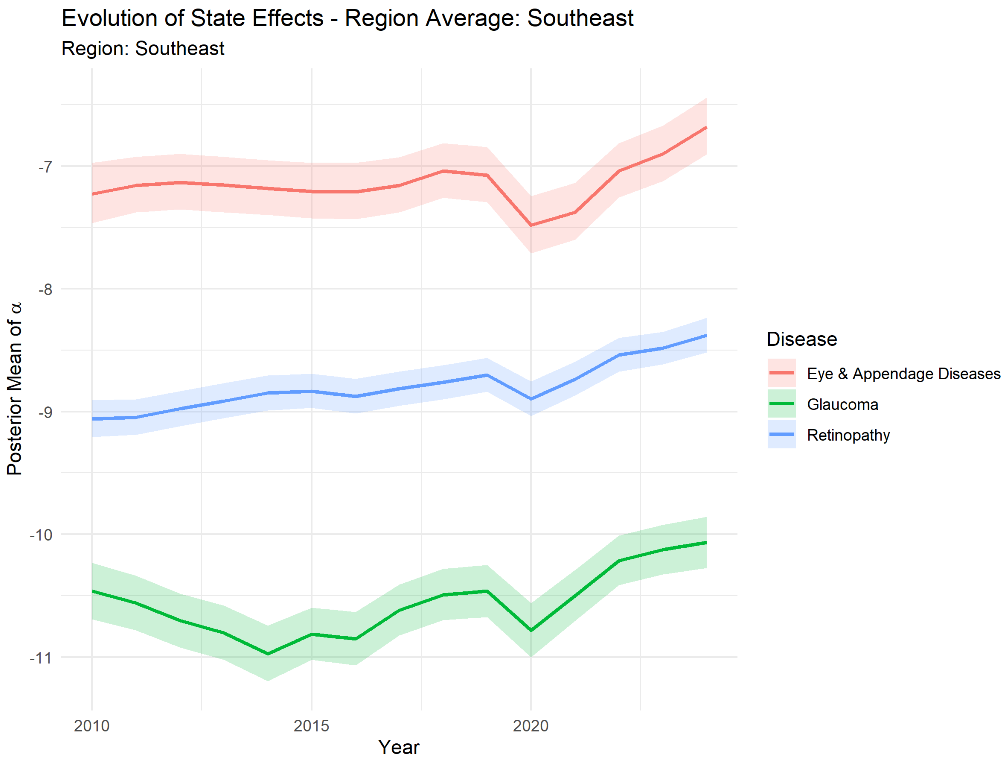

Figure 61: Southeast Posterior per Disease.

### 9 Observed State-level Change 2010-2024

The pooled national rate increases reported in the main paper (*Abstract* and Section *Results*) conceal substantial heterogeneity at the state level. Table 9 reports the observed 2010-to-2024 percentage change in hospitalization rate per million for each of the 27 federative units and each of the three diseases. Several small-population Northern states show very large percentage changes driven by very low 2010 baselines and should be interpreted as numerically unstable. Across stable units, median state-level changes are approximately +200%, +140% and +126% for retinopathy, eye-and-appendage diseases and glaucoma respectively, with between two and five states recording *decreases* over the period for each disease.

Table 9: State-level observed 2010-2024 hospitalization rate change (%) per disease. States are ordered alphabetically. "-" indicates undefined (zero or missing 2010 rate).

| State | Glaucoma (%) | Retinopathy (%) | Eye & App. (%) |
|---|---|---|---|
| AC | - | - | -24.6 |
| AL | -93.1 | +61.4 | -29.5 |
| AM | - | +197.4 | +455.9 |
| AP | - | - | +452.1 |
| BA | +126.0 | +140.7 | +33.4 |
| CE | +7.1 | +32.9 | -54.6 |
| DF | +49.1 | +206.8 | +5.7 |
| ES | +528.1 | -36.4 | +17.8 |
| GO | +120.3 | +83.2 | +176.4 |
| MA | - | +23.4 | +130.1 |
| MG | +620.8 | +318.5 | +138.6 |
| MS | +1698.2 | +228.5 | +475.0 |
| MT | +54.6 | +6855.8 | +150.2 |
| PA | +3588.4 | +2917.9 | +569.7 |
| PB | +202.1 | +6847.4 | +98.1 |
| PE | +2825.7 | +285.4 | +176.4 |
| PI | -40.3 | -14.2 | -65.5 |
| PR | +210.6 | +176.6 | +144.3 |
| RJ | +116.7 | +823.3 | +199.7 |
| RN | -27.7 | +115.0 | +282.2 |
| RO | +279.6 | +7612.0 | +1718.9 |
| RR | - | - | +67.0 |
| RS | +131.9 | +149.9 | +118.5 |
| SC | +216.2 | +713.7 | +298.9 |
| SE | -21.9 | +251.7 | +121.8 |
| SP | +22.6 | +59.4 | +160.4 |
| TO | +126.8 | +1022.3 | -68.6 |

### 10 Posterior Credible Intervals on 2024 Age × Region Cells

To contextualize the age- and region-specific trajectories shown in Fig. 2 of the main text, we report posterior summaries for the fitted 2024 hospitalization rates (per million inhabitants) across all disease × region × age-stratum combinations. These estimates represent the model-based counterparts to the observed rates plotted in the historical panels and are derived from the fitted `nb5_age_cat` hierarchical negative binomial model.

Tables 10-12 summarize posterior means together with their corresponding 95% credible intervals (CrIs). Because the hierarchical specification partially pools information across states and age strata within each disease category, the resulting intervals are generally narrower and more stable than naive cell-wise Poisson confidence intervals. Nevertheless, posterior uncertainty increases substantially in older age groups, particularly in the 75-89 and 90+ strata, where event counts per state become sparse and between-state heterogeneity is amplified.

The posterior summaries also illustrate the marked geographic heterogeneity discussed in the main manuscript. For example, retinopathy and eye-and-appendage disease rates remain consistently highest in the South and Southeast across nearly all age strata, whereas the North exhibits systematically lower estimated hospitalization rates. Importantly, the widening credible intervals in the oldest cohorts indicate that comparisons involving extreme-age groups should be interpreted cautiously, especially when discussing absolute rate magnitudes.

Table 10: Posterior mean and 95% credible intervals for 2024 glaucoma hospitalization rates (per million inhabitants) by region and age stratum.

| **Region** | **Age** | **Mean** | **95% CrI lo** | **95% CrI hi** |
|---|---|---|---|---|
| North | 25-34 | 2.8 | 2.3 | 3.4 |
| | 35-44 | 5.4 | 4.3 | 6.6 |
| | 45-54 | 13.9 | 11.3 | 17.0 |
| | 55-64 | 35.8 | 29.2 | 43.8 |
| | 65-74 | 81.6 | 66.4 | 100.0 |
| | 75-89 | 119.8 | 97.3 | 146.5 |
| | 90+ | 76.8 | 62.1 | 94.5 |
| Northeast | 25-34 | 3.9 | 3.4 | 4.5 |
| | 35-44 | 7.6 | 6.6 | 8.8 |
| | 45-54 | 20.4 | 17.7 | 23.6 |
| | 55-64 | 52.1 | 45.2 | 59.7 |
| | 65-74 | 119.5 | 103.9 | 136.9 |
| | 75-89 | 169.7 | 147.8 | 194.1 |
| | 90+ | 108.9 | 94.1 | 126.0 |
| Center-West | 25-34 | 5.4 | 4.6 | 6.3 |
| | 35-44 | 10.4 | 8.9 | 12.2 |
| | 45-54 | 26.7 | 23.0 | 31.3 |
| | 55-64 | 68.8 | 59.0 | 80.7 |
| | 65-74 | 155.7 | 133.2 | 181.1 |
| | 75-89 | 224.7 | 193.0 | 260.7 |
| | 90+ | 143.6 | 123.0 | 166.9 |
| Southeast | 25-34 | 5.8 | 5.1 | 6.6 |
| | 35-44 | 11.1 | 9.7 | 12.6 |
| | 45-54 | 28.3 | 25.0 | 32.1 |
| | 55-64 | 73.1 | 64.8 | 82.6 |
| | 65-74 | 165.8 | 147.0 | 188.2 |
| | 75-89 | 236.2 | 209.4 | 266.6 |
| | 90+ | 150.3 | 132.6 | 169.9 |
| South | 25-34 | 4.7 | 4.0 | 5.5 |
| | 35-44 | 9.1 | 7.8 | 10.5 |
| | 45-54 | 23.0 | 20.0 | 26.7 |
| | 55-64 | 59.1 | 50.9 | 68.4 |
| | 65-74 | 128.0 | 110.6 | 147.9 |
| | 75-89 | 178.7 | 155.4 | 206.8 |
| | 90+ | 111.0 | 95.9 | 128.8 |

Table 11: Posterior mean and 95% credible intervals for 2024 retinopathy hospitalization rates (per million inhabitants) by region and age stratum.

| **Region** | **Age** | **Mean** | **95% CrI lo** | **95% CrI hi** |
|---|---|---|---|---|
| North | 25-34 | 32.7 | 28.8 | 37.3 |
| | 35-44 | 57.1 | 50.0 | 64.7 |
| | 45-54 | 134.8 | 118.2 | 152.8 |
| | 55-64 | 359.5 | 316.5 | 407.7 |
| | 65-74 | 634.6 | 560.0 | 721.0 |
| | 75-89 | 684.3 | 600.7 | 776.9 |
| | 90+ | 367.7 | 322.5 | 417.9 |
| Northeast | 25-34 | 34.8 | 32.5 | 37.6 |
| | 35-44 | 61.0 | 56.8 | 65.3 |
| | 45-54 | 146.0 | 136.0 | 157.7 |
| | 55-64 | 386.9 | 360.0 | 416.8 |
| | 65-74 | 679.3 | 632.4 | 731.9 |
| | 75-89 | 712.4 | 663.9 | 766.9 |
| | 90+ | 381.6 | 354.8 | 411.4 |
| Center-West | 25-34 | 45.0 | 41.1 | 49.5 |
| | 35-44 | 78.5 | 71.4 | 86.4 |
| | 45-54 | 186.5 | 169.5 | 204.8 |
| | 55-64 | 490.8 | 447.8 | 539.2 |
| | 65-74 | 853.6 | 778.5 | 939.1 |
| | 75-89 | 906.9 | 826.9 | 998.9 |
| | 90+ | 483.8 | 441.2 | 533.8 |
| Southeast | 25-34 | 38.0 | 34.8 | 41.4 |
| | 35-44 | 66.2 | 60.7 | 72.2 |
| | 45-54 | 155.3 | 142.2 | 169.6 |
| | 55-64 | 413.9 | 380.3 | 451.6 |
| | 65-74 | 727.1 | 668.6 | 792.6 |
| | 75-89 | 766.7 | 703.3 | 833.0 |
| | 90+ | 409.6 | 376.0 | 446.9 |
| South | 25-34 | 61.2 | 56.1 | 66.5 |
| | 35-44 | 107.1 | 98.2 | 116.5 |
| | 45-54 | 250.7 | 230.2 | 271.8 |
| | 55-64 | 666.1 | 614.0 | 723.4 |
| | 65-74 | 1152.2 | 1060.5 | 1252.9 |
| | 75-89 | 1205.9 | 1107.3 | 1308.4 |
| | 90+ | 639.0 | 586.4 | 695.7 |

Table 12: Posterior mean and 95% credible intervals for 2024 eye and appendage disease hospitalization rates (per million inhabitants) by region and age stratum.

| Region | Age | Mean | 95% CrI lo | 95% CrI hi |
|---|---|---|---|---|
| North | 25-34 | 119.0 | 100.5 | 140.6 |
| | 35-44 | 160.0 | 135.6 | 189.2 |
| | 45-54 | 298.3 | 252.3 | 352.9 |
| | 55-64 | 732.7 | 620.8 | 870.5 |
| | 65-74 | 1731.1 | 1473.5 | 2051.7 |
| | 75-89 | 2606.0 | 2213.1 | 3098.2 |
| | 90+ | 1781.9 | 1503.6 | 2115.3 |
| Northeast | 25-34 | 113.8 | 100.4 | 128.8 |
| | 35-44 | 153.6 | 136.7 | 173.8 |
| | 45-54 | 290.4 | 255.9 | 328.1 |
| | 55-64 | 708.2 | 624.8 | 802.2 |
| | 65-74 | 1671.7 | 1476.2 | 1889.6 |
| | 75-89 | 2492.9 | 2201.0 | 2823.0 |
| | 90+ | 1721.6 | 1522.5 | 1953.3 |
| Center-West | 25-34 | 203.6 | 176.3 | 235.5 |
| | 35-44 | 273.0 | 237.4 | 315.3 |
| | 45-54 | 506.4 | 438.4 | 581.4 |
| | 55-64 | 1249.0 | 1084.8 | 1436.8 |
| | 65-74 | 2992.3 | 2585.4 | 3454.3 |
| | 75-89 | 4547.3 | 3926.5 | 5252.9 |
| | 90+ | 3204.3 | 2776.9 | 3715.7 |
| Southeast | 25-34 | 250.1 | 214.8 | 293.2 |
| | 35-44 | 337.2 | 289.1 | 395.0 |
| | 45-54 | 627.0 | 538.4 | 738.7 |
| | 55-64 | 1532.9 | 1316.1 | 1798.4 |
| | 65-74 | 3599.9 | 3104.8 | 4211.1 |
| | 75-89 | 5390.5 | 4650.1 | 6298.4 |
| | 90+ | 3684.9 | 3188.1 | 4303.5 |
| South | 25-34 | 163.1 | 140.4 | 190.1 |
| | 35-44 | 219.3 | 188.9 | 254.2 |
| | 45-54 | 405.8 | 349.6 | 468.8 |
| | 55-64 | 993.2 | 857.8 | 1151.8 |
| | 65-74 | 2298.2 | 1988.9 | 2676.3 |
| | 75-89 | 3415.4 | 2967.6 | 3954.5 |
| | 90+ | 2327.6 | 2012.6 | 2686.1 |

### 11 Forecasts

We forecast hospitalization rates from 2025 to 2036 by forward-simulating the state-level random walk on $\alpha_{s,t}$ for each posterior draw, holding the categorical age coefficients fixed and using IBGE projected populations as future denominators. For each draw, predicted state-year cases were summed across age groups; regional and national rates were obtained by aggregating predicted cases across the relevant states and dividing by the aggregated regional or national population. Posterior summaries are reported as means with 80% and 95% credible intervals. Sections below show aggregated regional and national forecasts (Sec. 11.1) followed by per-state forecasts (Sec. 11.2).

#### 11.1 Aggregated Forecasts

##### 11.1.1 Glaucoma

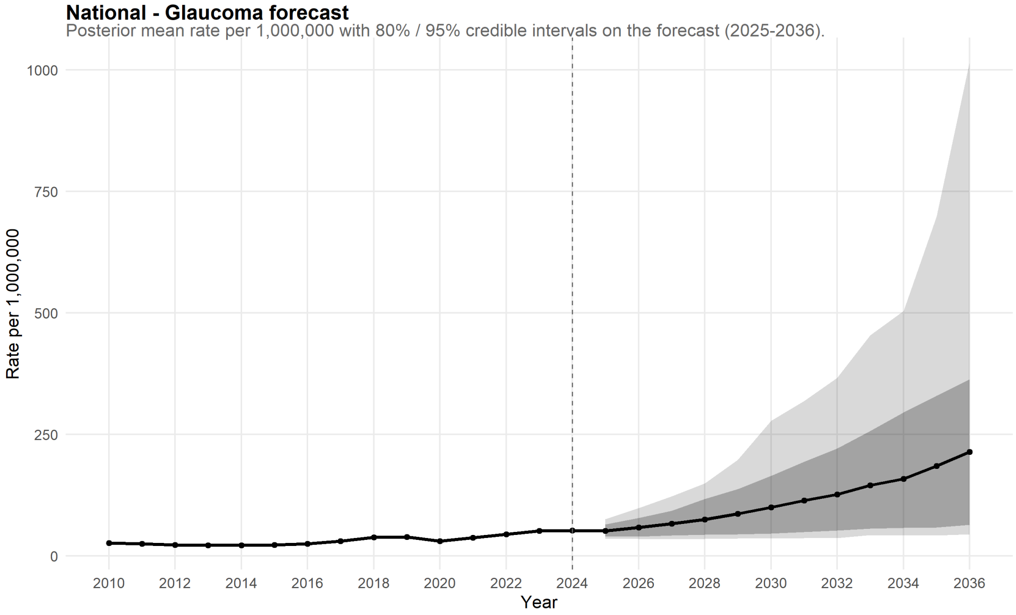

Figure 62: National Forecast for Glaucoma.

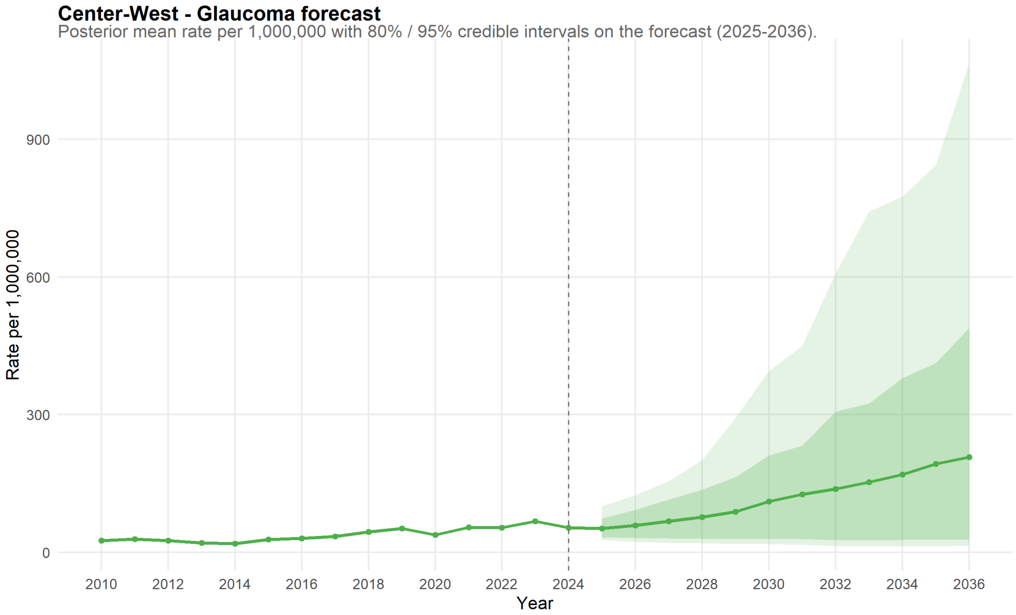

Figure 63: Center-West Forecast for Glaucoma.

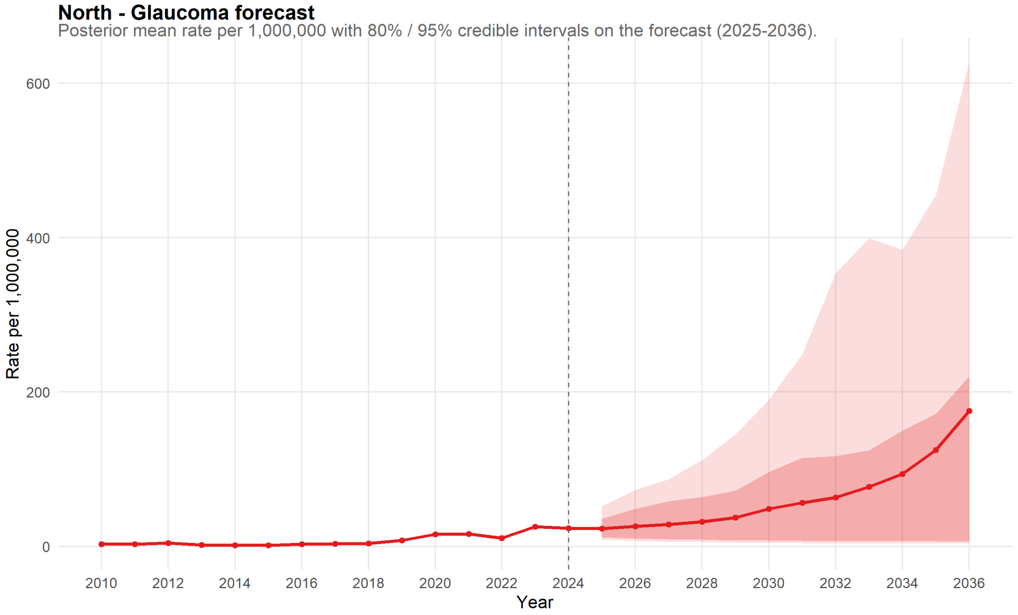

Figure 64: North Forecast for Glaucoma.

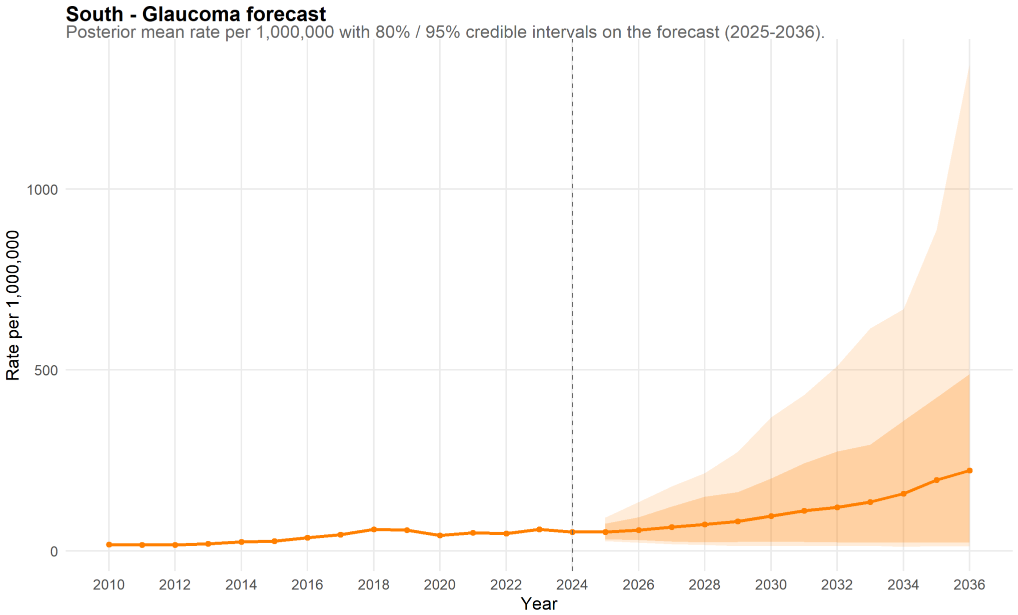

Figure 65: South Forecast for Glaucoma.

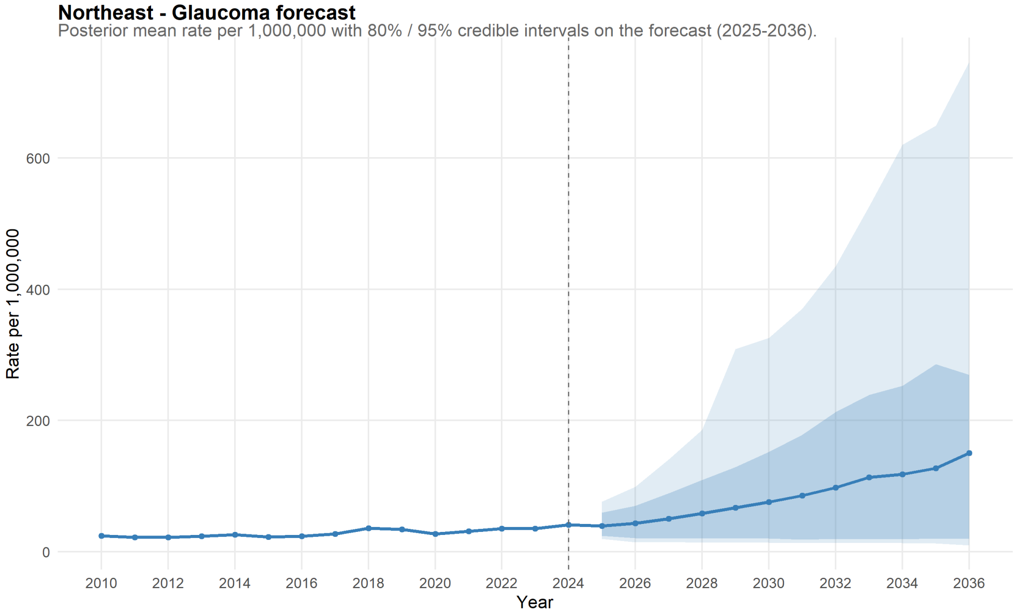

Figure 66: Northeast Forecast for Glaucoma.

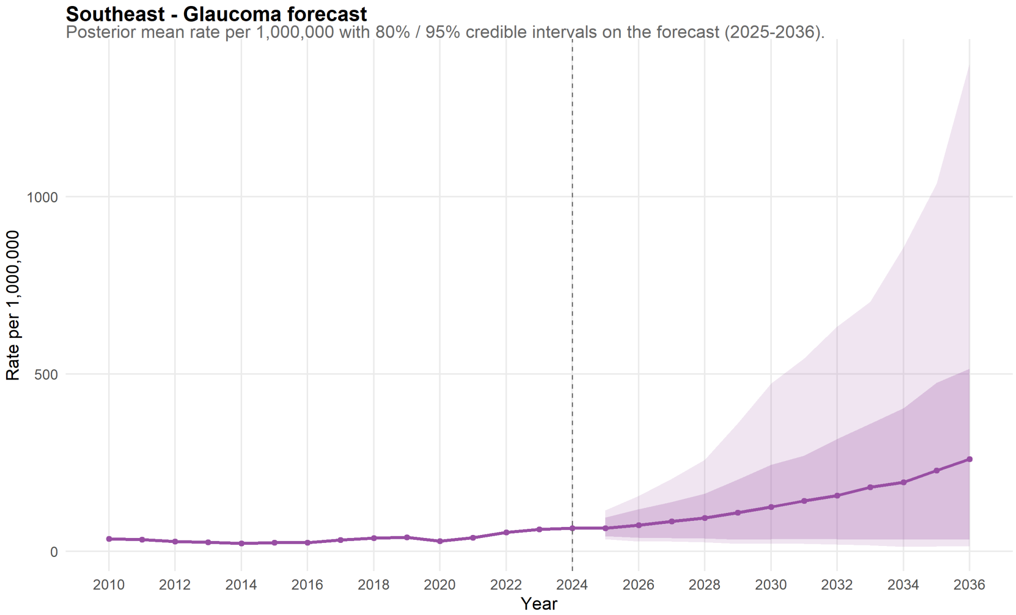

Figure 67: Southeast Forecast for Glaucoma.

##### 11.1.2 Retinopathy

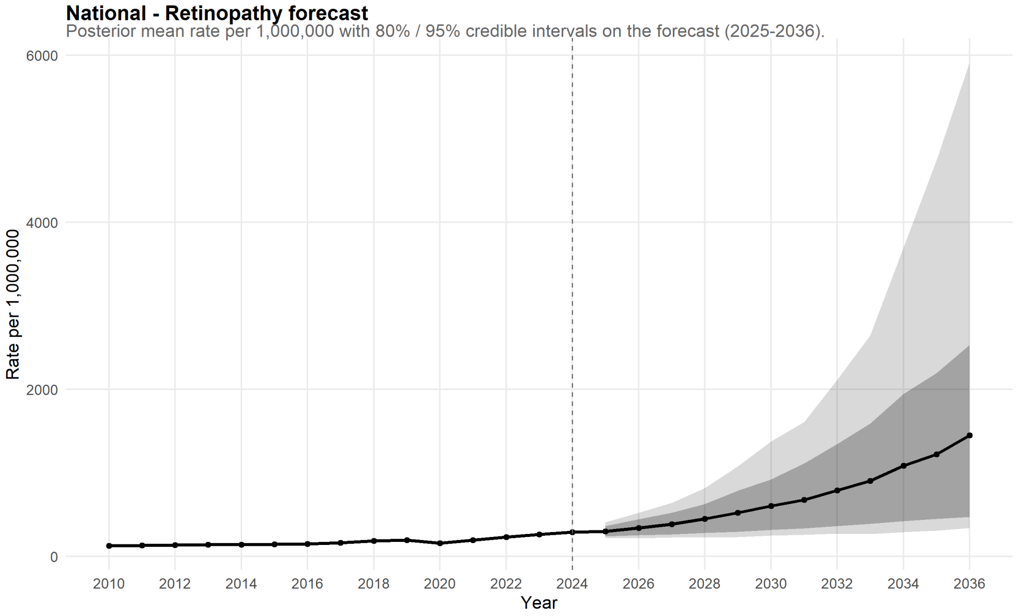

Figure 68: National Forecast for Retinopathy.

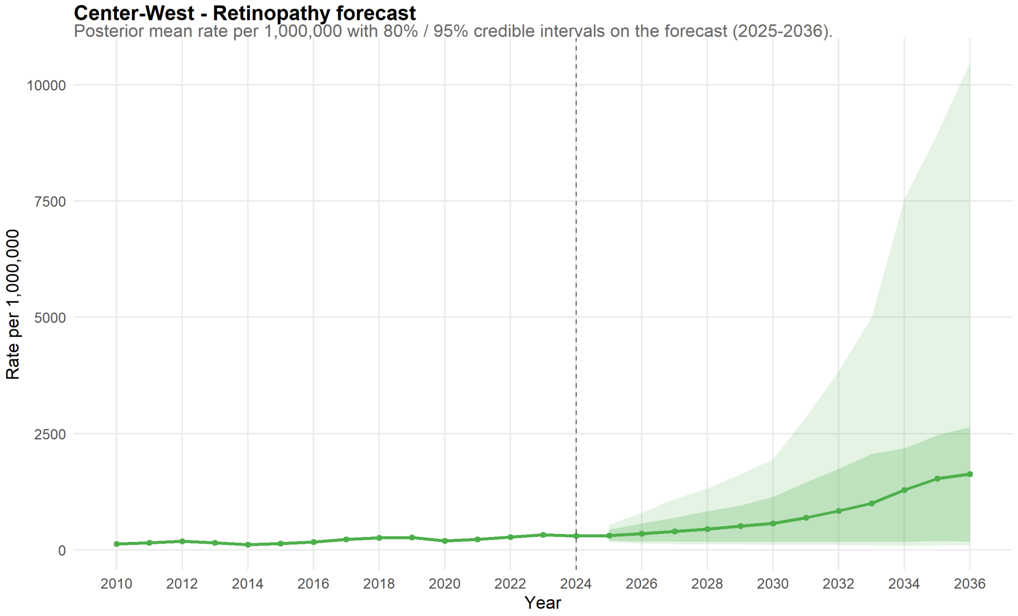

Figure 69: Center-West Forecast for Retinopathy.

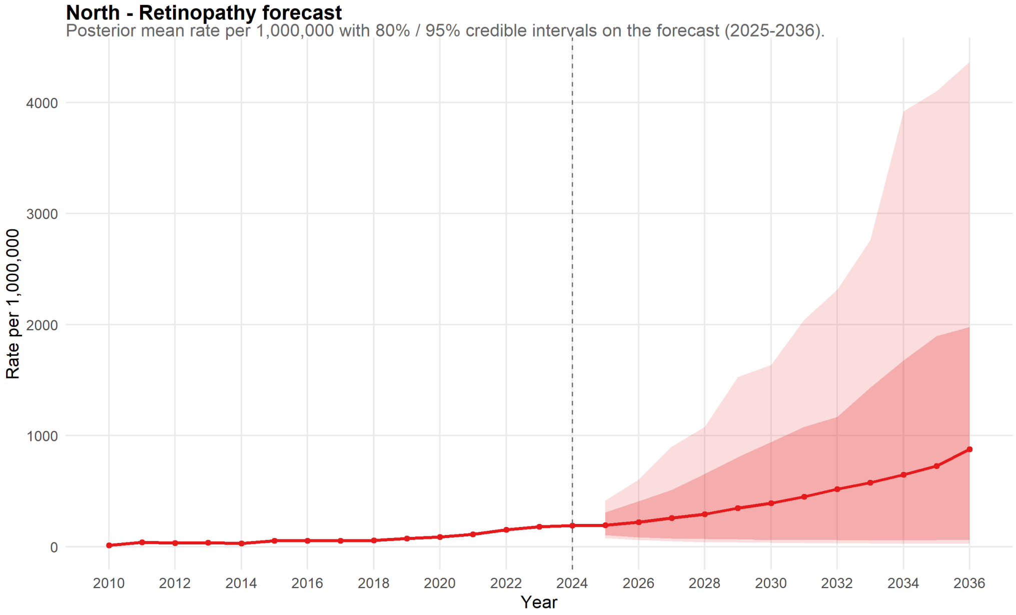

Figure 70: North Forecast for Retinopathy.

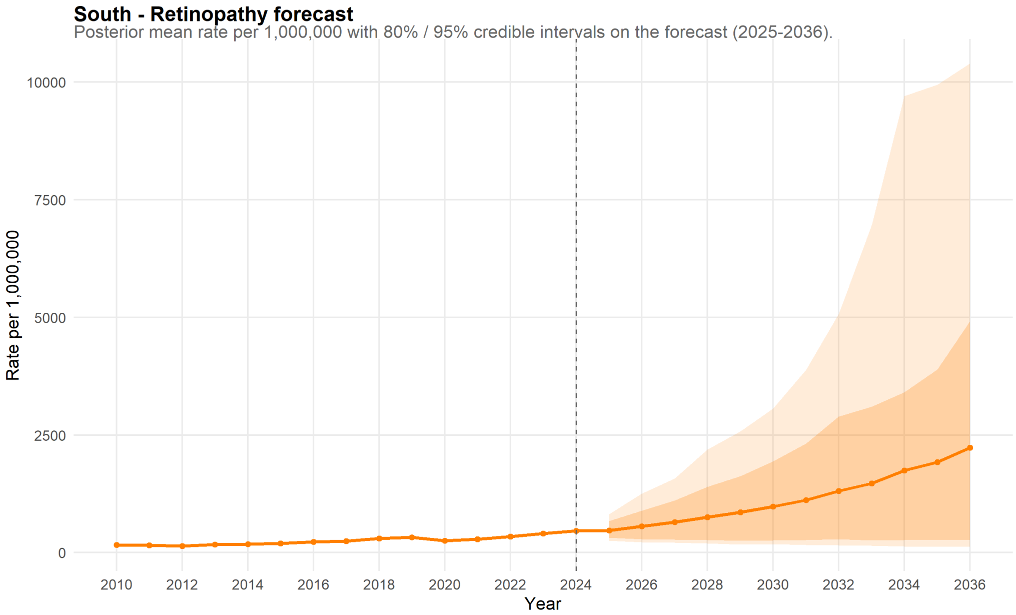

Figure 71: South Forecast for Retinopathy.

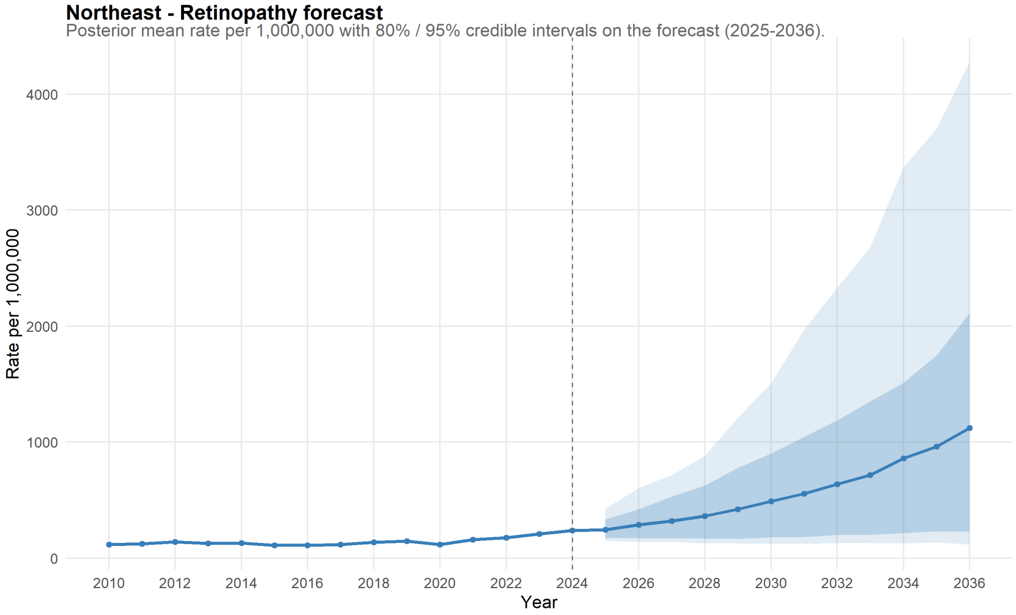

Figure 72: Northeast Forecast for Retinopathy.

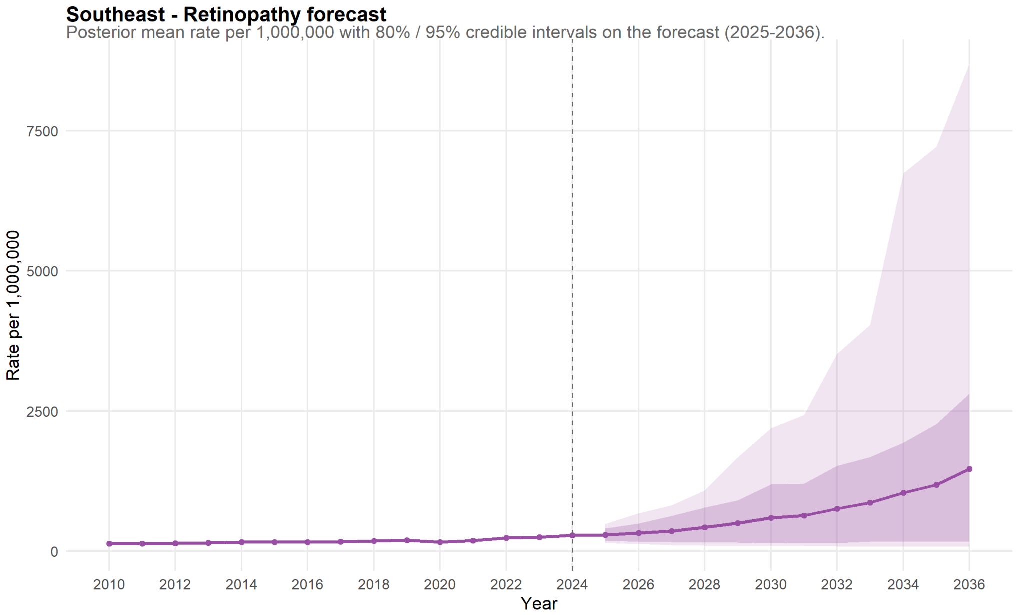

Figure 73: Southeast Forecast for Retinopathy.

##### 11.1.3 Eye & Appendage Diseases

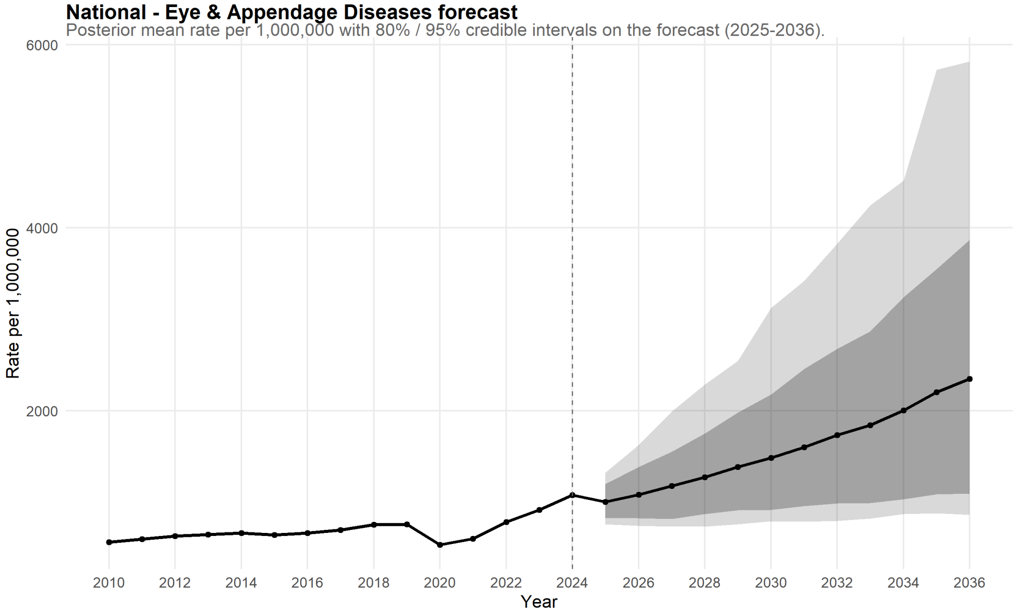

Figure 74: National Forecast for Eye & Appendage Diseases.

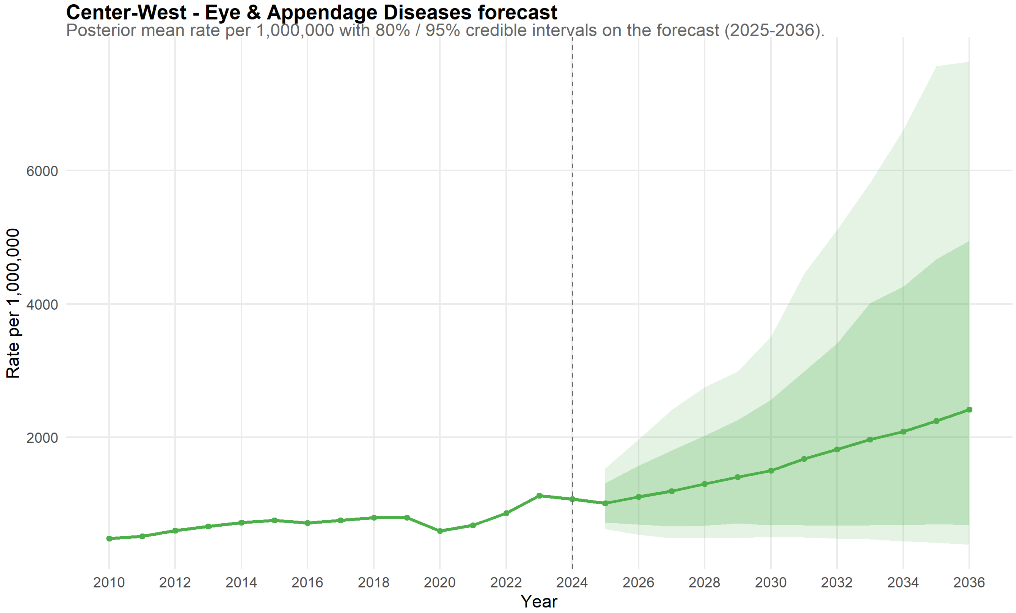

Figure 75: Center-West Forecast for Eye & Appendage Diseases.

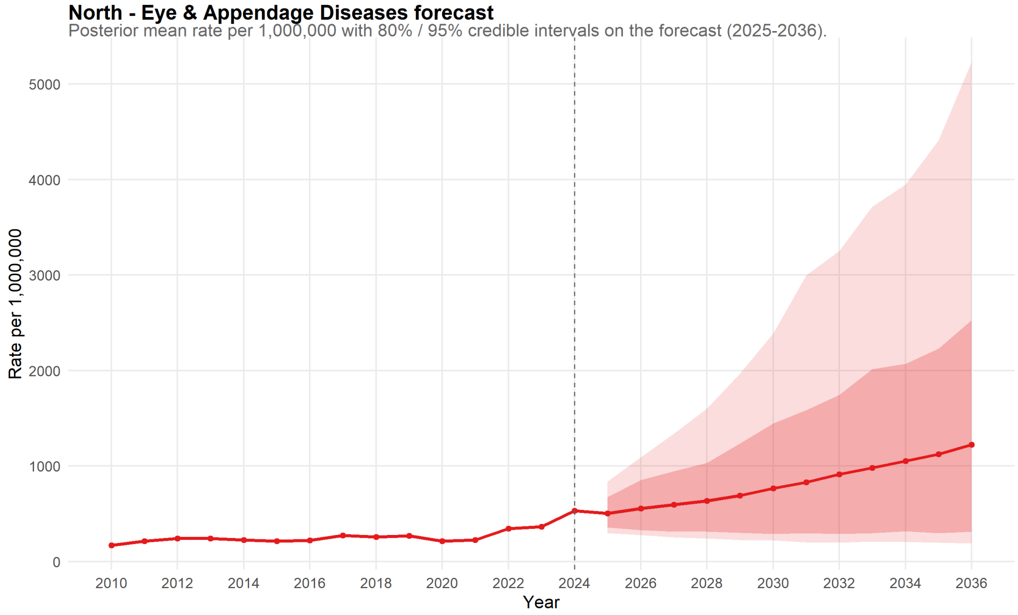

Figure 76: North Forecast for Eye & Appendage Diseases.

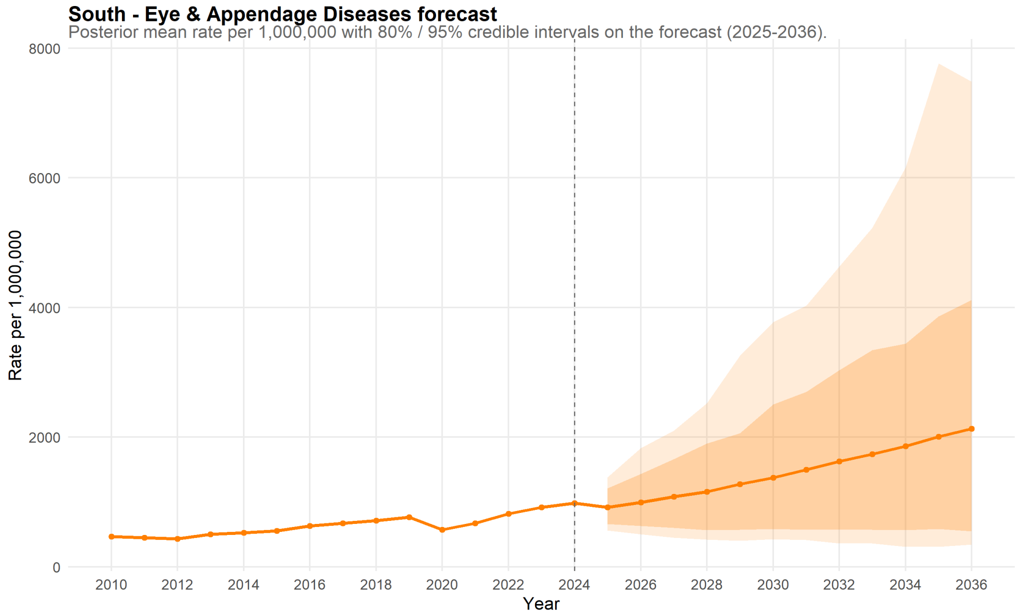

Figure 77: South Forecast for Eye & Appendage Diseases.

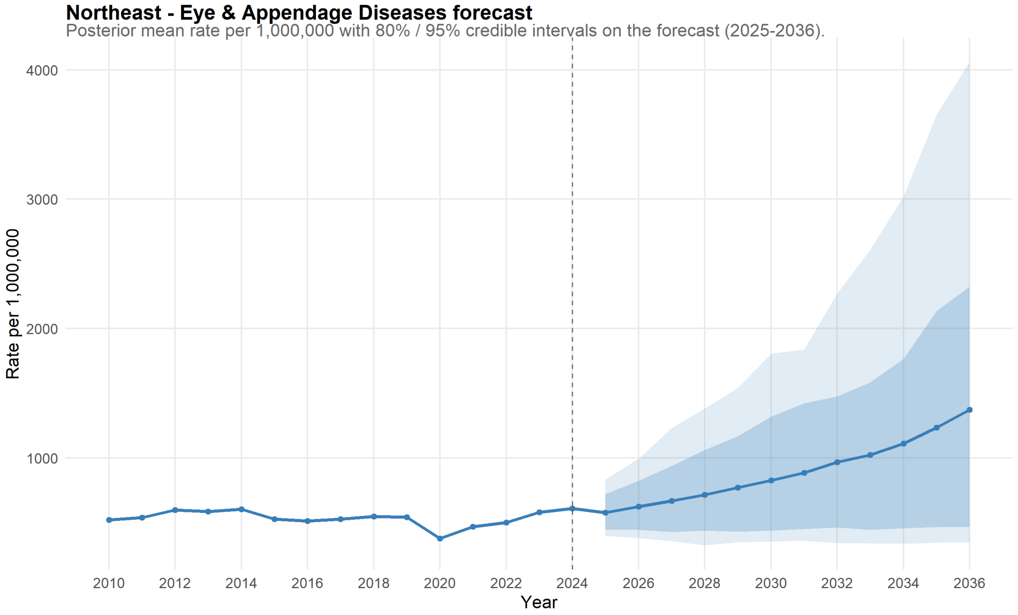

Figure 78: Northeast Forecast for Eye & Appendage Diseases.

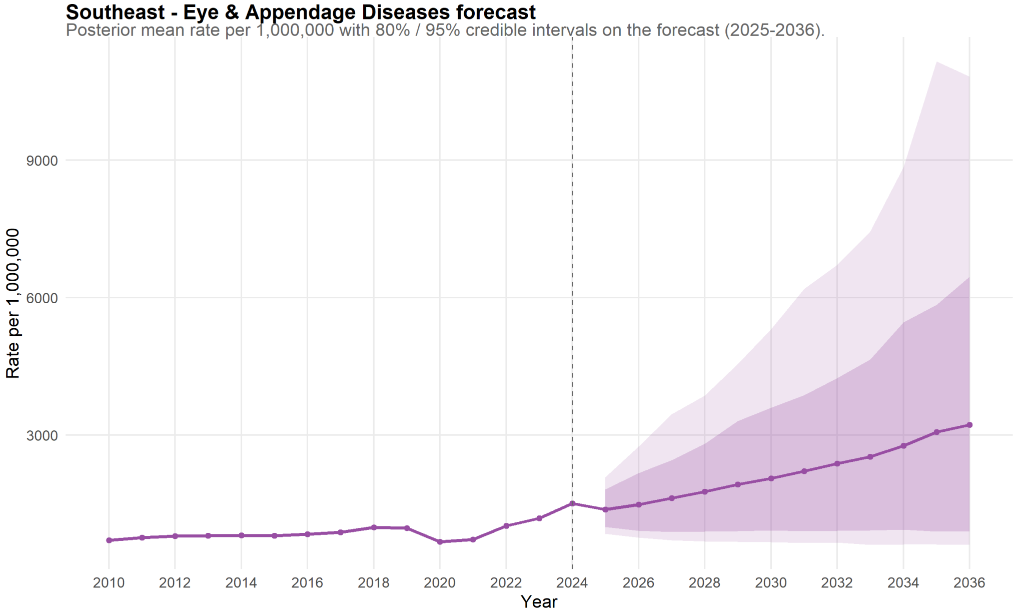

Figure 79: Southeast Forecast for Eye & Appendage Diseases.

#### 11.2 State-level Forecasts

##### 11.2.1 Glaucoma

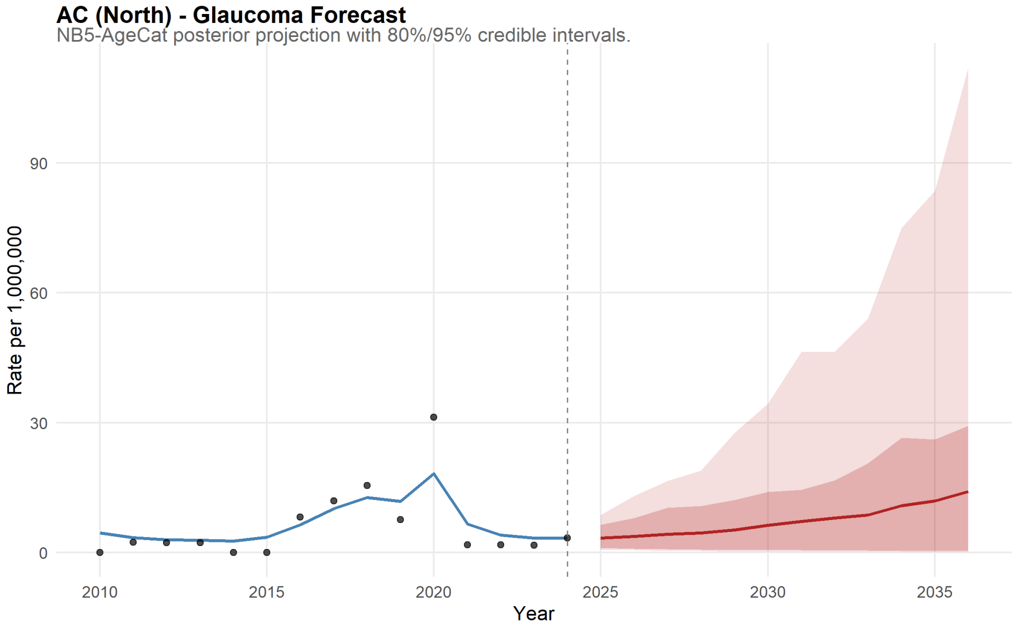

Figure 80: Acre Forecast for Glaucoma.

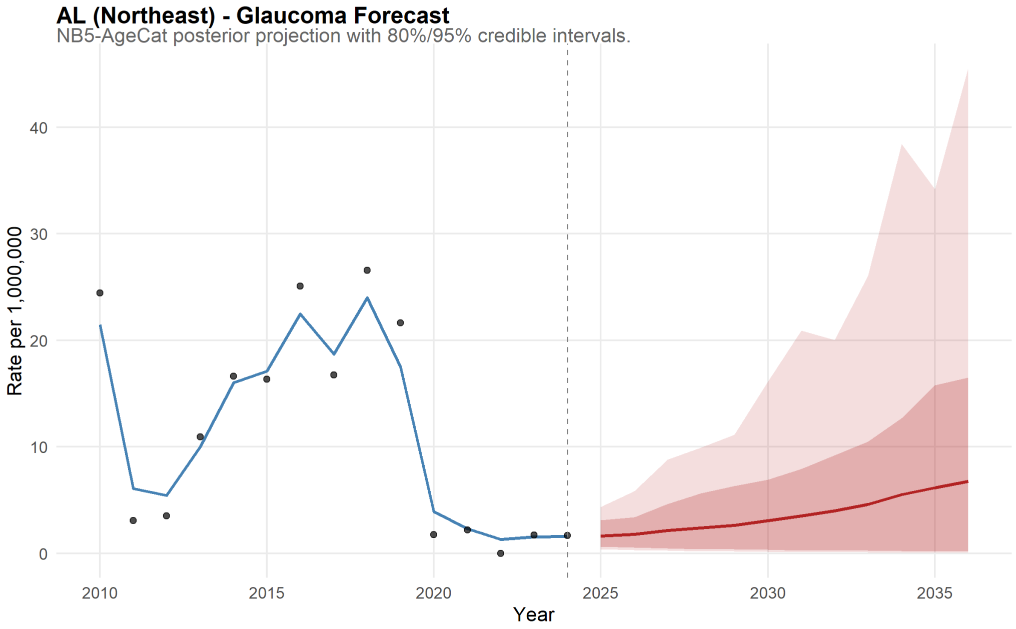

Figure 81: Alagoas Forecast for Glaucoma.

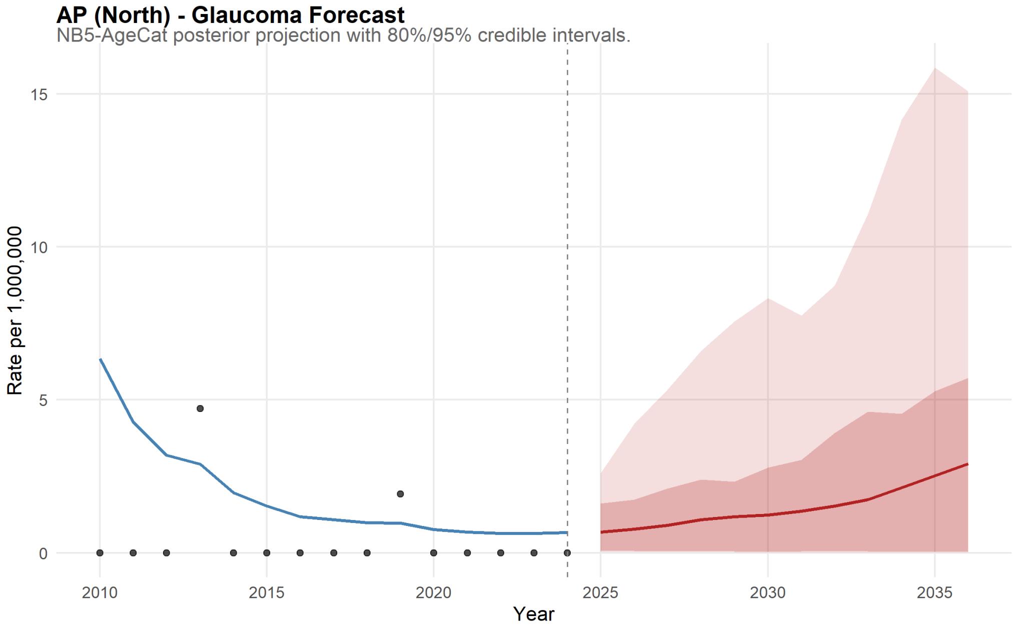

Figure 82: Amapá Forecast for Glaucoma.

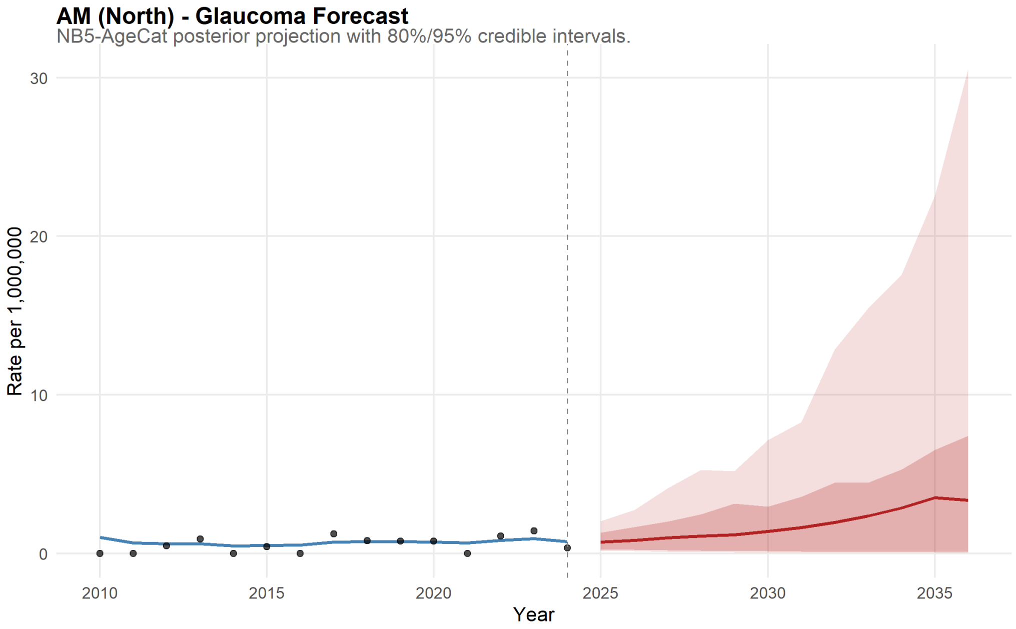

Figure 83: Amazonas Forecast for Glaucoma.

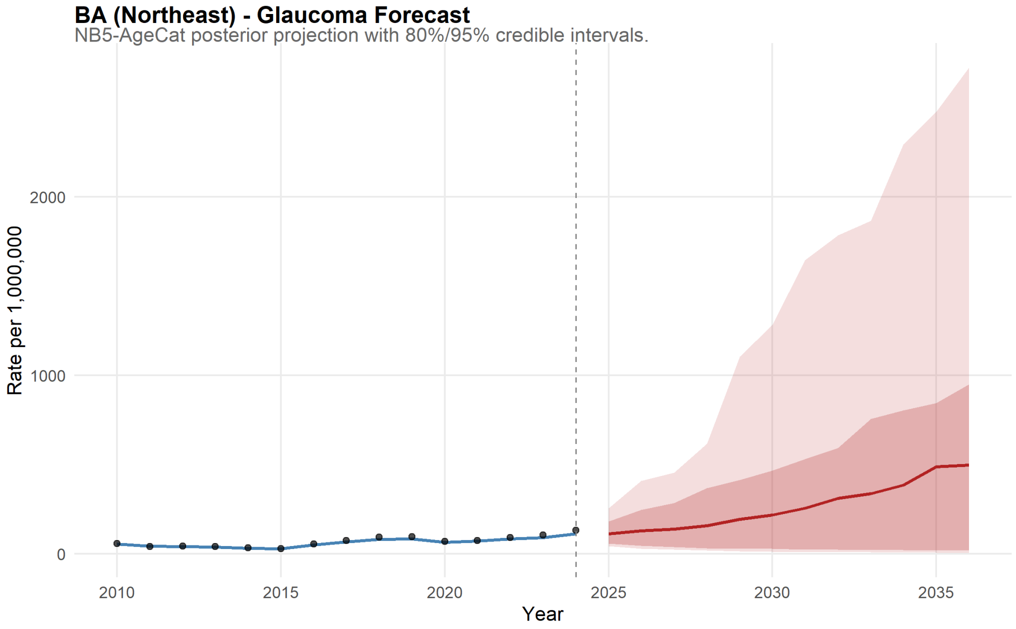

Figure 84: Bahia Forecast for Glaucoma.

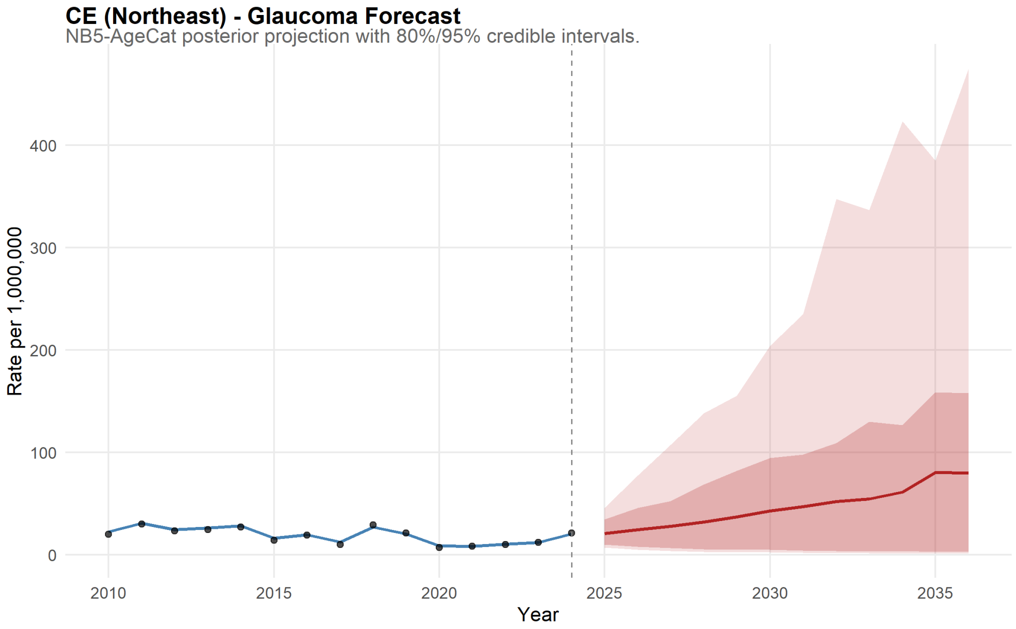

Figure 85: Ceará Forecast for Glaucoma.

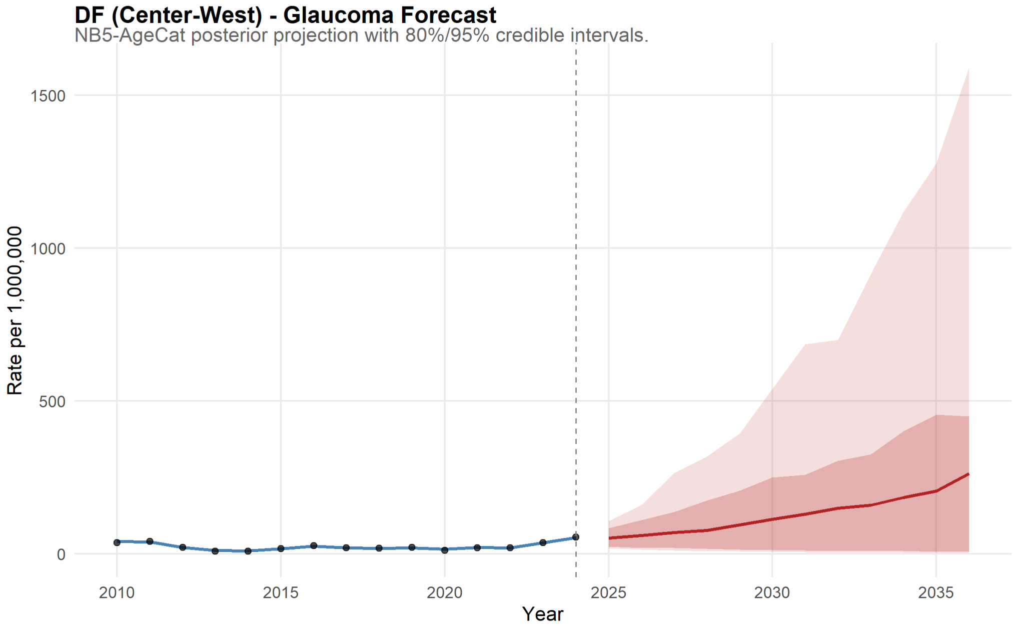

Figure 86: Distrito Federal Forecast for Glaucoma.

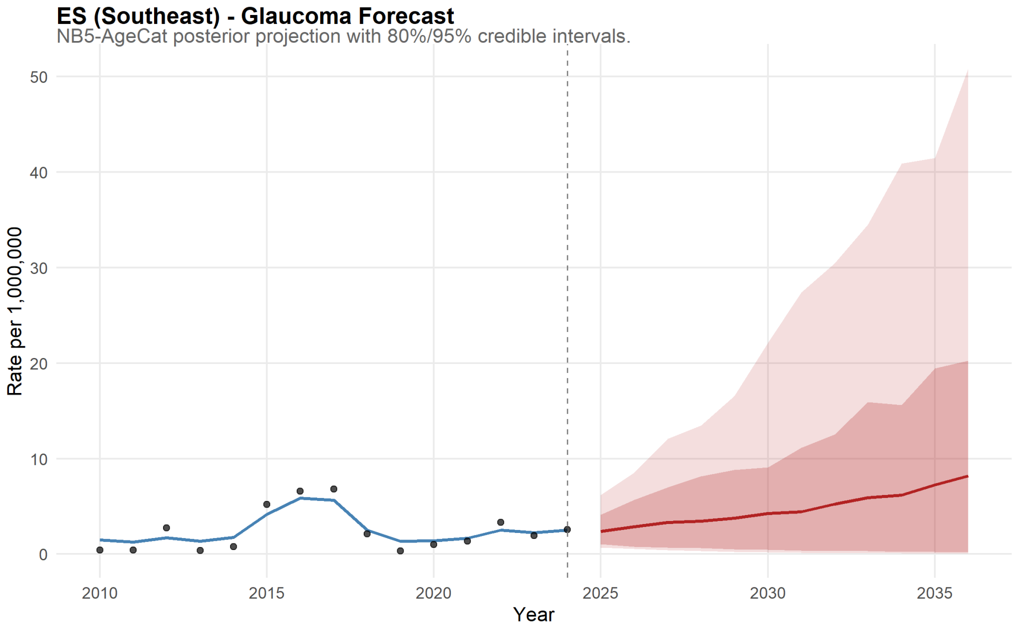

Figure 87: Espírito Santo Forecast for Glaucoma.

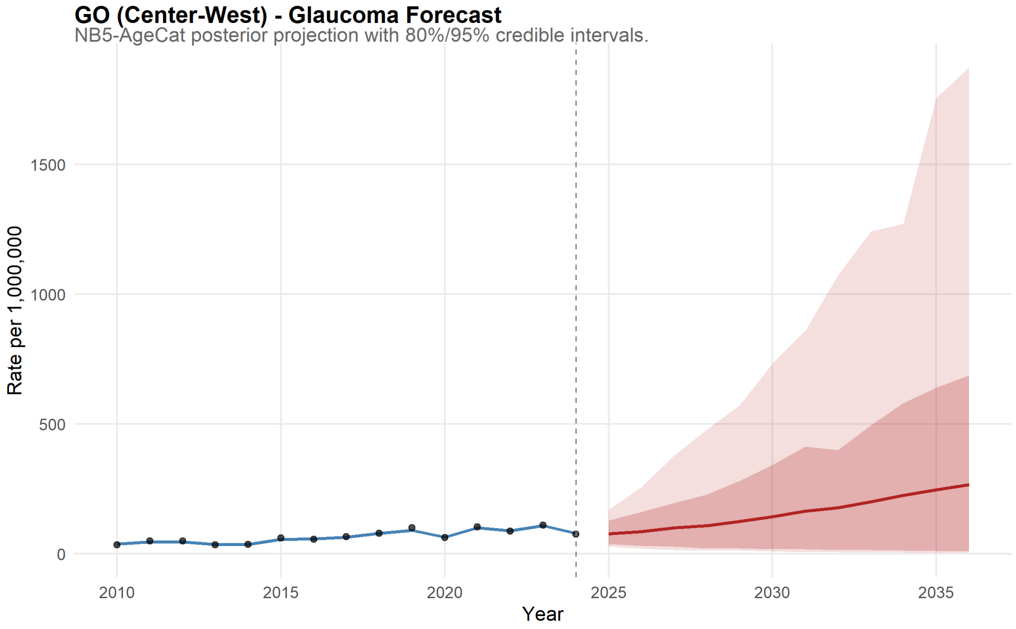

Figure 88: Goiás Forecast for Glaucoma.

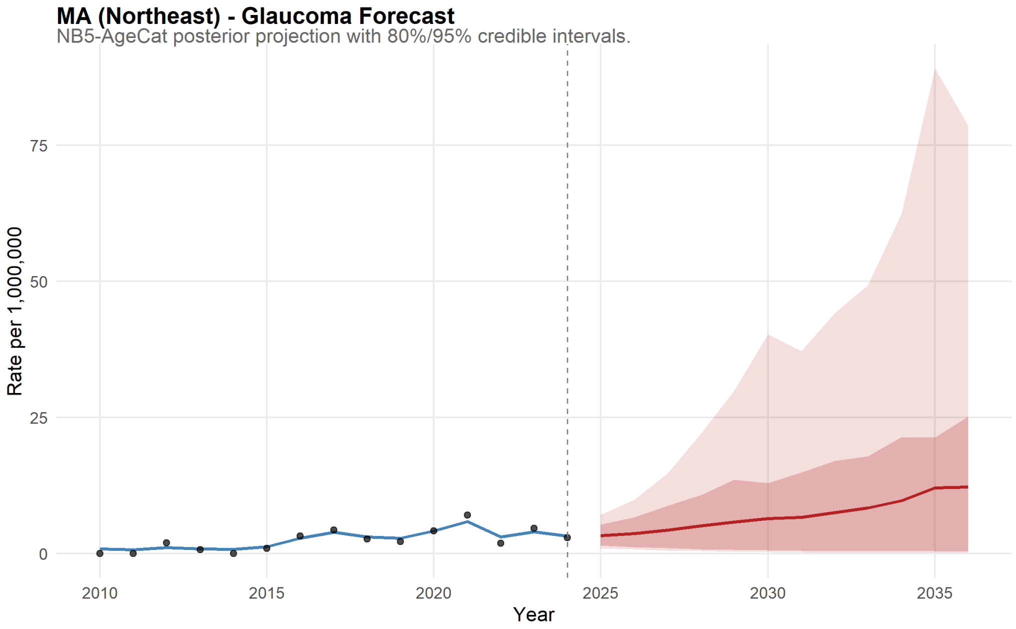

Figure 89: Maranhão Forecast for Glaucoma.

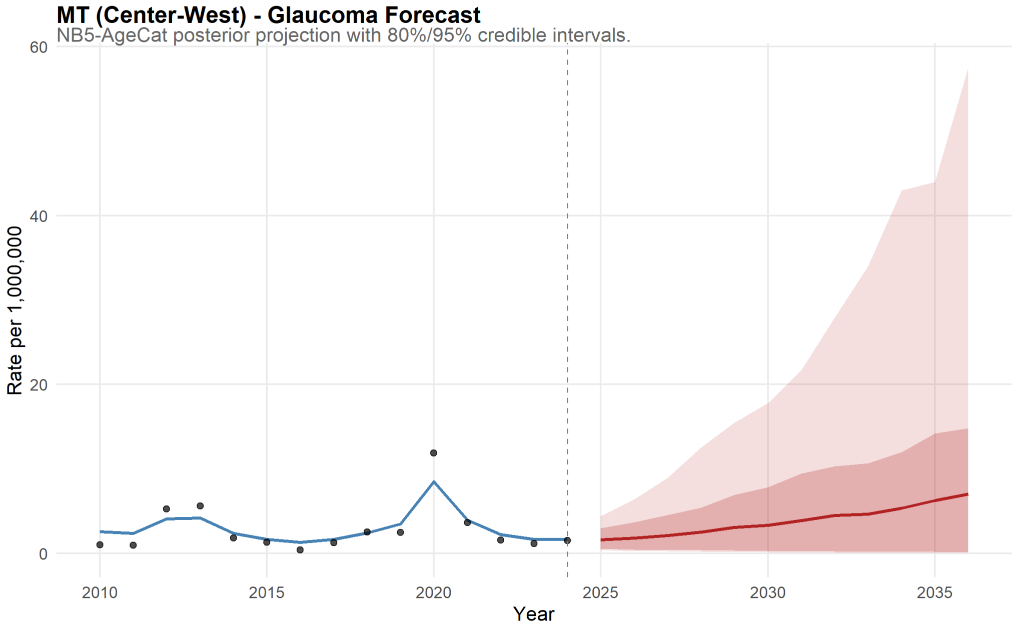

Figure 90: Mato Grosso Forecast for Glaucoma.

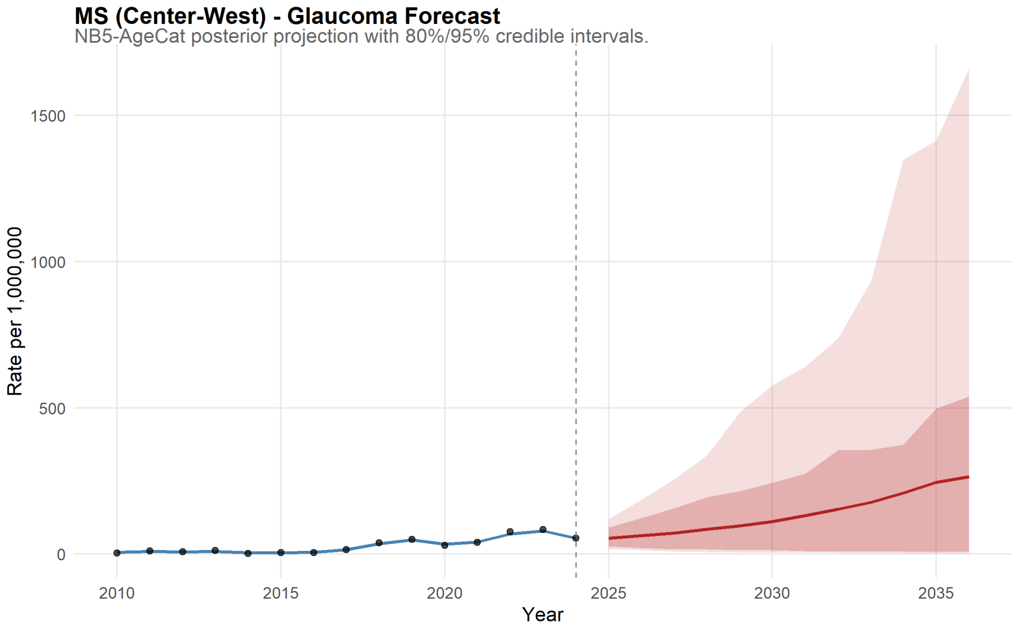

Figure 91: Mato Grosso do Sul Forecast for Glaucoma.

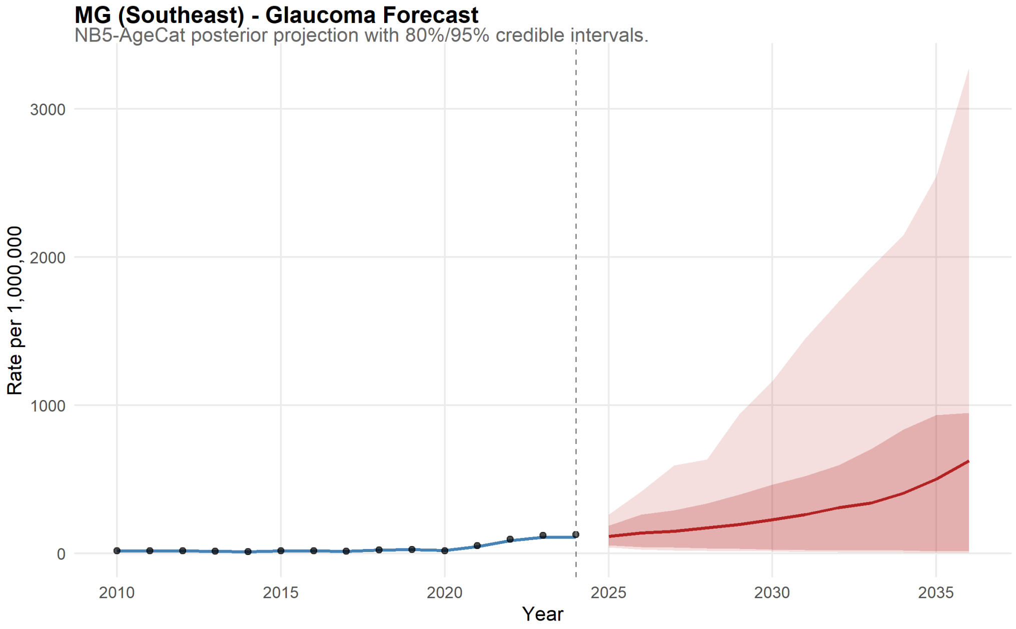

Figure 92: Minas Gerais Forecast for Glaucoma.

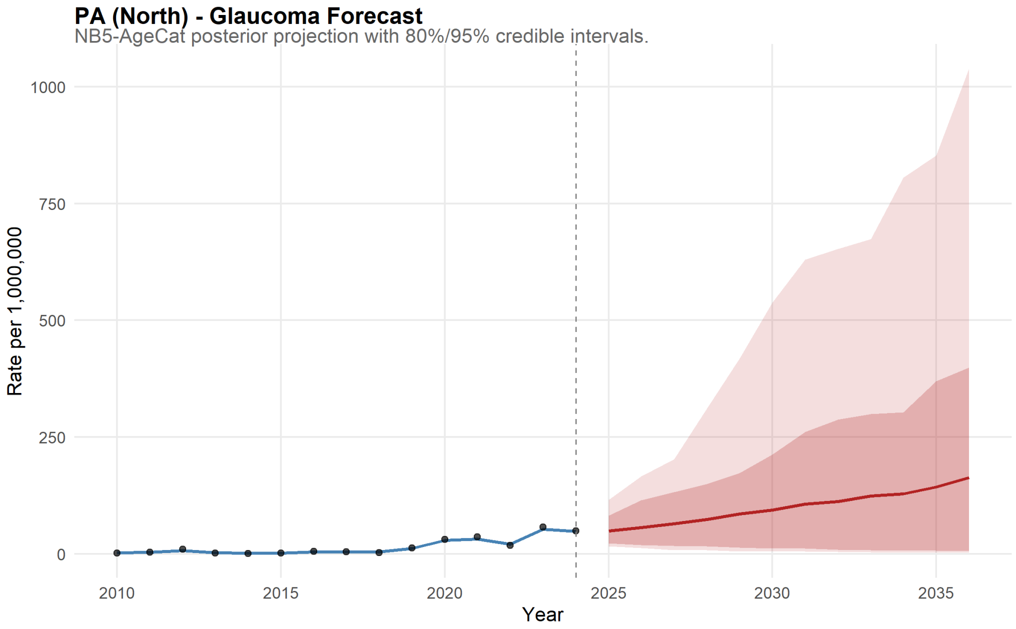

Figure 93: Pará Forecast for Glaucoma.

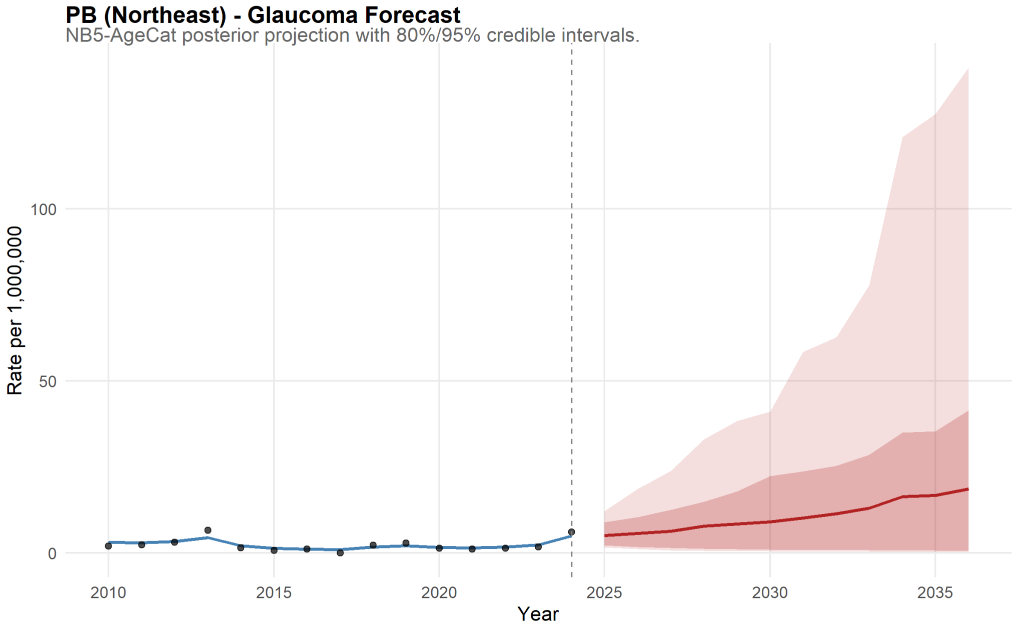

Figure 94: Paraíba Forecast for Glaucoma.

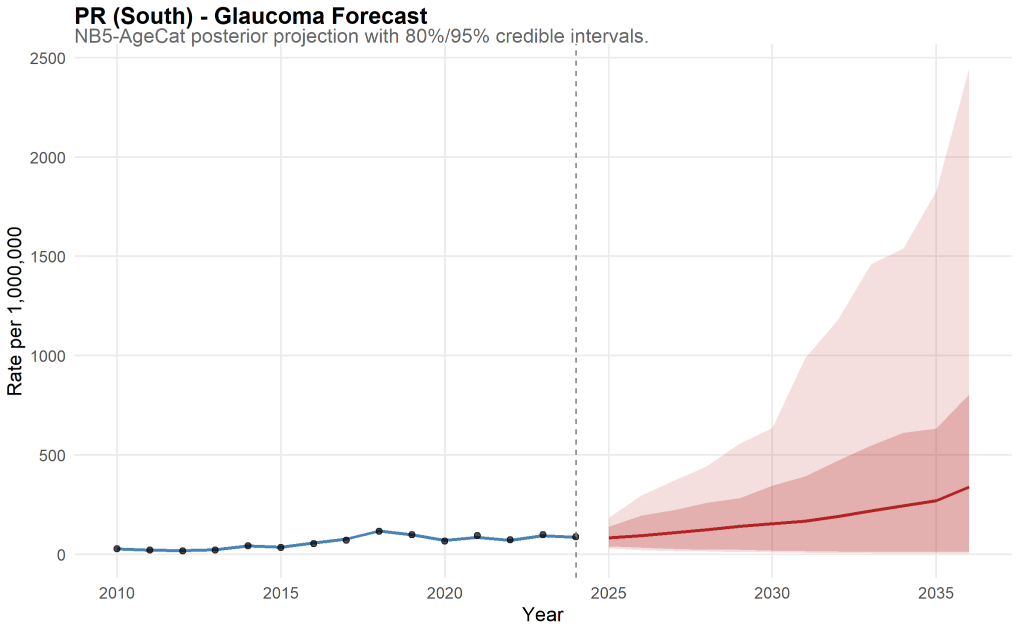

Figure 95: Paraná Forecast for Glaucoma.

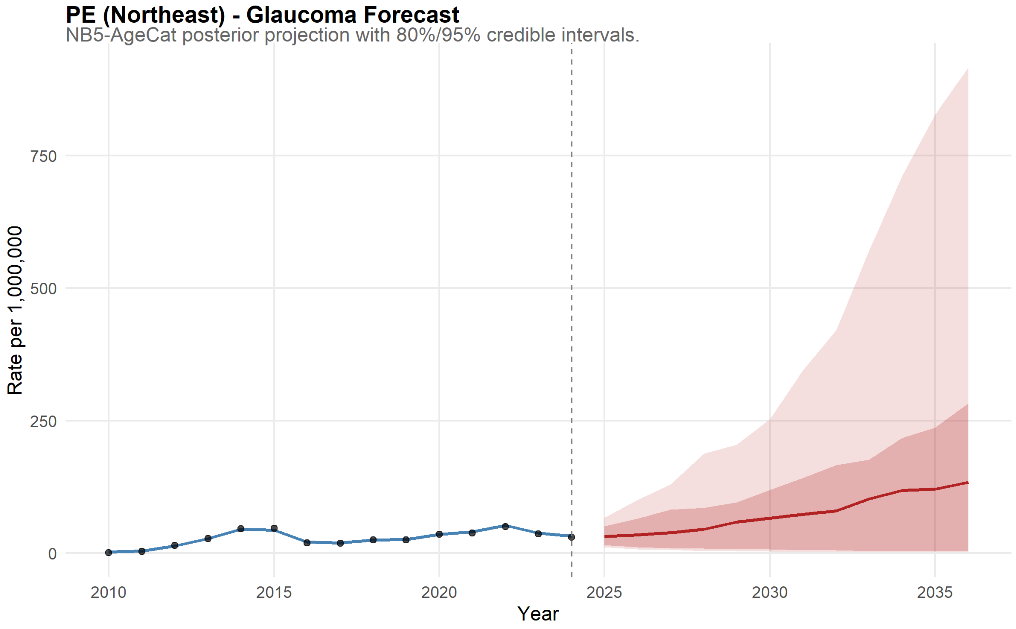

Figure 96: Pernambuco Forecast for Glaucoma.

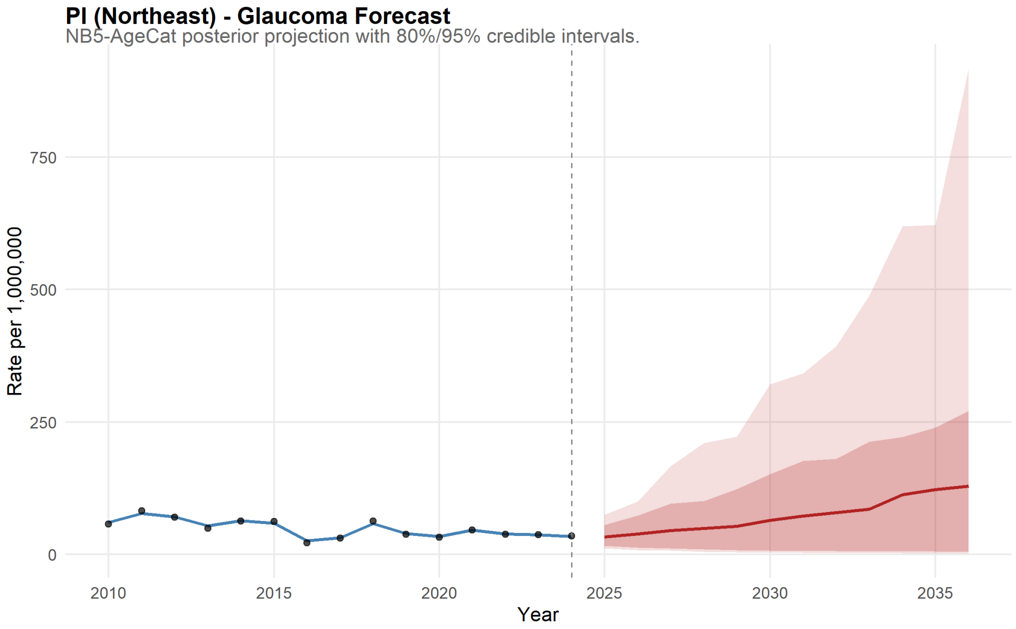

Figure 97: Piauí Forecast for Glaucoma.

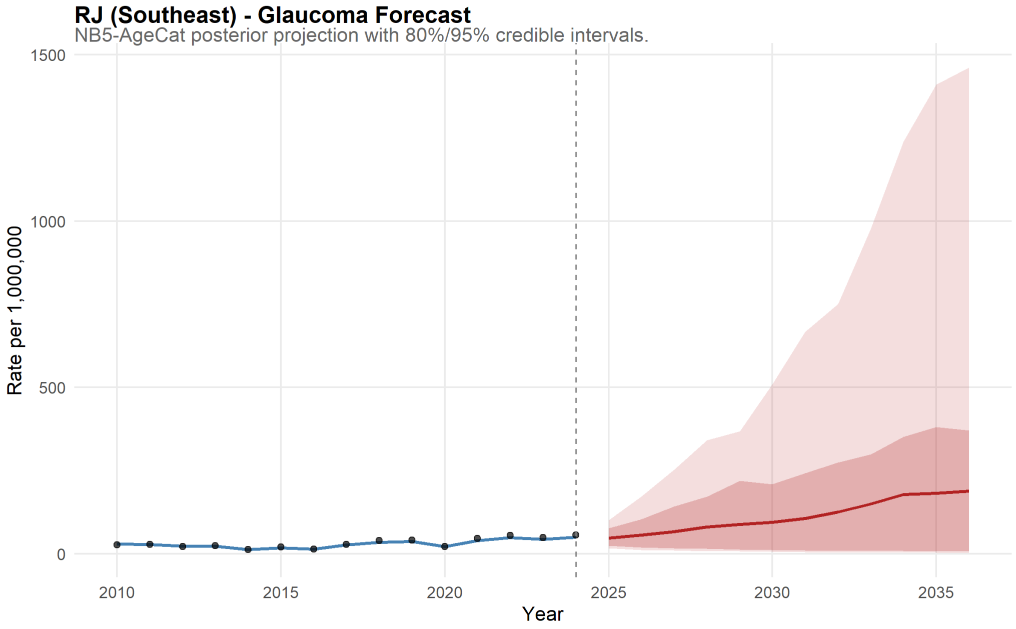

Figure 98: Rio de Janeiro Forecast for Glaucoma.

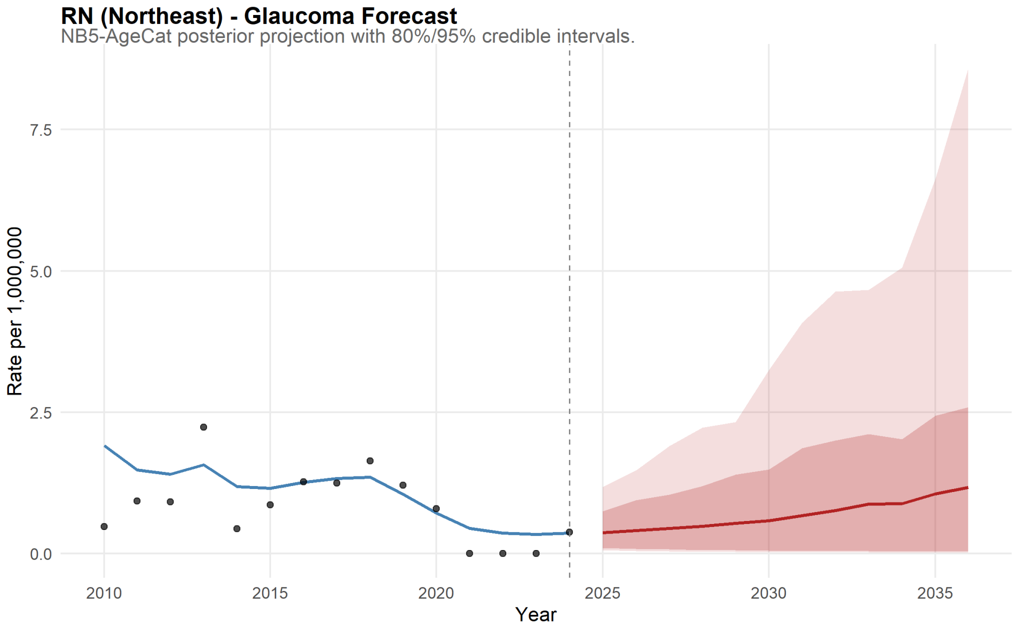

Figure 99: Rio Grande do Norte Forecast for Glaucoma.

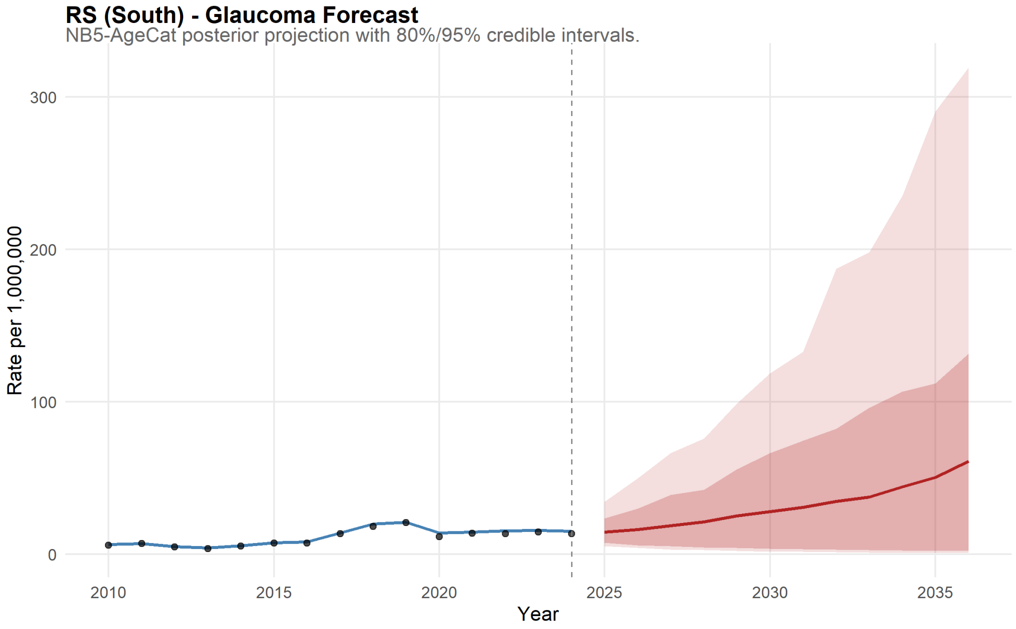

Figure 100: Rio Grande do Sul Forecast for Glaucoma.

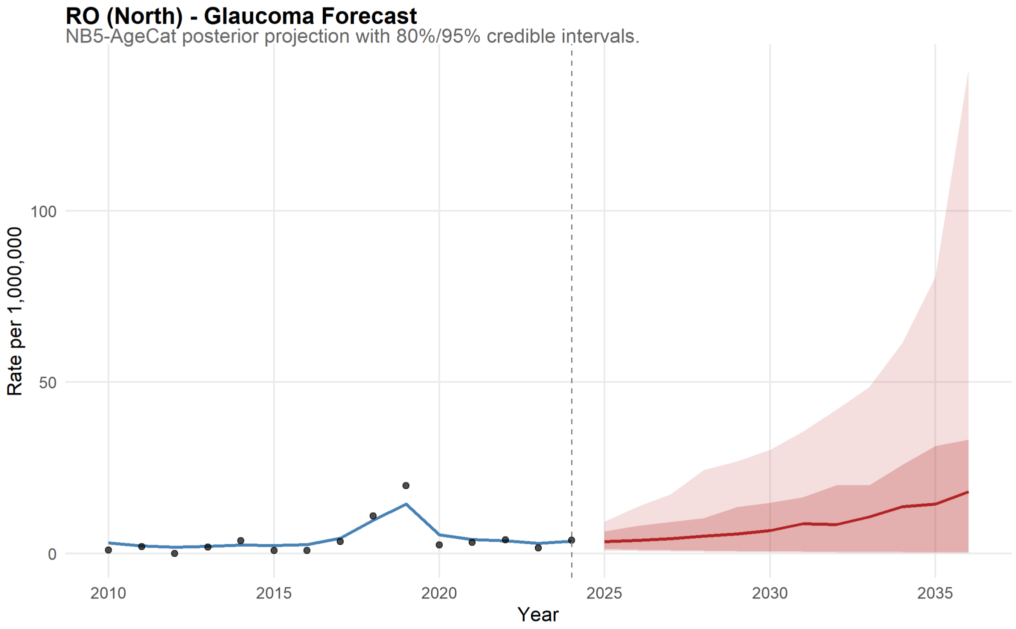

Figure 101: Rondônia Forecast for Glaucoma.

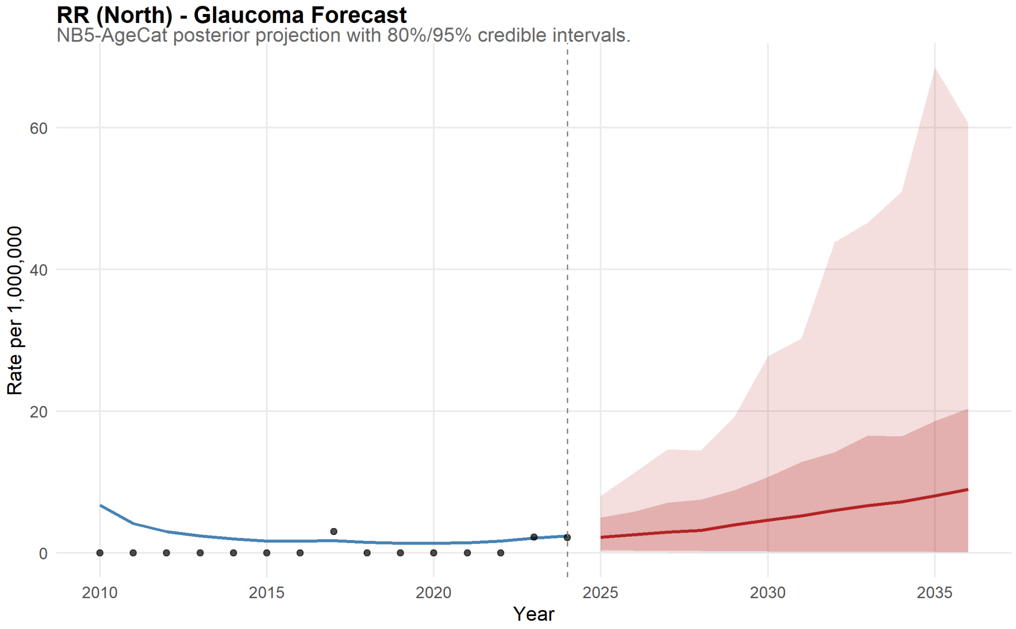

Figure 102: Roraima Forecast for Glaucoma.

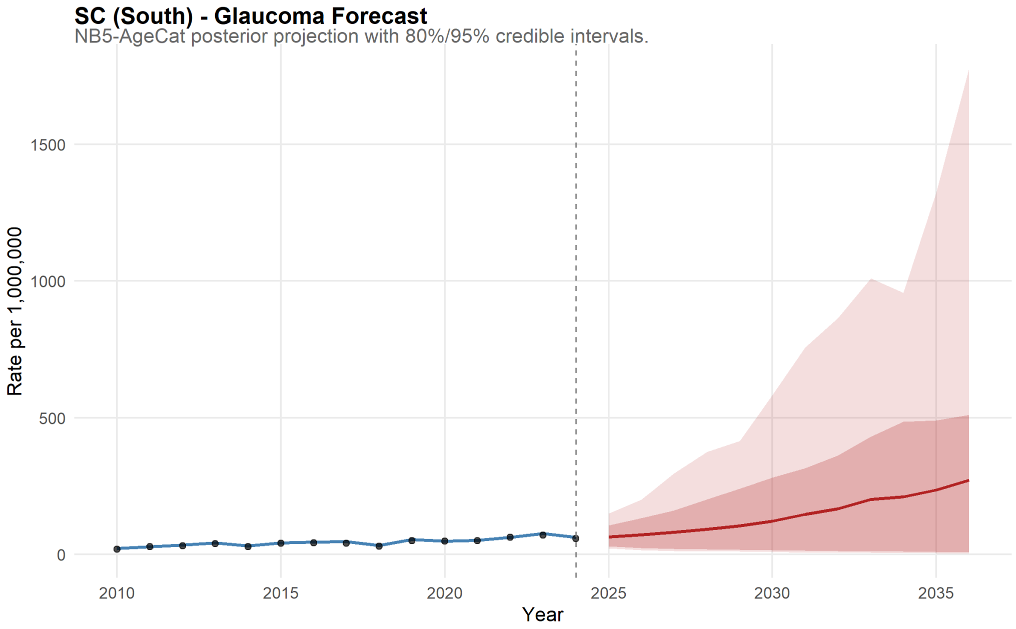

Figure 103: Santa Catarina Forecast for Glaucoma.

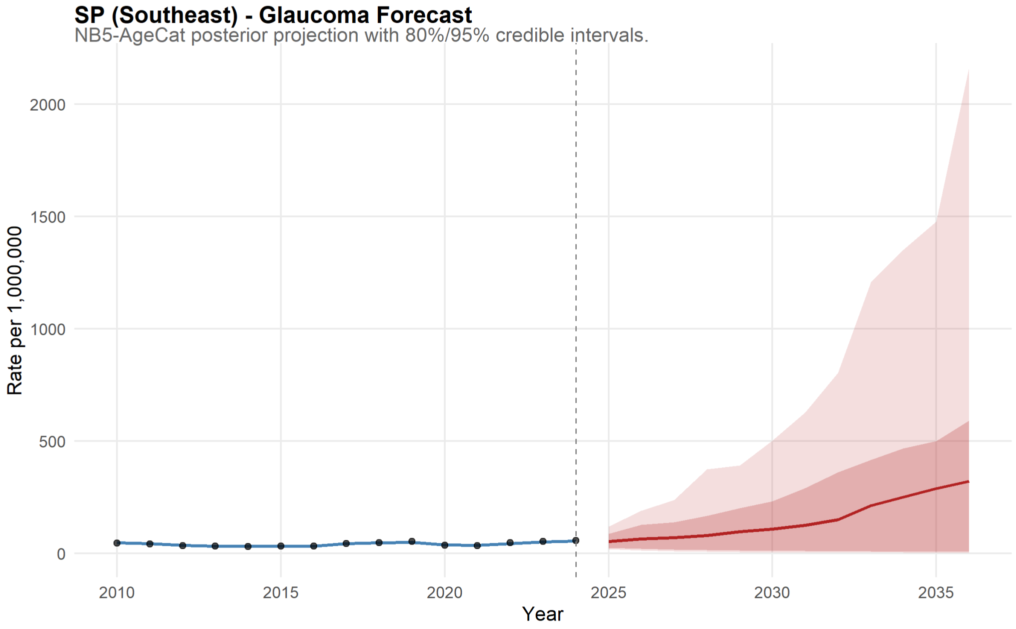

Figure 104: São Paulo Forecast for Glaucoma.

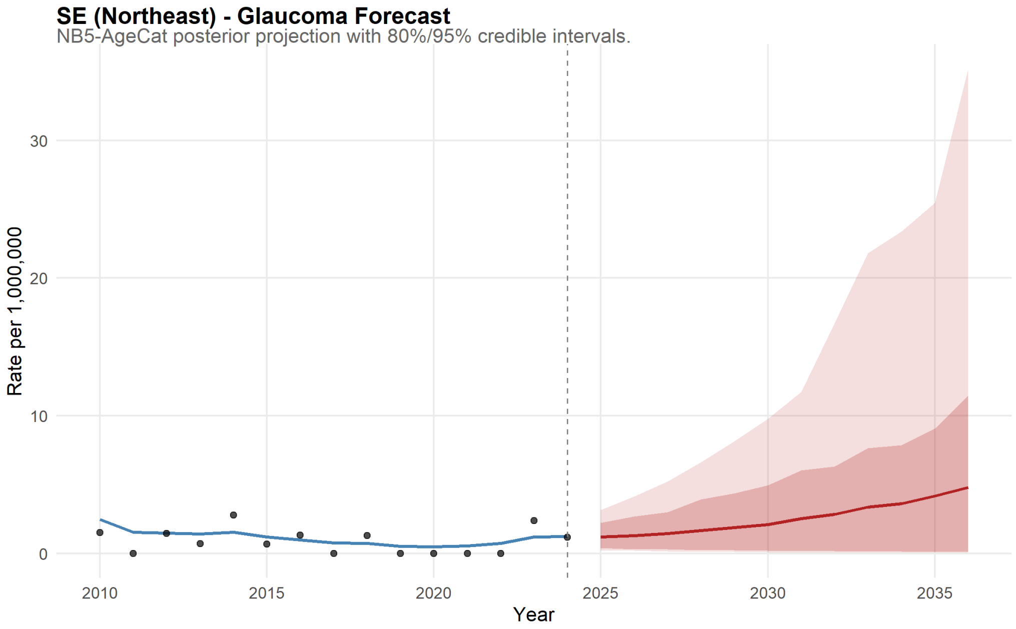

Figure 105: Sergipe Forecast for Glaucoma.

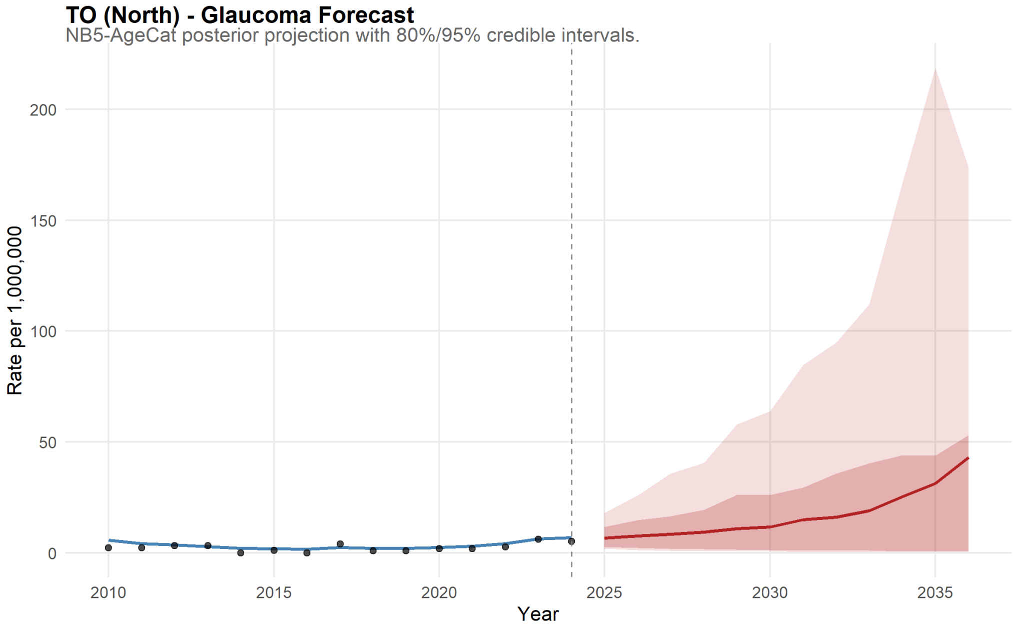

Figure 106: Tocantins Forecast for Glaucoma.

##### 11.2.2 Retinopathy

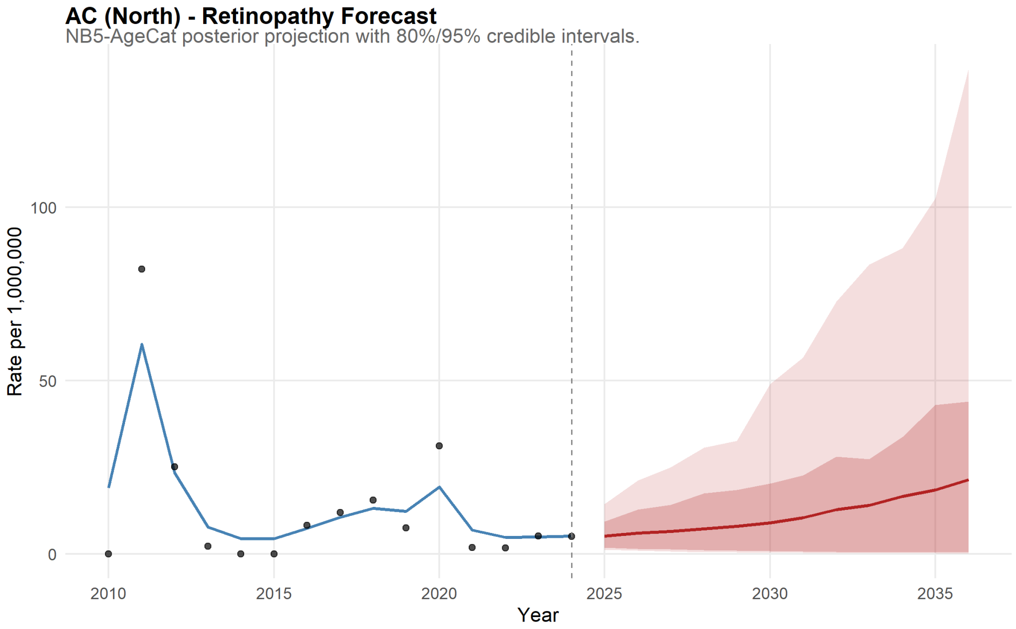

Figure 107: Acre Forecast for Retinopathy.

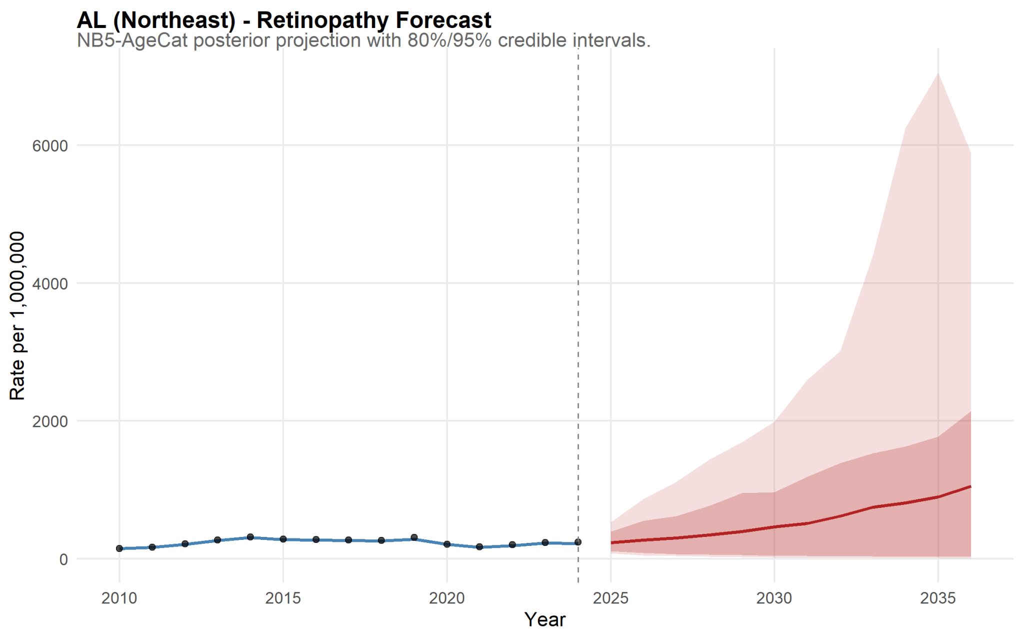

Figure 108: Alagoas Forecast for Retinopathy.

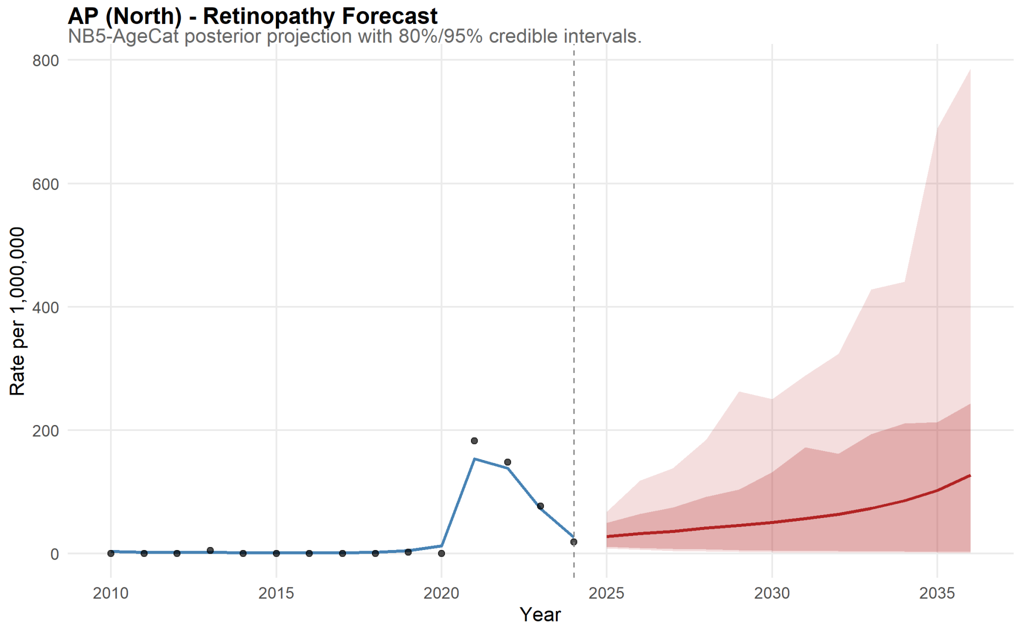

Figure 109: Amapá Forecast for Retinopathy.

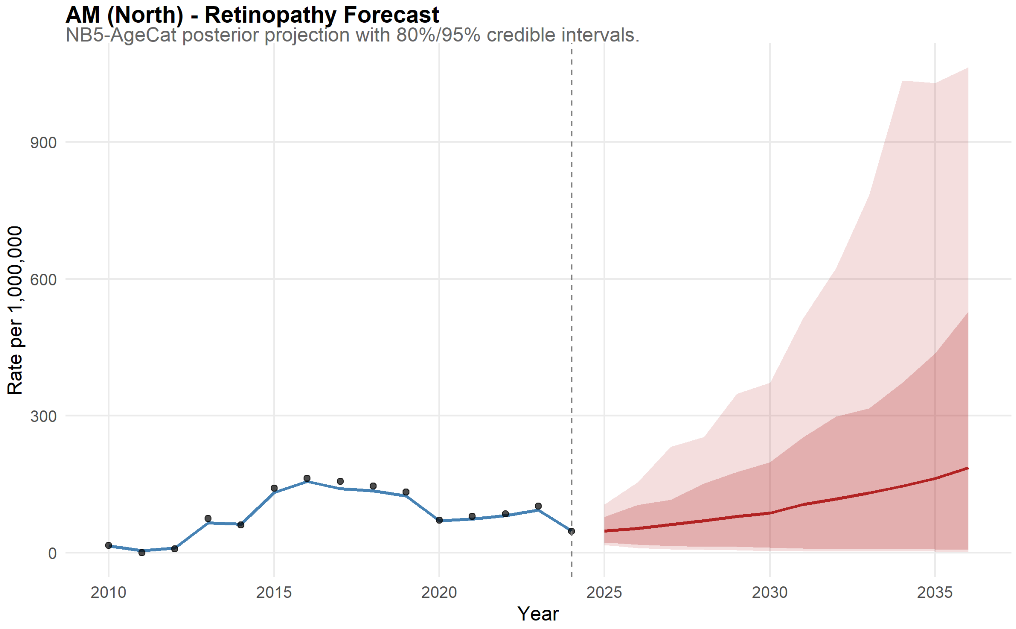

Figure 110: Amazonas Forecast for Retinopathy.

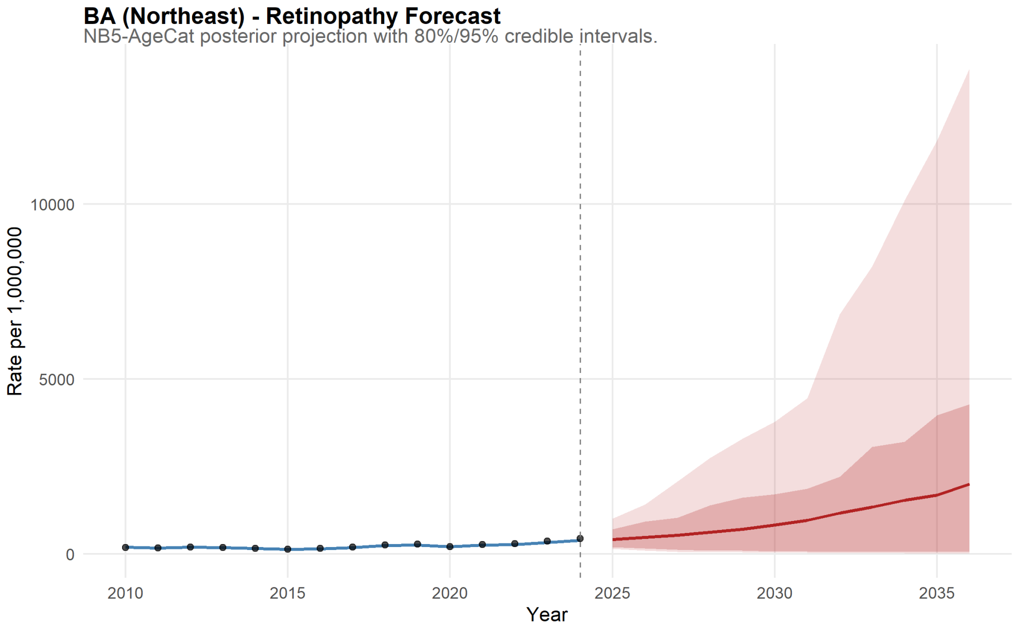

Figure 111: Bahia Forecast for Retinopathy.

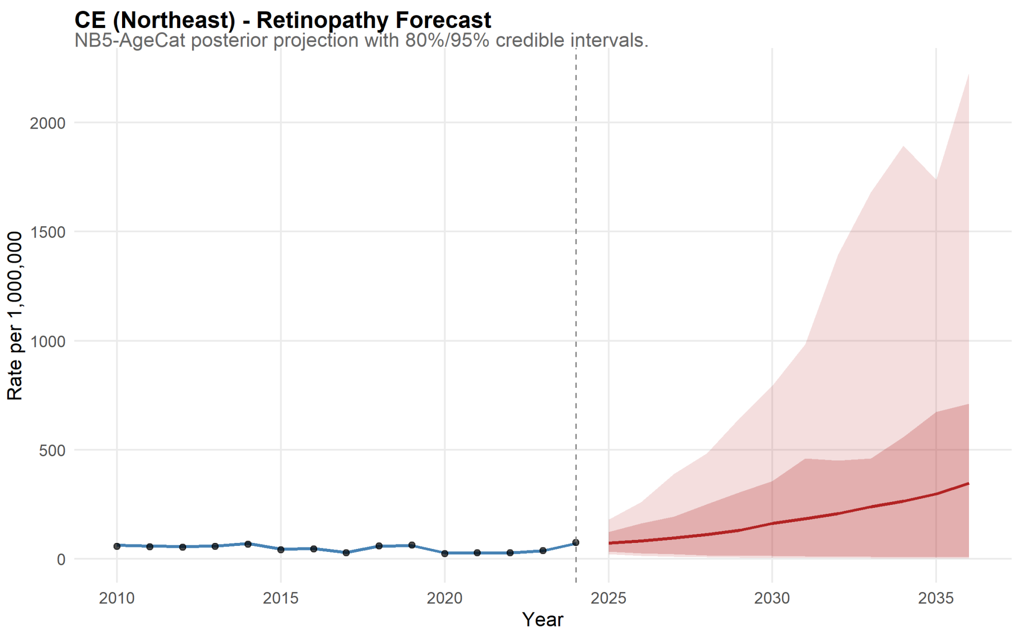

Figure 112: Ceará Forecast for Retinopathy.

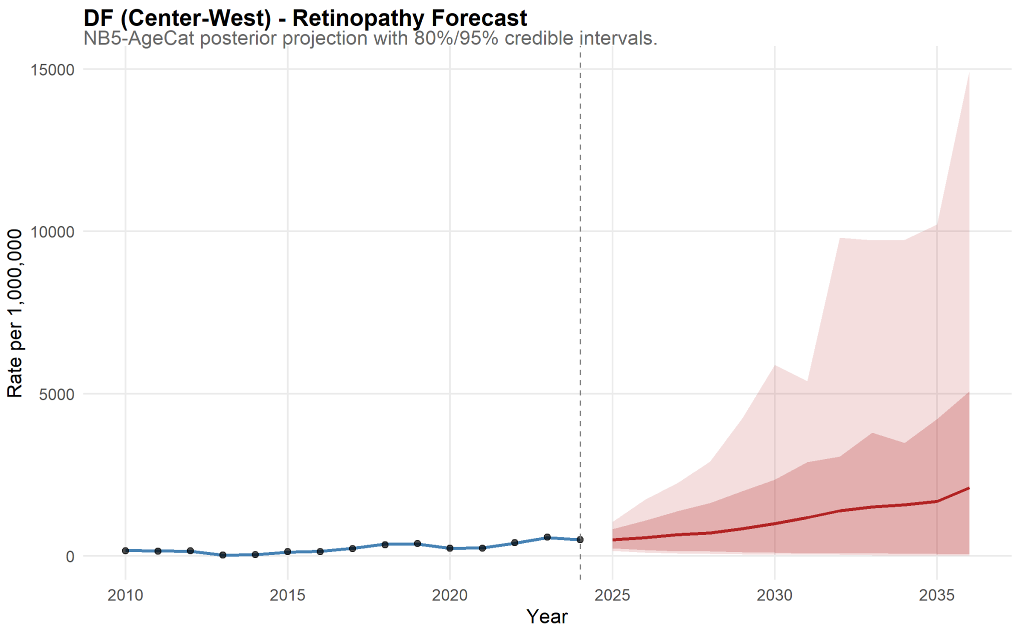

Figure 113: Distrito Federal Forecast for Retinopathy.

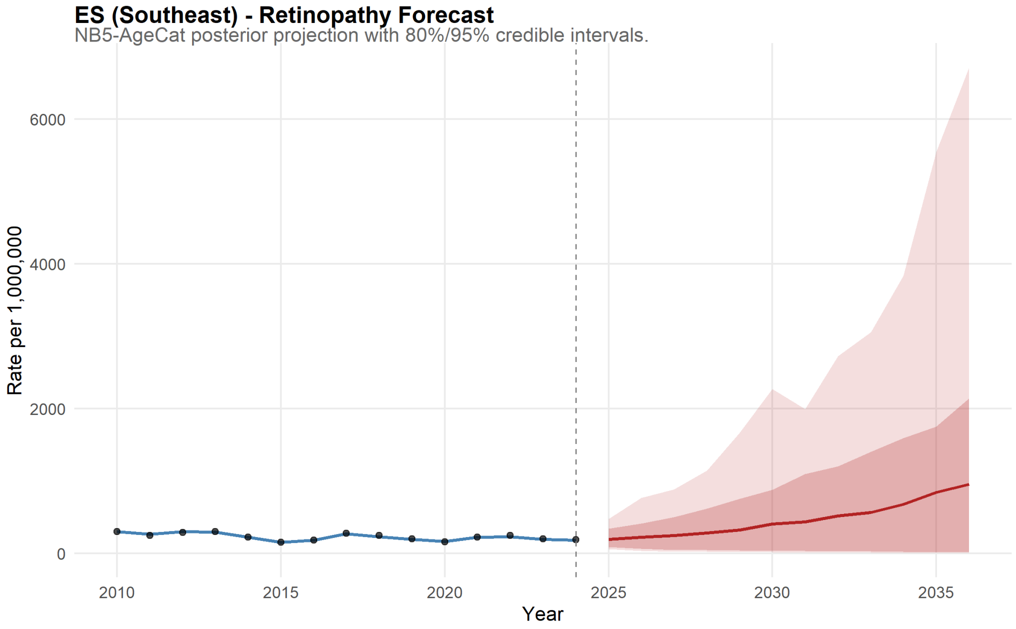

Figure 114: Espírito Santo Forecast for Retinopathy.

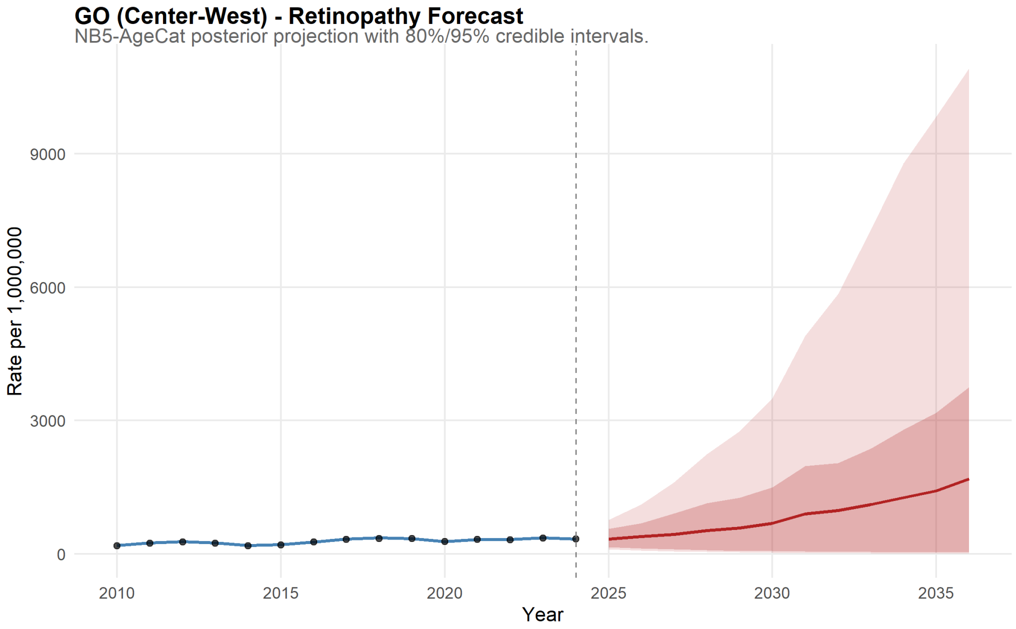

Figure 115: Goiás Forecast for Retinopathy.

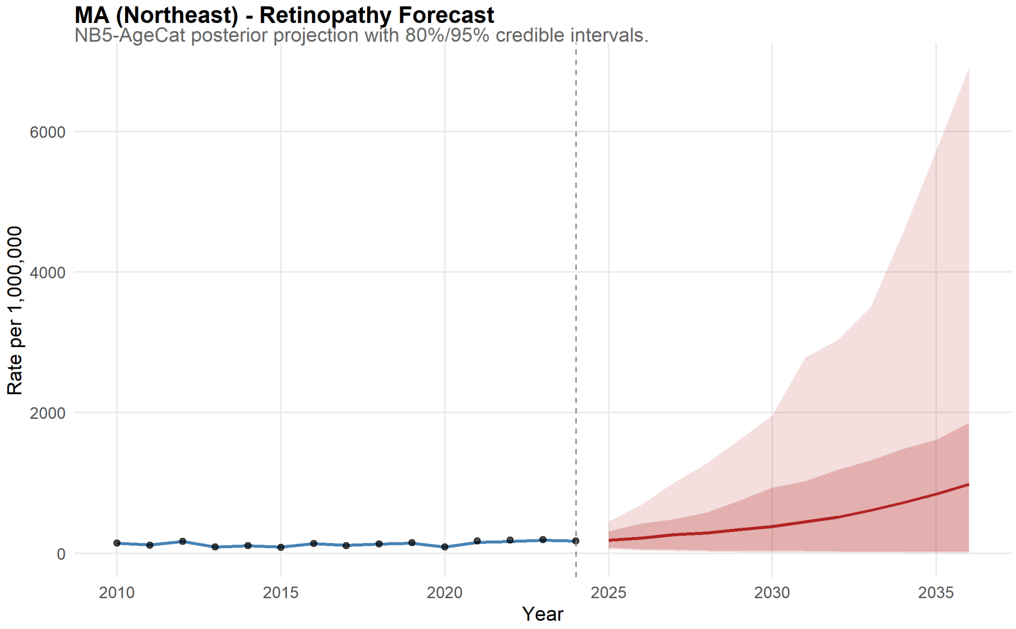

Figure 116: Maranhão Forecast for Retinopathy.

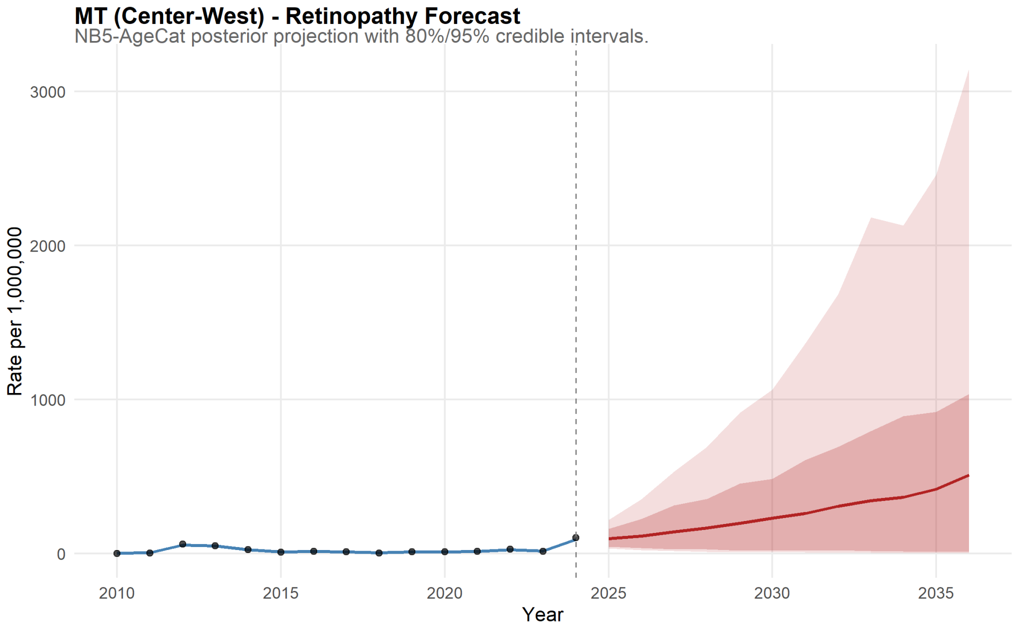

Figure 117: Mato Grosso Forecast for Retinopathy.

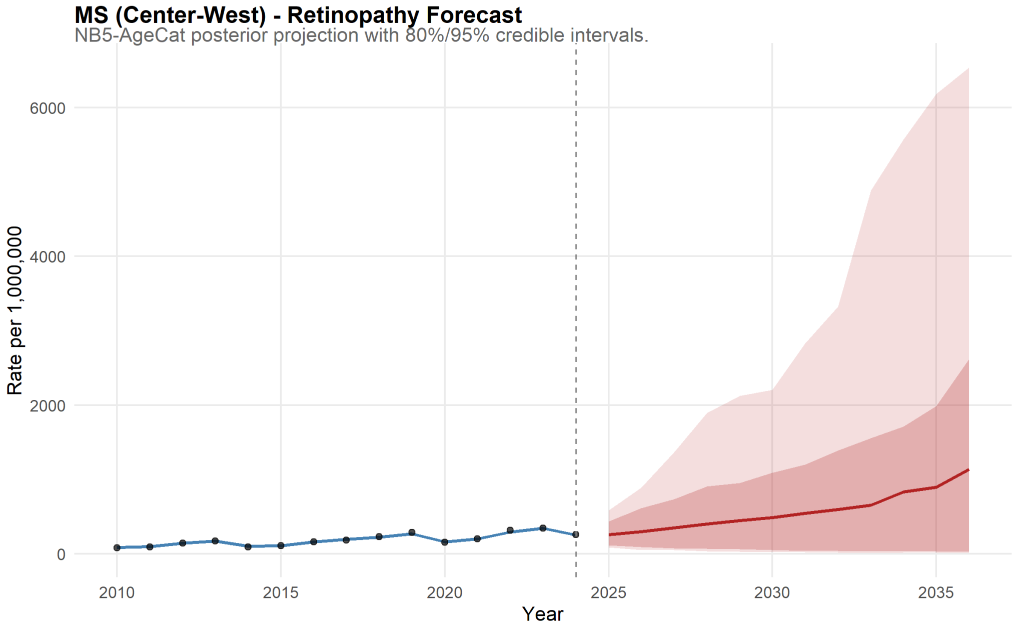

Figure 118: Mato Grosso do Sul Forecast for Retinopathy.

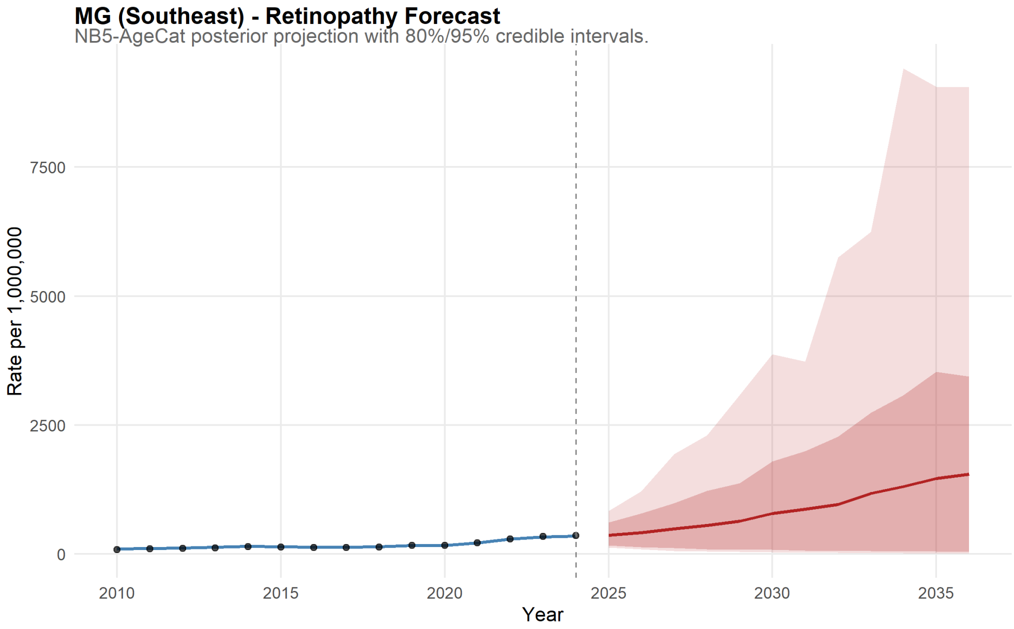

Figure 119: Minas Gerais Forecast for Retinopathy.

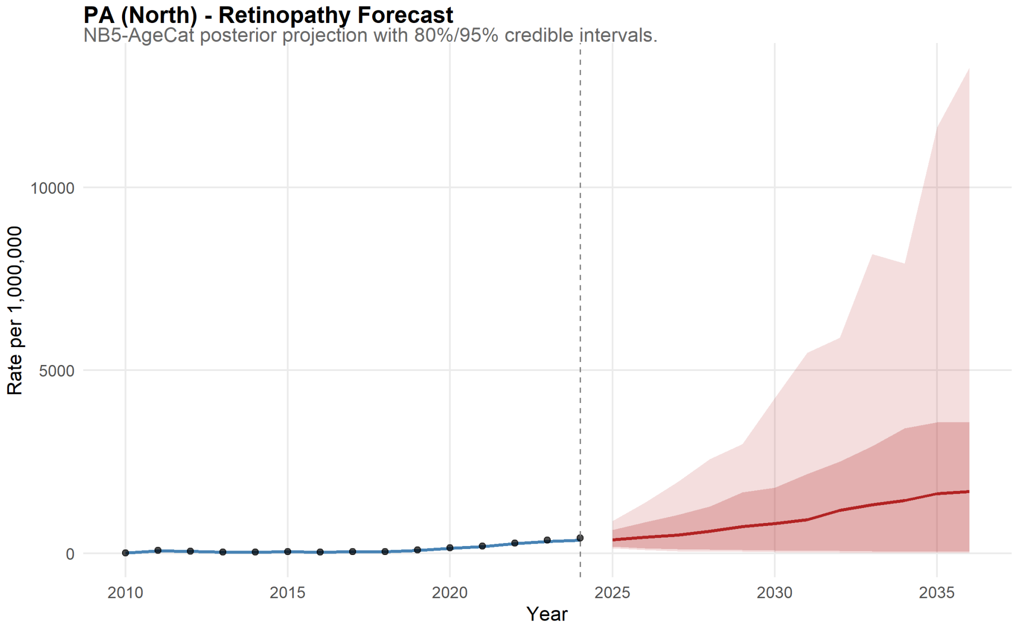

Figure 120: Pará Forecast for Retinopathy.

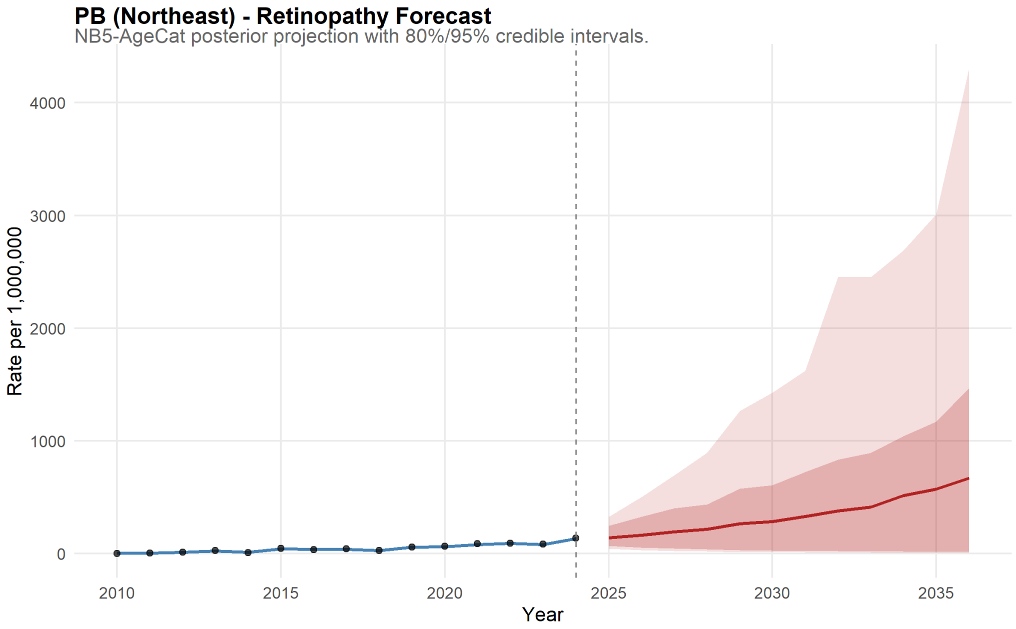

Figure 121: Paraíba Forecast for Retinopathy.

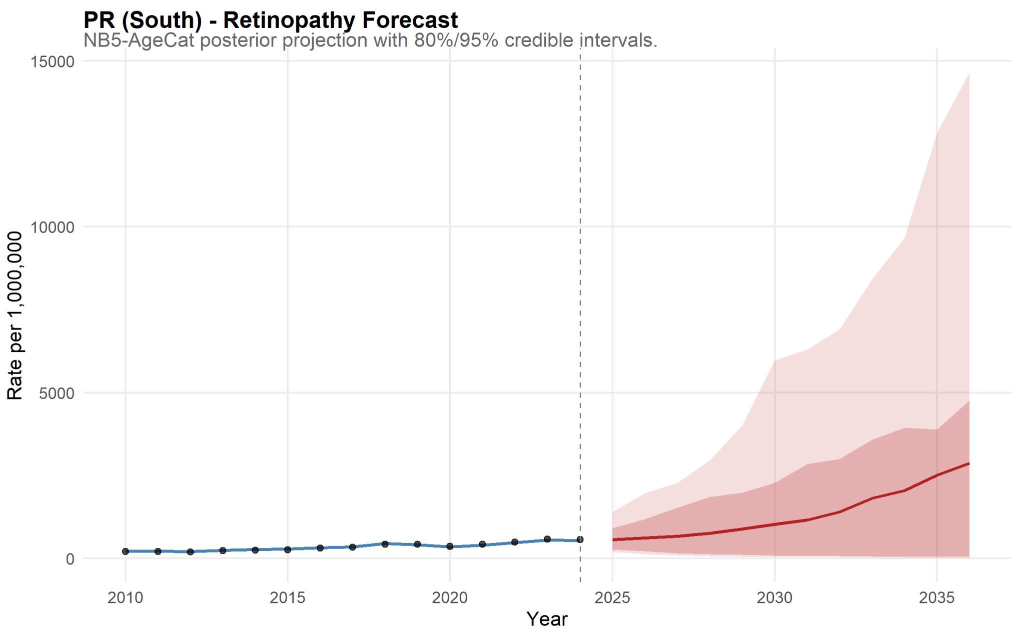

Figure 122: Paraná Forecast for Retinopathy.

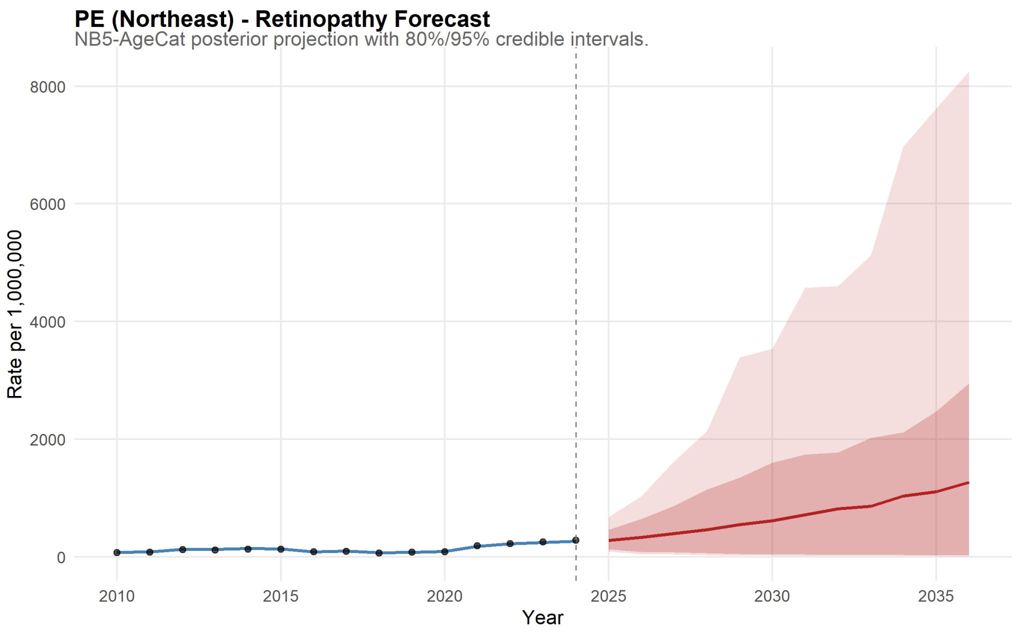

Figure 123: Pernambuco Forecast for Retinopathy.

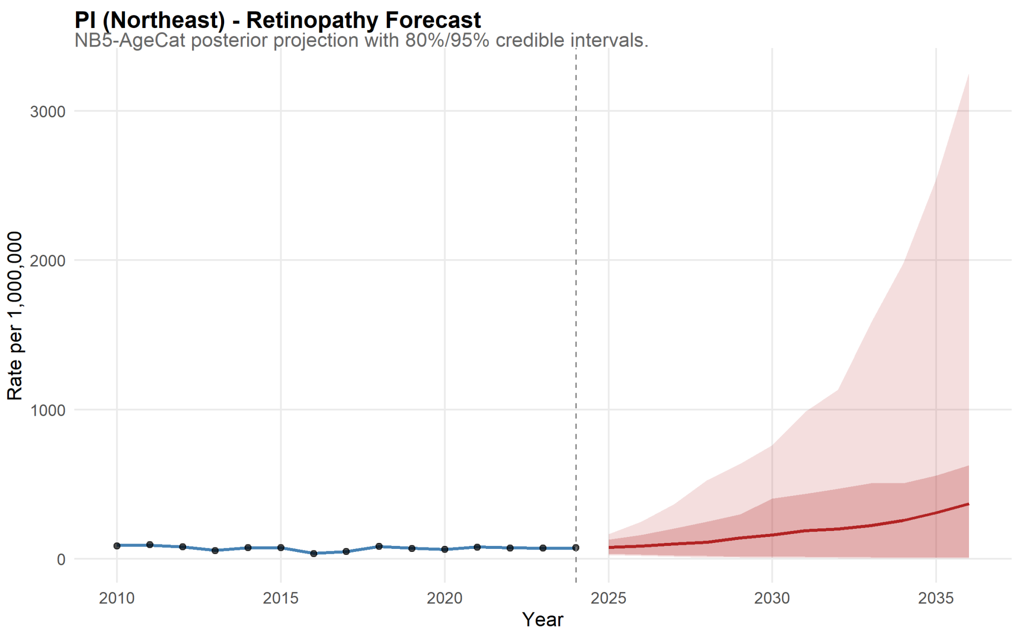

Figure 124: Piauí Forecast for Retinopathy.

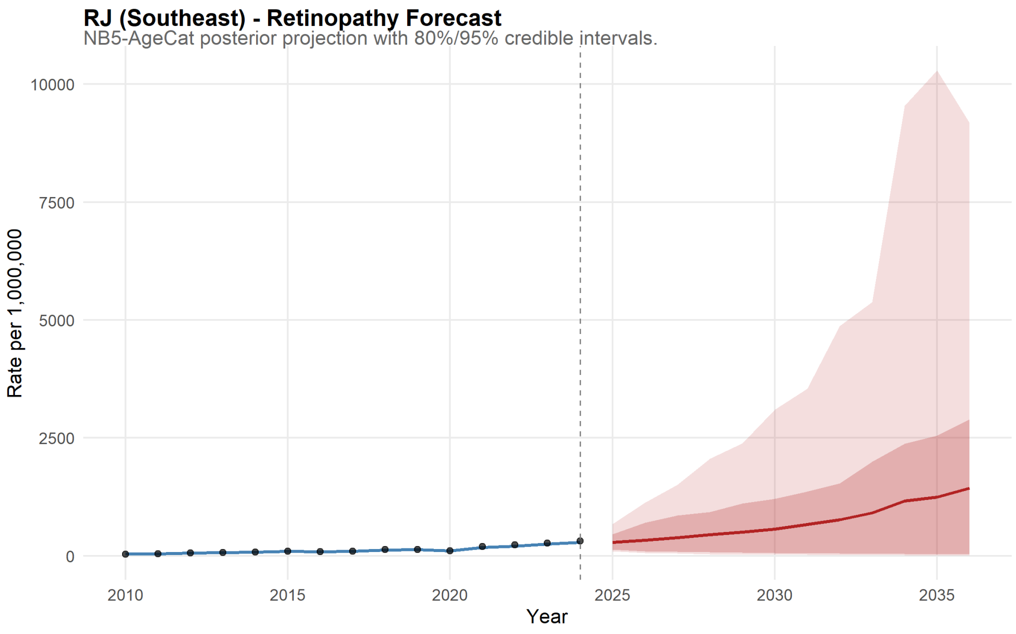

Figure 125: Rio de Janeiro Forecast for Retinopathy.

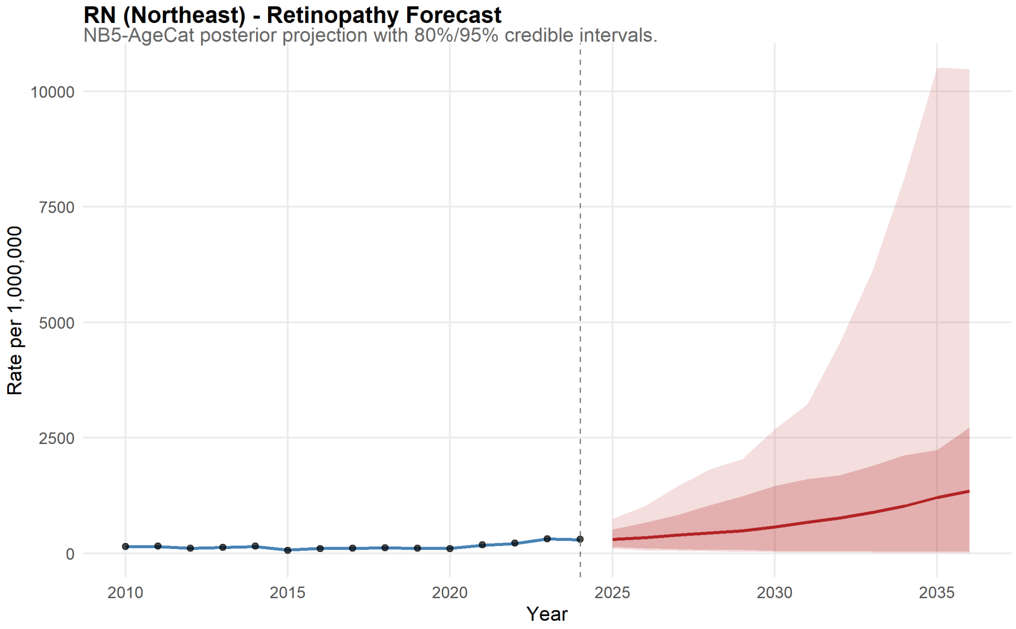

Figure 126: Rio Grande do Norte Forecast for Retinopathy.

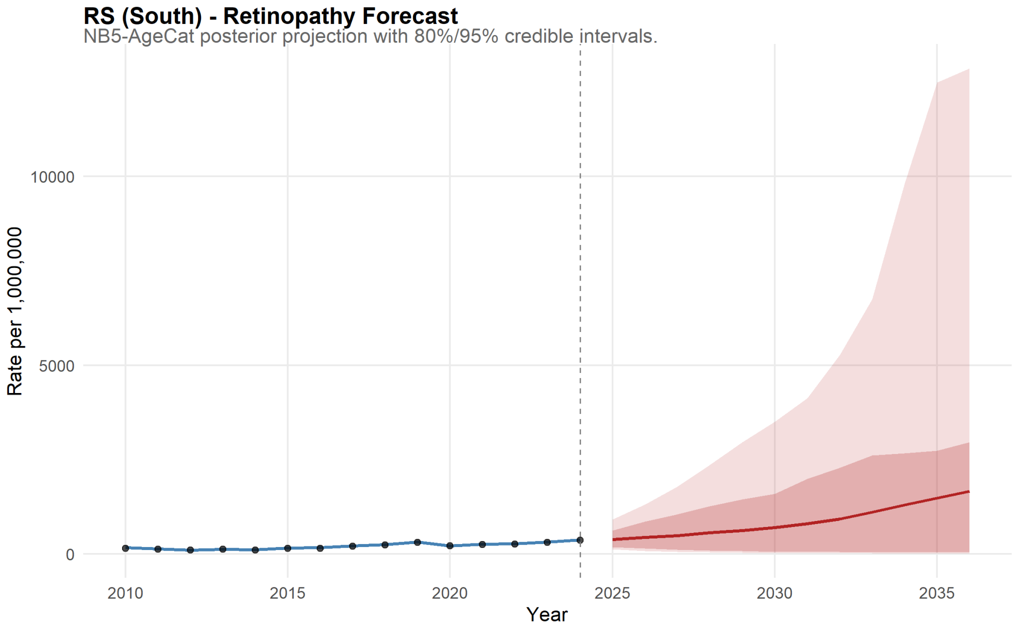

Figure 127: Rio Grande do Sul Forecast for Retinopathy.

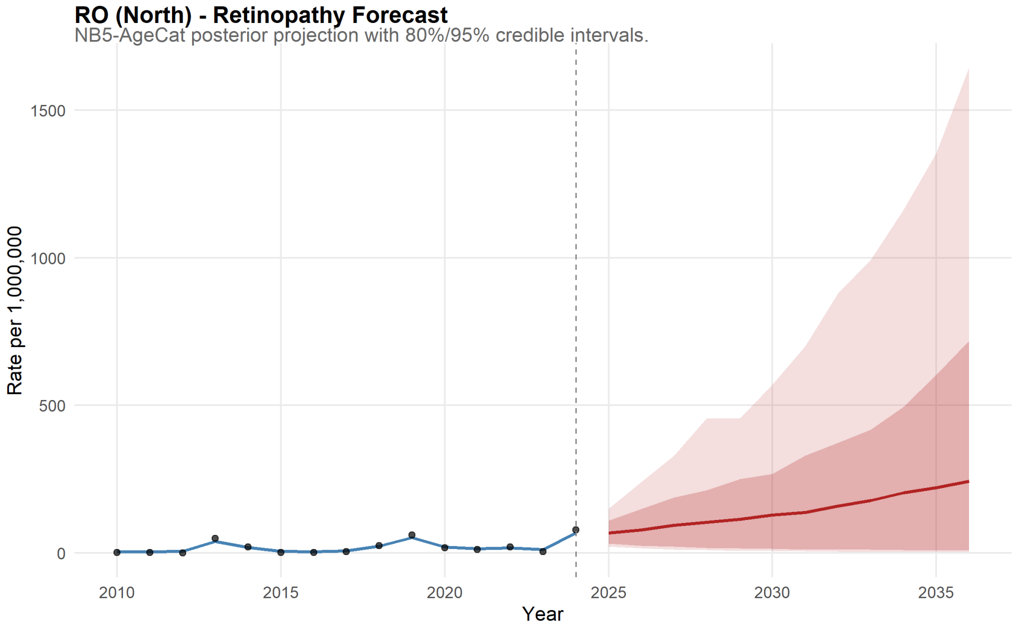

Figure 128: Rondônia Forecast for Retinopathy.

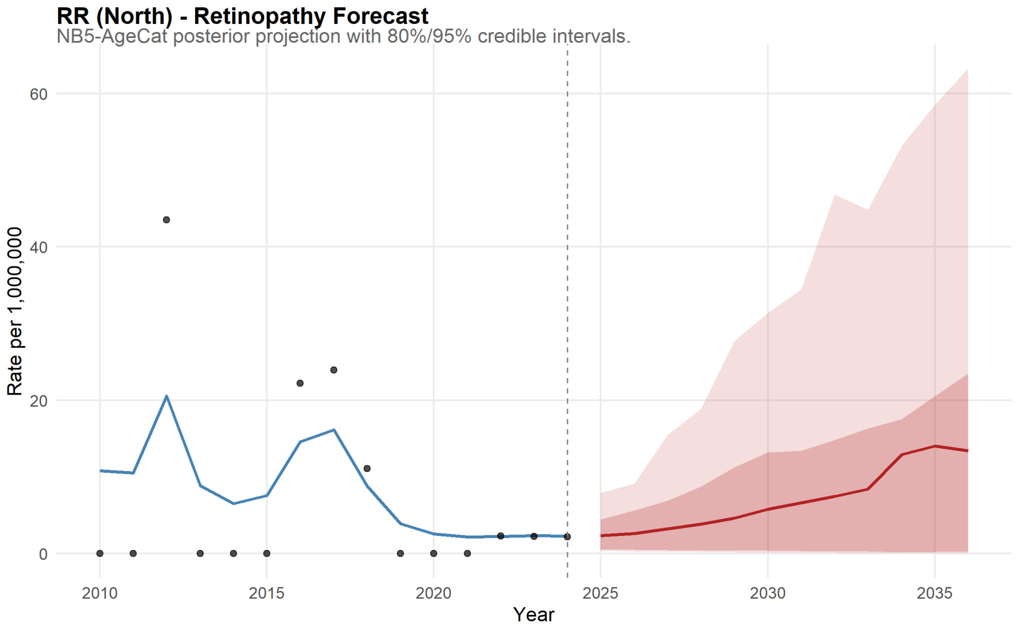

Figure 129: Roraima Forecast for Retinopathy.

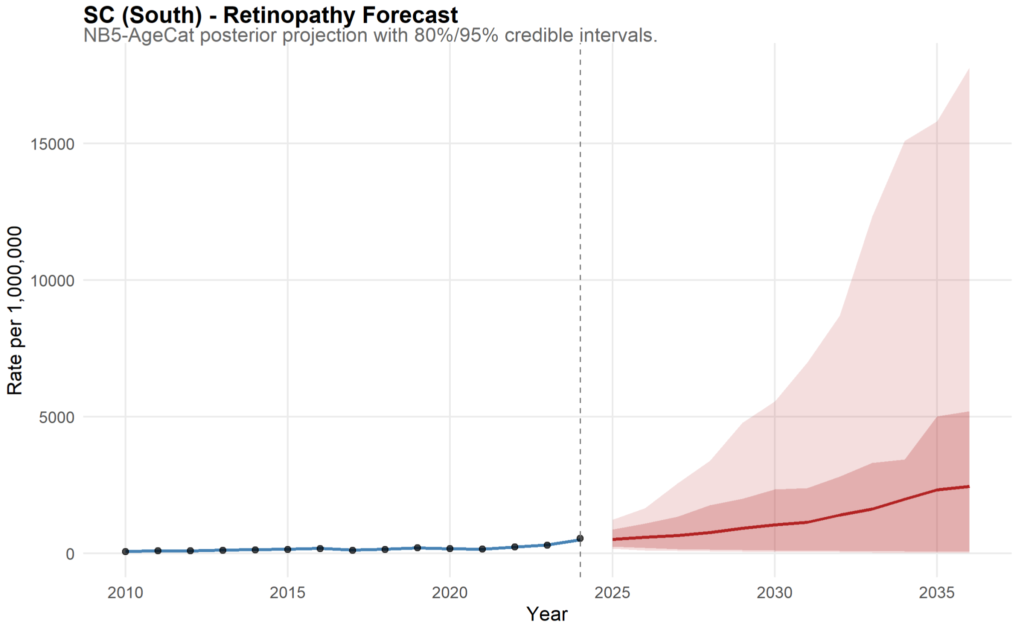

Figure 130: Santa Catarina Forecast for Retinopathy.

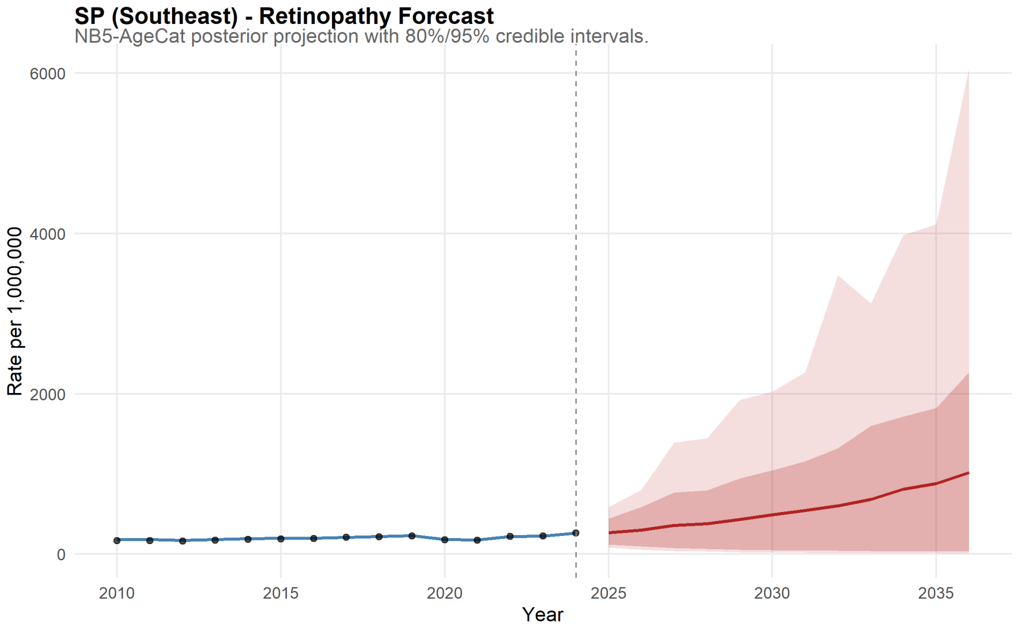

Figure 131: São Paulo Forecast for Retinopathy.

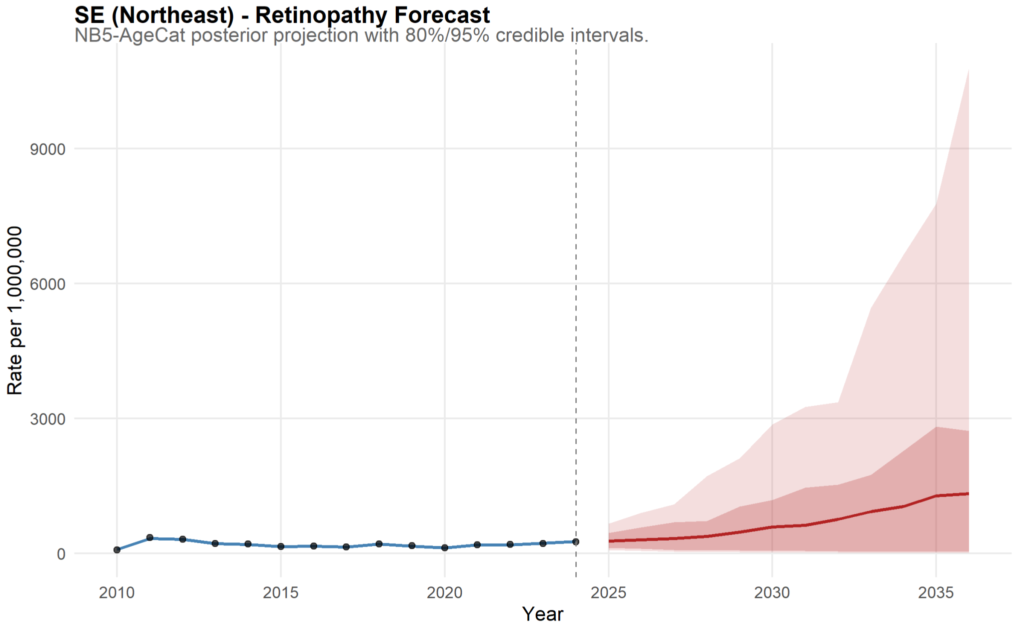

Figure 132: Sergipe Forecast for Retinopathy.

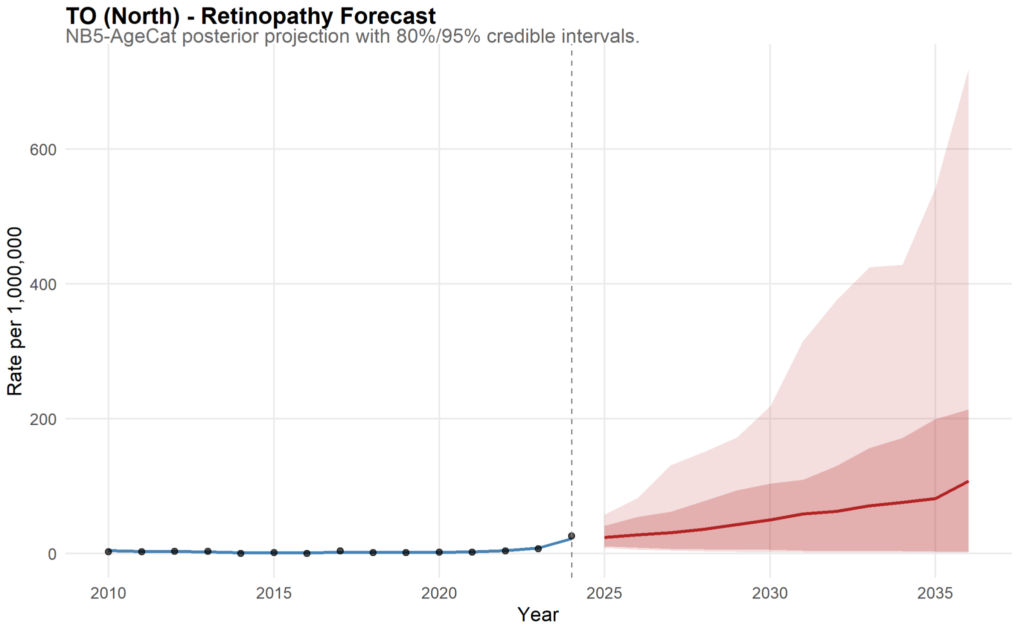

Figure 133: Tocantins Forecast for Retinopathy.

##### 11.2.3 Eye & Appendage Diseases

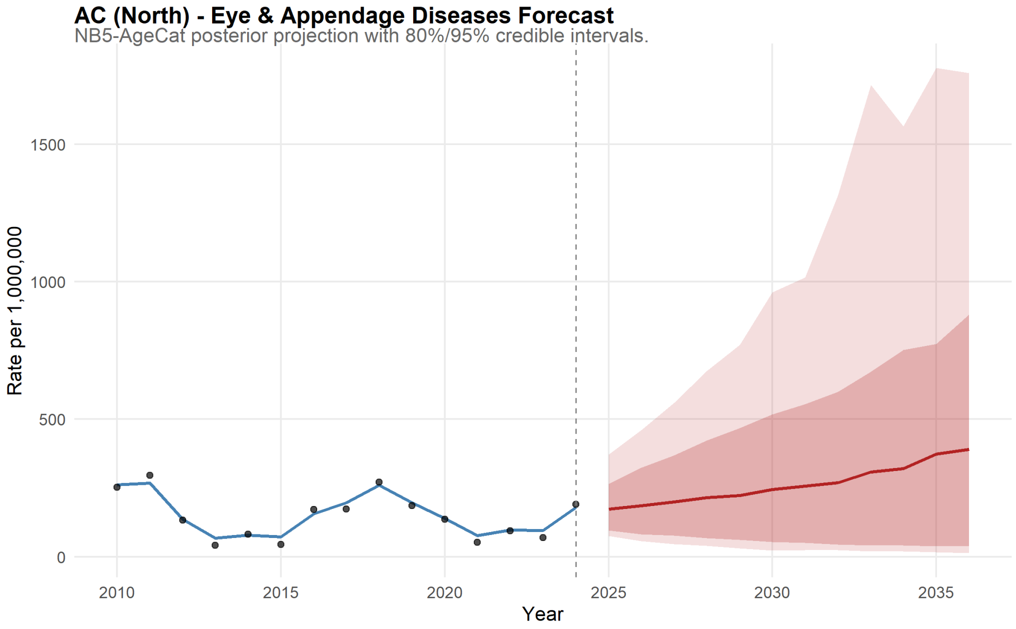

Figure 134: Acre Forecast for Eye & Appendage Diseases.

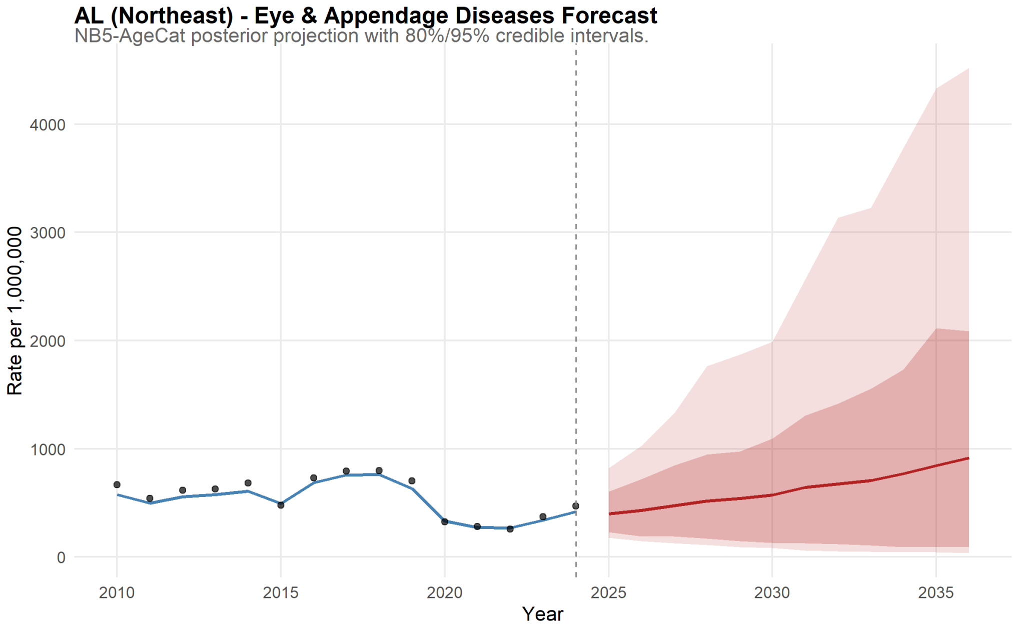

Figure 135: Alagoas Forecast for Eye & Appendage Diseases.

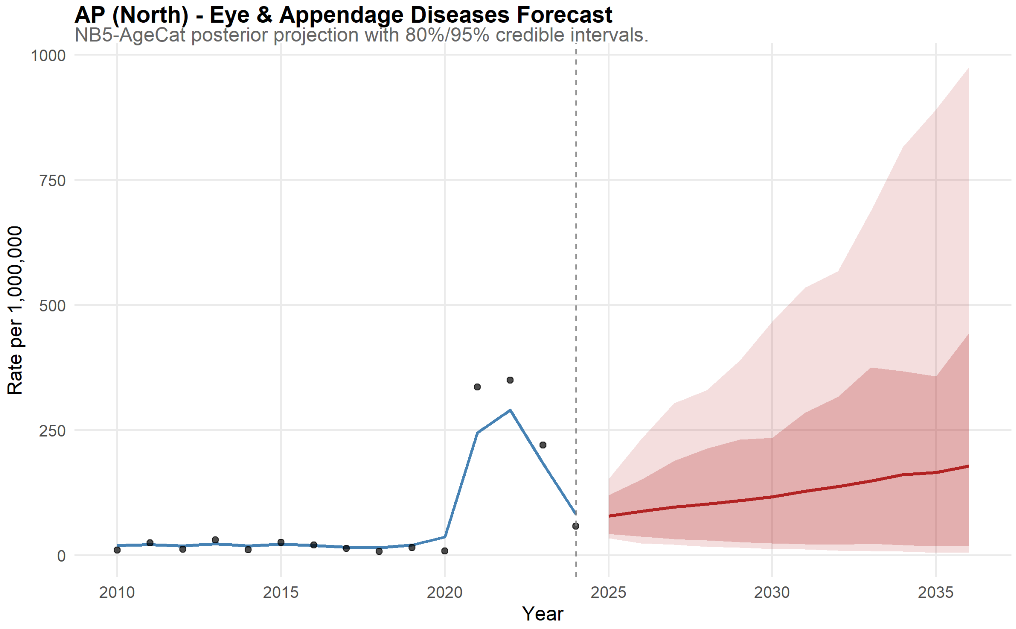

Figure 136: Amapá Forecast for Eye & Appendage Diseases.

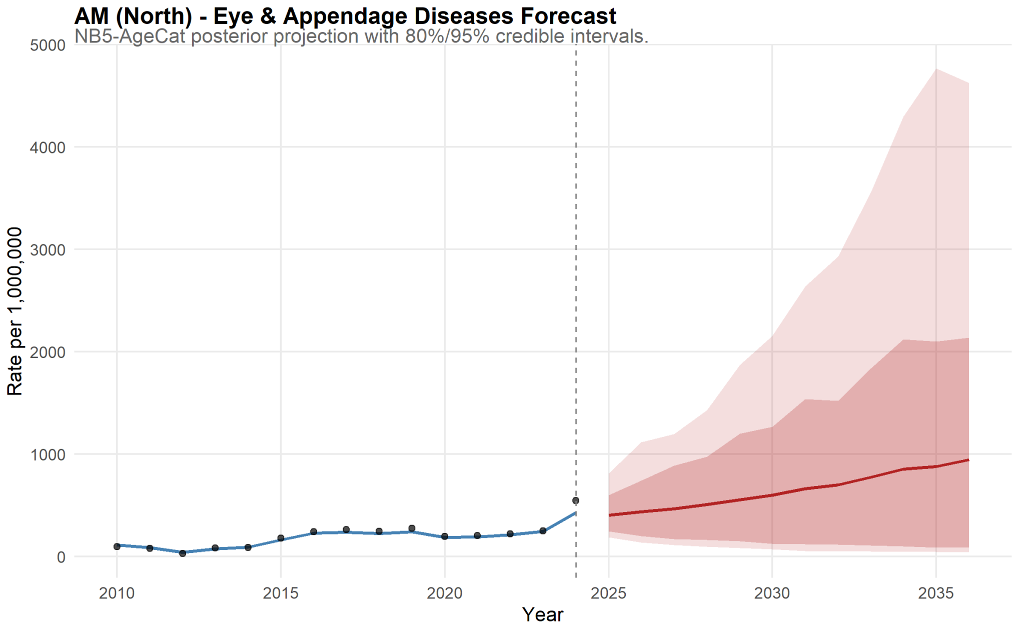

Figure 137: Amazonas Forecast for Eye & Appendage Diseases.

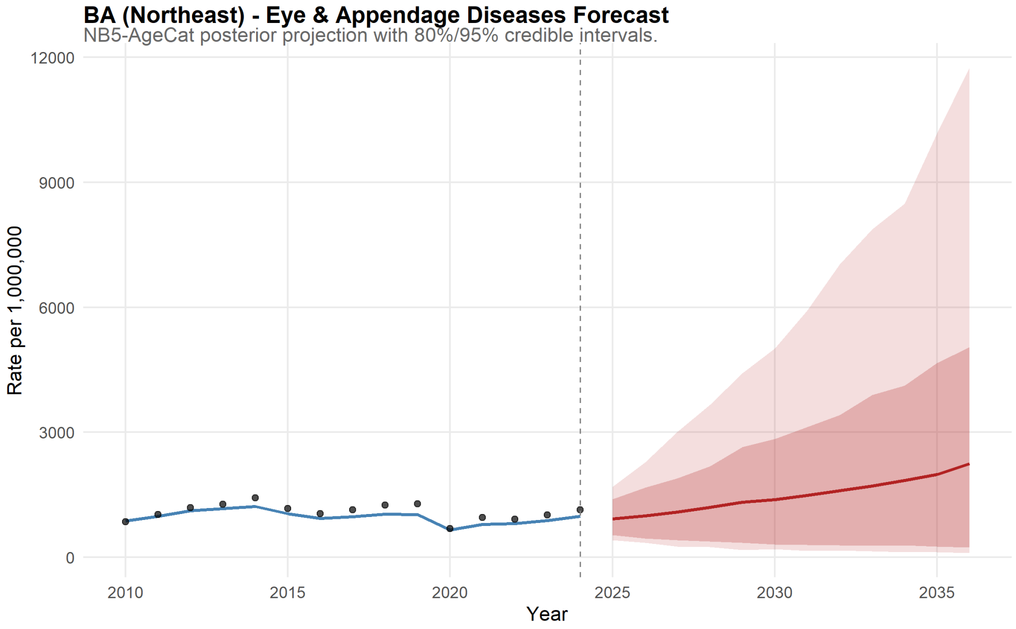

Figure 138: Bahia Forecast for Eye & Appendage Diseases.

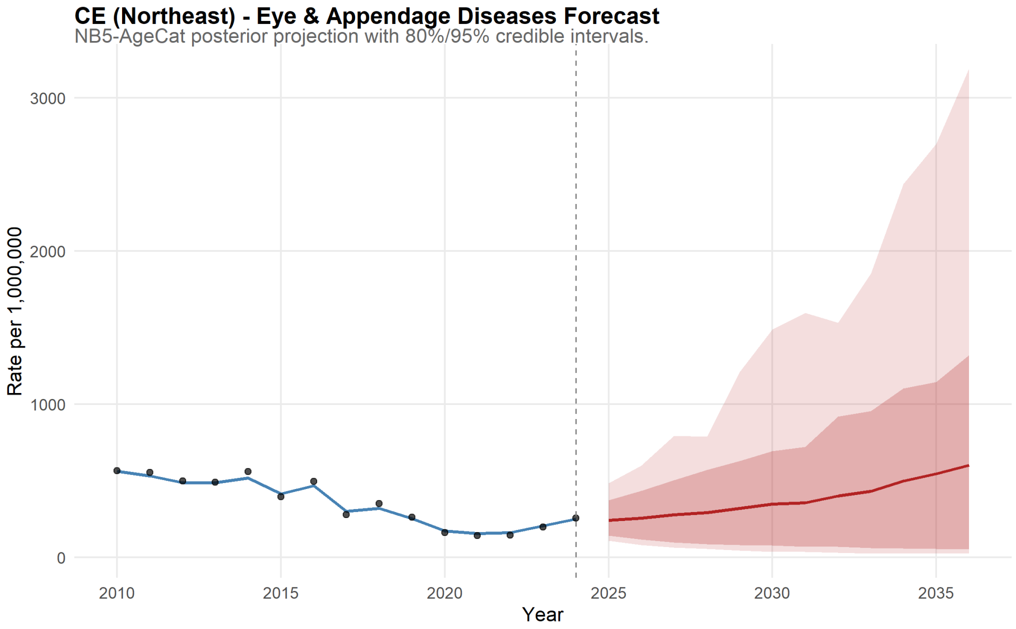

Figure 139: Ceará Forecast for Eye & Appendage Diseases.

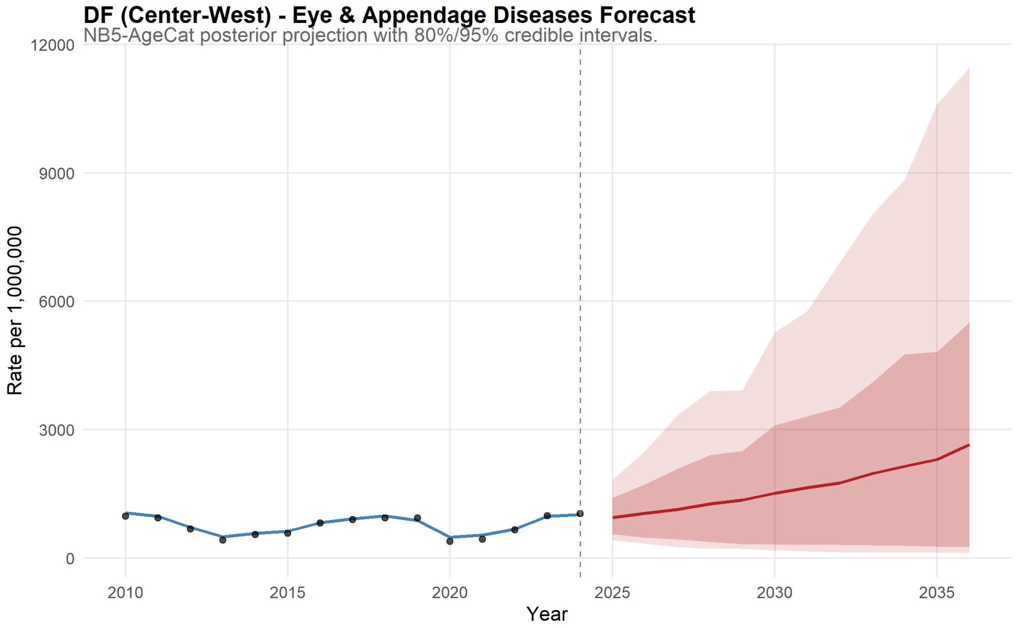

Figure 140: Distrito Federal Forecast for Eye & Appendage Diseases.

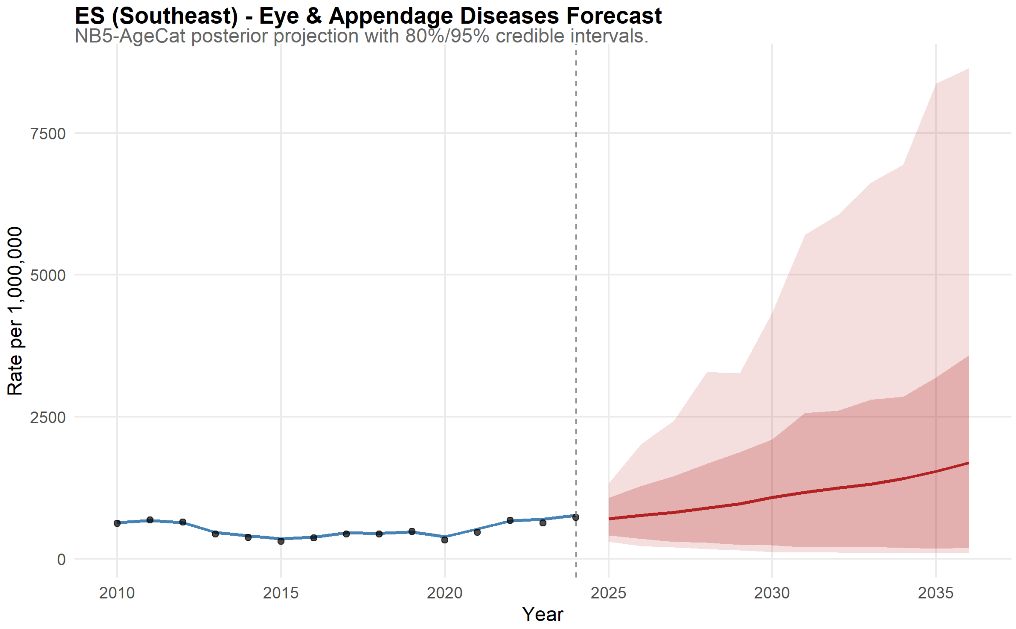

Figure 141: Espírito Santo Forecast for Eye & Appendage Diseases.

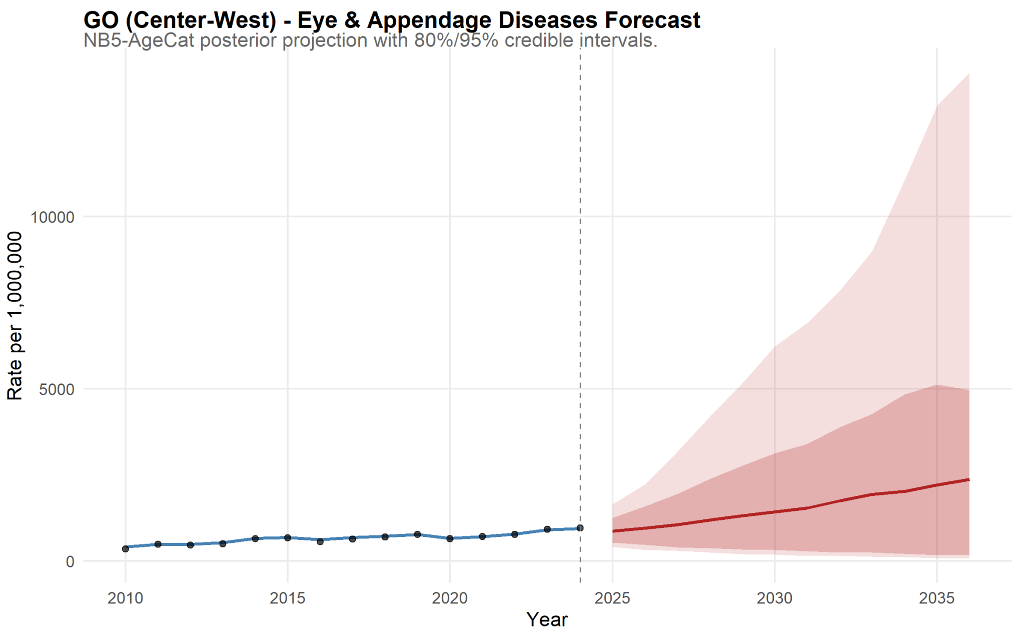

Figure 142: Goiás Forecast for Eye & Appendage Diseases.

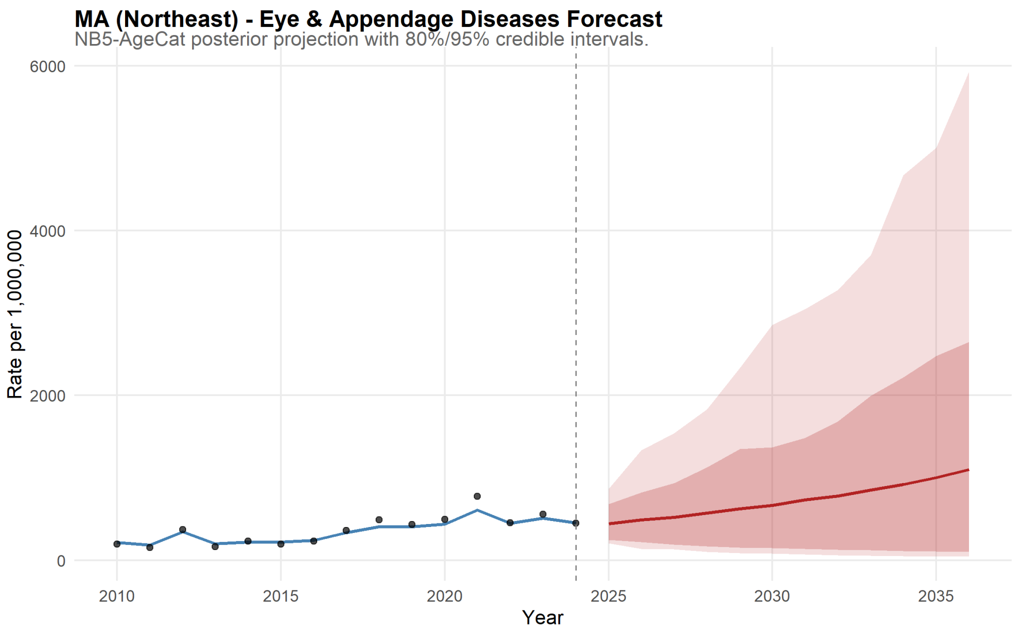

Figure 143: Maranhão Forecast for Eye & Appendage Diseases.

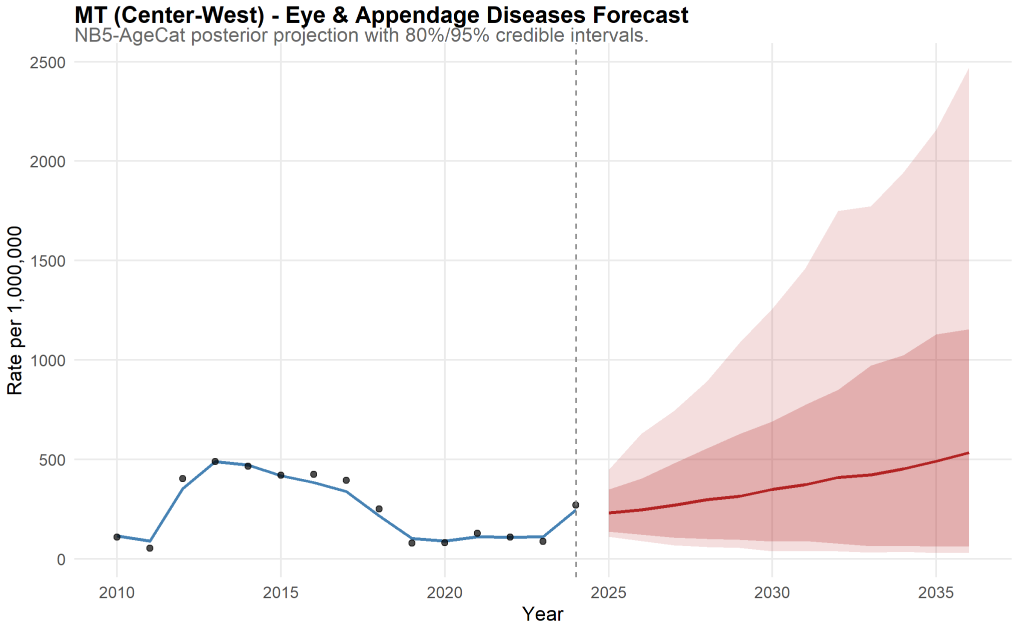

Figure 144: Mato Grosso Forecast for Eye & Appendage Diseases.

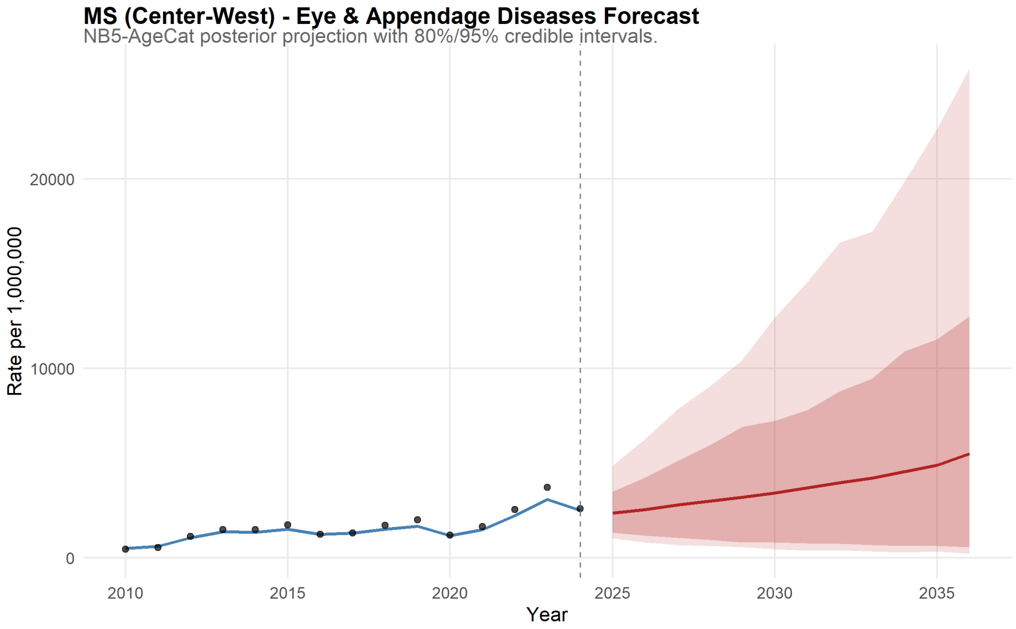

Figure 145: Mato Grosso do Sul Forecast for Eye & Appendage Diseases.

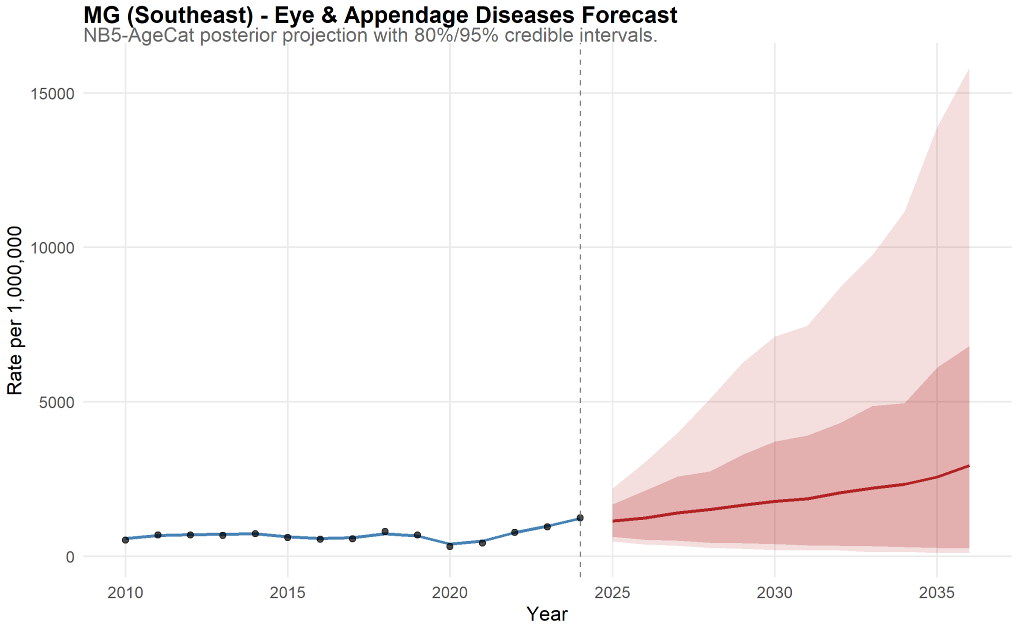

Figure 146: Minas Gerais Forecast for Eye & Appendage Diseases.

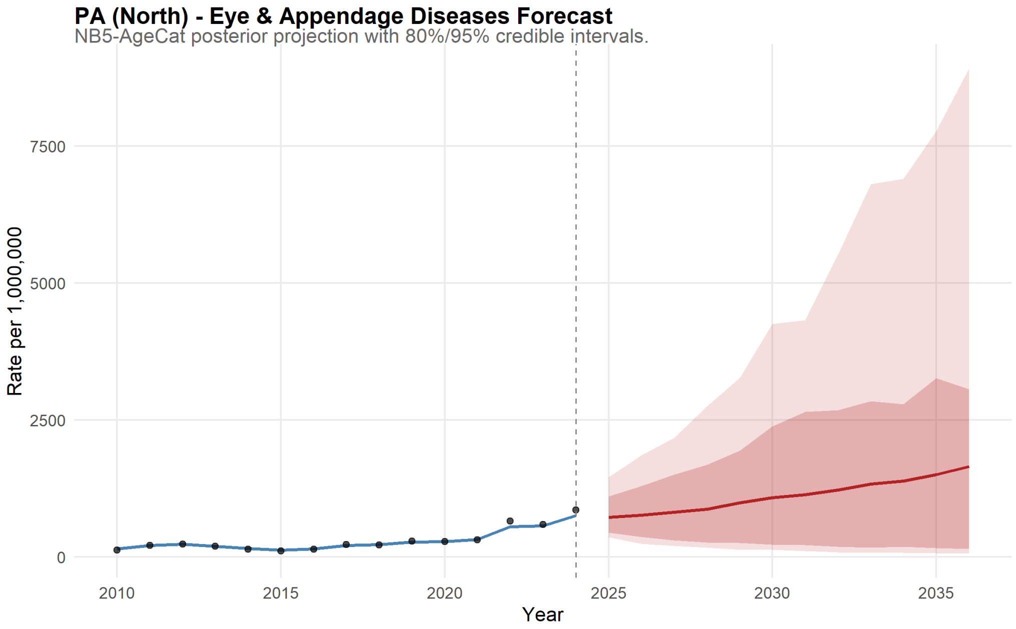

Figure 147: Pará Forecast for Eye & Appendage Diseases.

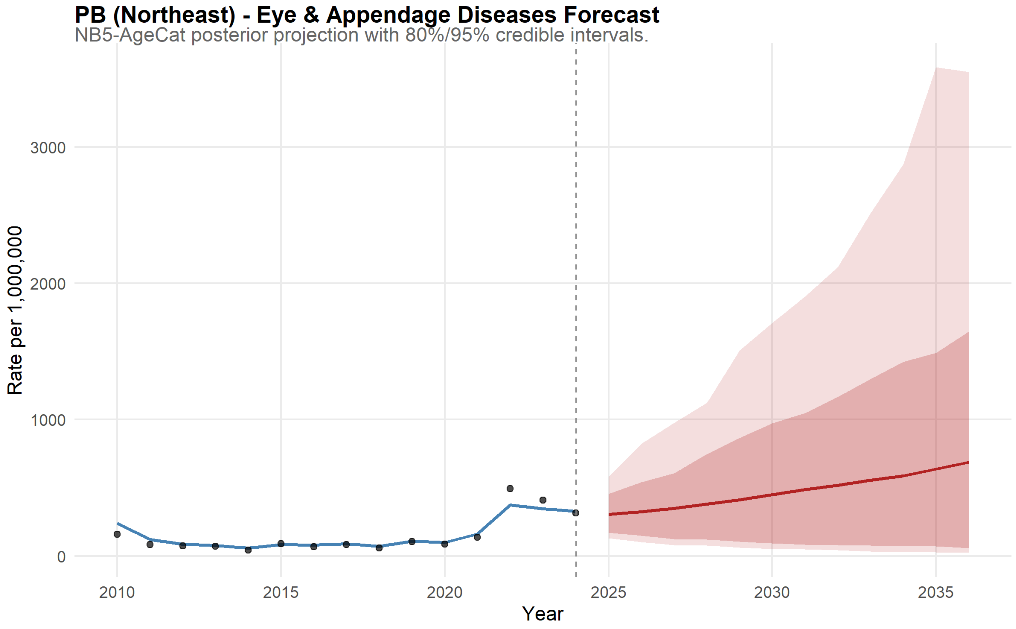

Figure 148: Paraíba Forecast for Eye & Appendage Diseases.

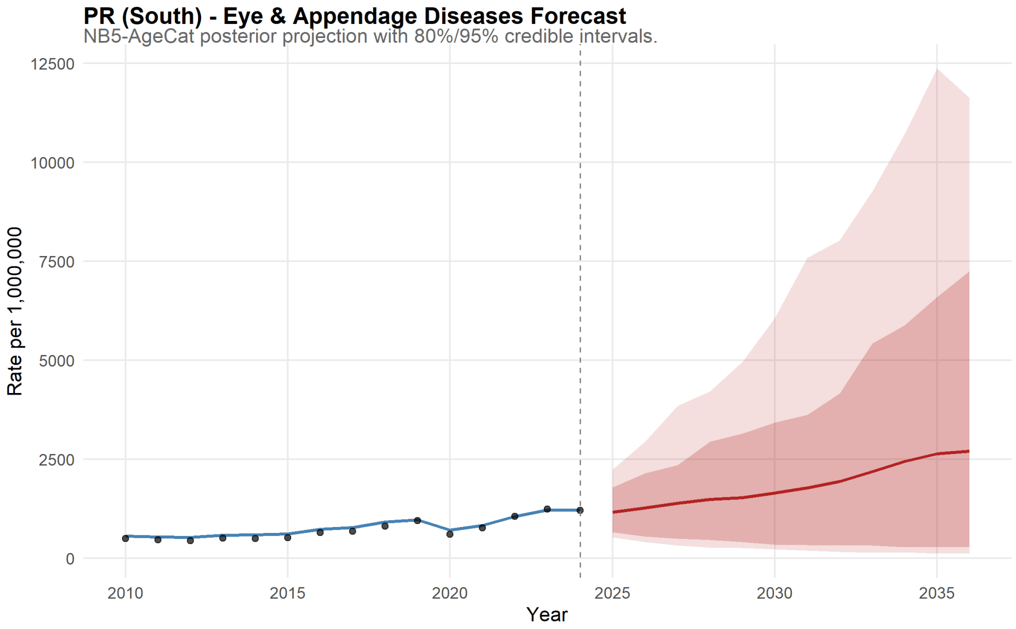

Figure 149: Paraná Forecast for Eye & Appendage Diseases.

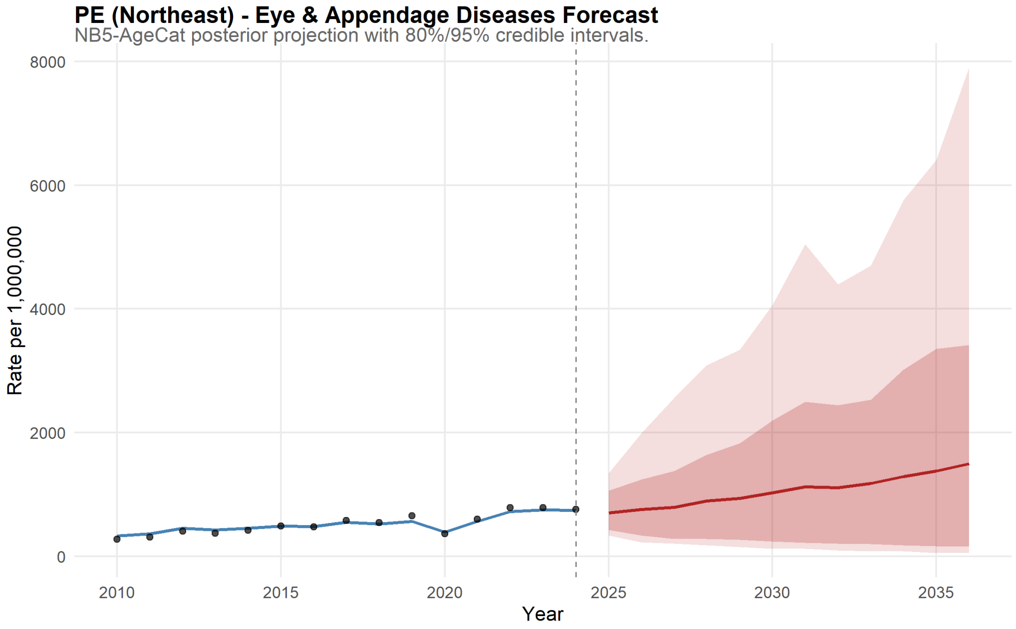

Figure 150: Pernambuco Forecast for Eye & Appendage Diseases.

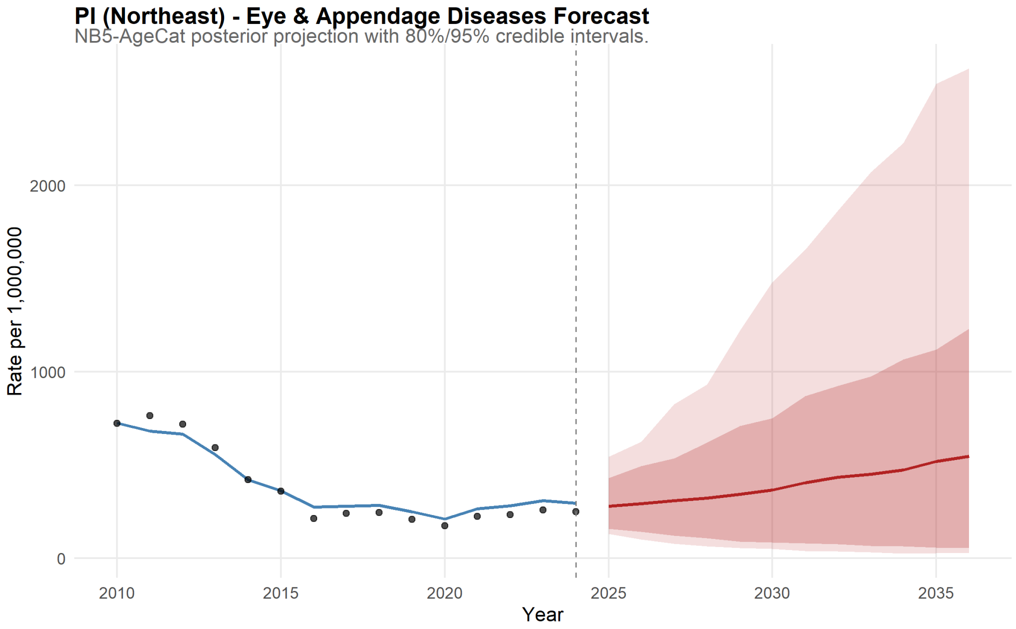

Figure 151: Piauí Forecast for Eye & Appendage Diseases.

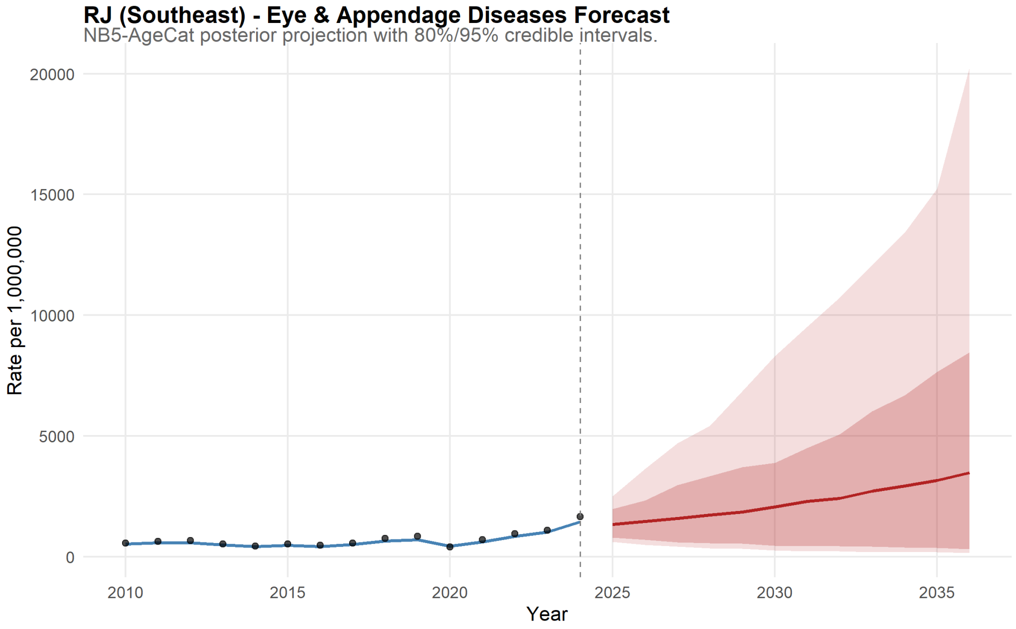

Figure 152: Rio de Janeiro Forecast for Eye & Appendage Diseases.

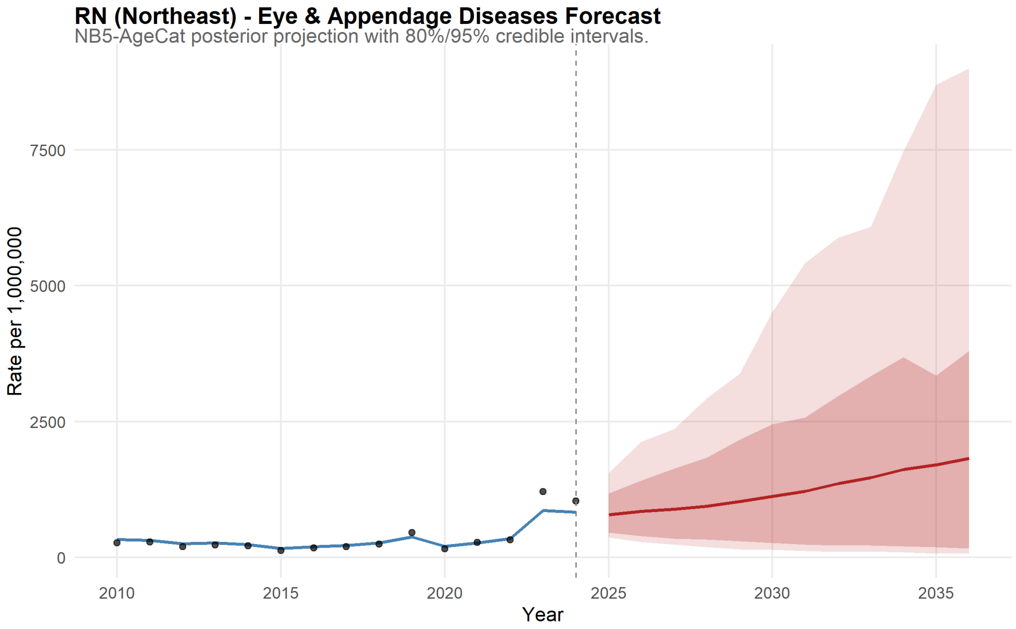

Figure 153: Rio Grande do Norte Forecast for Eye & Appendage Diseases.

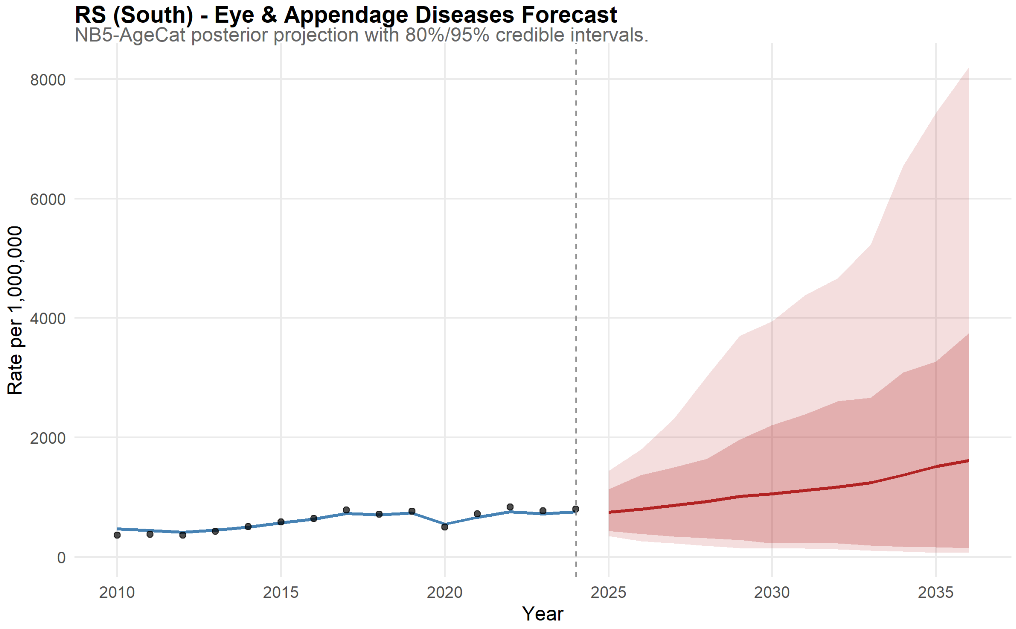

Figure 154: Rio Grande do Sul Forecast for Eye & Appendage Diseases.

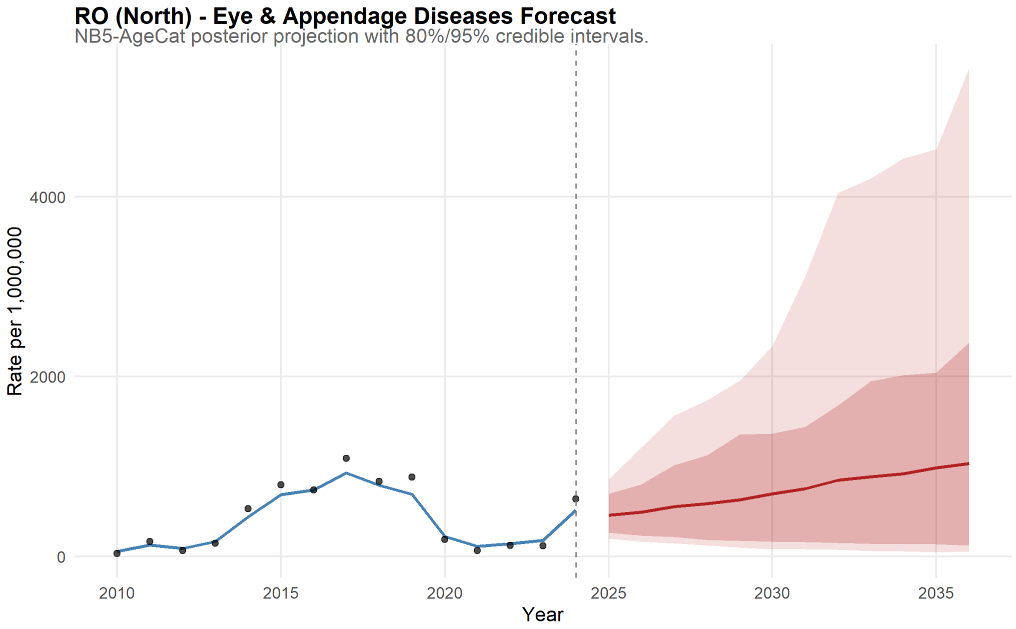

Figure 155: Rondônia Forecast for Eye & Appendage Diseases.

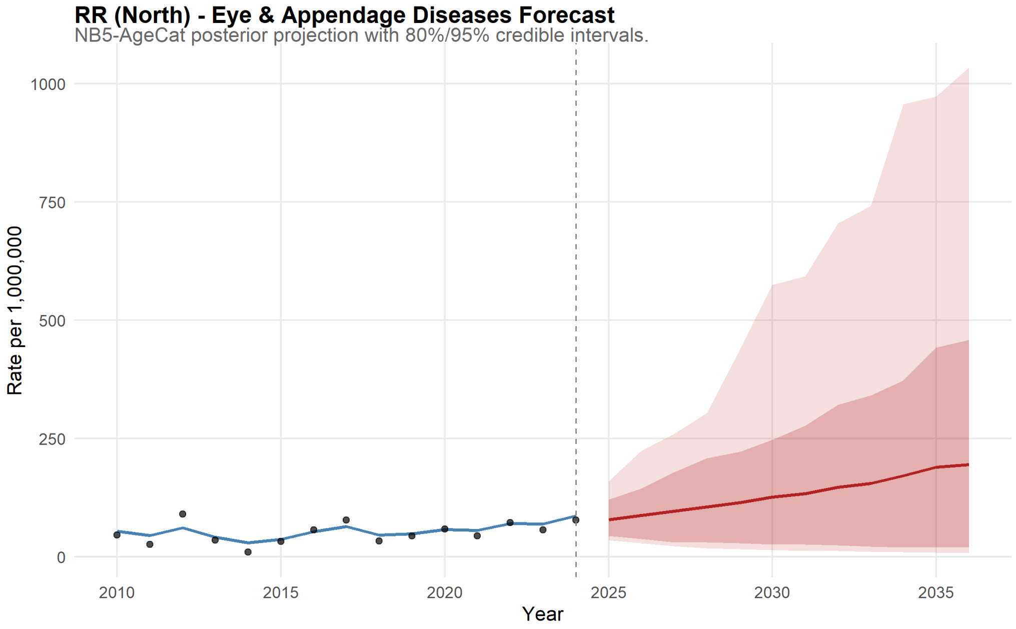

Figure 156: Roraima Forecast for Eye & Appendage Diseases.

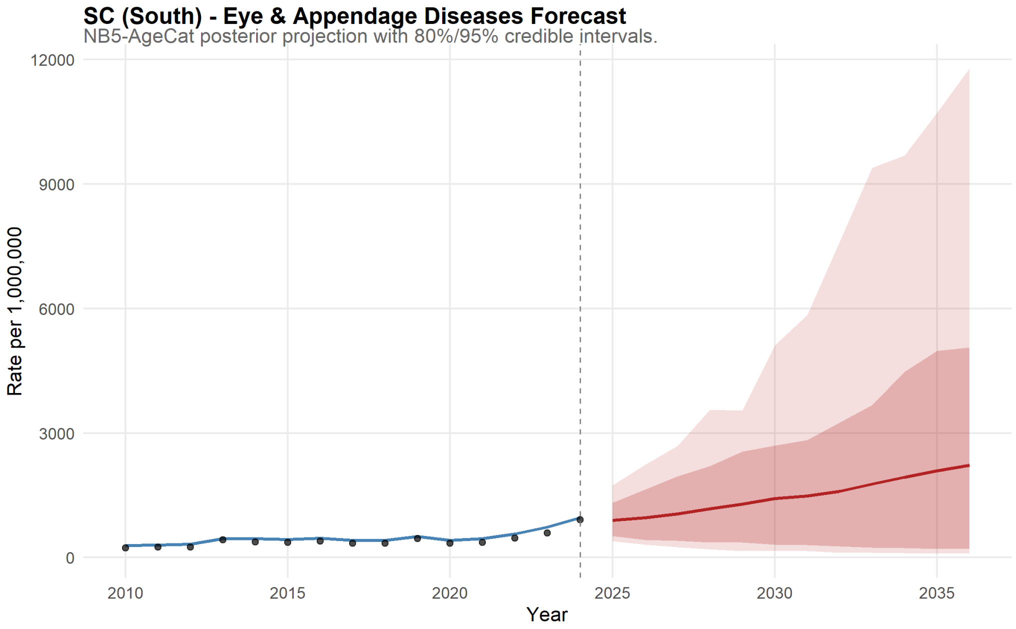

Figure 157: Santa Catarina Forecast for Eye & Appendage Diseases.

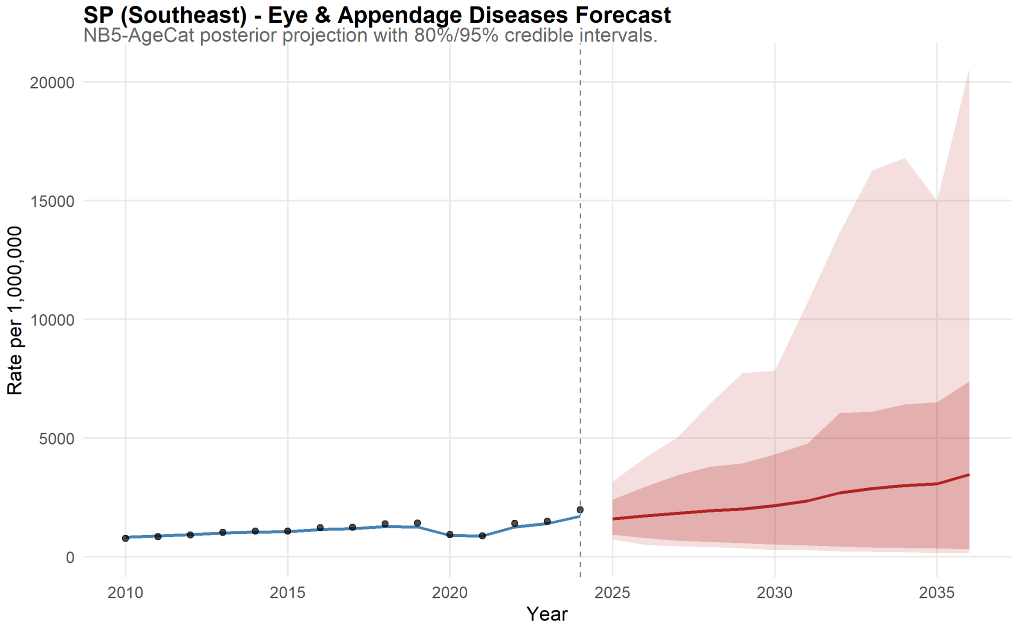

Figure 158: São Paulo Forecast for Eye & Appendage Diseases.

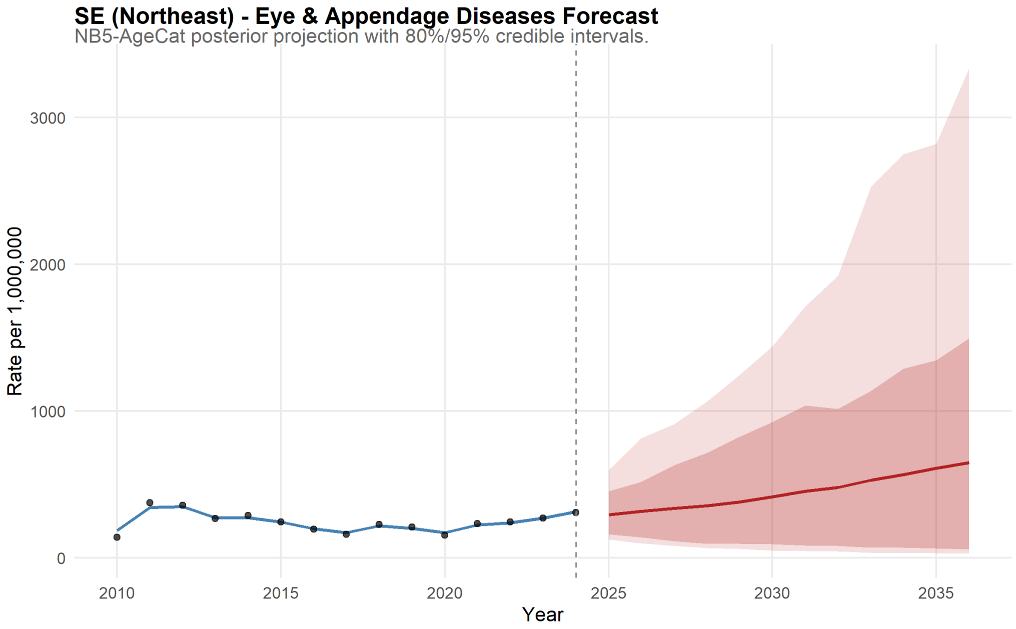

Figure 159: Sergipe Forecast for Eye & Appendage Diseases.

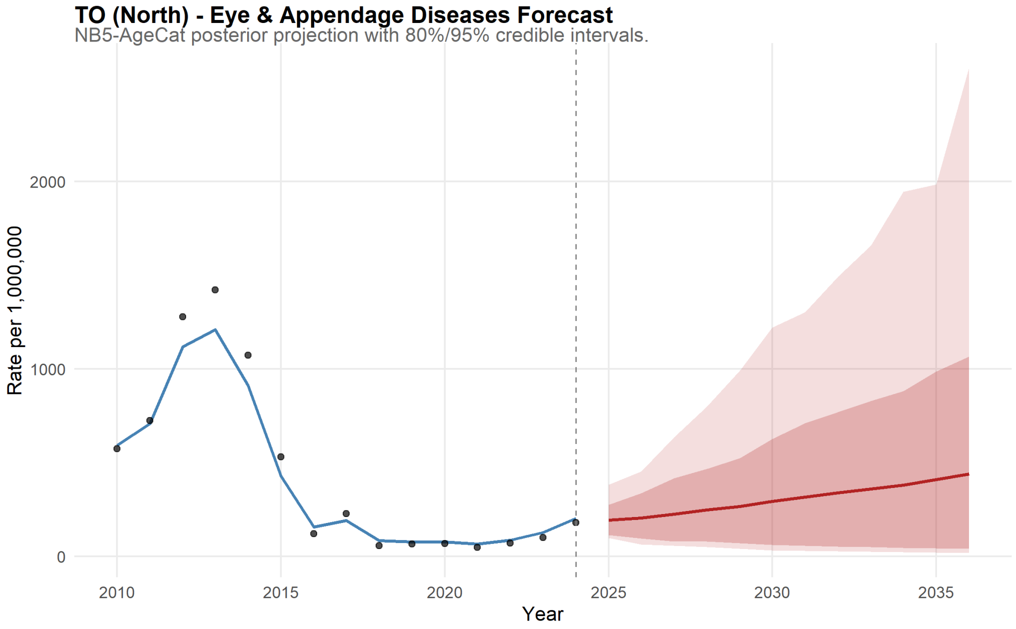

Figure 160: Tocantins Forecast for Eye & Appendage Diseases.
